# VascFlux-GEO: Comparator selection and auditability in vascular transcriptome module prediction

**DOI:** 10.64898/2026.10.25.26366405

**Authors:** Lingsen You, Zisu Peng, Yujun Guo, Li Shen, Junbo Ge

## Abstract

**Background:** Mathematical coupling and comparator selection are established concerns when a transcriptome response is defined as follow-up minus baseline. We assessed whether apparent within-cohort improvement survives matched constant-follow-up and scalar-shrinkage controls, and whether specified evaluation faults are detectable in an auditable vascular transcriptome benchmark.

**Results:** Using eight frozen modules, 72 rosuvastatin pairs, 14 peripheral artery disease pairs, 20 repeated subject-held-out five-fold partitions, and 5,000 subject bootstrap draws of fixed losses, we compared 15 explicit specifications. Supporting post hoc analyses included 800 known-truth training/test simulations, annotation and input-ablation sensitivities, complete-tuning integrity checks, and an independent-source cross-sectional artery description. In GSE86216, nested follow-up ridge had lower observed repeated-CV mean absolute error than mean follow-up, 0.3280 versus 0.3405 (difference 0.0126; descriptive central 95% percentile range 0.0033–0.0224), while scalar shrinkage had lower observed error (0.3256). These ranges have no validated coverage guarantee. No full-matrix advantage was established; equivalence was not tested. In peripheral artery disease, nested ridge (0.3449) had slightly higher observed MAE than the median follow-up comparator (0.3409), and no advantage was established. Simulations illustrated how mean follow-up can outperform mean change without temporal signal, while the GSE281841 histological stenosis-grade description remained limited by anatomical imbalance, unconfirmed participant identity and an unresolved sample-count discrepancy.

**Conclusions:** These retrospective, nonconfirmatory comparisons show modestly lower observed within-cohort error than constant-follow-up controls in one cohort, without established full-matrix advantage. They do not partition prediction into mathematical, technical and biological components. Independent longitudinal validation and clinical utility remain unestablished.

## INTRODUCTION

When response is defined as follow-up minus baseline, baseline participates in both the predictor and the target. Mathematical coupling, regularized prediction and nested validation are established concepts [1, 2, 3]. The unresolved empirical question in small paired vascular transcriptome cohorts is whether apparent prediction gains survive matched constant-follow-up and scalar-shrinkage controls. A full cross-module mapping must be compared directly with those controls to establish its additional value.

Vascular biology motivates the tissue context, but does not by itself establish predictive validity. Experiments identifying PECAM-1, VE-cadherin, and VEGFR2 as an endothelial mechanosensory complex demonstrate that flow can alter vascular signalling [4]. Predicting expression trajectories requires explicit tissue, timing, treatment and platform records; it does not establish why an intervention works.

Clinical treatment comparisons require further caution. A nationwide observational study of intra-aortic balloon pump use in acute myocardial infarction with Killip class III or IV identified residual indication confounding and unavailable insertion times as limits to causal interpretation [5]. This provides clinical context for distinguishing prediction from treatment comparison; it supplies no validation of the present transcriptomic representation.

VascFlux-GEO examines these empirical questions using a fixed module representation and inspectable analysis records. We assess within-cohort prediction in the statin cohort, repeat the fitting procedure in a small PAD cohort, and test specified evaluation faults. Known-truth simulations illustrate comparator behavior, while cross-sectional sources provide descriptive context. All extensions followed inspection of the initial fixed-penalty implementation of the same analysis package. The contribution is a retrospective comparison and execution audit in these data, not a new learning algorithm or model of vascular flow; statin-model parameters are not transported to PAD and comparative treatment effects are not estimated.

## METHODS

### Study scope and public-data provenance

This secondary analysis used processed, publicly accessible human Gene Expression Omnibus (GEO) matrices [6, 7]. Four accessions and their roles were retained from the original benchmark [8, 9, 10, 11]; a fifth source was added for the cross-sectional description [12, 13] (Table 1). No participants were recruited or contacted, no identifiable records were accessed, and no restricted clinical outcomes or device database was added. The original source studies supplied biospecimen collection and preprocessing; our denominators describe records retained by explicit parsing and pairing rules.

**Table 1:**
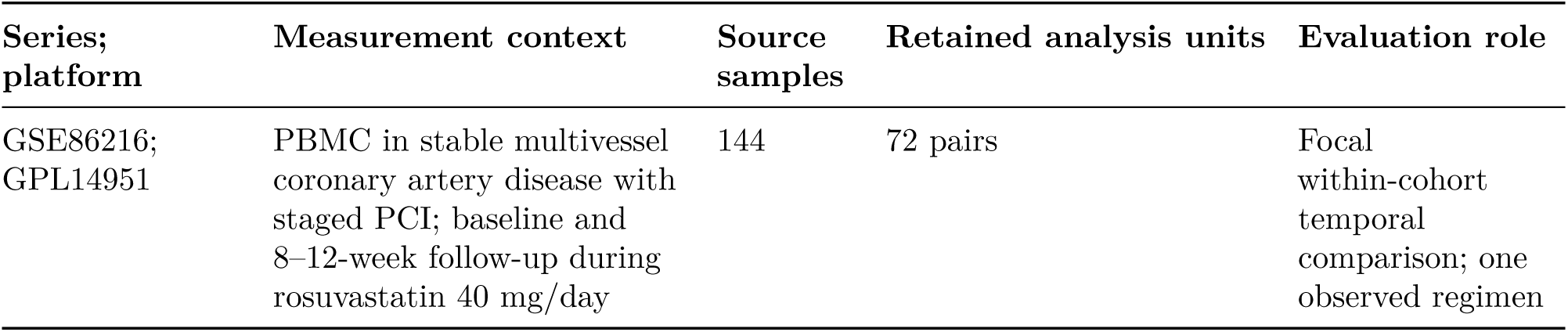
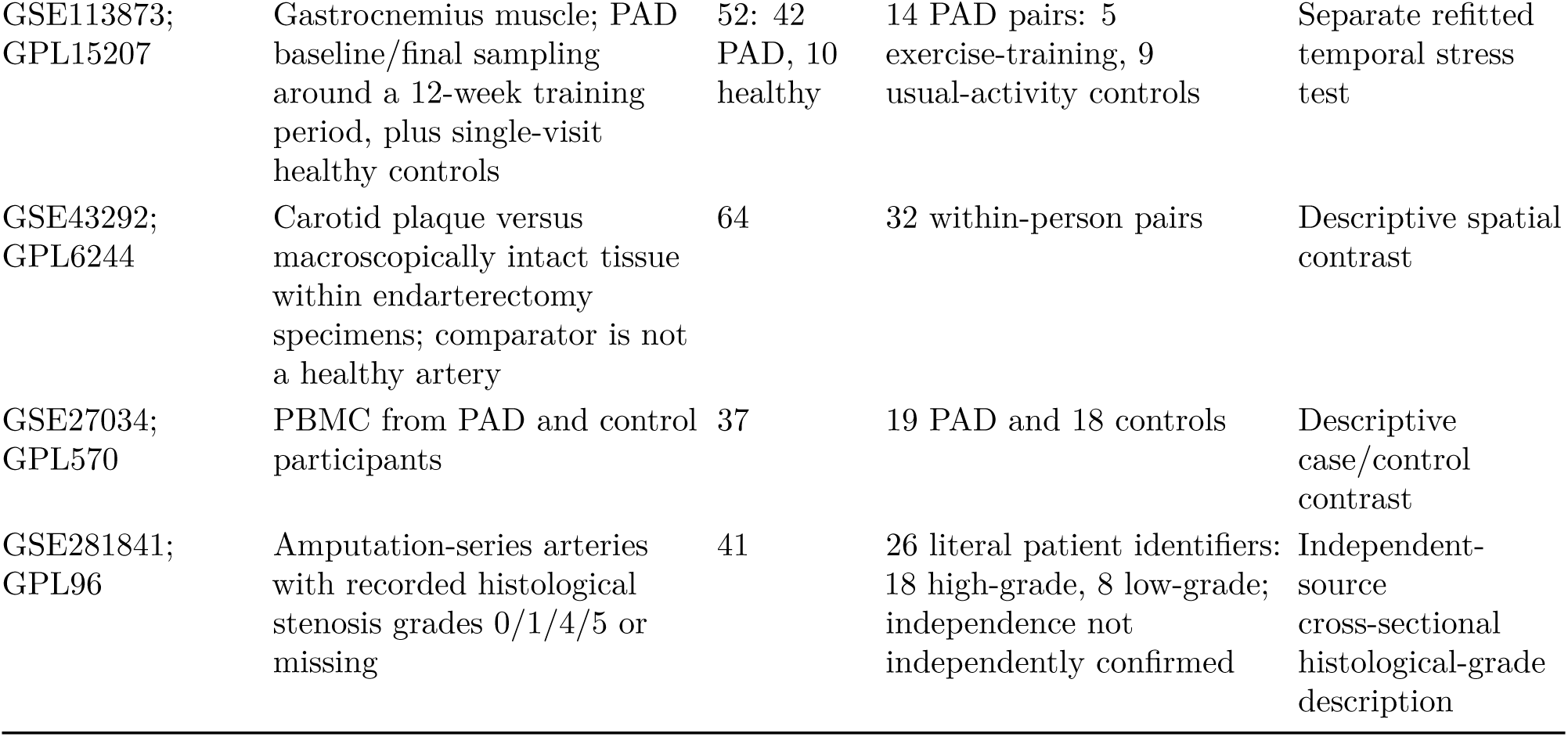
Dataset roles and analysis units. Source samples are matrix columns; longitudinal pairs, paired tissues, case/control participants and recorded patient-identifier clusters are different sampling units.

The extensions were designed after inspection of the earlier benchmark and were not prospectively registered. On 25 October 2026, all five complete processed matrices plus GPL15207 and GPL6244 were freshly downloaded and matched their frozen hashes. Complete first-platform tables for GPL14951 and GPL96 were separately obtained from official NCBI streams and matched the archived tables and module mappings; the full platform-family archives were not downloaded or verified. GPL570 initially failed the frozen-length download check; subsequent inspection identified an incomplete historical cache, and a complete official export replaced it with the resulting descriptive changes documented below. The current inputs therefore comprise eight freshly acquired complete files and two freshly validated complete platform tables, not ten freshly downloaded complete raw archives. Sample accessions were checked against matrix column order; repeated metadata rows were retained rather than overwritten. Prediction required exactly one eligible baseline/final pair per recorded subject, unique sample use, and metadata-consistent identity.

The GSE86216 pairing key was the deposited sibship (sampleid) field, not the numeric title prefix. Baseline records labelled control are the same participants before treatment, not an independent untreated arm. The cohort represents PBMC measurements in stable multivessel coronary artery disease during staged PCI and rosuvastatin treatment; it neither isolates a rosuvastatin effect nor measures coronary-wall expression. The parent study describes 85 participants, whereas the deposited matrix supplies 72 paired identifiers; the unavailable records cannot be assigned audited exclusion reasons here.

The 52 GSE113873 muscle samples represented 38 identifiers: 10 healthy participants sampled once by design and 28 participants labelled PAD. Fourteen PAD participants had eligible paired records; the remaining deposited PAD records comprised 12 baseline-only and 2 final-only participants. The archive does not establish clinical reasons for unavailable pairs. The source protocol specifies 12 weeks of treadmill training, three one-hour sessions per week with rest as needed; exact participant-level sampling intervals were not available for audit. GEO labels the control arm no exercise, but its protocol states that participants continued their usual activity. Machine-readable outputs retain the original label; the narrative uses usual-activity control. Exclusions and their reasons are exported, with the source-population distinction recorded in the accompanying dataset registry. The original source reports are cited alongside the accession records [14, 15, 16, 17] so that the biological design, sampling frame, and processed-matrix provenance can be independently checked. For GSE86216, the parent YELLOW II trial report describes the intervention design [14], whereas GEO directly links the diabetes-focused post hoc transcriptomic analysis [18]. These reports describe the same source study and do not constitute independent validation cohorts.

### Frozen modules and training-only transformations

The same eight modules were retained: endothelial activation, smooth-muscle contractile state, inflammation, extracellular matrix, coagulation/thrombosis, lipid handling, oxidative stress, and hypoxia/glycolysis. The manually curated lists were inherited unchanged from the earlier benchmark. No prospective timing claim is made; the revised analyses did not reselect genes using their outcomes. They overlap in genes and are neither independent replicates nor learned orthogonal factors. Module names label predefined gene sets. Their scores are not direct measurements of endothelial function, smooth-muscle contractility or pathway activity, and their interpretation may differ across PBMC, skeletal muscle and arterial tissue.

Platform probes were mapped using the original module-preferred first-symbol rule, with gene values summarized by the median across probes. Ambiguous annotations were retained for reproducibility. Annotation uncertainty was not propagated into the prediction ranges; two mapping rules were examined in a post hoc sensitivity analysis. For each predictive fit, gene means and population standard deviations came only from that training set’s baseline samples. Zero training variance received scale one. This transform was applied unchanged to baseline and follow-up samples, and each module was the median of its available standardized genes. Inner folds refitted these transformations from their own training baselines. Restricting processing to the frozen module-gene union preserved the original per-gene normalization numerically. GSEA and GSVA are established gene-set approaches [19, 20]; neither was used to construct these fixed median scores. Platform preprocessing was inherited from the deposited processed matrices rather than recomputed from raw probe intensities; robust multi-array normalization is an established method for that upstream task [21]. Training-only transformations refer to secondary gene/module scaling and model fitting; deposited preprocessing was not reconstructed within cross-validation. GSE113873, GSE43292, GSE27034 and GSE281841 report upstream RMA, while GSE86216 lists processing software without a complete reproducible sequence. The dataset registry records these inherited steps and unresolved input-scale details.

### Outcome identity, predictors, and context

Let B and F denote eight-module baseline and follow-up vectors, with Δ = F − B. Baseline–change association can reflect mathematical coupling and regression to the mean [1]. For any direct follow-up prediction, implied change is *F*^ − B, and its absolute error equals the follow-up error. We therefore compare all specifications on matched subjects and transforms rather than interpreting change and follow-up scores as independent evidence. For PAD, the predicted endpoint is follow-up muscle module expression, not functional improvement, limb events or survival.

Ridge models fit either Δ or F using baseline state and, where available, observed context [2]. The general predictor is *Y*^ = b + W x + C a, with an unpenalized intercept. Statin models omit a because all subjects share one regimen. PAD models include both unscaled treatment-group dummies, retaining the original redundant-intercept parameterization for reproducibility. Coefficients are not interpreted as treatment effects. The fixed-penalty anchor uses raw module features and *λ* = 0.01; nested ridge additionally standardizes module features within the current training set. Penalties across these feature scales are not directly comparable. Ridge, PCA and scalar shrinkage are standard prediction tools [22]; the empirical comparison concerns their behavior in these cohorts.

Eleven named specifications were evaluated: pooled mean change, pooled mean follow-up, persistence, group mean change, group mean follow-up, fixed change ridge, fixed follow-up ridge, nested change ridge, nested follow-up ridge, PCA-2 follow-up ridge, and scalar shrunken-state follow-up. Mean change predicts B_test + mean(F_train − B_train); mean follow-up predicts mean(F_train); persistence predicts B_test. Group controls use the observed group and fall back to a pooled mean if that training group is absent. In the single-regimen cohort, both group controls exactly duplicate their pooled counterparts. Fixed change/direct ridge are close reparameterizations, not independent architectures.

PCA-2 uses two components from singular-value decomposition of only the standardized training-baseline module matrix, followed by tuned follow-up ridge. It is a bounded module-level comparator, not a transcriptome-wide representation search. Scalar shrinkage predicts mean(F_train) + a[B_test − mean(B_train)], with a selected from {0, 0.25, 0.5, 0.75, 1}. It directly tests whether a full coefficient matrix is necessary for any modest prediction gain.

### Focal comparison and post hoc analysis hierarchy

The retrospectively designated focal comparison is the subject-level paired MAE difference between nested follow-up ridge and mean follow-up in GSE86216. This designation is retrospective and has no preregistration or confirmatory status. Positive differences favor the model. We retain all 11 original specifications and add four MAE-aligned controls: pooled and treatment-group median follow-up, and pooled and treatment-group median change. These use the identical exported splits and training-baseline transforms; group controls fall back to a pooled training median when the group is absent. They are additional specifications, not four new architectures.

The focal statin comparison, added median and scalar comparisons, separately refitted PAD analyses, simulations, representation sensitivities and descriptive contrasts address distinct questions. None is used to select a new primary winner. The lowest observed error among nine untuned mean, median and persistence controls is a descriptive ranking, not a locked selection strategy. All additions followed inspection of earlier results and remain exploratory; percentile resampling ranges do not account for comparator selection or multiple comparisons. Finite tuning samples and repeated comparator inspection can favor optimistic performance estimates [23].

The retrospective question map separates four claims. First, improvement beyond constant follow-up is assessed by comparator MAE minus model MAE on matched subjects and transforms; a nonpositive or uncertain difference does not support improvement. Second, additional value from a full matrix is assessed directly by scalar-shrinkage MAE minus ridge MAE; success against a constant control cannot substitute for this comparison. Third, known-truth simulations and coupling-preserving diagnostics ask whether an advantage over mean change can occur without temporal pairing information. Fourth, held-out-target perturbations and injected faults examine invariance and detection along the implemented evaluation paths. These questions organize post hoc evidence, not preregistered tests or a newly selected winning procedure. Failure to establish superiority is not evidence of equivalence; no clinically important MAE threshold has been established for these module scores.

### Repeated outer evaluation and nested selection

Each cohort underwent 20 repetitions of five-fold subject-held-out cross-validation. Every subject was tested once per repetition, with all visits held together and exact folds shared across methods. The first repetition reproduced the earlier random sequence, including PAD’s shuffle after the statin shuffle. The other seeds and assignments are exported. These are within-cohort random splits, not temporal, hospital, or external-study validation.

Hyperparameter selection within each specified candidate was confined to outer training data, following the separation of tuning and evaluation required for assessing a selected procedure [3]. This nesting did not include the post hoc development of candidate methods, comparator additions or focal-comparison designation. Statin fits used four inner folds; PAD and training subsets with fewer than 24 subjects used three. Candidate *λ* values were {0.01, 0.1, 1, 10, 100, 1000}. Hyperparameter selection minimized pooled inner-held-out module MAE; ties selected the first candidate. Standardization and PCA were refitted within each inner fit. Candidate losses and selected values are exported. PAD outer training sets contain only 11–12 subjects, so proper nesting prevents information sharing along these implemented tuning paths but cannot make parameter selection reliable in such small samples.

### Errors and uncertainty

MAE equally averages eight module errors, held-out subjects, and 20 repetitions. Positive improvement means comparator MAE minus method MAE. RMSE, module errors, and baseline/follow-up performance summaries are retained for inspection. Because gene scales vary by training split, aggregate errors are descriptive standardized-module quantities suited to matched method comparisons, rather than clinical endpoint scores.

For descriptive percentile resampling ranges, each subject’s out-of-fold (OOF) errors were averaged over modules and all 20 fitted repetitions before 5,000 subject bootstrap draws [24]. One weight followed each resampled subject across methods and repetitions. Modules, folds, and repeated predictions were not counted as independent observations. The ranges resample fixed observed subject-averaged losses while holding predictions, transforms, tuning and splits fixed; they omit model-refitting, training-set and selection uncertainty. Averaging within subjects does not remove dependence between their losses induced by shared training data. The nominal percentile level has no validated coverage guarantee, including conditional coverage given the fitted models, coverage for a fully refitted prediction procedure, or coverage in a new patient population. No universally unbiased K-fold variance estimator exists under all distributions [25]; that result does not validate these resampling ranges. Historical output fields named conditional_ci95_* are retained for traceability and denote these descriptive ranges, not calibrated confidence intervals. We separately report the central 95% range of complete-repeat MAEs as split variability, not an inferential confidence interval. Post hoc comparisons and module summaries receive no multiplicity-adjusted confirmatory interpretation.

### Known-truth simulation

A post hoc simulation examined when comparator choice rewards baseline subtraction, scalar persistence or cross-module dependence. The executed design is recorded in sim_protocol.json. Eight correlated Gaussian baseline states generated follow-up as F = A B + c + Gaussian noise under four scenarios: independent follow-up (A = 0), scalar persistence, off-diagonal state dependence, and simultaneous covariate/relationship transport shift. Complete coefficient matrices, covariance matrices, shifts and random seeds are supplied. Each scenario used 100 independent training/test datasets at training n = 72 and n = 300, with 2,000 independently generated test observations per dataset: 800 datasets in total.

Mean follow-up, median follow-up, mean change, persistence, scalar shrinkage and multivariate ridge were compared on every dataset. Scalar and ridge hyperparameters were selected by five-fold training-only MAE and refitted on the complete training set before test evaluation. All 48 scenario-by-size-by-method cells are reported. Monte Carlo standard errors (MCSEs) quantify simulation precision; they are distinct from patient confidence intervals and from the exported replicate-distribution quantiles. Full refitting in simulations does not establish coverage of the real-data resampling ranges. In the independent, unit-noise Gaussian scenario, population-oracle MAE is sqrt(2/pi) = 0.7979; finite-training expected MAE is sqrt(2/pi) × sqrt(1 + 1/n) for mean follow-up and sqrt(2/pi) × sqrt(2 + 2/n) for mean change. The historical test_population_oracle_MAE output field instead records the oracle’s empirical error on each finite generated test set; the statistical data dictionary distinguishes it from the analytic population expectation without renaming the saved column.

### Independent-source descriptions and external validity

No independently sampled longitudinal rosuvastatin cohort with the same paired PBMC outcome was included in this benchmark. GSE113873 is separately refitted, whereas GSE43292 and GSE27034 describe spatial and case/control contrasts. None is external validation of the statin prediction model. An independent longitudinal study-level holdout remains necessary.

GSE281841, a peripheral-artery series on GPL96 [12, 13], was added as a post hoc cross-sectional histological stenosis-grade description. The grouping refers to the histological grade of sampled arteries, not a validated patient-level clinical severity category. Grades 0–1 represent no stenosis or 1–25% stenosis, and grades 4–5 represent 76–100%; no grade 2 or 3 columns were present. SHA-256 checks preceded parsing. All 41 matrix columns and 28 literal patient identifiers were audited; source and title identifiers had to agree exactly, and differently prefixed forms were not merged without a source crosswalk. One unscored column was excluded, leaving 40 columns from 27 identifiers; two columns from one mixed-grade identifier were then excluded. The remaining 38 columns were averaged within literal identifier, yielding 18 identifiers contributing only high-grade and 8 contributing only low-grade retained arteries. Clustering assumes the recorded patient identity; label agreement does not independently confirm that all identifiers represent distinct participants. All 84 requested unique module genes mapped. The GEO summary reports 39 samples/27 patients; the 41-to-40-to-38 column and 28-to-27-to-26 identifier flow does not resolve that discrepancy.

The additional full-series gene z-scoring performed in this analysis was used only for descriptive scores; this restriction does not apply to inherited source preprocessing. Five thousand identifier-clustered bootstraps held normalization, mapping and modules fixed, without multiplicity adjustment and conditional on recorded patient identity. Exploratory restrictions compared grades within the same reported artery or title-derived prefix where each group had at least two recorded identifiers: peroneal (7 high/5 low), posterior tibial (6/3), prefix 0092 (5/4) and prefix 0521 (13/4). Prefixes are unverified technical proxies, not confirmed batches. Age and artery were available, but segment detail was limited and overlap sparse; no robust multivariable adjustment was possible or fitted. Lower-grade arteries belong to an end-stage PAD/amputation series, not healthy control participants.

### Coupling-preserving diagnostics and sensitivity analyses

Two hundred diagnostics independently re-paired follow-up vectors within each original outer training and test partition, retaining baselines in place. PAD re-pairings also remained within observed treatment groups. Recomputed targets were Δ_random = F_repaired − B, preserving the negative-baseline term while perturbing biological pairing where the strata permitted it. Training and test partitions never shared the same source subject within a fold. In the original PAD five-fold partition, at least 5/14 distinct test subjects necessarily retained their original follow-up pairing in every diagnostic because their test-fold treatment stratum contained only one person. Uniform permutations allowed additional fixed points in larger strata; this is partial re-pairing, not complete removal of individual matching.

These diagnostics use the fixed-penalty models and first five-fold partition. Different overlapping folds receive different re-pairings: the runs are split-conditional diagnostics, not globally coherent pseudo-cohorts or a calibrated exchangeability test. Their central ranges describe that diagnostic distribution. Exported plus-one fractions are not calibrated p values. The earlier 200 complete-CV permutations of already formed change targets are shown separately because they also destroy baseline–change coupling.

Regularization sensitivity evaluated every *λ* under raw and normalized features for change/direct targets in the first five repeats. These curves did not select the reported outer-test results. Training-size subsets used 12, 24, 36, 48, and all available statin training subjects, and 6, 8, 10, and all available PAD training subjects. Held-out subjects remained fixed; transformations and nested selection were refitted using each subset. Only one subset per fold/size was drawn, and gene-score scales changed with training size. These are descriptive sensitivities, not precise learning curves with independent intervals.

### Representation sensitivity, integrity checks and descriptive directions

Post hoc annotation sensitivity compared the frozen module-preferred mapping with retaining only probes containing one distinct parsed gene symbol before module filtering. The same first five repeated five-fold splits refitted ridge, scalar shrinkage, mean and componentwise-median follow-up controls. Probe aggregation, training-only gene normalization, module targets and inner folds were otherwise unchanged. Annotation changes both inputs and targets, so inference concerns improvement over a comparator within each scheme, not absolute MAE across schemes.

Two module sensitivities were kept separate. A fixed-prediction target-weight analysis removed one output module from the loss without refitting or deleting inputs. In contrast, actual input ablation removed each baseline module in turn while retaining all eight follow-up targets and fully retuning ridge in each fold: 400 outer fits across both cohorts and five repeats. Neither procedure selected a new primary model; correlated and overlapping modules preclude an inference of biological necessity.

The original integrity checks used the first fold of the first repetition in each cohort. Held-out follow-up expression was multiplied by 1,000 and increased by 10,000 with tuning fixed; clean predictions and training-baseline scalers should remain invariant. Six engineered faults per cohort tested all-visit scaling, duplicate subjects, sample reuse, cross-subject pairing, nonfinite training expression and train/test overlap. A stronger added check reran complete inner tuning after held-out follow-up perturbation in eight representative outer fits: folds 0 and 4 of repeat 0, both cohorts and both annotation schemes. Candidate losses, selected parameters and predictions were compared. Separate saved boundary checks perturbed inner-validation baseline or follow-up values in the first inner fold of outer fold 0/repeat 0 under the frozen mapping in each cohort. Training gene scalers, training module states and training feature matrices were expected to remain invariant. Validation baseline changes could alter validation inputs and predictions; validation follow-up changes could alter targets and losses but should leave fixed-parameter predictions unchanged. These additional checks did not rerun complete tuning or expand the eight representative checks. All perturbations start after the deposited processed matrices and do not audit source-study normalization. These finite checks test specified computational paths, not general verifier sensitivity or biological validity.

Descriptive directions used additional full-series gene z-scoring, separate from secondary predictive transforms: follow-up minus baseline for statin, exercise training and usual-activity controls; plaque minus macroscopically intact carotid tissue; and PAD minus control PBMC. The intact carotid comparator came from the same endarterectomy specimens and was described as Stary stage I–II, not healthy donor tissue. Exact action values separate five exercise-training from nine usual-activity control pairs. Five thousand draws resampled whole pairs within paired groups and subjects independently for PBMC, holding normalization and annotations fixed. Modules were not independently resampled. Pairwise directional concordance compared the two group-mean contrast vectors in the same eight-module order. For vectors d_a and d_b, the implementation computes (d_a · d_b)/(||d_a|| ||d_b|| + 10*−*12), both for the observed vectors and for each subject-bootstrap draw. The denominator offset prevents division by zero; an exactly zero vector returns zero as a computational convention, not evidence of biological concordance.

### Implementation and AI assistance

The supplied Python code uses NumPy, pandas, and Matplotlib. OpenAI Codex assisted with planning, code development and execution, statistical and consistency checks, figure generation, prose revision, and document assembly. Numerical results were generated by executing the supplied scripts; the extension records Python 3.12.14, NumPy 2.3.5, and pandas 3.0.1. Human review remains necessary. Figure 1 separates the implemented interfaces and evaluation boundaries; no neural architecture, recursive agent training, device simulation, or causal policy evaluation was executed.

**Figure 1:**
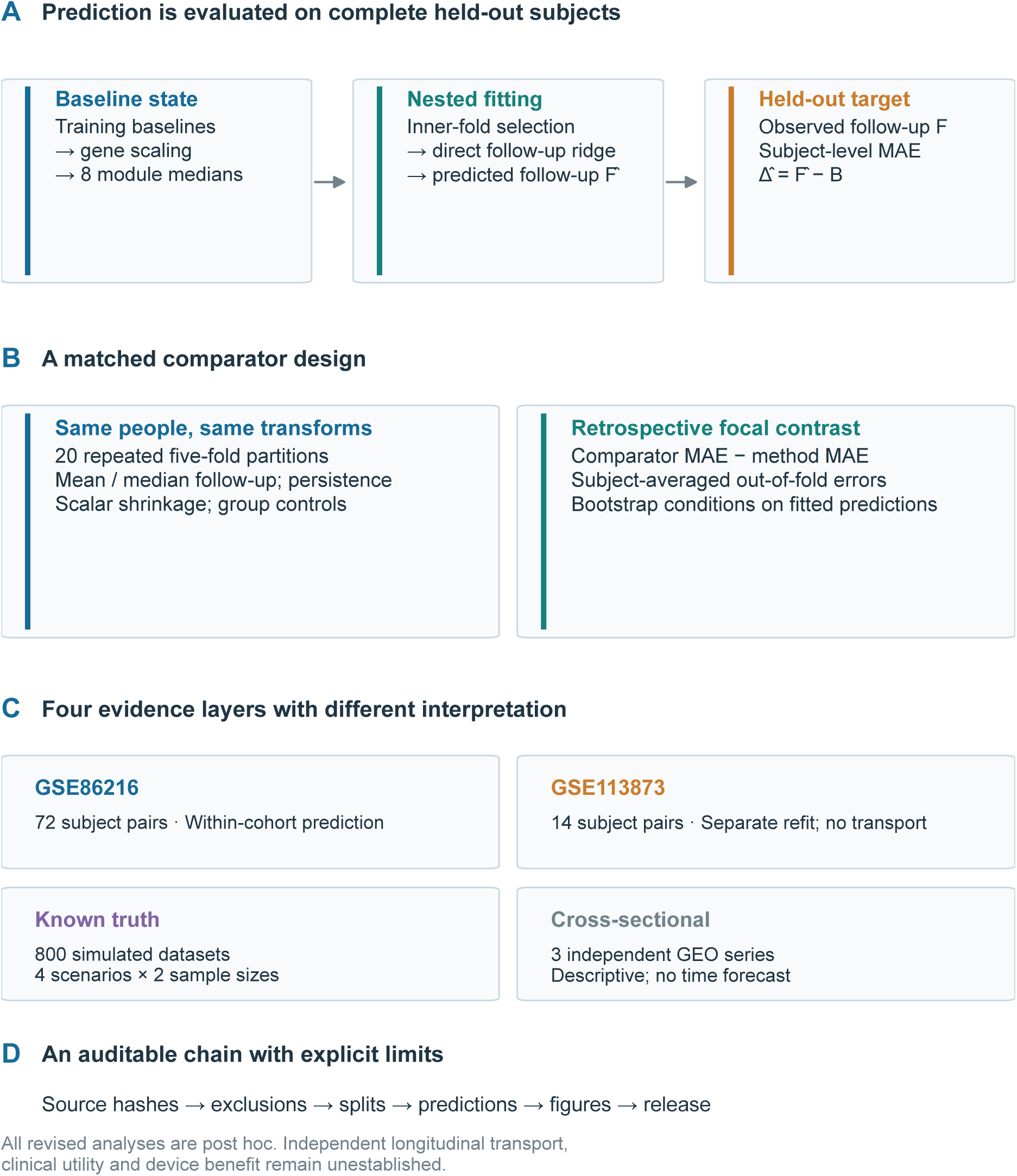
Implemented direct-follow-up prediction workflow and four evidence layers. The focal statin comparison is designated retrospectively; PAD is separately refitted. Simulation and cross-sectional descriptions do not validate longitudinal transport. The estimand is comparator MAE minus method MAE; positive values favor the method. Bootstrap ranges describe fixed observed OOF losses and have no validated coverage guarantee. All revised analyses are post hoc.

## RESULTS

### Data audit and reproducibility

The temporal cohorts retained 72 statin pairs and 14 PAD pairs; descriptive source cohorts retained 32 carotid pairs and 19 PAD/18 control PBMC participants. The added artery series contributed 26 literal patient identifiers after exclusions. Module coverage and exclusions are shown in Figure 2. The original statin module-gene union contained 83 available genes and PAD contained 84; overlapping modules are not independent biological tests.

**Figure 2:**
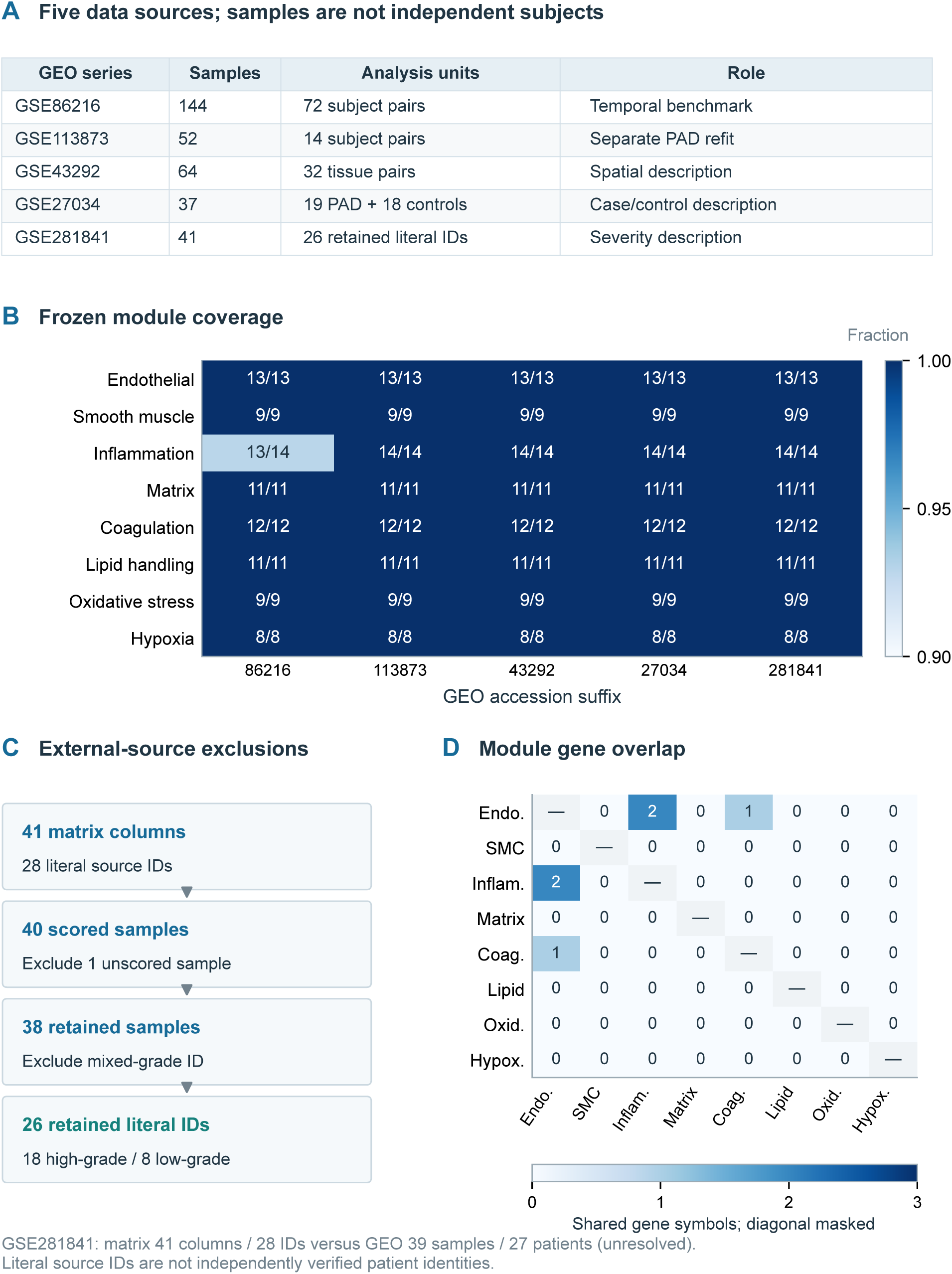
Five GEO sources, frozen module coverage, source exclusions and module-gene overlap. The GSE281841 matrix has 41 columns and 28 literal source IDs, versus 39 samples and 27 patients in the GEO summary; this discrepancy is unresolved. Excluding one unscored sample and two samples from one mixed-grade source ID leaves 38 samples and 26 literal IDs (18 high-grade, 8 low-grade). Source IDs are not independently confirmed patient identities. All 84 unique module genes map in GPL96; overlap diagonals are masked.

The original first partition reproduced 688 held-out subject-module predictions within 1.33 × 10*−*13 maximum discrepancy. Fixed change-ridge statin MAE was 0.327045 in that partition and 0.336481 over 20 partitions; the difference reflects aggregation, not failure to reproduce. All 200 cohort/repeat/fold reconstructions for the four added median controls passed identity, disjointness, finite-value and transformed-target checks.

The source-integrity audit found that the historical GPL570 cache stopped at 44,562 data rows without a platform-table end marker, whereas the complete official export contained 54,675 rows. Replacing it increased module-mapped probes from 214 to 229 and changed only GSE27034 descriptive scores and the cross-context cosines involving that source. Paired prediction, simulation, annotation/input-ablation and GSE281841 results were unchanged. The historical input and the correction impact are documented rather than treating the mismatch as header-only source drift.

After this correction, a reconstruction in a new output directory ran all six analysis stages and reproduced 48 corrected reference CSV tables with zero numerical difference. It used a newly created virtual environment on the same host and Python base runtime, with the eight freshly acquired complete input files and two freshly validated complete platform tables described above. Historical results were copied only as the frozen original-prediction comparison anchor; no current result directories were preloaded, and all current fits and six-stage outputs were regenerated. This verifies the supplied entry point against corrected inputs; it is not a test on an independent host, operating system or base runtime, nor a reconstruction from raw arrays. Current execution evidence is supplied in provenance/corrected_full_reconstruction_20261025.json; source-acquisition and GPL570 correction records distinguish complete files from complete platform-table validation.

### Matched comparators in both temporal cohorts

Table 2 retains all 11 original specifications. Across 20 statin repeats, fixed change ridge had lower observed MAE than mean change by 0.0497 (descriptive central 95% percentile range 0.0298–0.0702), but the observed difference from mean follow-up was 0.0040 (−0.0140 to 0.0222). The nested-change-ridge difference from mean follow-up was 0.0097 (−0.0091 to 0.0290). The designated focal nested direct-follow-up comparison yielded an observed difference of 0.0126 (0.0033–0.0224), with MAE 0.3280. Scalar shrinkage had lower observed mean MAE, 0.3256, and no ridge advantage over scalar shrinkage was established (Table 3). The reported ranges describe fixed-loss resampling and do not establish population superiority.

**Table 2:**
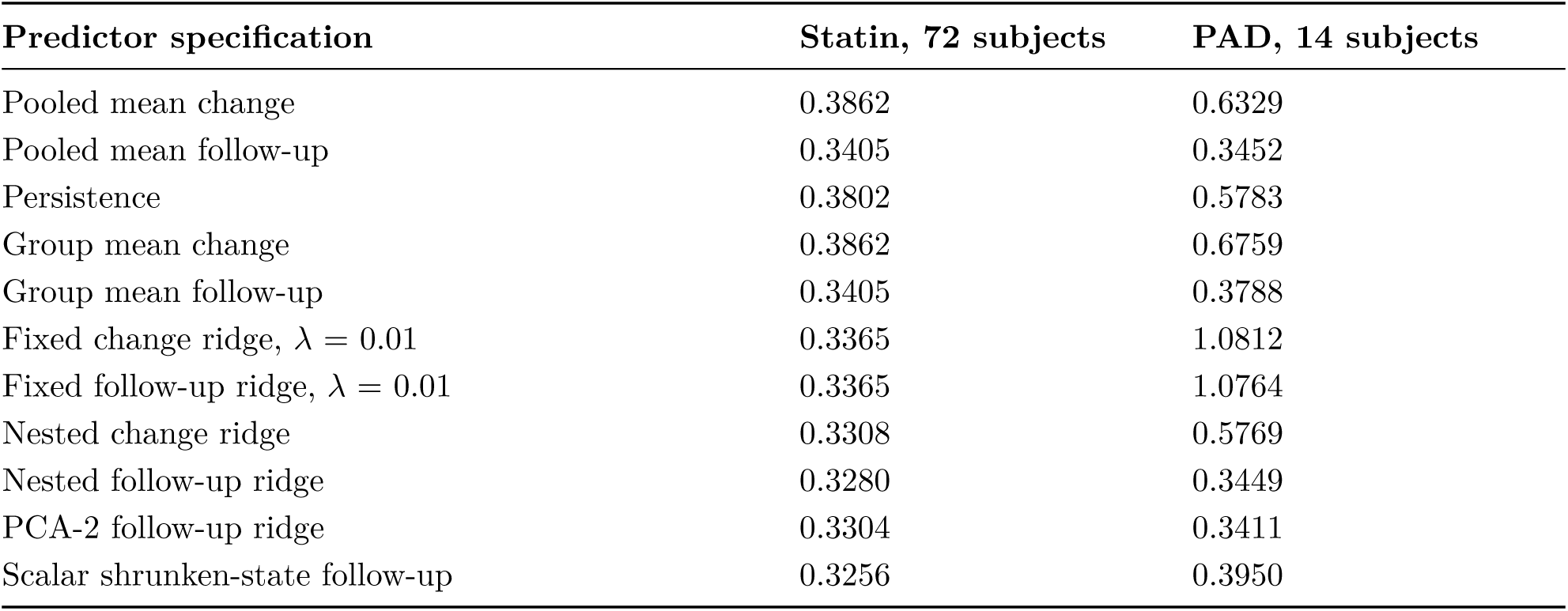
Mean subject-held-out MAE across 20 five-fold repetitions. Specifications within each cohort share held-out subjects and gene-score transformations; module-feature processing differs by method. Lower is better; units depend on training-baseline gene standardization. Absolute errors across these differently transformed tissues do not rank performance between cohorts.

**Table 3:** Added median controls and paired comparator analyses. Rows marked A give repeated MAE. Rows marked B give comparator MAE minus model MAE and descriptive central 95% percentile ranges from resampling fixed subject-averaged OOF losses; positive values favor the named model in these observations. The nominal level is not a validated coverage guarantee. All comparisons are post hoc, without full refitting, multiplicity or comparator-selection adjustment.

| Analysis | Statin, 72 subjects | PAD, 14 subjects |
| --- | --- | --- |
| A: Pooled median follow-up MAE | 0.3405 | 0.3409 |
| A: Group median follow-up MAE | 0.3405 | 0.3705 |
| A: Pooled median change MAE | 0.3861 | 0.5889 |
| A: Group median change MAE | 0.3861 | 0.6146 |
| B: Median follow-up minus nested follow-up ridge | 0.0125 (0.0021, 0.0232) | −0.0040 (−0.0194, 0.0080) |
| B: Median follow-up minus scalar shrinkage | 0.0149 (0.0033, 0.0268) | −0.0541 (−0.1787, 0.0149) |
| B: Scalar shrinkage minus nested follow-up ridge | −0.0024 (−0.0091, 0.0040) | 0.0501 (−0.0081, 0.1600) |

Group means duplicate pooled means in the single-regimen cohort. The median specifications are reported separately in Table 3; median follow-up had the lowest empirical MAE among the nine untuned mean, median and persistence controls in each cohort. Its small difference from mean follow-up in statin does not alter the focal descriptive ranking. The observed median-minus-ridge MAE difference was 0.0125 (0.0021–0.0232), while the median-minus-scalar difference was 0.0149 (0.0033–0.0268). These are outcome-informed exploratory comparisons with descriptive percentile ranges from fixed-loss resampling, without validated coverage.

PAD demonstrates a limitation rather than model transport (Figure 3). Fixed change ridge averaged MAE 1.0812; nested direct ridge had an observed MAE numerically close to mean follow-up (0.3449 versus 0.3452) and higher than median follow-up (0.3409). The observed median-minus-ridge difference was −0.0040 (−0.0194 to 0.0080). Scalar shrinkage and group-specific controls also had higher observed MAE than the pooled median. No advantage over the median was established; these comparisons do not establish equivalence or absence of predictive information. PAD direct ridge chose *λ* = 100 or 1000 in 99/100 outer fits; scalar shrinkage chose zero in 80/100. In statin, scalar tuning chose a = 0.25 in 73/100 fits and 0.5 in 27/100, while direct ridge selected *λ* = 100 in 84/100 and 10 in 16/100. These correlated fit counts describe strong shrinkage, not independent support for optimal parameters (Figure S4).

**Figure 3:**
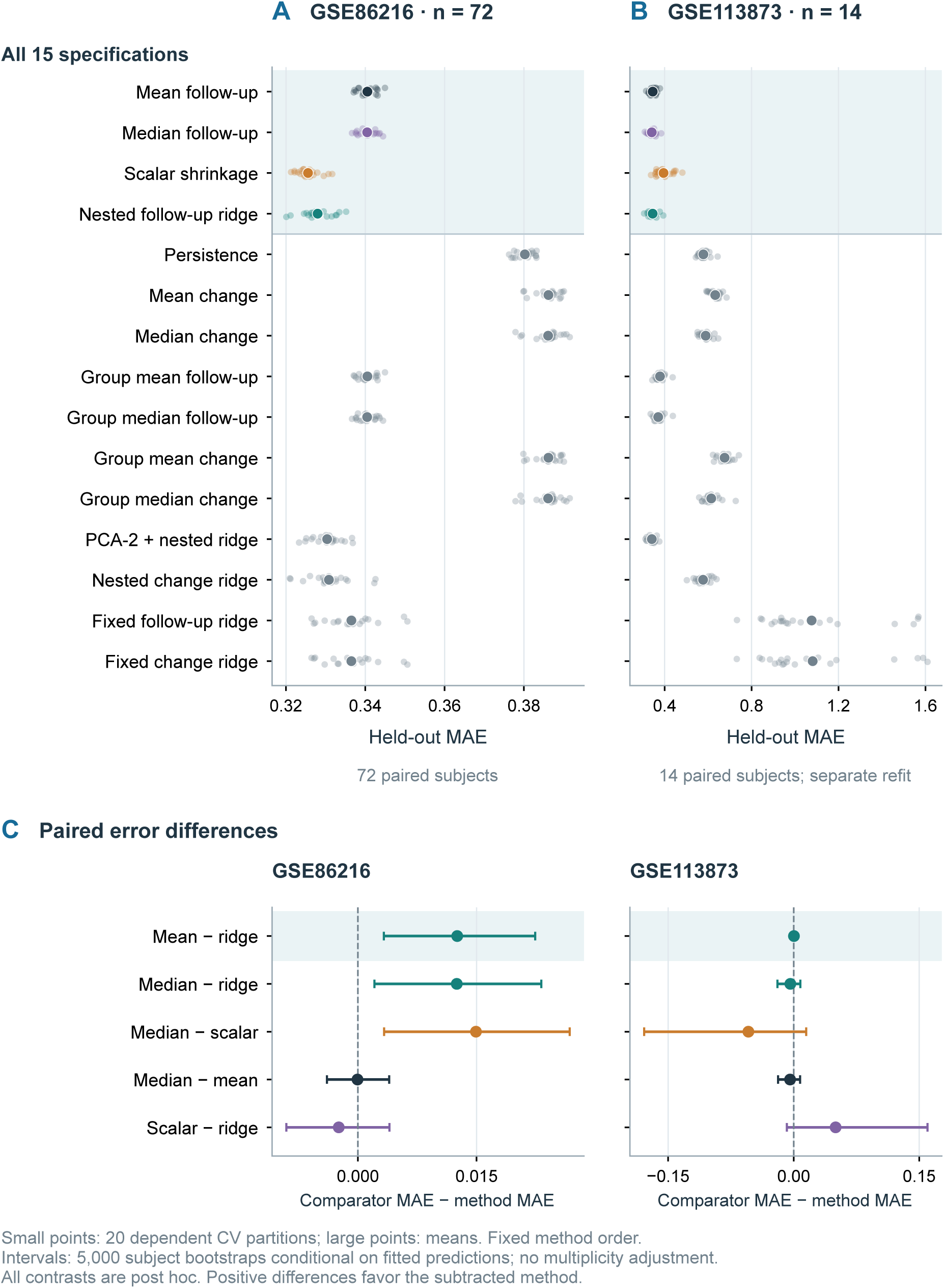
All 15 executed specifications and paired error differences in two temporal cohorts. Method order is fixed. Small dots are 20 dependent partitions; large dots are their mean. A and B use different x-axis ranges. C shows matched comparisons; shading marks the retrospectively designated mean-follow-up versus nested-ridge contrast. Descriptive percentile ranges use 5,000 subject bootstraps of fixed OOF losses after averaging errors across modules and repetitions, holding fitted predictions, transformations and tuning fixed. They omit refitting and multiplicity adjustment. Shared training data induce dependence between subject losses; the nominal percentile level has no validated coverage guarantee. Positive differences favor the subtracted method. PAD is separately refitted, not transported.

### Known-truth simulations explain comparator behavior

The 800 generated training/test datasets produced all 48 planned simulation cells (Figure 4). Without temporal signal at training n = 72, mean-follow-up MAE was 0.8033 (MCSE 0.0006), compared with 1.1359 (MCSE 0.0008) for mean change. At n = 300, mean and median follow-up achieved 0.7993 (MCSE 0.0005) and 0.8000 (MCSE 0.0005), close to the Gaussian population oracle of 0.7979. Thus a large advantage over mean change can occur without any temporal signal.

**Figure 4:**
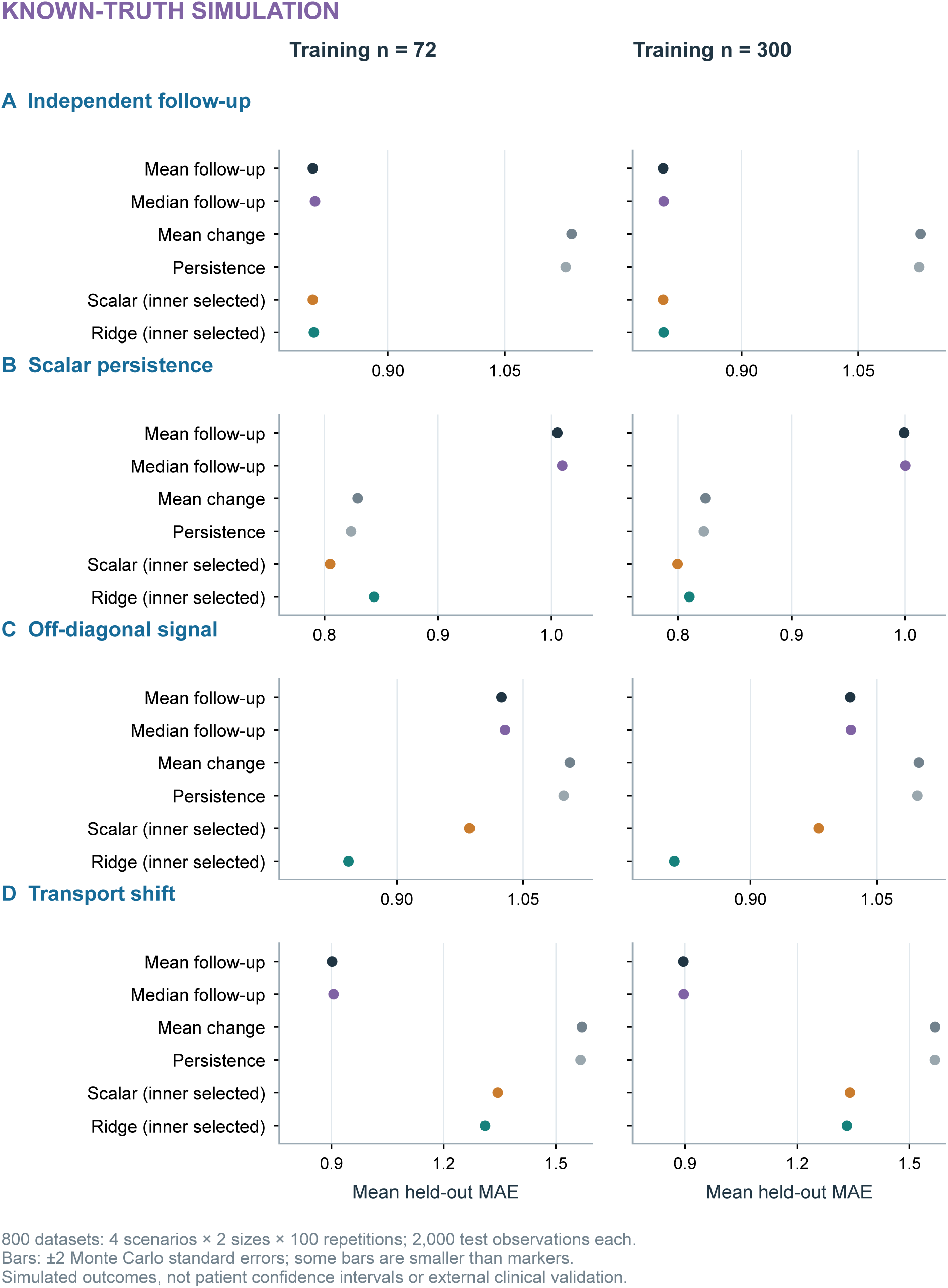
SIMULATION: four known-truth scenarios, each at training n = 72 and 300. Six methods share each generated training/test dataset; scalar and ridge tuning is confined to training data. Points average 100 independent repetitions, with bars of plus or minus 2 Monte Carlo standard errors, which can be smaller than markers. Each repetition has 2,000 test observations. There are 800 generated datasets, each evaluated by all six methods. The two subplots in each scenario share x limits. These post hoc demonstrations are not patient confidence intervals or evidence of human transport.

At n = 300 under scalar-persistence truth, scalar and ridge MAE were 0.7996 (MCSE 0.0005) and 0.8100 (MCSE 0.0005); under off-diagonal truth, they were 0.9811 (MCSE 0.0006) and 0.8097 (MCSE 0.0005). Full-matrix structure is useful in the latter constructed setting, but the real-data comparison does not establish that setting. Under the fixed transport shift, mean follow-up, scalar and ridge MAE were 0.8958 (MCSE 0.0011), 1.3413 (MCSE 0.0010) and 1.3332 (MCSE 0.0024), respectively. All n = 72 results and 40 paired contrasts are retained. Analytic null expectations and the follow-up/change residual identity passed numerical checks. These four illustrative scenarios test comparator behavior, not vascular biology or exhaustive transport robustness.

### Coupling, representation and integrity diagnostics

In the original statin partition, fixed-ridge improvement over mean change was 0.0609. Coupling-preserving re-pairing gave mean improvement 0.1013, with diagnostic central 95% range 0.0712–0.1262. Apparent benefit therefore persisted after subject matching was disrupted. Against mean follow-up, observed improvement was 0.0130, whereas re-pairing yielded −0.0283 (−0.0454 to −0.0160). The earlier change-target permutation gave mean MAE 0.418 versus observed 0.327, but removed baseline coupling as well as subject pairing. These fixed-model diagnostics do not test a calibrated null for the nested focal procedure (Figure 5; Figure S2).

The annotation sensitivity was an identity control in statin: both rules retained 141 probes and 83/84 module-union genes, producing identical predictions. In PAD, unique-symbol filtering reduced 290 probes/84 genes to 282 probes/83 genes and oxidative-stress coverage from 9 to 8 genes. Across five repeats, the observed mean-follow-up-minus-ridge MAE difference was 0.001640 under the frozen rule and 0.002895 under unique-symbol mapping; both descriptive percentile ranges included zero. Ridge had higher observed MAE than the corresponding median baseline (differences −0.001966 and −0.002417). Annotation robustness in statin therefore does not demonstrate insensitivity to a changed representation.

Removing one target module from fixed-prediction loss left statin ridge improvements over mean follow-up between 0.008642 and 0.017487. This reweighting is distinct from input deletion. The 400 fully retuned input-ablation fits (Figure 7) produced 27,520 target predictions; full-input MAE minus ablated-input MAE ranged from −0.004018 to −0.000866 in statin and −0.002898 to 0.004701 in PAD. No ablated input set was selected as a new primary model. Original penalty and training-size sensitivities remain descriptive (Figure 8): for example, fixed change-ridge statin MAE fell from 0.6003 with 12 training subjects to 0.3414 with 48, versus 0.3624 to 0.3285 for nested direct ridge.

**Figure 5:**
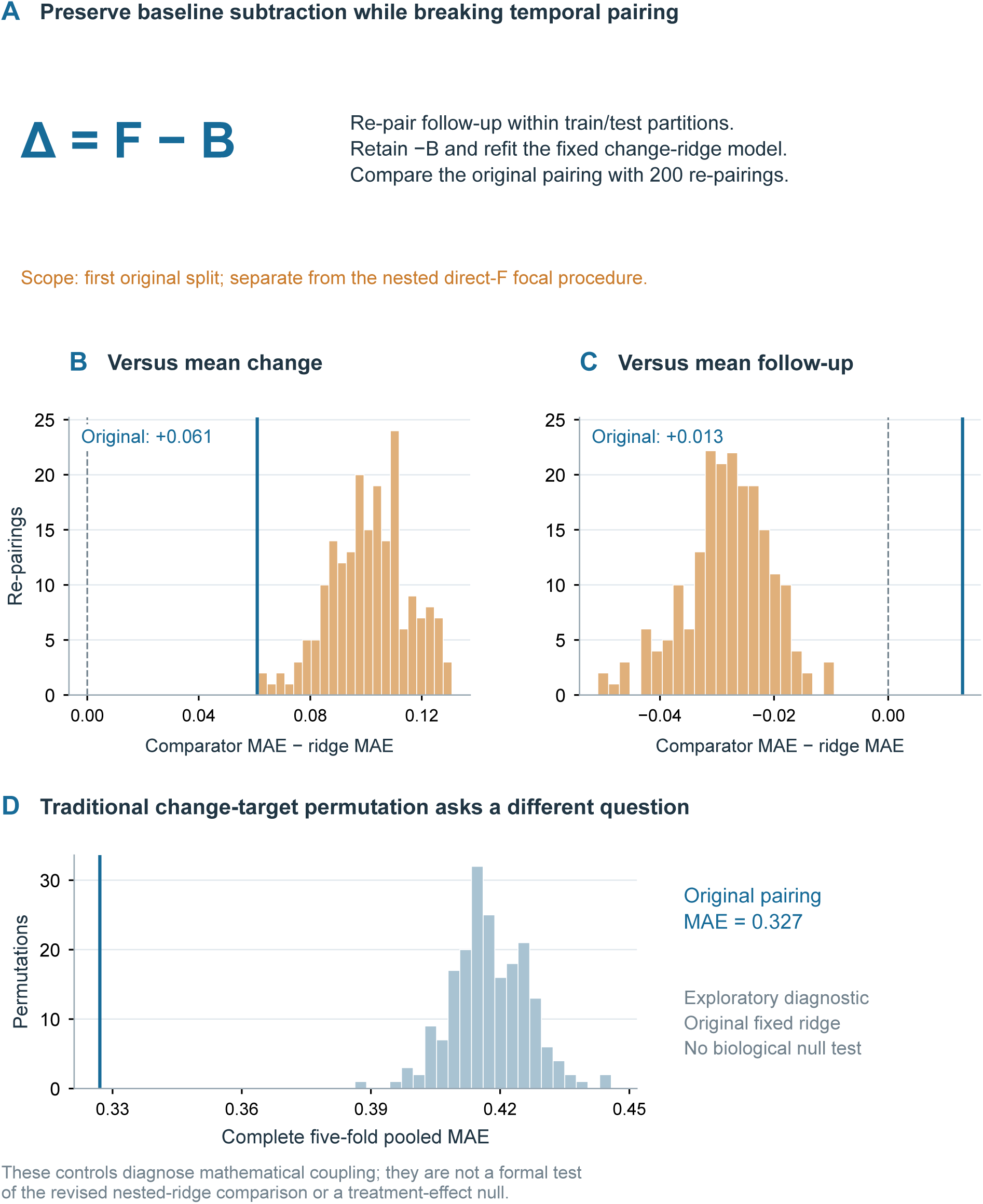
Fixed-change-ridge coupling diagnostics using the first original five-fold partition. Follow-up is re-paired within train and test partitions 200 times while retaining baseline subtraction. B and C compare the observed pairing with these re-pairings using different comparators. D permutes the change target and answers a different question. These diagnostics do not test a calibrated null for the nested direct-follow-up focal procedure or a biological treatment-effect null.

**Figure 6:**
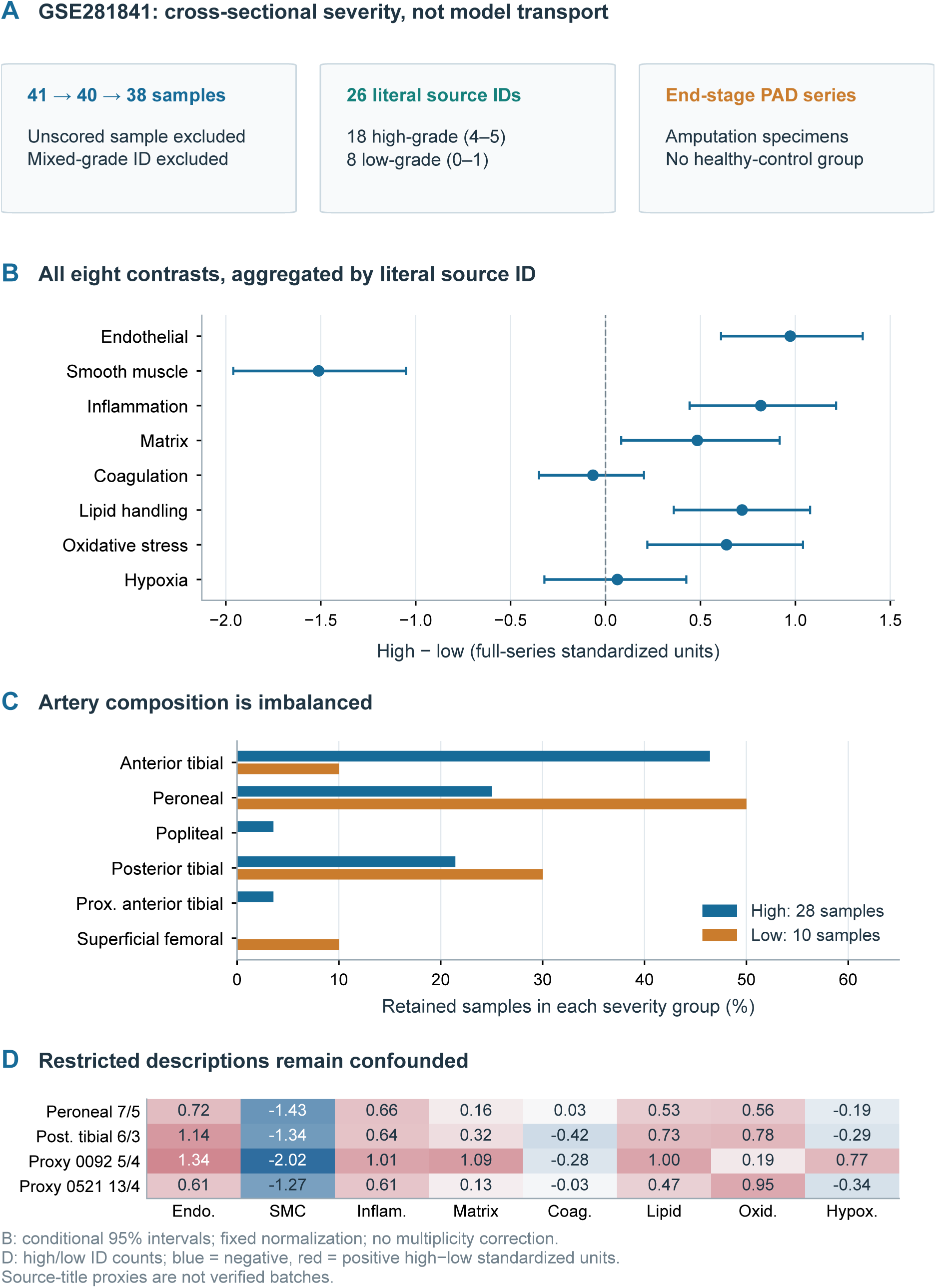
Post hoc GSE281841 cross-sectional severity description. B uses 18 high-grade and 8 low-grade literal source IDs, averaging retained samples within each ID; descriptive percentile ranges use 5,000 identifier-clustered bootstrap draws with normalization, mapping and modules fixed. Their nominal level has no validated population coverage guarantee. C describes 28 high-grade and 10 low-grade samples, not independent people. D shows all eight modules in four restriction rows, each with at least two IDs per grade; labels give high/low ID counts. Values are high minus low in full-series standardized units, on a symmetric blue (negative) to red (positive) scale. Full unadjusted descriptive ranges are in the source CSV. Anatomical imbalance, unverified identity and title-prefix proxies, and residual confounding prevent longitudinal, causal or device claims.

**Figure 7:**
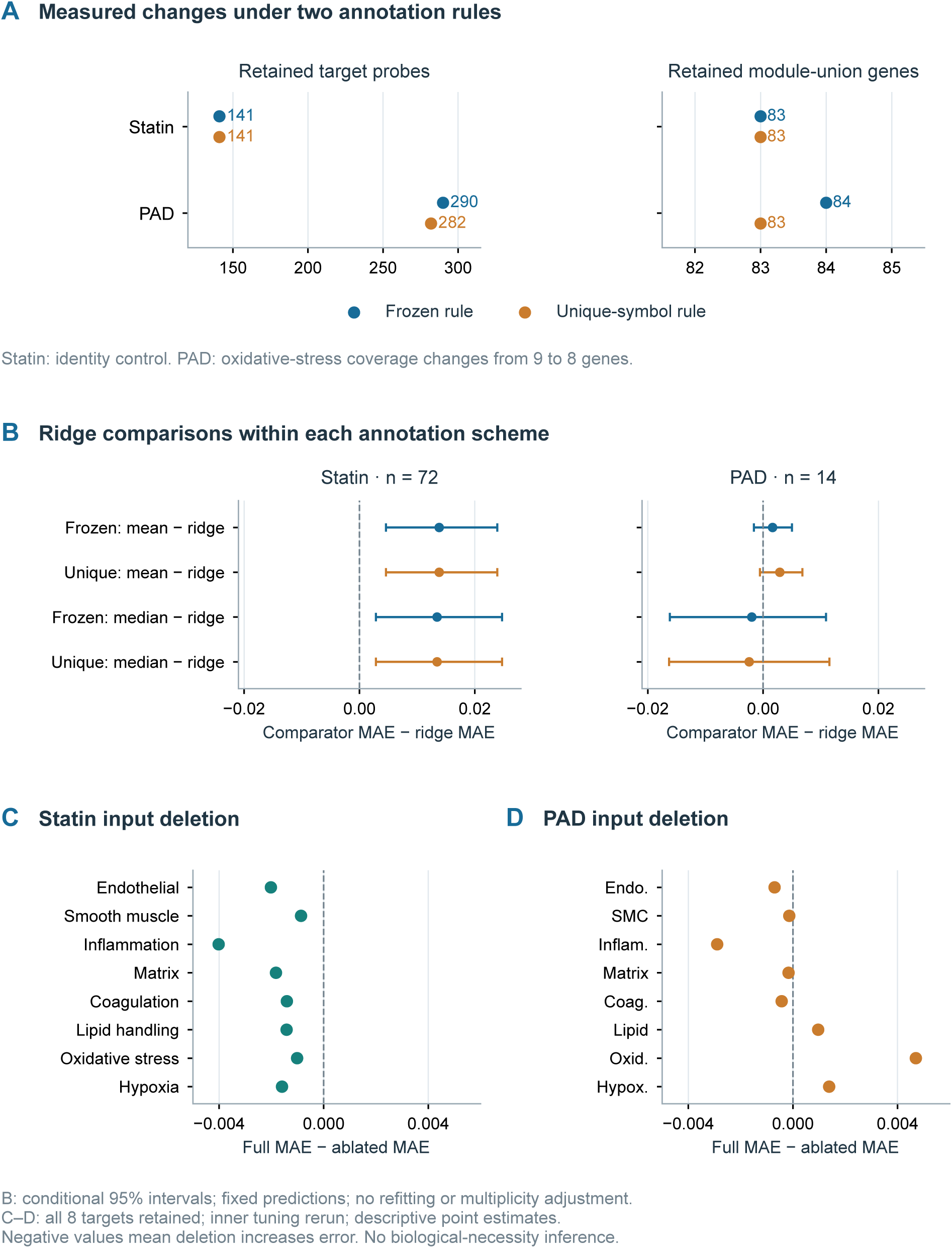
Post hoc representation and input sensitivities using five repeated five-fold partitions. A quantifies annotation changes: statin is an identity control (141 probes and 83 genes under both rules); PAD changes from 290/84 to 282/83, with oxidative-stress coverage 9 to 8. B compares nested ridge with mean and median follow-up within each annotation scheme; mapping changes the targets, so absolute errors across schemes are not directly compared. Descriptive percentile ranges use 5,000 subject bootstrap draws of fixed OOF losses, without refitting or multiplicity adjustment. The nominal percentile level has no validated coverage guarantee; shared training data induce dependence between subject losses. C and D delete each input module, retain all eight targets and rerun inner tuning. Negative full-minus-ablated MAE means deletion increases error. Ablation points have no inferential intervals; overlapping modules preclude biological-necessity claims.

**Figure 8:**
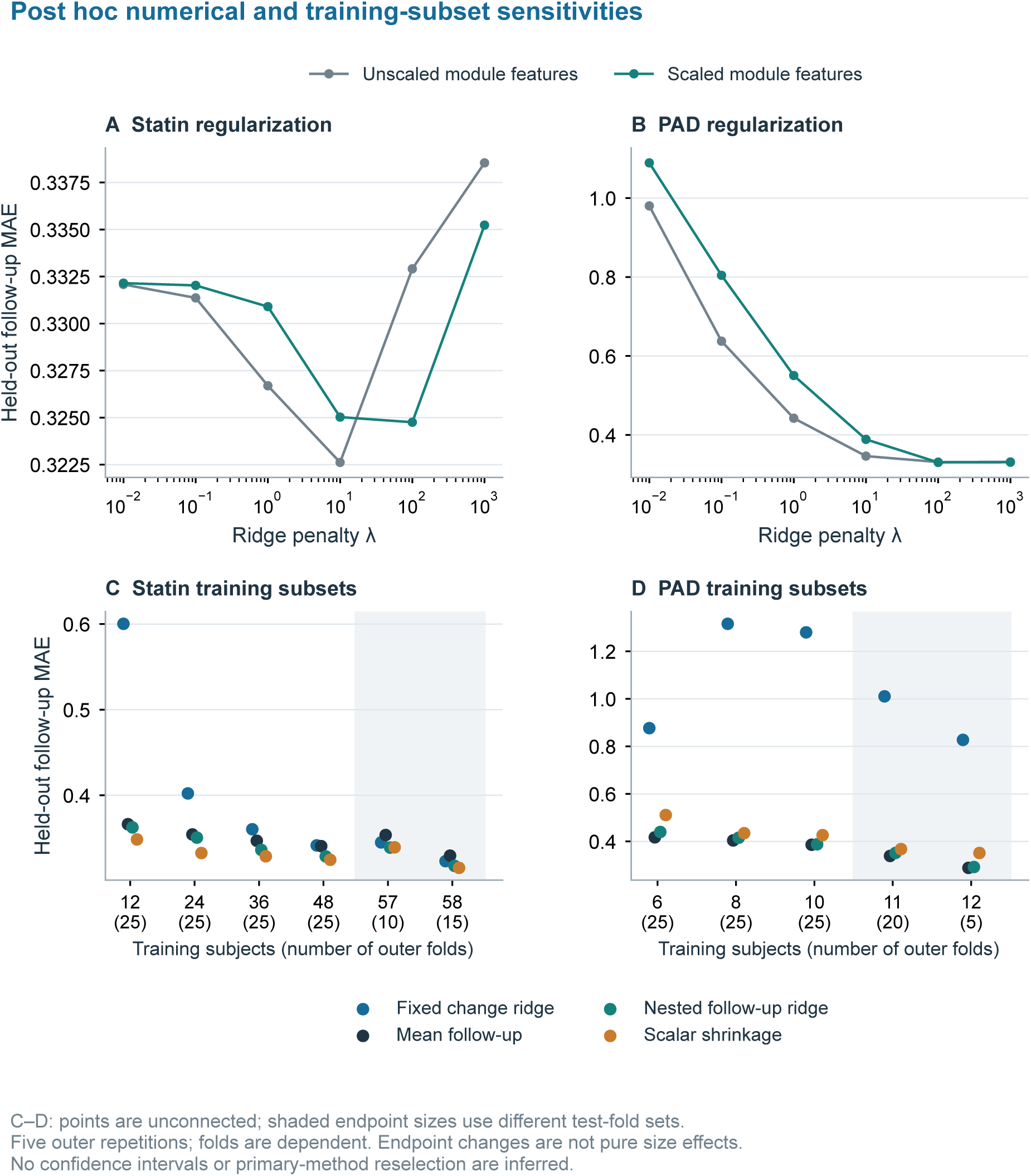
Original post hoc sensitivity analyses. A and B compare fixed follow-up-ridge penalties with and without additional module-feature scaling; gene preprocessing remains within training folds. C and D show descriptive held-out errors across training subsets and methods, using five outer repetitions. Points are deliberately unconnected: statin sizes 57/58 use different sets of 10/15 folds, and PAD sizes 11/12 use different sets of 20/5 folds, whereas smaller sizes use all 25 folds. Their test-subject composition is not common, so endpoint differences are not pure training-size effects. Repeated folds are dependent; no confidence interval or primary-method reselection is inferred.

The original two clean fixed-tuning mutation checks remained invariant and all 12 injected faults were detected (Figure S6). In the stronger eight representative checks, complete inner tuning was rerun for all four originally tuned families. All 184 candidate-loss comparisons, selected parameters and predictions from the 11 original specifications were unchanged within 10*−*12 while held-out targets changed. This tests future-target use along those exercised paths, not every possible leak. The frozen annotation reproduced 10,320 original five-repeat OOF module predictions within 4.44 × 10*−*16, and all newly fitted annotation predictions were uniquely keyed and finite.

The additional inner-validation checks found zero change in the tested training transformations for both perturbation types in each cohort. Follow-up perturbations changed validation targets while leaving the 11 fixed-parameter predictions unchanged; baseline perturbations changed validation inputs and some predictions, as permitted by the check. The saved script also reconstructed the original PAD folds and confirmed the 5/14 singleton constraint; replayed permutation-assignment counts agreed with the archived diagnostics. These checks cover only the specified secondary-analysis paths.

### Independent-source histological stenosis-grade and cross-context descriptions

In GSE281841, high-minus-low histological-grade contrasts were positive for the endothelial-activation, inflammation, extracellular-matrix, lipid-handling and oxidative-stress module scores, and negative for the contractile-gene module score (Figure 6). Six descriptive percentile ranges excluded zero: endothelial-activation module score 0.973 (0.609–1.355), contractile-gene module score −1.511 (−1.960 to −1.051), inflammation module score 0.818 (0.442–1.215), extracellular-matrix module score 0.484 (0.083–0.917), lipid-handling module score 0.719 (0.359–1.078), and oxidative-stress module score 0.637 (0.220–1.041). The coagulation/thrombosis module score was −0.066 (−0.350 to 0.203), and hypoxia/glycolysis module score 0.063 (−0.322 to 0.426). These are unadjusted histological-grade descriptions conditional on recorded patient identity, not follow-up prediction validation or calibrated population confidence statements.

Mean recorded age across retained identifiers was 64.67 (SD 10.17) years in the high-grade group and 61.63 (SD 12.50) in the low-grade group. Artery composition was imbalanced, including 13 high/1 low anterior-tibial samples; retained title-prefix sample counts were 5/5 for 0092 and 23/5 for 0521. The endothelial-activation, contractile-gene, inflammation and lipid-handling module scores retained their respective contrast signs in both estimable artery strata and both prefix strata. Other directions were unstable: hypoxia/glycolysis changed from +0.768 in prefix 0092 to −0.344 in 0521; coagulation/thrombosis changed from +0.025 in peroneal to −0.420 in posterior-tibial arteries. The 32 restricted module contrasts are exploratory and correlated. Anatomical and technical composition, cell mixture and imperfect identity information remain plausible explanations; lower-grade arteries are not healthy controls.

The other cross-context descriptions were also uncertain. Exact action matching had corrected an earlier substring filter that combined all 14 exercise-training/usual-activity pairs; the subsequent GPL570 completeness correction further changed PBMC-based contrasts. With both corrections, the exercise-training/PAD PBMC cosine was 0.191 (−0.265 to 0.620). Statin/carotid, statin/PAD PBMC and carotid/PAD PBMC cosines were 0.741 (−0.113 to 0.874), 0.555 (−0.323 to 0.803) and 0.503 (−0.079 to 0.746). All ten cross-group descriptive percentile ranges included zero (Figures S5 and S7); these observations do not establish a universal response or device suitability.

## DISCUSSION

The empirical result is that comparator choice changes the assessment of prediction in these cohorts. In statin, nested direct ridge had modestly lower observed repeated-CV MAE than mean and median follow-up. Its observed error was higher than scalar shrinkage, and no ridge advantage over scalar shrinkage was established. This finding does not establish equivalence or prove that cross-module structure is absent. PAD ridge had an observed MAE numerically close to mean follow-up and higher than the median; no advantage over the median was established. The distinction between lower observed error than a constant control and additional value from a full matrix is central: the former cannot establish the latter. These results apply established statistical tools to a specific empirical question; they do not establish a new prediction algorithm.

The independent-follow-up simulation checks an analytically expected comparator effect: mean-change prediction adds baseline variability and can lose to constant follow-up prediction without temporal signal. Other constructed scenarios illustrate when scalar or cross-module structure can help and when transport can fail. Their data-generating mechanisms cannot be inferred from the vascular cohorts. In the real-data fixed-model diagnostic, improvement over mean change survived disruption of biological matching while baseline subtraction remained. This supplies a counterexample to interpreting that advantage alone as temporal information; it does not show that the real data contain no such information. These analyses identify insufficiently supported interpretations without partitioning observed prediction into mathematical, technical and biological components. The statin series has one regimen and no untreated counterfactual, so response mechanisms and treatment effects remain unidentified.

A second contribution is the inspectable implementation of this benchmark: subject and split manifests, fitted-prediction records, candidate tuning losses, scoped fault tests and source-to-result checks. Complete-tuning perturbation checks strengthen the fixed-tuning checks, and reconstruction from a new output directory examines the analysis entry point. Their value is traceability of this analysis; the exercised paths do not constitute a generally complete leakage detector. Computational reproducibility and independent replication are distinct evidentiary properties [26, 27]. These checks do not establish general source-annotation correctness, cross-platform transport, reproduction on an independent host or base runtime, or biological validity.

Representation uncertainty remains consequential. Statin’s alternative annotation rule did not change its representation, whereas PAD lost one module gene and retained no demonstrated advantage over median follow-up. Input ablation and target reweighting answer different questions, and overlap between curated modules prevents claims about indispensable pathways. Empirical-Bayes batch adjustment and surrogate-variable analysis address technical heterogeneity [28, 29], but neither was fitted here; cross-platform and latent technical variation remain possible explanations of descriptive contrasts. PAD’s 11–12 outer-training subjects, noise and tissue differences are not resolved by correct nesting. The representativeness of its complete-pair subset relative to the source PAD population is unknown. Its role is to expose a limitation of the fixed recipe, not prove cross-tissue generalization. Source matrices inherited upstream preprocessing that may use information across samples; the training-only checks cover the secondary analysis and do not establish an end-to-end single-future-sample deployment workflow. Evaluation is conditional on the frozen processed matrices, without demonstrating or ruling out upstream information sharing.

The independent artery series provides descriptive context from another source and answers a cross-sectional question. Several histological-grade module-score directions persisted within sparse anatomical and prefix strata, whereas others changed sign. Low-grade arteries from patients with end-stage PAD cannot substitute for healthy controls; the unresolved source sample count, unconfirmed distinct-person identities, artery composition and cell mixture further limit interpretation. Neither these contrasts nor the uncertain cross-context cosines validate baseline-to-follow-up prediction. A decisive next step requires an independently collected paired cohort, a comparator and acceptable gain specified before outcome access, and uncertainty from repeated full fitting. Clinical translation additionally requires treatment timing, anatomy, haemodynamics, device variables and clinical outcomes absent here.

All extensions are post hoc and nonconfirmatory. Descriptive percentile ranges from fixed subject-averaged OOF losses omit model-refitting, representation and selection uncertainty; their nominal level has no validated coverage guarantee. Correlated repeated folds do not create additional subjects, and averaging within subjects does not remove dependence induced by shared training data. Simulations refit models on independent synthetic datasets but cannot supply missing patient evidence or an exhaustive robustness assessment. Reproducing outputs from the same inputs and seeds adds no patients or training-set sampling uncertainty. The code package is prepared for review but has not yet been deposited with a persistent public identifier. A publicly archived release remains necessary for findability and reuse in line with FAIR principles [30] and open research practice [31]. The present evidence supports an inspectable comparator benchmark with modest observed cohort-specific error differences; it does not establish clinical usefulness or independent longitudinal validity.

## CONCLUSIONS

VascFlux-GEO demonstrates how comparator choice changes the interpretation of vascular transcriptome module prediction. Nested follow-up ridge had modestly lower observed repeated-CV MAE than constant-follow-up controls in one statin cohort, with no advantage over scalar shrinkage established; PAD ridge had slightly higher observed MAE than the median, with no advantage over that comparator established. These exploratory comparisons do not establish equivalence or population superiority. Known-truth simulations, representation sensitivities and executable checks clarify these limits. Independent longitudinal validation, treatment mechanisms, clinical utility and device suitability remain unestablished.

## RESOURCE AVAILABILITY

### Lead contact

Questions about the computational package should be directed to Junbo Ge.

### Materials availability

This study is computational and generated no new reagents or materials.

### Data and code availability

GEO source matrices and platform annotations are public through the accession links in Table 1 and References. The private author-review package contains executable scripts, frozen module definitions, source endpoints and hashes, exact subject folds, tuning losses, held-out predictions, simulation protocols/results, annotation and input-ablation outputs, descriptive percentile ranges, cross-sectional restrictions and fault evidence. The submission code-and-aggregate attachment omits source patient identifiers, sample-level linked records, and individual ages; those retained audit records belong only to the private package. Eight main and seven supplementary figures summarize these analyses. The supplied six-stage entry point was rerun in a new output directory and a newly created virtual environment on the same host and Python base runtime after correcting the incomplete GPL570 cache. It reproduced 48 corrected CSV tables with zero numerical difference using eight freshly acquired complete input files and two freshly validated complete platform tables; the source acquisition, correction and reconstruction scopes are documented in the provenance records. No public code repository, archived release DOI or unrestricted download URL has yet been created, and the local review package should not be described as a public release.

## ETHICS AND CONSENT

This secondary analysis used publicly deposited processed gene-expression matrices and accompanying study metadata. No new participants were recruited, no directly identifying personal data were accessed, and no contact with source-study participants occurred. Recruitment, consent, and ethics oversight for the original biospecimen collections belong to the source studies; this manuscript makes no claim of a new institutional ethics approval or exemption.

## AUTHOR CONTRIBUTIONS

Lingsen You: Conceptualization, Methodology, Writing - original draft. Zisu Peng: Data curation, Visualization, Validation, Writing - review & editing. Yujun Guo: Methodology, Software, Validation, Writing - review & editing. Li Shen: Supervision, Validation, Writing - review & editing. Junbo Ge: Conceptualization, Supervision, Project administration, Writing - review & editing. Lingsen You, Zisu Peng, and Yujun Guo contributed equally to this work.

## DECLARATION OF GENERATIVE AI AND AI-ASSISTED TECHNOLOGIES

Codex assisted with LaTeX assembly, prose editing, and consistency checks. The authors reviewed and revised all AI-assisted text and take responsibility for the submitted manuscript.

## Supporting information

Analysis code and aggregate results

## Data Availability

Source processed matrices and platform annotations are publicly accessible through NCBI GEO accessions GSE86216, GSE113873, GSE43292, GSE27034 and GSE281841. Analysis code and aggregate results are included in the supplementary code-and-aggregate attachment. No separate public code repository or archive DOI has been created.

https://www.ncbi.nlm.nih.gov/geo/query/acc.cgi?acc=GSE86216

https://www.ncbi.nlm.nih.gov/geo/query/acc.cgi?acc=GSE113873

https://www.ncbi.nlm.nih.gov/geo/query/acc.cgi?acc=GSE43292

https://www.ncbi.nlm.nih.gov/geo/query/acc.cgi?acc=GSE27034

https://www.ncbi.nlm.nih.gov/geo/query/acc.cgi?acc=GSE281841

## Supplementary files

The supplementary PDF contains aggregate methods, tables and Figures S1–S7. The code-and-aggregate submission attachment is a screened subset of the private author-review package. The directory inventory below describes that full private package; per-person predictions, fold assignments, linked source records and ages are not submission attachments.

- data_sources.tsv, inputs/ and provenance/: source accessions, acquisition endpoints, hashes, module definitions and reconstruction evidence.
- results/: original numerical artifacts and first-partition reproduction anchor.
- results_extended/: original 11-specification predictions, folds, tuning, conditional intervals, re-pairing, sensitivity and descriptive results.
- results_reviewer_revision/fair_* and summary_fair_comparators.json: four added median controls and paired comparisons, preserving original specifications.
- sim_protocol.json and results_reviewer_revision/simulation_*: results from all 800 known-truth datasets, 48-cell summaries, 40 paired contrasts and analytic checks.
- results_reviewer_revision/robustness_*: annotation comparisons, fixed-prediction target weighting, fully retuned input ablation and eight complete-tuning mutation checks.
- audit_inner_validation_boundaries.py and provenance/inner_validation_audit/: scoped inner-validation boundary checks and PAD test-stratum/fixed-pair accounting after source preprocessing.
- results_extended/external_GSE281841_*: all 41 source columns, exact exclusions, patient scores, module coverage, confounding audit and 32 restricted cross-sectional contrasts. The historical filename direction_holdout.csv does not indicate longitudinal model validation.
- Figures S1 and S3: first-partition statin and PAD prediction diagnostics; S2: outcome identities; S4: tuning; S5: descriptive directions; S6: original fixed-tuning fault controls; S7: cross-context directions and all ten cosine ranges. Original regularization and training-size sensitivities are in main Figure 8.

## References

[1] Vickers AJ, Altman DG. Analysing controlled trials with baseline and follow up measurements. BMJ. 2001;323:1123–1124. doi:10.1136/bmj.323.7321.1123. https://www.bmj.com/content/323/ 7321/1123

[2] Hoerl AE, Kennard RW. Ridge Regression: Biased Estimation for Nonorthogonal Problems. Technometrics. 1970;12(1):55–67. doi:10.1080/00401706.1970.10488634. https://www.tandfonline.com/doi/abs/10.1080/00401706.1970.10488634

[3] Varma S, Simon R. Bias in error estimation when using cross-validation for model selection. BMC Bioinformatics. 2006;7:91. doi:10.1186/1471-2105-7-91. https://link.springer.com/article/10.1186/1471-2105-7-91

[4] Tzima E, Irani-Tehrani M, Kiosses WB, et al. A mechanosensory complex that mediates the endothelial cell response to fluid shear stress. Nature. 2005;437:426–431. doi:10.1038/nature03952. https://www.nature.com/articles/nature03952

[5] You L, Li H, Lu Z, et al. Intra-aortic balloon pump use and in-hospital mortality in acute myocardial infarction with Killip class III or IV: a nationwide cohort study. Biomarker Research. 2026;14:97. doi:10.1186/s40364-026-00993-1. https://link.springer.com/article/10.1186/ s40364-026-00993-1

[6] Clough E, Barrett T. The Gene Expression Omnibus Database. Methods in Molecular Biology. 2016;1418:93–110. doi:10.1007/978-1-4939-3578-9_5. https://link.springer.com/protocol/1 0.1007/978-1-4939-3578-9_5

[7] Edgar R, Domrachev M, Lash AE. Gene Expression Omnibus: NCBI gene expression and hybridization array data repository. Nucleic Acids Res. 2002;30(1):207–210. doi:10.1093/nar/30.1.207.

[8] National Center for Biotechnology Information. Gene Expression Omnibus dataset GSE86216. https://www.ncbi.nlm.nih.gov/geo/query/acc.cgi?acc=GSE86216 (accessed 9 October 2026).

[9] National Center for Biotechnology Information. Gene Expression Omnibus dataset GSE113873. https://www.ncbi.nlm.nih.gov/geo/query/acc.cgi?acc=GSE113873 (accessed 9 October 2026).

[10] National Center for Biotechnology Information. Gene Expression Omnibus dataset GSE43292. https://www.ncbi.nlm.nih.gov/geo/query/acc.cgi?acc=GSE43292 (accessed 9 October 2026).

[11] National Center for Biotechnology Information. Gene Expression Omnibus dataset GSE27034. https://www.ncbi.nlm.nih.gov/geo/query/acc.cgi?acc=GSE27034 (accessed 9 October 2026).

[12] NCBI Gene Expression Omnibus. Gene expression data from patients with peripheral arterial disease stenosis grade 1 versus 4 and 5. Series GSE281841, GPL96. https://www.ncbi.nlm.nih.gov/geo/query/acc.cgi?acc=GSE281841.

[13] Zhao F, Li F, Sedaghati F, et al. Piezo-type mechanosensitive ion channel component 1 (PIEZO1) is upregulated in peripheral arterial disease (PAD) and a novel murine PAD model. JVS Vasc Sci. 2025;6:100394. doi:10.1016/j.jvssci.2025.100394.

[14] Kini AS, Vengrenyuk Y, Shameer K, et al. Intracoronary Imaging, Cholesterol Efflux, and Transcriptomes After Intensive Statin Treatment: The YELLOW II Study. J Am Coll Cardiol. 2017;69(6):628–640. doi:10.1016/j.jacc.2016.10.029. Source series: GSE86216.

[15] Parmer C, De Sousa-Coelho AL, Cheng HS, et al. Skeletal muscle expression of adipose-specific phospholipase in peripheral artery disease. Vasc Med. 2020;25(5):401–410. doi:10.1177/1358863X20947467. 10.1177/1358863X20947467. Source series: GSE113873.

[16] Ayari H, Bricca G. Identification of two genes potentially associated in iron-heme homeostasis in human carotid plaque using microarray analysis. J Biosci. 2013;38(2):311–315. doi:10.1007/s12038-013-9310-2. 10.1007/s12038-013-9310-2. Source series: GSE43292.

[17] Masud R, Shameer K, Dhar A, Ding K, Kullo IJ. Gene expression profiling of peripheral blood mononuclear cells in the setting of peripheral arterial disease. J Clin Bioinforma. 2012;2:6. doi:10.1186/2043-9113-2-6. 10.1186/2043-9113-2-6. Source series: GSE27034.

[18] Chamaria S, Johnson KW, Vengrenyuk Y, et al. Intracoronary Imaging, Cholesterol Efflux, and Transcriptomics after Intensive Statin Treatment in Diabetes. Sci Rep. 2017;7(1):7001. doi:10.1038/s41598-017-07029-7. 10.1038/s41598-017-07029-7. Source series: GSE86216.

[19] Subramanian A, Tamayo P, Mootha VK, et al. Gene set enrichment analysis: a knowledge-based approach for interpreting genome-wide expression profiles. Proc Natl Acad Sci U S A. 2005;102(43):15545–15550. doi:10.1073/pnas.0506580102. 10.1073/pnas.0506580102

[20] Hänzelmann S, Castelo R, Guinney J. GSVA: gene set variation analysis for microarray and RNA-seq data. BMC Bioinformatics. 2013;14:7. doi:10.1186/1471-2105-14-7. 10.1186/1471-2105-14-7

[21] Irizarry RA, Hobbs B, Collin F, et al. Exploration, normalization, and summaries of high density oligonucleotide array probe level data. Biostatistics. 2003;4(2):249–264. doi:10.1093/biostatistics/4.2.249. 10.1093/biostatistics/4.2.249

[22] Hastie T, Tibshirani R, Friedman J. The elements of statistical learning: data mining, inference, and prediction. 2nd ed. New York: Springer; 2009. doi:10.1007/978-0-387-84858-7. 10.1007/978-0-387-84858-7

[23] Cawley GC, Talbot NLC. On over-fitting in model selection and subsequent selection bias in performance evaluation. J Mach Learn Res. 2010;11:2079–2107. https://www.jmlr.org/papers/ v11/cawley10a.html

[24] Efron B, Tibshirani RJ. An introduction to the bootstrap. New York: Chapman & Hall; 1993. doi:10.1007/978-1-4899-4541-9. 10.1007/978-1-4899-4541-9

[25] Bengio Y, Grandvalet Y. No Unbiased Estimator of the Variance of K-Fold Cross-Validation. Journal of Machine Learning Research. 2004;5:1089–1105. https://www.jmlr.org/papers/v5/grandvalet04a.html

[26] Peng RD. Reproducible research in computational science. Science. 2011;334(6060):1226–1227. doi:10.1126/science.1213847. 10.1126/science.1213847

[27] Goodman SN, Fanelli D, Ioannidis JPA. What does research reproducibility mean? Sci Transl Med. 2016;8(341):341ps12. doi:10.1126/scitranslmed.aaf5027. 10.1126/scitranslmed.aaf5027

[28] Johnson WE, Li C, Rabinovic A. Adjusting batch effects in microarray expression data using empirical Bayes methods. Biostatistics. 2007;8(1):118–127. doi:10.1093/biostatistics/kxj037. 10.1093/biostatistics/kxj037

[29] Leek JT, Storey JD. Capturing heterogeneity in gene expression studies by surrogate variable analysis. PLoS Genet. 2007;3(9):e161. doi:10.1371/journal.pgen.0030161. 10.1371/journal.pgen.0030161

[30] Wilkinson MD, Dumontier M, Aalbersberg IJJ, et al. The FAIR Guiding Principles for scientific data management and stewardship. Sci Data. 2016;3:160018. doi:10.1038/sdata.2016.18. 10.1038/sdata.2016.18

[31] Nosek BA, Alter G, Banks GC, et al. Promoting an open research culture. Science. 2015;348(6242):1422–1425. doi:10.1126/science.aab2374. 10.1126/science.aab2374

