## Supplementary Methods, Tables S1-S12 and Figures S1-S7 for "VascFlux-GEO: Comparator selection and auditability in vascular transcriptome module prediction"

### VascFlux-GEO supplementary material

This file contains aggregate methods and numerical detail. Public study accessions identify the source studies; source patient identifiers, sample-level linked records and individual ages are excluded from this submission supplement. The private review package retains the full computational provenance for author inspection. All extensions are post hoc.

#### S1. Data structure and analytical units

**Table S1:** Source and analysis denominators. Counts are not summed as independent patients.

| Study and tissue | Deposited and retained records | Role and time |
|---|---|---|
| GSE86216; PBMC | 144 samples; 72 paired individuals | Within-cohort baseline to 8-12-week follow-up during rosuvastatin 40 mg/day |
| GSE113873; gastrocnemius | 52 samples; 38 identifiers: 28 PAD and 10 healthy single-visit participants. 14 PAD pairs retained (5 training, 9 usual activity). | Separately refitted baseline/final comparison; 12-week protocol, not audited individual visit dates |
| GSE43292; carotid tissue | 64 specimens; 32 within-person pairs | Plaque versus macroscopically intact tissue in endarterectomy specimens; no temporal outcome |
| GSE27034; PBMC | 37 samples: 19 PAD and 18 controls | Cross-sectional case/control description; corrected complete GPL570 annotation |
| GSE281841; peripheral arteries | 41 columns; 28 literal identifiers before exclusions. 38 retained columns in 26 literal identifiers (18 high, 8 low). | Histological grade 4-5 versus 0-1; unresolved 41/28 versus source summary 39/27 discrepancy |

In GSE113873, 10 healthy participants were sampled once by design. Of 28 deposited PAD identifiers, 12 had baseline-only and 2 final-only records. These counts describe archive availability and do not establish clinical loss to follow-up. In GSE281841, one unscored column and two columns from the identifier contributing both grade groups were excluded; within-identifier specimens were then averaged. Literal identifiers remain an imperfect proxy for independent people.

### S2. Frozen module definitions

**Table S2:** Frozen curated gene sets.

| Module label | Genes (n) | Frozen genes |
|---|---|---|
| endothelial activation | 13 | PECAM1, VWF, ENG, KDR, NOS3, EDNRB, RAMP2, KLF2, KLF4, ICAM1, VCAM1, SELE, SELP |
| smooth muscle contractile | 9 | ACTA2, TAGLN, CNN1, MYH11, CALD1, DES, TPM2, LMOD1, MYLK |
| inflammation | 14 | IL1B, TNF, NFKB1, NFKBIA, CCL2, CCL5, CXCL8, CXCL10, NLRP3, TLR4, ICAM1, VCAM1, CSF1R, ITGAM |
| extracellular matrix | 11 | COL1A1, COL3A1, COL4A1, FN1, MMP2, MMP9, TIMP1, LUM, DCN, SPARC, ELN |
| coagulation thrombosis | 12 | F3, F5, F7, FGA, FGB, FGG, SERPINE1, THBS1, VWF, PF4, PPBP, PLAU |
| lipid handling | 11 | APOE, APOB, LDLR, ABCA1, ABCG1, LPL, CD36, SREBF2, HMGCR, MSR1, LIPA |
| oxidative stress | 9 | NQO1, HMOX1, SOD1, SOD2, GPX1, TXN, GCLC, GCLM, PRDX1 |
| hypoxia glycolysis | 8 | HIF1A, VEGFA, EGLN1, CA9, LDHA, PGK1, ENO1, ALDOA |

The lists contain 84 unique genes and overlap. Labels do not directly measure cellular function or pathway activity. Scores are the median of available standardized gene values. For prediction, standardization is fitted only on each training-baseline subset; this restriction starts after the source-study processed matrix and does not certify upstream RMA, lumi, limma or sva processing. No learned orthogonal factorization or neural pretraining was performed.

#### S3. Selected hyperparameters

**Table S3:** Selected values across 20 repeated five-fold partitions.

| Cohort | Candidate family | Selected value and count |
|---|---|---|
| GSE113873 | delta | 10: 59/100 |
| GSE113873 | delta | 100: 34/100 |
| GSE113873 | delta | 1000: 7/100 |
| GSE113873 | followup | 10: 1/100 |
| GSE113873 | followup | 100: 48/100 |
| GSE113873 | followup | 1000: 51/100 |
| GSE113873 | pca2 | 10: 3/100 |
| GSE113873 | pca2 | 100: 61/100 |
| GSE113873 | pca2 | 1000: 36/100 |
| GSE113873 | shrunken | 0: 80/100 |
| GSE113873 | shrunken | 0.25: 14/100 |
| GSE113873 | shrunken | 0.5: 6/100 |
| GSE86216 | delta | 10: 100/100 |
| GSE86216 | followup | 10: 16/100 |
| GSE86216 | followup | 100: 84/100 |
| GSE86216 | pca2 | 0.01: 2/100 |
| GSE86216 | pca2 | 1: 1/100 |
| GSE86216 | pca2 | 10: 53/100 |
| GSE86216 | pca2 | 100: 44/100 |
| GSE86216 | shrunken | 0.25: 73/100 |
| GSE86216 | shrunken | 0.5: 27/100 |

These 100 fits per cohort/family share participants and overlapping training sets. Counts are descriptive tuning frequencies, not 100 independent confirmations. Hyperparameter selection was nested within each specified candidate; the choice and presentation of candidates were post hoc.

### S4. Complete known-truth simulation summaries

#### independent followup

**Table S4:** Simulation summary: independent followup.

| Training n / method | Mean MAE | MCSE |
|---|---|---|
| 72 / mean followup | 0.8033 | 0.0006 |
| 72 / median followup | 0.8062 | 0.0006 |
| 72 / mean change | 1.1359 | 0.0008 |
| 72 / persistence | 1.1283 | 0.0007 |
| 72 / scalar inner selected | 0.8033 | 0.0006 |
| 72 / ridge inner selected | 0.8048 | 0.0007 |
| 300 / mean followup | 0.7993 | 0.0005 |
| 300 / median followup | 0.8000 | 0.0005 |
| 300 / mean change | 1.1300 | 0.0007 |
| 300 / persistence | 1.1283 | 0.0007 |
| 300 / scalar inner selected | 0.7993 | 0.0005 |
| 300 / ridge inner selected | 0.7999 | 0.0005 |

#### scalar persistence

**Table S5:** Simulation summary: scalar persistence.

| Training n / method | Mean MAE | MCSE |
|---|---|---|
| 72 / mean followup | 1.0052 | 0.0007 |
| 72 / median followup | 1.0095 | 0.0007 |
| 72 / mean change | 0.8293 | 0.0006 |
| 72 / persistence | 0.8235 | 0.0004 |
| 72 / scalar inner selected | 0.8050 | 0.0007 |
| 72 / ridge inner selected | 0.8439 | 0.0010 |
| 300 / mean followup | 0.9992 | 0.0006 |
| 300 / median followup | 1.0003 | 0.0006 |
| 300 / mean change | 0.8243 | 0.0005 |
| 300 / persistence | 0.8228 | 0.0005 |
| 300 / scalar inner selected | 0.7996 | 0.0005 |
| 300 / ridge inner selected | 0.8100 | 0.0005 |

#### off diagonal state

**Table S6:** Simulation summary: off diagonal state.

| Training n / method | Mean MAE | MCSE |
|---|---|---|
| 72 / mean followup | 1.0242 | 0.0007 |
| 72 / median followup | 1.0284 | 0.0009 |
| 72 / mean change | 1.1054 | 0.0008 |
| 72 / persistence | 1.0982 | 0.0007 |
| 72 / scalar inner selected | 0.9864 | 0.0007 |
| 72 / ridge inner selected | 0.8425 | 0.0011 |
| 300 / mean followup | 1.0187 | 0.0006 |
| 300 / median followup | 1.0195 | 0.0006 |
| 300 / mean change | 1.1002 | 0.0007 |
| 300 / persistence | 1.0984 | 0.0007 |
| 300 / scalar inner selected | 0.9811 | 0.0006 |
| 300 / ridge inner selected | 0.8097 | 0.0005 |

#### transport shift

**Table S7:** Simulation summary: transport shift.

| Training n / method | Mean MAE | MCSE |
|---|---|---|
| 72 / mean followup | 0.9017 | 0.0020 |
| 72 / median followup | 0.9057 | 0.0022 |
| 72 / mean change | 1.5695 | 0.0013 |
| 72 / persistence | 1.5659 | 0.0013 |
| 72 / scalar inner selected | 1.3445 | 0.0024 |
| 72 / ridge inner selected | 1.3105 | 0.0041 |
| 300 / mean followup | 0.8958 | 0.0011 |
| 300 / median followup | 0.8966 | 0.0012 |
| 300 / mean change | 1.5690 | 0.0012 |
| 300 / persistence | 1.5683 | 0.0012 |
| 300 / scalar inner selected | 1.3413 | 0.0010 |
| 300 / ridge inner selected | 1.3332 | 0.0024 |

Each cell contains 100 independent synthetic training/test replicates, with 2,000 synthetic test observations per replicate. There are 800 generated datasets and 48 scenario-by-size-by-method cells. MCSE is the replicate standard deviation divided by the square root of 100; it is distinct from patient uncertainty and from the spread of replicate errors. The simulation demonstrates configured behavior and implementation checks, not clinical findings.

#### S5. Cross-context directions after source correction

**Table S8:** All ten cross-context comparisons using complete GPL570 annotation.

| Contrast pair | Cosine | Conditional percentile interval |
|---|---|---|
| Statin PBMC / Exercise muscle | 0.4529 | [-0.3604, 0.7576] |
| Statin PBMC / Usual-activity muscle | 0.2680 | [-0.5121, 0.7256] |
| Statin PBMC / Carotid tissue | 0.7407 | [-0.1132, 0.8739] |
| Statin PBMC / PAD PBMC | 0.5553 | [-0.3229, 0.8034] |
| Exercise muscle / Usual-activity muscle | -0.0416 | [-0.5158, 0.5488] |
| Exercise muscle / Carotid tissue | -0.0732 | [-0.3255, 0.1421] |
| Exercise muscle / PAD PBMC | 0.1912 | [-0.2652, 0.6202] |
| Usual-activity muscle / Carotid tissue | 0.3811 | [-0.1183, 0.6163] |
| Usual-activity muscle / PAD PBMC | -0.3055 | [-0.7358, 0.2440] |
| Carotid tissue / PAD PBMC | 0.5029 | [-0.0792, 0.7458] |

All ten ranges include zero. These are descriptive comparisons of differently defined, differently sampled tissue contrasts. Normalization and annotation are held fixed during 5,000 whole-unit bootstrap draws. They do not establish longitudinal transfer, common treatment mechanisms or device suitability.

### S6. Restricted histological-grade descriptions

#### artery / peroneal

Retained literal identifiers: 7 high grade and 5 low grade.

**Table S9:** Restricted description: artery / peroneal.

| Module | High minus low | Conditional percentile interval |
|---|---|---|
| endothelial activation | 0.7233 | [0.3923, 1.0646] |
| smooth muscle contractile | -1.4260 | [-2.1042, -0.7896] |
| inflammation | 0.6568 | [0.2975, 1.0424] |
| extracellular matrix | 0.1615 | [-0.2077, 0.5388] |
| coagulation thrombosis | 0.0254 | [-0.3357, 0.3874] |
| lipid handling | 0.5290 | [0.0669, 1.0329] |
| oxidative stress | 0.5597 | [-0.1165, 1.2507] |
| hypoxia glycolysis | -0.1944 | [-1.0592, 0.5933] |

#### artery / posterior tibial

Retained literal identifiers: 6 high grade and 3 low grade.

**Table S10:** Restricted description: artery / posterior tibial.

| Module | High minus low | Conditional percentile interval |
|---|---|---|
| endothelial activation | 1.1423 | [0.5538, 1.6651] |
| smooth muscle contractile | -1.3446 | [-2.0517, -0.6252] |
| inflammation | 0.6385 | [0.0148, 1.3588] |
| extracellular matrix | 0.3190 | [-0.3912, 1.1666] |
| coagulation thrombosis | -0.4205 | [-0.7493, -0.1178] |
| lipid handling | 0.7298 | [0.1450, 1.2320] |
| oxidative stress | 0.7804 | [0.2221, 1.3868] |
| hypoxia glycolysis | -0.2946 | [-0.7218, 0.0987] |

#### title prefix proxy / 0092

Retained literal identifiers: 5 high grade and 4 low grade.

**Table S11:** Restricted description: title prefix proxy / 0092.

| Module | High minus low | Conditional percentile interval |
|---|---|---|
| endothelial activation | 1.3441 | [0.8624, 1.7885] |
| smooth muscle contractile | -2.0160 | [-2.6702, -1.4274] |
| inflammation | 1.0073 | [0.4408, 1.5702] |
| extracellular matrix | 1.0924 | [0.5154, 1.7731] |
| coagulation thrombosis | -0.2787 | [-0.4895, -0.0692] |
| lipid handling | 1.0017 | [0.3451, 1.6900] |
| oxidative stress | 0.1921 | [-0.3431, 0.6484] |
| hypoxia glycolysis | 0.7678 | [0.4119, 1.1163] |

#### title prefix proxy / 0521

Retained literal identifiers: 13 high grade and 4 low grade.

**Table S12:** Restricted description: title prefix proxy / 0521.

| Module | High minus low | Conditional percentile interval |
|---|---|---|
| endothelial activation | 0.6092 | [0.3250, 0.9358] |
| smooth muscle contractile | -1.2681 | [-1.8777, -0.5820] |
| inflammation | 0.6061 | [0.2838, 0.9246] |
| extracellular matrix | 0.1280 | [-0.2355, 0.4834] |
| coagulation thrombosis | -0.0346 | [-0.5320, 0.4474] |
| lipid handling | 0.4731 | [0.1607, 0.7736] |
| oxidative stress | 0.9482 | [0.4071, 1.4745] |
| hypoxia glycolysis | -0.3440 | [-0.8457, 0.1812] |

All 32 estimates are shown without multiplicity adjustment. Source-title prefixes are unverified technical proxies. Histological grade describes the sampled arteries, not a validated clinical severity category; lower-grade specimens are from the same end-stage PAD sampling context and are not healthy controls. No individual ages or original subject IDs are included here.

#### S7. Execution, source completeness and reproducibility

On 25 October 2026 the six numerical stages were reconstructed in a new virtual environment on the same host and Python base runtime. Historical files in results/ were retained only as original comparison anchors; current result directories started empty. Before the input correction, all 48 generated CSV tables agreed with the archived outputs. This reproducibility result did not prove that the archived sources were complete.

Fresh acquisition exposed an incomplete historical GPL570 export: 44,562 table rows and no end marker, versus 54,675 rows in the complete official export. Module-mapped probes increased from 214 to 229. GSE27034 descriptive outputs and their cross-context cosines were regenerated. Paired prediction, simulations, representation sensitivities and the independent-source artery description were unaffected. A complete corrected six-stage reconstruction then reproduced all 48 updated reference tables exactly.

The new entry point checks hashes, table begin/end markers, and expected row counts and normalized table digests where specified. This is a bounded source-integrity guard: it does not authenticate the truth of source annotations or exclude unreported upstream processing. The complete GPL14951 and GPL96 platform tables were obtained through official family streams; their entire family archives were not downloaded or verified. Download and reconstruction timestamps, expected hashes, package versions and random seeds are in the accompanying provenance records.

```
python -m pip install -r requirements-analysis.txt
python reproduce.py --verify-only
# For refitting, acquire complete inputs first (see README).
python reproduce.py --full --data-dir canonical_cache \
  --source-manifest canonical_cache/canonical_manifest.json
```

Output-only verification does not acquire sources or refit models. A public repository URL, permanent release identifier and distribution licence have not been approved or issued by this preparation. The review package must not be described as a published software release.

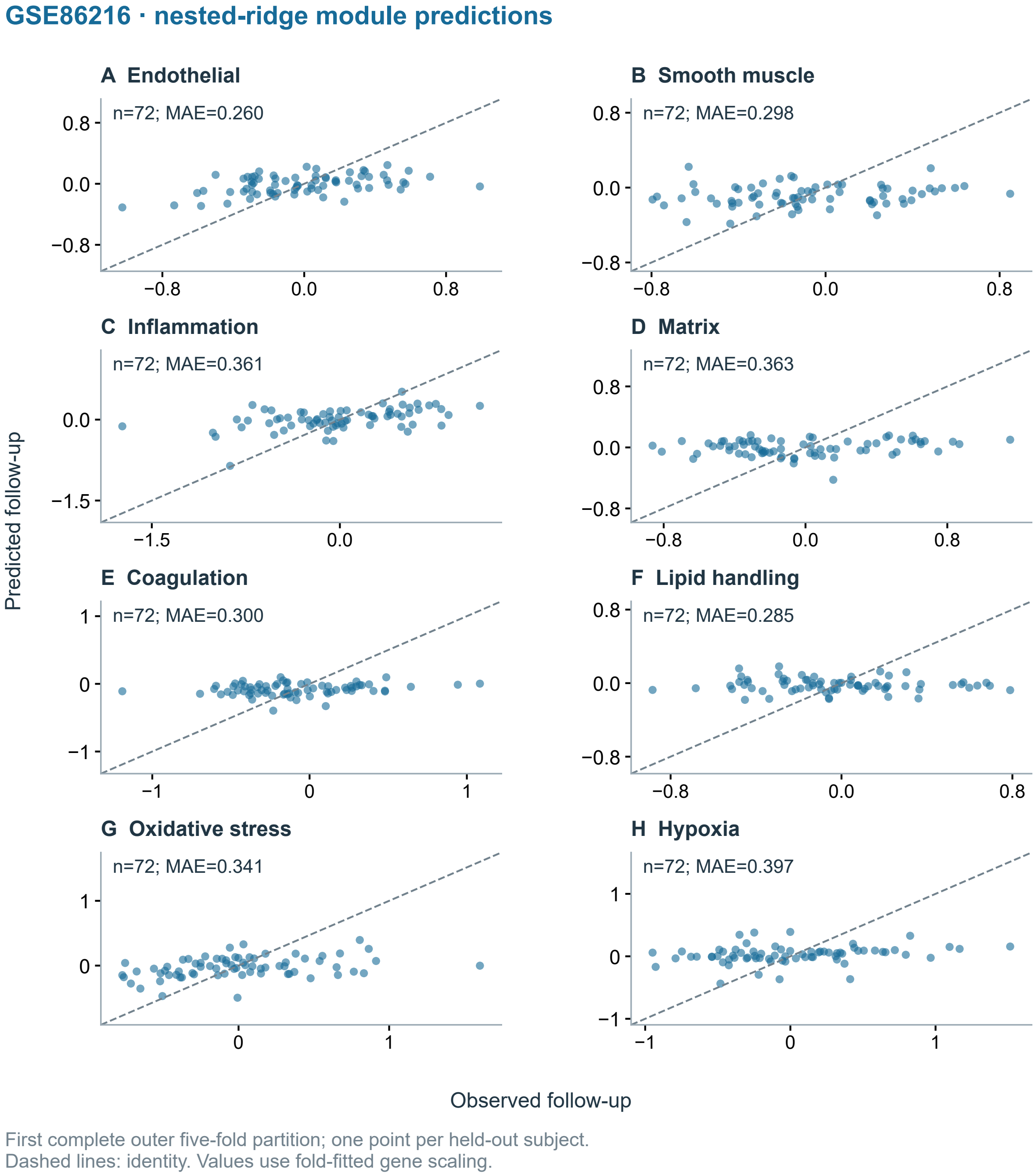


**Figure S1:** Observed and nested-ridge predicted follow-up modules for GSE86216, using the first complete outer five-fold partition only. Each point represents one held-out subject; the diagonal is identity, and per-module MAE uses the fold-fitted gene scale. This is a diagnostic visualization, not treatment-mechanism evidence.

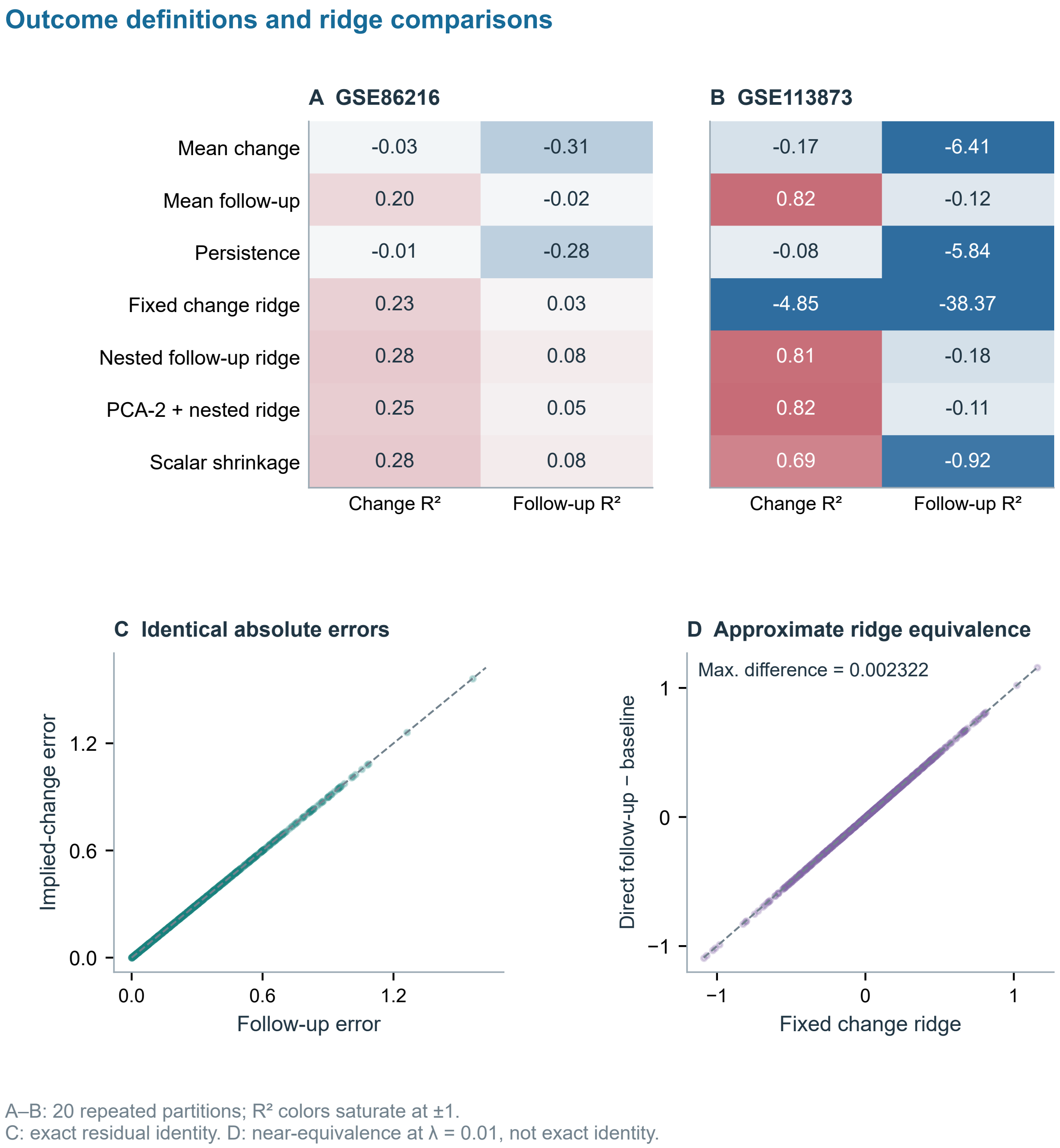


**Figure S2:** Outcome-dependent R-squared and prediction identities. A and B average 20 complete outer repetitions with separate module centering; colors saturate at minus/plus 1 while numerical labels preserve the values. C shows the exact equality of absolute follow-up and implied-change residuals. D compares fixed change ridge with direct follow-up ridge minus baseline: at lambda = 0.01 these are close, not exact, parameterizations (maximum absolute difference 0.00232210). C and D use original statin five-fold predictions and supply no independent evidence of response biology.

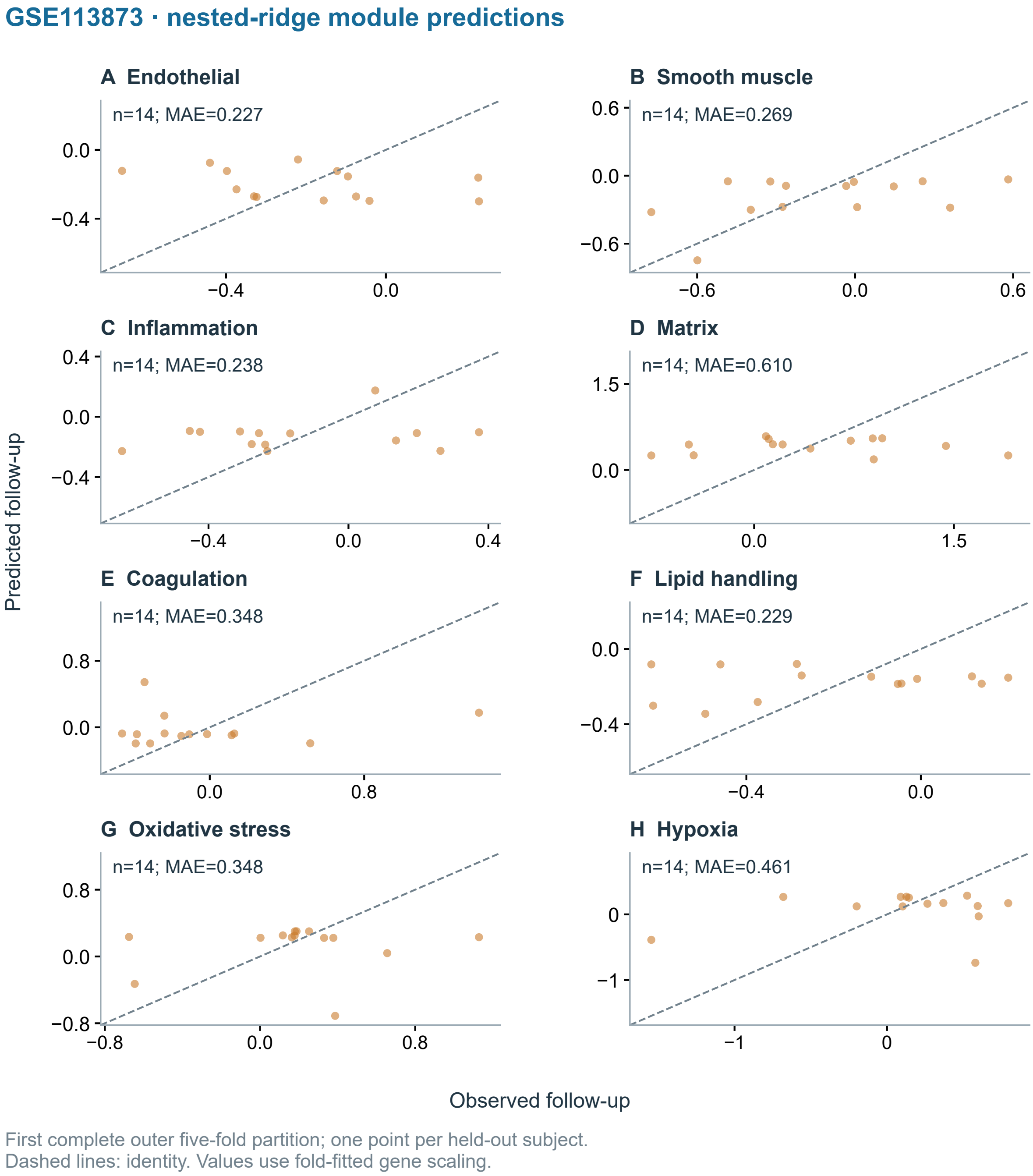


**Figure S3:** Observed and nested-ridge predicted follow-up modules for GSE113873, using the first complete outer five-fold partition only. Each point represents one held-out subject; the diagonal is identity, and per-module MAE uses the fold-fitted gene scale. This is a diagnostic visualization, not treatment-mechanism evidence.

#### Inner-fold tuning selections across repeated partitions

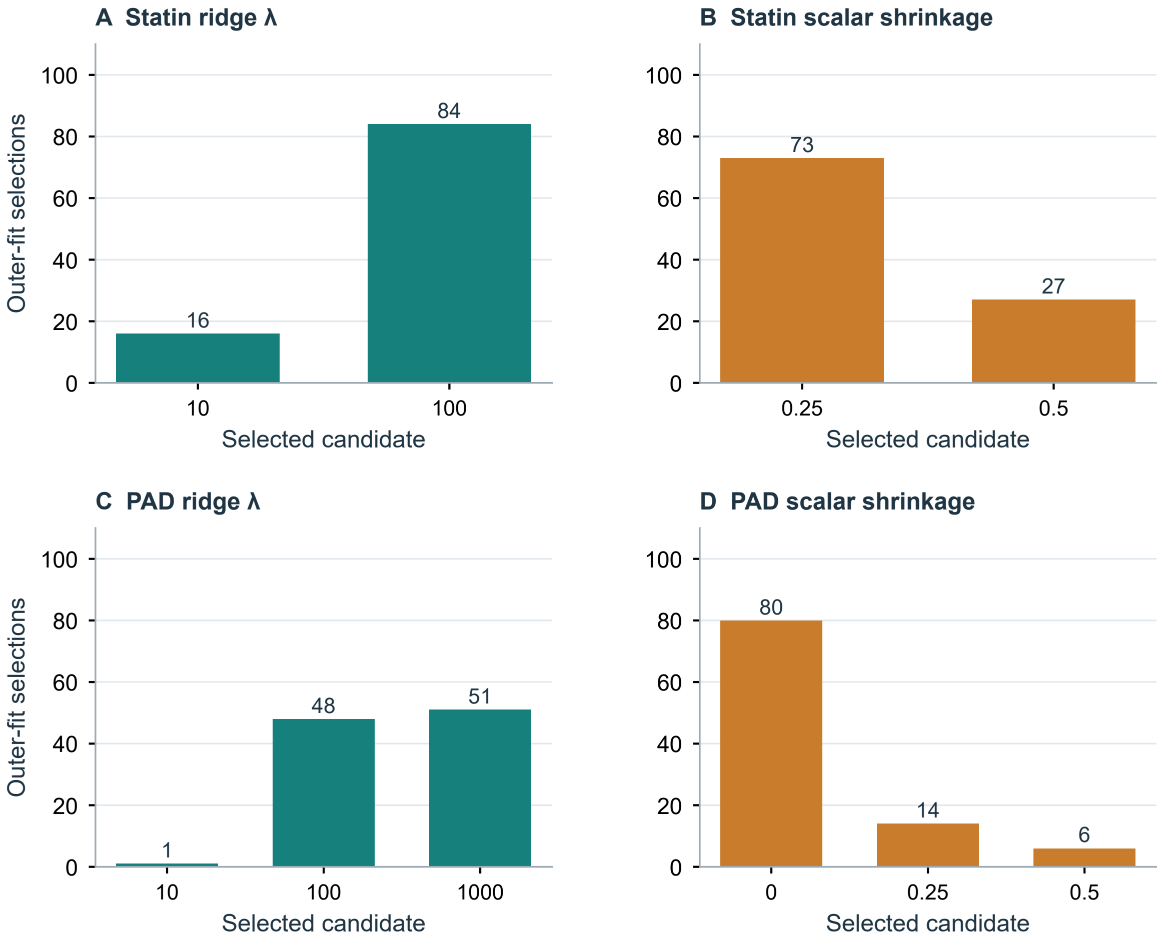


100 outer fits per cohort and model. Fits share subjects; frequencies are diagnostic.
Candidate selection is confined to the training folds.

**Figure S4:** Selected ridge and scalar-shrinkage candidates across 100 outer fits per cohort. Candidate frequencies expose tuning behavior; fits share subjects and are not independent biological observations. Severe PAD shrinkage frequently approximates the mean-follow-up comparator.

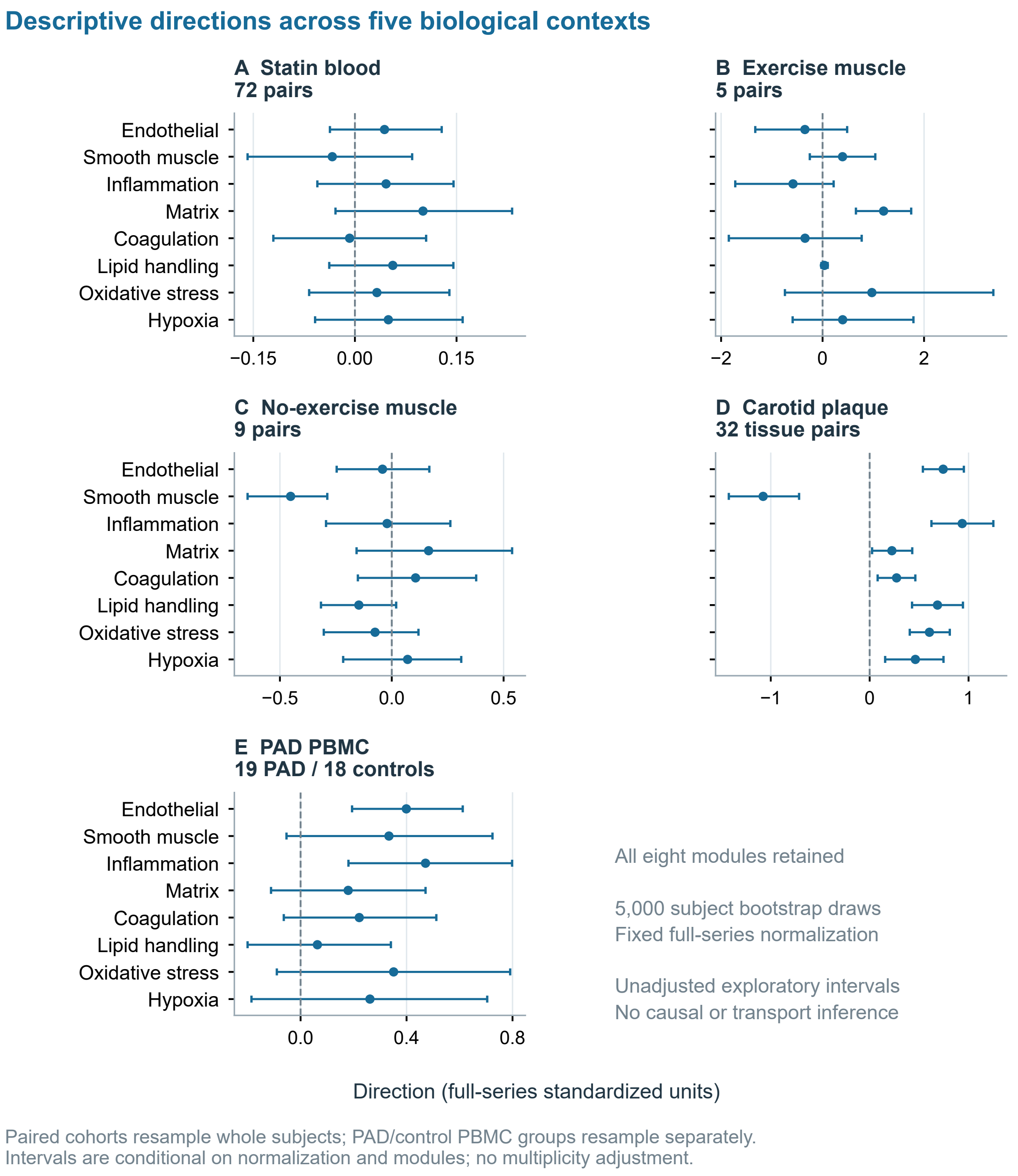


**Figure S5:** All eight module directions in five separately defined contexts: statin 72 pairs, exercise 5 pairs, no exercise 9 pairs, carotid 32 tissue pairs, and PAD PBMC 19 cases versus 18 controls. Descriptive percentile ranges use 5,000 subject bootstrap draws holding full-series normalization and modules fixed; paired observations are resampled together and independent PBMC groups separately. The PBMC results use the complete GPL570 annotation. All contrasts are exploratory and unadjusted; no calibrated population coverage is established, and none validates longitudinal transport or causal treatment effects.

#### Engineered control outcomes and positive-control effect

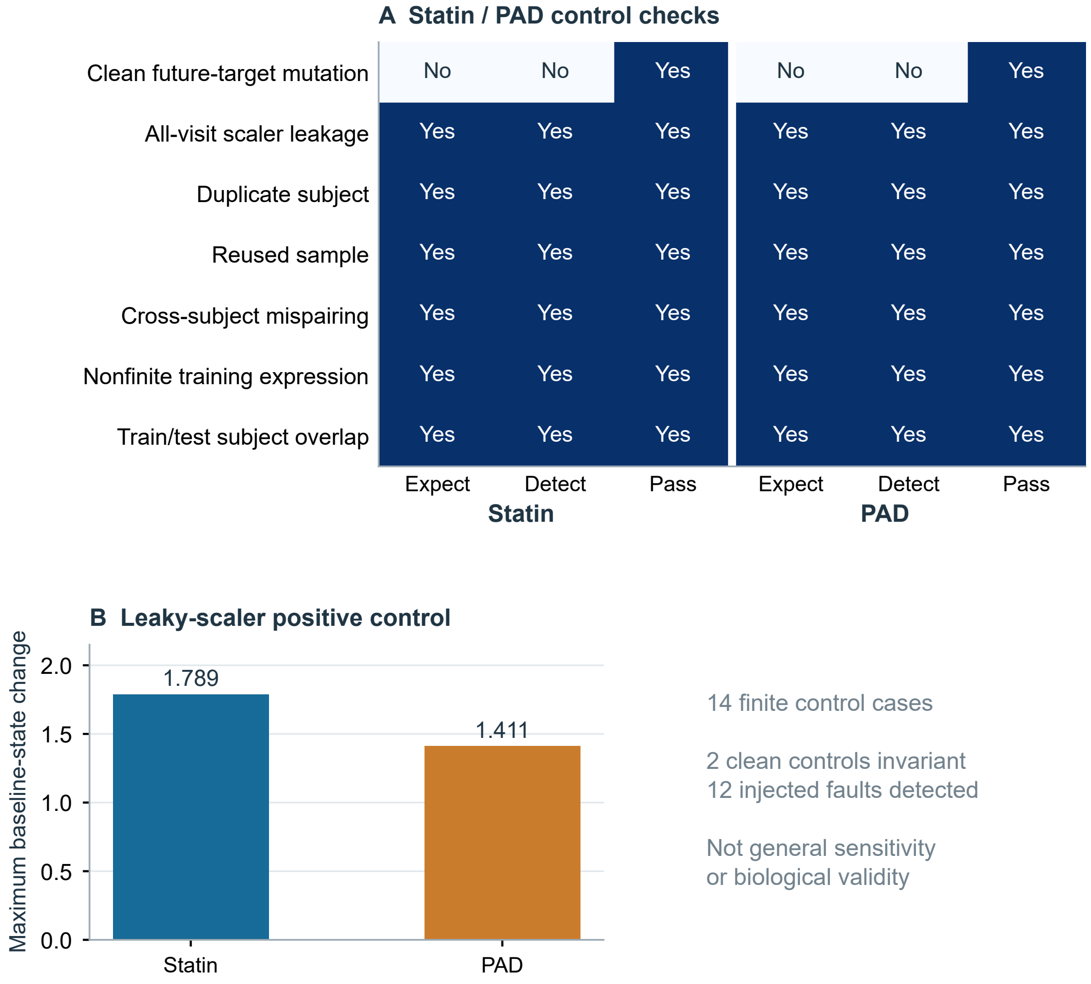


Clean mutation checks: first fold of the first repetition; tuning held fixed.
Positive control deliberately contaminates scaling with future expression.

**Figure S6:** All fourteen engineered control outcomes: two clean mutation cases correctly remain invariant and twelve injected faults are detected. Clean mutation checks use the first fold of the first repetition per cohort with tuning parameters held fixed. The positive all-visit-scaling control measures altered training-baseline module states after future expression mutation. These finite cases do not estimate general verifier sensitivity or establish biological validity.

#### Cross-context comparisons and their uncertainty

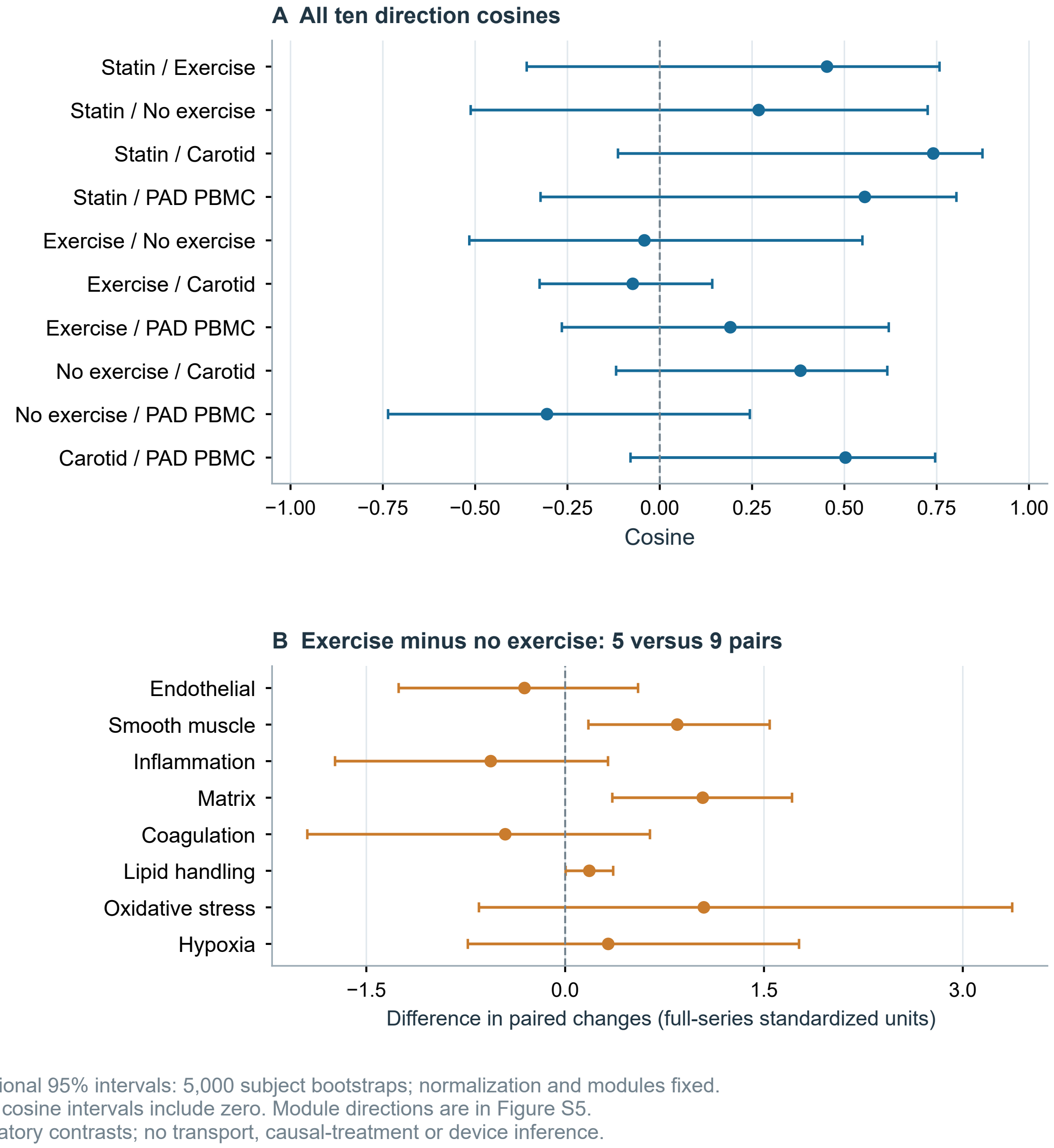


**Figure S7:** A shows all ten cross-context cosine estimates and their descriptive central 95% percentile ranges; B shows exercise-minus-no-exercise paired-change contrasts for all eight modules. Five thousand subject bootstrap draws hold full-series normalization and modules fixed. Exercise and no-exercise groups contain 5 and 9 pairs; PBMC contains 19 cases and 18 controls. Complete GPL570 annotation is used for PBMC directions. All ten cosine ranges include zero; the nominal level has no validated population coverage guarantee. Module-direction estimates are in Figure S5; duplicated heatmaps have been removed. These exploratory contrasts do not validate transported prediction, causal treatment effects or device suitability.
