## Supplementary material for "VascFlux-GEO: Comparator selection and auditability in vascular transcriptome module prediction": Analysis code and aggregate results: finalize_figures.py

```
"""Verify V07 export inventories and publish the combined manifest and captions.

Run after redraw_main_figures.py and redraw_supplement_figures.py.
This checks existing exports and data hashes; it does not fit or simulate models.
"""
from pathlib import Path
from datetime import datetime, timezone
import hashlib
import json
import platform
import re
import numpy as np
import pandas as pd
import matplotlib
from PIL import Image

ROOT=Path(__file__).resolve().parent
SRC=ROOT.parent/'reproducibility'
sha=lambda path:hashlib.sha256(path.read_bytes()).hexdigest()

def main():
    main_figures=json.loads((ROOT/'figure_manifest.json').read_text(encoding='utf-8'))
    supplementary=json.loads((ROOT/'supplementary_figure_manifest.json').read_text(encoding='utf-8'))
    specs=json.loads((ROOT/'figure_specs.json').read_text(encoding='utf-8'))
    all_figures=main_figures+supplementary
    assert len(main_figures)==8 and len(supplementary)==7
    assert [r['figure'] for r in all_figures]==[r['figure'] for r in specs]
    checks=[]
    for row,spec in zip(all_figures,specs):
        name=row['figure']
        assert row['caption']==spec['caption']
        assert row['minimum_font_pt']>=8 and row['text_within_canvas']
        assert row['nominal_width_mm']==170
        for relative,expected in row['source_sha256'].items():
            assert sha(SRC/relative)==expected,(name,relative,'source drift')
        for ext,expected in row['sha256'].items():
            assert sha(ROOT/f'{name}.{ext}')==expected,(name,ext,'export drift')
        with Image.open(ROOT/f'{name}.png') as im:
            png_size=im.size
            assert abs(png_size[0]-170/25.4*600)<=1
            assert abs(im.info['dpi'][0]-600)<.1
        svg=(ROOT/f'{name}.svg').read_text(encoding='utf-8')
        width_pt=float(re.search(r'<svg[^>]*\bwidth="([0-9.]+)pt"',svg).group(1))
        assert abs(width_pt/72*25.4-170)<.001
        checks.append({'figure':name,'png_pixels':png_size,'svg_width_mm':round(width_pt/72*25.4,6),'minimum_font_pt':row['minimum_font_pt'],'source_and_export_hashes_match':True})
    assert len(list(ROOT.glob('Figure*.png')))==15,'Retired assets must be kept in history only'
    (ROOT/'all15_figure_manifest.json').write_text(json.dumps(all_figures,indent=2,ensure_ascii=False)+'\n',encoding='utf-8')
    mapping={'main_figures':8,'supplementary_figures':7,'main_labelled_panels':sum(r['panels'] for r in main_figures),'main_effective_quantitative_panels':sum(r['effective_quantitative_panels'] for r in main_figures),'supplementary_labelled_panels':sum(r['panels'] for r in supplementary),'counting_scope':'Descriptive presentation counts; not independent experiments or external validations.','figure_mapping':[{'current':r['figure'],'former':r['former_figure'],'panels':r['panels'],'caption':r['caption']} for r in all_figures]}
    (ROOT/'figure_mapping.json').write_text(json.dumps(mapping,indent=2,ensure_ascii=False)+'\n',encoding='utf-8')
    qa={'completed_utc':datetime.now(timezone.utc).isoformat(),'runtime':{'python':platform.python_version(),'numpy':np.__version__,'pandas':pd.__version__,'matplotlib':matplotlib.__version__},'figures':checks,'main_labelled_panels':mapping['main_labelled_panels'],'main_effective_quantitative_panels':mapping['main_effective_quantitative_panels'],'supplementary_labelled_panels':mapping['supplementary_labelled_panels'],'visual_inspection_scope':'All 15 exported PNGs actually opened and inspected in V07; final assembled manuscript pages remain for the assembly task.','assembly_requirement':'Use native 170 mm width without a height cap that shrinks the asset. Place caption on the same figure page.','script_sha256':{name:sha(ROOT/name) for name in ['redraw_main_figures.py','redraw_supplement_figures.py','finalize_figures.py','figure_specs.json']}}
    (ROOT/'V07_figure_validation.json').write_text(json.dumps(qa,indent=2,ensure_ascii=False)+'\n',encoding='utf-8')
    (ROOT/'REDESIGN_QA.txt').write_text('V07: 8 main and 7 supplementary figures. All 15 PNGs visually inspected.\nAll exports use 170 mm native width and at least 8 pt visible type.\nFormats: 600 dpi PNG, vector PDF, editable SVG, LZW TIFF.\n31 labelled and 23 effective quantitative main panels; these are not independent experiments.\nComplete GPL570 descriptive results are bound through ../reproducibility source hashes.\nSee V07_figure_validation.json for checks; final manuscript page assembly requires separate visual QA.\n',encoding='utf-8')
    print(json.dumps({'verified_figures':len(checks),'main':8,'supplementary':7,'main_labelled_panels':mapping['main_labelled_panels'],'main_effective_quantitative_panels':mapping['main_effective_quantitative_panels'],'minimum_font_pt':min(r['minimum_font_pt'] for r in all_figures)},indent=2))

if __name__=='__main__':
    main()
```
