## Supplementary material for "VascFlux-GEO: Comparator selection and auditability in vascular transcriptome module prediction": Analysis code and aggregate results: redraw_main_figures.py

```
"""Eight publication-width scientific figures from existing VascFlux-GEO results.
No numerical re-analysis or invented evidence. Output width = 170 mm; fonts >= 8 pt.
"""
from pathlib import Path
import ast, hashlib, json
import numpy as np
import pandas as pd
import matplotlib
matplotlib.use('Agg')
import matplotlib.pyplot as plt
from matplotlib.patches import FancyBboxPatch, FancyArrowPatch
from matplotlib.colors import LinearSegmentedColormap
from matplotlib.ticker import MaxNLocator

OUT=Path(__file__).resolve().parent
SRC=OUT.parent/'reproducibility'
R=SRC/'results_reviewer_revision'; E=SRC/'results_extended'
W=170/25.4
C=dict(ink='#203543',blue='#176B98',teal='#16817C',orange='#CA7C2D',red='#B75260',gray='#72828D',light='#EDF3F6',purple='#8063A4')
plt.rcParams.update({'font.family':'Arial','font.size':9,'axes.titlesize':10,'axes.labelsize':9,'xtick.labelsize':8,'ytick.labelsize':8.5,'text.color':C['ink'],'axes.labelcolor':C['ink'],'axes.edgecolor':'#9BAAB4','axes.spines.top':False,'axes.spines.right':False,'pdf.fonttype':42,'svg.fonttype':'none','savefig.facecolor':'white','axes.linewidth':.65})
DIV=LinearSegmentedColormap.from_list('vascular',['#2F6D9F','#F8F9FA','#BD4F5C'])
S=json.loads((SRC/'results/summary.json').read_text()); MOD=S['module_names']
LAB=['Endothelial','Smooth muscle','Inflammation','Matrix','Coagulation','Lipid handling','Oxidative stress','Hypoxia']
ABBR=['Endo.','SMC','Inflam.','Matrix','Coag.','Lipid','Oxid.','Hypox.']
OLD={x['figure']:x for x in json.loads((OUT/'figure_specs.json').read_text(encoding='utf-8'))}
MAN=[]

def title(fig,letter,text,x,y):
    fig.text(x,y,letter,fontsize=12,fontweight='bold',va='top',color=C['blue'])
    fig.text(x+.037,y-.002,text,fontsize=10,fontweight='bold',va='top')
def clean(ax,axis='x'):
    ax.set_axisbelow(True); ax.grid(axis=axis,color='#E1E8ED',lw=.6)
    ax.tick_params(length=2.5,pad=4)
def canvas(h):
    return plt.figure(figsize=(W,h),facecolor='white')
def note(fig,text,y=.018,color=None):
    fig.text(.035,y,text,fontsize=8.2,va='bottom',linespacing=1.3,color=color or C['gray'])
def card(ax,x,y,w,h,head,body,color='blue',hs=9.4,bs=8.7):
    col=C.get(color,color)
    ax.add_patch(FancyBboxPatch((x,y),w,h,boxstyle='round,pad=.008,rounding_size=.012',fc='#F8FAFB',ec='#CBD8E0',lw=.75))
    ax.plot([x+.018,x+.018],[y+.035,y+h-.035],color=col,lw=2.4)
    ax.text(x+.045,y+h-.045,head,fontsize=hs,fontweight='bold',va='top',color=col,linespacing=1.15)
    ax.text(x+.045,y+h-.16,body,fontsize=bs,va='top',linespacing=1.35)
def off(ax):
    ax.set(xlim=(0,1),ylim=(0,1)); ax.axis('off')
def arrow(ax,a,b):
    ax.add_patch(FancyArrowPatch(a,b,arrowstyle='-|>',mutation_scale=11,color=C['gray'],lw=1))
def save(fig,name):
    # Fixed canvas preserves the nominal 170 mm width, unlike tight-bbox export.
    fig.canvas.draw()
    text_sizes=[x.get_fontsize() for x in fig.findobj(matplotlib.text.Text) if x.get_text()]
    assert min(text_sizes)>=8, (name,min(text_sizes))
    renderer=fig.canvas.get_renderer();fw,fh=fig.canvas.get_width_height();outside=[];skip=set()
    for a in fig.axes:
        xl=sorted(a.get_xlim());yl=sorted(a.get_ylim())
        for tick in a.xaxis.get_major_ticks()+a.xaxis.get_minor_ticks():
            if not xl[0] <= tick.get_loc() <= xl[1]:skip.update([id(tick.label1),id(tick.label2)])
        for tick in a.yaxis.get_major_ticks()+a.yaxis.get_minor_ticks():
            if not yl[0] <= tick.get_loc() <= yl[1]:skip.update([id(tick.label1),id(tick.label2)])
    for t in fig.findobj(matplotlib.text.Text):
        if not t.get_visible() or not t.get_text() or id(t) in skip:continue
        b=t.get_window_extent(renderer)
        if b.x0 < -1 or b.y0 < -1 or b.x1 > fw+1 or b.y1 > fh+1:
            outside.append({'text':t.get_text(),'bbox_px':[round(v,2) for v in b.extents]})
    assert not outside, (name,outside)
    for ext in ('pdf','svg','png'):
        fig.savefig(OUT/f'{name}.{ext}',dpi=600,facecolor='white')
    fig.savefig(OUT/f'{name}.tiff',dpi=600,facecolor='white',pil_kwargs={'compression':'tiff_lzw'})
    meta=dict(OLD[name]);meta['formats']=['png_600dpi','pdf_vector','svg_editable','tiff_600dpi_lzw'];meta['nominal_width_mm']=170
    meta['height_mm']=round(fig.get_figheight()*25.4,1);meta['minimum_font_pt']=min(text_sizes)
    meta['text_within_canvas']=not outside
    meta['source_sha256']={p:hashlib.sha256((SRC/p).read_bytes()).hexdigest() for p in meta['source_files']}
    meta['source_root']='../reproducibility'
    meta['sha256']={ext:hashlib.sha256((OUT/f'{name}.{ext}').read_bytes()).hexdigest() for ext in ('png','pdf','svg','tiff')}
    meta['redesign']='V07: portable redraw of executed results; no new fitted experiments. Updated descriptive CSVs include complete GPL570 annotation.'
    MAN.append(meta);plt.close(fig)
    print(name, 'saved', flush=True)

def framework():
    fig=canvas(7.6)
    title(fig,'A','Prediction is evaluated on complete held-out subjects',.035,.97)
    ax=fig.add_axes([.035,.725,.93,.19]);off(ax)
    card(ax,.008,.10,.29,.85,'Baseline state','Training baselines\n→ gene scaling\n→ 8 module medians',bs=8.8)
    card(ax,.355,.10,.29,.85,'Nested fitting','Inner-fold selection\n→ direct follow-up ridge\n→ predicted follow-up F̂','teal',bs=8.8)
    card(ax,.702,.10,.29,.85,'Held-out target','Observed follow-up F\nSubject-level MAE\nΔ̂ = F̂ − B','orange',bs=8.8)
    arrow(ax,(.305,.52),(.345,.52));arrow(ax,(.652,.52),(.692,.52))
    title(fig,'B','A matched comparator design',.035,.69)
    ax=fig.add_axes([.035,.445,.93,.205]);off(ax)
    card(ax,.008,.06,.475,.90,'Same people, same transforms','20 repeated five-fold partitions\nMean / median follow-up; persistence\nScalar shrinkage; group controls',hs=9.5,bs=9)
    card(ax,.517,.06,.475,.90,'Retrospective focal contrast','Comparator MAE − method MAE\nSubject-averaged out-of-fold errors\nBootstrap conditions on fitted predictions','teal',hs=9.5,bs=9)
    title(fig,'C','Four evidence layers with different interpretation',.035,.407)
    ax=fig.add_axes([.035,.16,.93,.205]);off(ax)
    rows=[('GSE86216','72 subject pairs','Within-cohort prediction','blue'),('GSE113873','14 subject pairs','Separate refit; no transport','orange'),('Known truth','800 simulated datasets','4 scenarios × 2 sample sizes','purple'),('Cross-sectional','3 independent GEO series','Descriptive; no time forecast','gray')]
    for i,(h,n,b,c) in enumerate(rows):
        x=.008+(i%2)*.509;y=.54-(i//2)*.52
        ax.add_patch(FancyBboxPatch((x,y),.475,.43,boxstyle='round,pad=.008,rounding_size=.012',fc='#F8FAFB',ec='#CBD8E0',lw=.75))
        ax.text(x+.025,y+.34,h,fontsize=9.4,fontweight='bold',color=C[c],va='top')
        ax.text(x+.025,y+.20,n+' · '+b if i<2 else n+'\n'+b,fontsize=8.4,va='top',linespacing=1.3)
    title(fig,'D','An auditable chain with explicit limits',.035,.128)
    fig.text(.072,.085,'Source hashes → exclusions → splits → predictions → figures → release',fontsize=9.1,va='top')
    note(fig,'All revised analyses are post hoc. Independent longitudinal transport,\nclinical utility and device benefit remain unestablished.',y=.022)
    save(fig,'Figure1_framework')

def atlas():
    ext=json.loads((E/'summary_external_GSE281841.json').read_text())
    expected={'source_samples':41,'source_patient_ids':28,'usable_extreme_samples':40,
              'retained_samples':38,'patient_count_after_excluding_mixed':26,
              'high_4_5_patients':18,'low_0_1_patients':8}
    assert all(ext[k]==v for k,v in expected.items()), 'Update the displayed source-flow labels'
    assert [S['datasets'][a]['samples'] for a in ['GSE86216','GSE113873','GSE43292','GSE27034']]==[144,52,64,37]
    assert [S['paired_counts'][a] for a in ['statin_coronary','exercise_PAD','carotid_plaque']]==[72,14,32]
    assert S['paired_counts']['PAD_PBMC']=={'PAD':19,'control':18}
    fig=canvas(8.8)
    title(fig,'A','Five data sources; samples are not independent subjects',.035,.978)
    ax=fig.add_axes([.035,.785,.93,.15]);ax.axis('off')
    rows=[['GSE86216','144','72 subject pairs','Temporal benchmark'],['GSE113873','52','14 subject pairs','Separate PAD refit'],['GSE43292','64','32 tissue pairs','Spatial description'],['GSE27034','37','19 PAD + 18 controls','Case/control description'],['GSE281841','41','26 retained literal IDs','Severity description']]
    tb=ax.table(cellText=rows,colLabels=['GEO series','Samples','Analysis units','Role'],colWidths=[.18,.12,.30,.40],cellLoc='left',bbox=[0,0,1,1]);tb.auto_set_font_size(False);tb.set_fontsize(8.4)
    for (i,j),cell in tb.get_celld().items():
        cell.set_edgecolor('#D3DFE7');cell.set_linewidth(.5);cell.set_facecolor('#E6EFF4' if i==0 else ('#F7F9FA' if i%2==0 else 'white'));cell.PAD=.08
        if i==0:cell.get_text().set_fontweight('bold')
    title(fig,'B','Frozen module coverage',.035,.746)
    ax=fig.add_axes([.235,.48,.66,.225]);total=np.array([13,9,14,11,12,11,9,8]);acc=['GSE86216','GSE113873','GSE43292','GSE27034']
    coverage=pd.read_csv(E/'external_GSE281841_module_coverage.csv').set_index('module').reindex(MOD)
    assert np.array_equal(coverage.requested_genes.to_numpy(),total)
    v=np.array([[S['datasets'][a]['module_genes'][m] for a in acc]+[int(coverage.loc[m,'mapped_genes'])] for i,m in enumerate(MOD)])
    im=ax.imshow(v/total[:,None],cmap='Blues',vmin=.9,vmax=1,aspect='auto');ax.set_yticks(range(8),LAB);ax.set_xticks(range(5),['86216','113873','43292','27034','281841']);ax.set_xlabel('GEO accession suffix',labelpad=5);ax.tick_params(length=0)
    for i in range(8):
        for j in range(5):ax.text(j,i,f'{v[i,j]}/{total[i]}',ha='center',va='center',color='white' if v[i,j]/total[i]>.95 else C['ink'],fontsize=8.5)
    cax=fig.add_axes([.918,.48,.017,.225]);cb=fig.colorbar(im,cax=cax);cb.set_ticks([.9,.95,1]);fig.text(.88,.72,'Fraction',fontsize=8.2,color=C['gray'])
    title(fig,'C','External-source exclusions',.035,.418)
    ax=fig.add_axes([.035,.072,.37,.303]);off(ax)
    steps=[('41 matrix columns','28 literal source IDs'),('40 scored samples','Exclude 1 unscored sample'),('38 retained samples','Exclude mixed-grade ID'),('26 retained literal IDs','18 high-grade / 8 low-grade')]
    for i,(h,b) in enumerate(steps):
        y=.77-i*.245
        ax.add_patch(FancyBboxPatch((.01,y),.98,.20,boxstyle='round,pad=.008,rounding_size=.012',fc='#F5F9FB',ec='#CBD8E0',lw=.75))
        ax.text(.055,y+.155,h,fontsize=9,fontweight='bold',va='top',color=C['blue'] if i<3 else C['teal']);ax.text(.055,y+.067,b,fontsize=8.2,va='top')
        if i<3:arrow(ax,(.5,y-.006),(.5,y-.042))
    title(fig,'D','Module gene overlap',.475,.418)
    tree=ast.parse((SRC/'run_vascflux_public.py').read_text());genes=next(ast.literal_eval(n.value) for n in tree.body if isinstance(n,ast.Assign) and any(isinstance(t,ast.Name) and t.id=='MODULES' for t in n.targets))
    mat=np.array([[len(set(genes[a])&set(genes[b])) for b in MOD] for a in MOD],float);np.fill_diagonal(mat,np.nan);cm=plt.get_cmap('Blues').copy();cm.set_bad('#EFF3F6')
    ax=fig.add_axes([.565,.16,.385,.215]);im=ax.imshow(mat,cmap=cm,vmin=0,vmax=3,aspect='auto');ax.set_yticks(range(8),ABBR);ax.set_xticks(range(8),ABBR,rotation=55,ha='right');ax.tick_params(length=0)
    for i in range(8):
        for j in range(8):ax.text(j,i,'—' if i==j else f'{mat[i,j]:.0f}',ha='center',va='center',fontsize=8,color='white' if mat[i,j]>=2 else C['ink'])
    cax=fig.add_axes([.565,.081,.385,.013]);cb=fig.colorbar(im,cax=cax,orientation='horizontal');cb.set_ticks([0,1,2,3]);cb.set_label('Shared gene symbols; diagonal masked',fontsize=8.2,labelpad=3)
    note(fig,'GSE281841: matrix 41 columns / 28 IDs versus GEO 39 samples / 27 patients (unresolved).\nLiteral source IDs are not independently verified patient identities.',y=.012)
    save(fig,'Figure2_data_atlas')

METHODS=['mean_followup','median_followup','shrunken_state_followup_nested','followup_ridge_nested','persistence','mean_delta','median_delta','group_mean_followup','group_median_followup','group_mean_delta','group_median_delta','pca2_followup_nested','delta_ridge_nested','followup_ridge_fixed_0.01','delta_ridge_fixed_0.01']
ML=['Mean follow-up','Median follow-up','Scalar shrinkage','Nested follow-up ridge','Persistence','Mean change','Median change','Group mean follow-up','Group median follow-up','Group mean change','Group median change','PCA-2 + nested ridge','Nested change ridge','Fixed follow-up ridge','Fixed change ridge']
COL={m:C['gray'] for m in METHODS};COL.update(mean_followup=C['ink'],median_followup=C['purple'],shrunken_state_followup_nested=C['orange'],followup_ridge_nested=C['teal'])
def primary():
    rep=pd.read_csv(R/'fair_repeat_metrics.csv');dif=pd.read_csv(R/'fair_paired_differences.csv');old=pd.read_csv(E/'conditional_subject_intervals.csv')
    fig=canvas(8.8)
    for k,(cohort,n) in enumerate([('GSE86216',72),('GSE113873',14)]):
        left=.30+k*.35
        title(fig,'AB'[k],f'{cohort} · n = {n}',left,.974)
        ax=fig.add_axes([left,.465,.305,.46]);rng=np.random.default_rng(31)
        for j,m in enumerate(METHODS):
            vals=rep[(rep.cohort==cohort)&(rep.method==m)].MAE.to_numpy();ax.scatter(vals,j+rng.uniform(-.10,.10,len(vals)),s=9,color=COL[m],alpha=.30,edgecolors='none');ax.scatter(vals.mean(),j,s=25,color=COL[m],edgecolor='white',lw=.35,zorder=5)
        ax.set_yticks(range(15),ML if k==0 else ['']*15);ax.set_ylim(14.6,-.6);ax.axhspan(-.5,3.5,color='#EAF3F5',zorder=0);ax.axhline(3.5,color='#BCCAD3',lw=.7);ax.set_xlabel('Held-out MAE',labelpad=7);ax.xaxis.set_major_locator(MaxNLocator(4));clean(ax)
        ax.text(.5,-.135,'72 paired subjects' if k==0 else '14 paired subjects; separate refit',transform=ax.transAxes,fontsize=8.2,ha='center',va='top',color=C['gray'])
    fig.text(.035,.94,'All 15 specifications',fontsize=9.2,fontweight='bold',va='top')
    title(fig,'C','Paired error differences',.035,.365)
    fig.text(.30,.322,'GSE86216',fontsize=9.5,fontweight='bold');fig.text(.65,.322,'GSE113873',fontsize=9.5,fontweight='bold')
    labs=['Mean − ridge','Median − ridge','Median − scalar','Median − mean','Scalar − ridge']
    for k,cohort in enumerate(['GSE86216','GSE113873']):
        rows=[old[(old.cohort==cohort)&(old.method=='followup_ridge_nested')&(old.comparator=='mean_followup')].iloc[0]]
        for m,c in [('followup_ridge_nested','median_followup'),('shrunken_state_followup_nested','median_followup'),('mean_followup','median_followup'),('followup_ridge_nested','shrunken_state_followup_nested')]:rows.append(dif[(dif.cohort==cohort)&(dif.method==m)&(dif.comparator==c)].iloc[0])
        ax=fig.add_axes([.30+k*.35,.115,.305,.185])
        for j,(r,col) in enumerate(zip(rows,[C['teal'],C['teal'],C['orange'],C['ink'],C['purple']])):
            ax.errorbar(r.estimate,j,xerr=[[r.estimate-r.conditional_ci95_low],[r.conditional_ci95_high-r.estimate]],fmt='o',color=col,ms=4.5,lw=1.25,capsize=2)
        ax.axhspan(-.5,.5,color='#EAF3F5',zorder=0);ax.set_yticks(range(5),labs if k==0 else ['']*5);ax.set_ylim(4.6,-.6);ax.axvline(0,color=C['gray'],ls='--',lw=.8);ax.set_xlabel('Comparator MAE − method MAE',fontsize=8.2);ax.xaxis.set_major_locator(MaxNLocator(3));clean(ax)
    note(fig,'Small points: 20 dependent CV partitions; large points: means. Fixed method order.\nIntervals: 5,000 subject bootstraps conditional on fitted predictions; no multiplicity adjustment.\nAll contrasts are post hoc. Positive differences favor the subtracted method.',y=.018)
    save(fig,'Figure3_primary_benchmark')

def simulation():
    d=pd.read_csv(R/'simulation_summary_metrics.csv');sc=['independent_followup','scalar_persistence','off_diagonal_state','transport_shift'];names=['Independent follow-up','Scalar persistence','Off-diagonal signal','Transport shift'];ms=['mean_followup','median_followup','mean_change','persistence','scalar_inner_selected','ridge_inner_selected'];labs=['Mean follow-up','Median follow-up','Mean change','Persistence','Scalar (inner selected)','Ridge (inner selected)'];cols=[C['ink'],C['purple'],C['gray'],'#9AA7AE',C['orange'],C['teal']]
    fig=canvas(8.85);fig.text(.035,.978,'KNOWN-TRUTH SIMULATION',fontsize=11.5,fontweight='bold',color=C['purple'],va='top')
    fig.text(.32,.936,'Training n = 72',fontsize=10,fontweight='bold');fig.text(.66,.936,'Training n = 300',fontsize=10,fontweight='bold')
    for i,(s,name) in enumerate(zip(sc,names)):
        y=.716-i*.198
        fig.text(.035,y+.182,chr(65+i)+'  '+name,fontsize=10,fontweight='bold',color=C['blue'])
        sub=d[d.scenario==s];lo=min(.76,(sub.mc_mean-2*sub.mc_standard_error).min()-.025);hi=(sub.mc_mean+2*sub.mc_standard_error).max()+.025
        for j,n in enumerate([72,300]):
            ax=fig.add_axes([.31+j*.35,y,.31,.15])
            for k,m in enumerate(ms):
                r=d[(d.scenario==s)&(d.train_n==n)&(d.method==m)].iloc[0];ax.errorbar(r.mc_mean,k,xerr=2*r.mc_standard_error,fmt='o',color=cols[k],ms=4,capsize=2,lw=1.1)
            ax.set_yticks(range(6),labs if j==0 else ['']*6);ax.set_ylim(5.55,-.55);ax.set_xlim(lo,hi);ax.xaxis.set_major_locator(MaxNLocator(3));clean(ax)
            if i==3:ax.set_xlabel('Mean held-out MAE',labelpad=6)
    note(fig,'800 datasets: 4 scenarios × 2 sizes × 100 repetitions; 2,000 test observations each.\nBars: ±2 Monte Carlo standard errors; some bars are smaller than markers.\nSimulated outcomes, not patient confidence intervals or external clinical validation.',y=.015)
    save(fig,'Figure4_known_truth_simulation')

def coupling():
    d=pd.read_csv(E/'coupling_preserving_null.csv');p=d[d.cohort=='GSE86216'].pivot(index='null_repetition',columns='method',values='MAE')
    fig=canvas(8.1)
    title(fig,'A','Preserve baseline subtraction while breaking temporal pairing',.035,.975)
    ax=fig.add_axes([.035,.745,.93,.17]);off(ax)
    ax.text(.02,.79,'Δ = F − B',fontsize=23,color=C['blue'],fontweight='bold',va='top')
    ax.text(.36,.82,'Re-pair follow-up within train/test partitions.\nRetain −B and refit the fixed change-ridge model.\nCompare the original pairing with 200 re-pairings.',fontsize=9.3,va='top',linespacing=1.55)
    ax.text(.02,.10,'Scope: first original split; separate from the nested direct-F focal procedure.',fontsize=8.7,color=C['orange'],va='top')
    for k,(comp,name) in enumerate([('mean_delta','Versus mean change'),('mean_followup','Versus mean follow-up')]):
        left=.10+k*.49;title(fig,'BC'[k],name,left,.697)
        ax=fig.add_axes([left,.425,.385,.22]);diff=p[comp]-p['delta_ridge_fixed_0.01'];vals=diff[diff.index>=0];obs=diff.loc[-1]
        ax.hist(vals,bins=22,color='#E0B07B',edgecolor='white',lw=.45);ax.axvline(obs,color=C['blue'],lw=1.5);ax.axvline(0,color=C['gray'],ls='--',lw=.8);ax.set_xlabel('Comparator MAE − ridge MAE',fontsize=8.5,labelpad=6);ax.set_ylabel('Re-pairings' if k==0 else '');ax.xaxis.set_major_locator(MaxNLocator(4));clean(ax,'y')
        ax.text(.03,.96,f'Original: {obs:+.3f}',transform=ax.transAxes,fontsize=8.6,color=C['blue'],va='top',bbox=dict(facecolor='white',edgecolor='none',pad=2))
    title(fig,'D','Traditional change-target permutation asks a different question',.035,.345)
    original_mae=S['permutation_negative_control']['true_pooled_oof_MAE']
    ax=fig.add_axes([.105,.113,.54,.185]);vals=pd.read_csv(SRC/'results/permutation_pooled_mae.csv').pooled_oof_mae;ax.hist(vals,bins=22,color='#A9C3D2',edgecolor='white',lw=.45);ax.axvline(original_mae,color=C['blue'],lw=1.5);ax.set_xlabel('Complete five-fold pooled MAE',labelpad=5);ax.set_ylabel('Permutations');clean(ax,'y');ax.xaxis.set_major_locator(MaxNLocator(5))
    fig.text(.69,.273,f'Original pairing\nMAE = {original_mae:.3f}',fontsize=9.5,color=C['blue'],va='top',linespacing=1.4)
    fig.text(.69,.191,'Exploratory diagnostic\nOriginal fixed ridge\nNo biological null test',fontsize=8.7,color=C['gray'],va='top',linespacing=1.5)
    note(fig,'These controls diagnose mathematical coupling; they are not a formal test\nof the revised nested-ridge comparison or a treatment-effect null.',y=.023)
    save(fig,'Figure5_coupling_diagnostics')

def external():
    d=pd.read_csv(E/'external_GSE281841_direction_holdout.csv').set_index('module').reindex(MOD);c=pd.read_csv(E/'external_GSE281841_confounding.csv');sens=pd.read_csv(E/'external_GSE281841_restricted_sensitivities.csv')
    fig=canvas(8.85)
    title(fig,'A','GSE281841: cross-sectional severity, not model transport',.035,.978)
    ax=fig.add_axes([.035,.817,.93,.115]);off(ax)
    for x,w,h,b,col in [(.008,.30,'41 → 40 → 38 samples','Unscored sample excluded\nMixed-grade ID excluded','blue'),(.35,.30,'26 literal source IDs','18 high-grade (4–5)\n8 low-grade (0–1)','teal'),(.692,.30,'End-stage PAD series','Amputation specimens\nNo healthy-control group','orange')]:
        ax.add_patch(FancyBboxPatch((x,.02),w,.96,boxstyle='round,pad=.008,rounding_size=.012',fc='#F8FAFB',ec='#CBD8E0',lw=.75));ax.text(x+.02,.83,h,fontsize=8.9,fontweight='bold',color=C[col],va='top');ax.text(x+.02,.56,b,fontsize=8.7,va='top',linespacing=1.4)
    title(fig,'B','All eight contrasts, aggregated by literal source ID',.035,.78)
    ax=fig.add_axes([.235,.526,.69,.215])
    for i,(_,r) in enumerate(d.iterrows()):ax.errorbar(r.mean_high_minus_low,i,xerr=[[r.mean_high_minus_low-r.conditional_ci95_low],[r.conditional_ci95_high-r.mean_high_minus_low]],fmt='o',color=C['blue'],ms=4.5,lw=1.3,capsize=2)
    ax.set_yticks(range(8),LAB);ax.set_ylim(7.6,-.6);ax.axvline(0,color=C['gray'],ls='--',lw=.8);ax.set_xlabel('High − low (full-series standardized units)',labelpad=7);clean(ax)
    title(fig,'C','Artery composition is imbalanced',.035,.457)
    ax=fig.add_axes([.27,.255,.66,.165]);art=c[(c.variable=='artery')&(c.unit=='sample')];cats=art.category.unique().tolist();y=np.arange(len(cats))
    for z,g,col in [(-.18,'high_4_5',C['blue']),(.18,'low_0_1',C['orange'])]:
        p=art[art.group==g].set_index('category').reindex(cats);v=100*p.value/p.denominator;ax.barh(y+z,v,height=.30,color=col,label='High: 28 samples' if g=='high_4_5' else 'Low: 10 samples')
    yl=[a.capitalize().replace('Proximal anterior tibial','Prox. anterior tibial') for a in cats];ax.set_yticks(y,yl);ax.invert_yaxis();ax.set_xlim(0,65);ax.set_xlabel('Retained samples in each severity group (%)',labelpad=5);clean(ax);ax.legend(frameon=False,fontsize=8.3,loc='lower right')
    title(fig,'D','Restricted descriptions remain confounded',.035,.187)
    strata=[('artery','peroneal','Peroneal\n7 / 5'),('artery','posterior tibial','Post. tibial\n6 / 3'),('title_prefix_proxy','0092','Proxy 0092\n5 / 4'),('title_prefix_proxy','0521','Proxy 0521\n13 / 4')]
    arr=np.column_stack([sens[(sens.restriction==r)&(sens.stratum==s)].set_index('module').reindex(MOD).mean_high_minus_low.to_numpy() for r,s,_ in strata]);lim=max(2,np.abs(arr).max())
    # Horizontal compact heatmap retains all 32 contrasts and avoids tiny eight-row text.
    ax=fig.add_axes([.18,.075,.75,.075]);im=ax.imshow(arr.T,cmap=DIV,vmin=-lim,vmax=lim,aspect='auto');ax.set_xticks(range(8),ABBR);ax.set_yticks(range(4),['Peroneal 7/5','Post. tibial 6/3','Proxy 0092 5/4','Proxy 0521 13/4']);ax.tick_params(length=0,pad=3,labelsize=8)
    for i in range(4):
        for j in range(8):ax.text(j,i,f'{arr[j,i]:.2f}',ha='center',va='center',fontsize=8,color='white' if abs(arr[j,i])>lim*.65 else C['ink'])
    note(fig,'B: conditional 95% intervals; fixed normalization; no multiplicity correction.\nD: high/low ID counts; blue = negative, red = positive high−low standardized units.\nSource-title proxies are not verified batches.',y=.009)
    save(fig,'Figure6_external_severity')

def representation():
    coverage=pd.read_csv(R/'robustness_annotation_coverage.csv')
    summary=pd.read_csv(R/'robustness_annotation_summary.csv')
    ablation=pd.read_csv(R/'robustness_input_module_ablation_summary.csv')
    fig=canvas(8.6)
    title(fig,'A','Measured changes under two annotation rules',.035,.975)
    schemes=[('module_preferred','Frozen rule',C['blue'],-.12),('unique_symbol_only','Unique-symbol rule',C['orange'],.12)]
    for j,(field,heading,limits) in enumerate([
        ('retained_target_probes_in_platform','Retained target probes',(120,315)),
        ('union_observed_genes','Retained module-union genes',(81.5,85.5))]):
        ax=fig.add_axes([.22+j*.47,.785,.29 if j==0 else .26,.12])
        ax.set_title(heading,fontsize=9,pad=7)
        for scheme,lab,col,off_y in schemes:
            vals=[]
            for i,cohort in enumerate(['GSE86216','GSE113873']):
                rows=coverage[(coverage.cohort==cohort)&(coverage.mapping_scheme==scheme)]
                assert rows[field].nunique()==1
                value=int(rows.iloc[0][field]);vals.append(value)
                ax.text(value+(limits[1]-limits[0])*.025,i+off_y,str(value),va='center',fontsize=8,color=col)
            ax.scatter(vals,np.arange(2)+off_y,s=23,color=col,label=lab,zorder=3)
        ax.set_yticks([0,1],['Statin','PAD'] if j==0 else ['','']);ax.set_ylim(1.4,-.4);ax.set_xlim(limits);ax.xaxis.set_major_locator(MaxNLocator(4,integer=True));clean(ax)
        if j==0:fig.legend(*ax.get_legend_handles_labels(),frameon=False,fontsize=8.3,ncol=2,loc='upper center',bbox_to_anchor=(.55,.753))
    fig.text(.035,.695,'Statin: identity control. PAD: oxidative-stress coverage changes from 9 to 8 genes.',fontsize=8.3,color=C['gray'])
    title(fig,'B','Ridge comparisons within each annotation scheme',.035,.65)
    row_defs=[('module_preferred','mean_followup','Frozen: mean − ridge'),('unique_symbol_only','mean_followup','Unique: mean − ridge'),('module_preferred','median_followup','Frozen: median − ridge'),('unique_symbol_only','median_followup','Unique: median − ridge')]
    for k,cohort in enumerate(['GSE86216','GSE113873']):
        ax=fig.add_axes([.27+k*.40,.445,.28,.145]);ax.set_title('Statin · n = 72' if k==0 else 'PAD · n = 14',fontsize=9,pad=8)
        for i,(scheme,comp,lab) in enumerate(row_defs):
            rows=summary[(summary.cohort==cohort)&(summary.mapping_scheme==scheme)&(summary.method=='followup_ridge_nested')&(summary.comparator==comp)]
            assert len(rows)==1
            row=rows.iloc[0];value=row.within_scheme_MAE_improvement
            ax.errorbar(value,i,xerr=[[value-row.conditional_subject_ci95_low],[row.conditional_subject_ci95_high-value]],fmt='o',ms=3.5,lw=1.1,capsize=2,color=C['blue'] if scheme=='module_preferred' else C['orange'])
        ax.set_yticks(range(4),[r[2] for r in row_defs] if k==0 else ['']*4,fontsize=8);ax.set_ylim(3.6,-.6);ax.set_xlim(-.021,.028);ax.axvline(0,color=C['gray'],lw=.8,ls='--');ax.set_xlabel('Comparator MAE − ridge MAE',fontsize=8.2);ax.set_xticks([-.02,0,.02]);clean(ax)
    for k,cohort in enumerate(['GSE86216','GSE113873']):
        title(fig,'CD'[k],('Statin' if k==0 else 'PAD')+' input deletion',.035+k*.50,.353)
        ax=fig.add_axes([.225+k*.465,.125,.285,.18]);q=ablation[(ablation.cohort==cohort)&(ablation.comparator=='followup_ridge_nested')].set_index('removed_input_module').reindex(MOD)
        assert len(q)==8 and q.complete_inner_tuning_rerun.all() and (q.retained_target_modules==8).all()
        ax.scatter(q.comparator_MAE_minus_ablated_MAE,np.arange(8),s=24,color=C['teal'] if k==0 else C['orange'],zorder=3)
        ax.set_yticks(range(8),LAB if k==0 else ABBR,fontsize=8);ax.set_ylim(7.6,-.6);ax.set_xlim(-.005,.006);ax.set_xticks([-.004,0,.004]);ax.axvline(0,color=C['gray'],lw=.8,ls='--');ax.set_xlabel('Full MAE − ablated MAE',fontsize=8.2);clean(ax)
    note(fig,'B: conditional 95% intervals; fixed predictions; no refitting or multiplicity adjustment.\nC–D: all 8 targets retained; inner tuning rerun; descriptive point estimates.\nNegative values mean deletion increases error. No biological-necessity inference.',y=.015)
    save(fig,'Figure7_representation')

def robustness():
    reg=pd.read_csv(E/'regularization_sensitivity_summary.csv');size=pd.read_csv(E/'sample_size_sensitivity_summary.csv')
    fig=canvas(7.3)
    fig.text(.035,.975,'Post hoc numerical and training-subset sensitivities',fontsize=11,fontweight='bold',color=C['blue'],va='top')
    for k,cohort in enumerate(['GSE86216','GSE113873']):
        ax=fig.add_axes([.12+k*.47,.585,.355,.27]);ax.set_title('AB'[k]+'  '+('Statin' if k==0 else 'PAD')+' regularization',loc='left',fontsize=9,fontweight='bold',pad=7)
        q=reg[(reg.cohort==cohort)&(reg.target=='followup')]
        for flag,col,lab in [(False,C['gray'],'Unscaled module features'),(True,C['teal'],'Scaled module features')]:
            v=q[q.normalized_module_features==flag].sort_values('lambda');ax.plot(v['lambda'],v.MAE,'-o',ms=3,lw=1.1,color=col,label=lab)
        ax.set_xscale('log');ax.set_xlabel('Ridge penalty λ');ax.set_ylabel('Held-out follow-up MAE' if k==0 else '');clean(ax,'y')
        if k==0:fig.legend(*ax.get_legend_handles_labels(),frameon=False,fontsize=8,ncol=2,loc='upper center',bbox_to_anchor=(.55,.935))
        ax=fig.add_axes([.12+k*.47,.255,.355,.22]);ax.set_title('CD'[k]+'  '+('Statin' if k==0 else 'PAD')+' training subsets',loc='left',fontsize=9,fontweight='bold',pad=7)
        q=size[size.cohort==cohort];sizes=sorted(q.n_train.unique());positions=np.arange(len(sizes));folds=q[q.method=='mean_followup'].set_index('n_train').reindex(sizes).n_folds
        for j,(method,col,lab) in enumerate([('delta_ridge_fixed_0.01',C['blue'],'Fixed change ridge'),('mean_followup',C['ink'],'Mean follow-up'),('followup_ridge_nested',C['teal'],'Nested follow-up ridge'),('shrunken_state_followup_nested',C['orange'],'Scalar shrinkage')]):
            v=q[q.method==method].set_index('n_train').reindex(sizes)
            ax.scatter(positions+(j-1.5)*.07,v.MAE,s=19,color=col,label=lab,zorder=3)
        ax.axvspan(len(sizes)-2.5,len(sizes)-.5,color='#F0F3F6',zorder=0)
        ax.set_xticks(positions,[f'{n}\n({int(f)})' for n,f in zip(sizes,folds)],fontsize=8);ax.set_xlabel('Training subjects (number of outer folds)',fontsize=8.2);ax.set_ylabel('Held-out follow-up MAE' if k==0 else '');clean(ax,'y')
        if k==1:fig.legend(*ax.get_legend_handles_labels(),frameon=False,fontsize=8,ncol=2,loc='upper center',bbox_to_anchor=(.55,.17))
    note(fig,'C–D: points are unconnected; shaded endpoint sizes use different test-fold sets.\nFive outer repetitions; folds are dependent. Endpoint changes are not pure size effects.\nNo confidence intervals or primary-method reselection are inferred.',y=.014)
    save(fig,'Figure8_robustness')

if __name__=='__main__':
    framework();atlas();primary();simulation();coupling();external();representation();robustness()
    (OUT/'figure_manifest.json').write_text(json.dumps(MAN,indent=2),encoding='utf-8')
    print(json.dumps([{'figure':m['figure'],'width_mm':m['nominal_width_mm'],'height_mm':m['height_mm'],'minimum_font_pt':m['minimum_font_pt']} for m in MAN],indent=2))
```
