## Supplementary material for "VascFlux-GEO: Comparator selection and auditability in vascular transcriptome module prediction": Analysis code and aggregate results: redraw_supplement_figures.py

```
"""Redesign seven supplementary diagnostics at final 170-mm publication width.
Reads only previously executed CSV results. No new numerical experiments.
"""
from pathlib import Path
import json
import numpy as np
import pandas as pd
import matplotlib
matplotlib.use('Agg')
import matplotlib.pyplot as plt
from matplotlib.ticker import MaxNLocator
import redraw_main_figures as p
SRC,OUT,E=p.SRC,p.OUT,p.E
C,LAB,MOD,ABBR=p.C,p.LAB,p.MOD,p.ABBR
METHODS=['mean_delta','mean_followup','persistence','delta_ridge_fixed_0.01','followup_ridge_nested','pca2_followup_nested','shrunken_state_followup_nested']
ML=['Mean change','Mean follow-up','Persistence','Fixed change ridge','Nested follow-up ridge','PCA-2 + nested ridge','Scalar shrinkage']
GROUPS=['statin','exercise_only','no_exercise_only','carotid_plaque_minus_intact','PAD_PBMC_minus_control']
GL=['Statin blood','Exercise muscle','No-exercise muscle','Carotid plaque','PAD PBMC']
SHORT=['Statin','Exercise','No exercise','Carotid','PAD PBMC']

def header(fig,text):
    fig.text(.035,.975,text,fontsize=11,fontweight='bold',color=C['blue'],va='top')
def label(ax,letter,title):
    ax.set_title(letter+'  '+title,loc='left',pad=7,fontsize=9,fontweight='bold')
def save(fig,name):
    assert fig.get_figheight()*25.4<=185
    p.save(fig,name)

def scatter(cohort,name):
    d=pd.read_csv(E/'oof_predictions.csv')
    d=d[(d.cohort==cohort)&(d['repeat']==0)&(d.method=='followup_ridge_nested')]
    fig=p.canvas(7.2);header(fig,cohort+' · nested-ridge module predictions')
    for i,m in enumerate(MOD):
        col=i%2;row=i//2
        ax=fig.add_axes([.12+col*.47,.756-row*.207,.355,.142]);q=d[d.module==m]
        ax.scatter(q.observed_followup,q.predicted_followup,s=12,alpha=.60,color=C['blue'] if cohort=='GSE86216' else C['orange'],edgecolors='none')
        lo=min(q.observed_followup.min(),q.predicted_followup.min());hi=max(q.observed_followup.max(),q.predicted_followup.max());span=hi-lo;lo-=span*.06;hi+=span*.06
        ax.plot([lo,hi],[lo,hi],ls='--',lw=.8,color=C['gray']);ax.set(xlim=(lo,hi),ylim=(lo,hi))
        label(ax,chr(65+i),LAB[i]);ax.text(.03,.96,f'n={len(q)}; MAE={q.absolute_error.mean():.3f}',transform=ax.transAxes,fontsize=8,va='top')
        ax.xaxis.set_major_locator(MaxNLocator(3));ax.yaxis.set_major_locator(MaxNLocator(3));ax.tick_params(pad=2,length=2.5)
    fig.text(.035,.55,'Predicted follow-up',rotation=90,fontsize=9,va='center')
    fig.text(.51,.067,'Observed follow-up',fontsize=9,ha='center')
    p.note(fig,'First complete outer five-fold partition; one point per held-out subject.\nDashed lines: identity. Values use fold-fitted gene scaling.',y=.014)
    save(fig,name)

def outcome():
    rep=pd.read_csv(E/'repeat_metrics.csv');o=pd.read_csv(SRC/'results/oof_subject_errors.csv')
    fig=p.canvas(6.9);header(fig,'Outcome definitions and ridge comparisons')
    for k,c in enumerate(['GSE86216','GSE113873']):
        ax=fig.add_axes([.30+k*.35,.57,.30,.31]);q=rep[rep.cohort==c].groupby('method')[['pooled_delta_R2','pooled_followup_R2']].mean().reindex(METHODS)
        ax.imshow(q,vmin=-1,vmax=1,cmap=p.DIV,aspect='auto');ax.set_yticks(range(7),ML if k==0 else ['']*7);ax.set_xticks([0,1],['Change R²','Follow-up R²']);ax.tick_params(length=0)
        label(ax,'AB'[k],c)
        for i in range(7):
            for j in range(2):ax.text(j,i,f'{q.iloc[i,j]:.2f}',ha='center',va='center',fontsize=8.5,color='white' if abs(q.iloc[i,j])>.65 else C['ink'])
    d=o[o.cohort=='GSE86216_statin'];a=abs(d.observed_followup_level-d.direct_followup_level_prediction);base=d.observed_followup_level-d.observed_delta;b=abs(d.observed_delta-(d.direct_followup_level_prediction-base))
    ax=fig.add_axes([.13,.16,.34,.27]);label(ax,'C','Identical absolute errors');ax.scatter(a,b,s=5,alpha=.25,color=C['teal'],rasterized=True);lim=max(a.max(),b.max())*1.04;ax.plot([0,lim],[0,lim],ls='--',color=C['gray'],lw=.8);ax.set_xlabel('Follow-up error');ax.set_ylabel('Implied-change error');ax.xaxis.set_major_locator(MaxNLocator(3));ax.yaxis.set_major_locator(MaxNLocator(3))
    ax=fig.add_axes([.63,.16,.32,.27]);label(ax,'D','Approximate ridge equivalence');implied=d.direct_followup_level_prediction-base;ax.scatter(d.delta_ridge_prediction,implied,s=5,alpha=.25,color=C['purple'],rasterized=True);lo=min(d.delta_ridge_prediction.min(),implied.min());hi=max(d.delta_ridge_prediction.max(),implied.max());ax.plot([lo,hi],[lo,hi],ls='--',color=C['gray'],lw=.8);ax.set_xlabel('Fixed change ridge');ax.set_ylabel('Direct follow-up − baseline',fontsize=8.4);ax.xaxis.set_major_locator(MaxNLocator(3));ax.yaxis.set_major_locator(MaxNLocator(3))
    maxdiff=float(np.max(np.abs(d.delta_ridge_prediction-implied)))
    ax.text(.03,.97,f'Max. difference = {maxdiff:.6f}',transform=ax.transAxes,fontsize=8,va='top')
    p.note(fig,'A–B: 20 repeated partitions; R² colors saturate at ±1.\nC: exact residual identity. D: near-equivalence at λ = 0.01, not exact identity.',y=.015)
    save(fig,'FigureS2_outcome_identity')

def tuning():
    d=pd.read_csv(E/'nested_tuning.csv');d=d[d.selected.astype(bool)]
    fig=p.canvas(6.3);header(fig,'Inner-fold tuning selections across repeated partitions')
    for i,(c,t) in enumerate([(c,t) for c in ['GSE86216','GSE113873'] for t in ['followup','shrunken']]):
        ax=fig.add_axes([.12+(i%2)*.47,.57-(i//2)*.40,.355,.28]);label(ax,chr(65+i),('Statin' if c=='GSE86216' else 'PAD')+(' ridge λ' if t=='followup' else ' scalar shrinkage'))
        q=d[(d.cohort==c)&(d.target_model==t)];v=q.candidate.value_counts().sort_index();assert v.sum()==100
        ax.bar(range(len(v)),v.values,color=C['teal'] if t=='followup' else C['orange'],width=.65);ax.set_xticks(range(len(v)),[f'{x:g}' for x in v.index],rotation=35 if len(v)>5 else 0,ha='right' if len(v)>5 else 'center');ax.set_ylim(0,110);ax.set_ylabel('Outer-fit selections' if i%2==0 else '');ax.set_xlabel('Selected candidate');p.clean(ax,'y')
        for j,x in enumerate(v.values):ax.text(j,x+2,str(x),ha='center',fontsize=8)
    p.note(fig,'100 outer fits per cohort and model. Fits share subjects; frequencies are diagnostic.\nCandidate selection is confined to the training folds.',y=.014)
    save(fig,'FigureS4_tuning_choices')

def directions():
    d=pd.read_csv(E/'corrected_group_module_directions.csv');fig=p.canvas(7.2);header(fig,'Descriptive directions across five biological contexts')
    for i,g in enumerate(GROUPS):
        ax=fig.add_axes([.24+(i%2)*.43,.705-(i//2)*.282,.26,.185]);label(ax,chr(65+i),GL[i]+'\n'+['72 pairs','5 pairs','9 pairs','32 tissue pairs','19 PAD / 18 controls'][i]);q=d[d.group==g].set_index('module').reindex(MOD)
        for j,(_,r) in enumerate(q.iterrows()):ax.errorbar(r.mean_direction,j,xerr=[[r.mean_direction-r.conditional_ci95_low],[r.conditional_ci95_high-r.mean_direction]],fmt='o',color=C['blue'],ms=3,lw=.9,capsize=1.5)
        ax.set_yticks(range(8),LAB if i%2==0 else ['']*8);ax.set_ylim(7.6,-.6);ax.axvline(0,color=C['gray'],ls='--',lw=.8);ax.xaxis.set_major_locator(MaxNLocator(3));p.clean(ax)
    fig.text(.58,.28,'All eight modules retained\n\n5,000 subject bootstrap draws\nFixed full-series normalization\n\nUnadjusted exploratory intervals\nNo causal or transport inference',fontsize=8.6,color=C['gray'],va='top',linespacing=1.45)
    fig.text(.51,.072,'Direction (full-series standardized units)',fontsize=9,ha='center')
    p.note(fig,'Paired cohorts resample whole subjects; PAD/control PBMC groups resample separately.\nIntervals are conditional on normalization and modules; no multiplicity adjustment.',y=.014)
    save(fig,'FigureS5_direction_intervals')

def controls():
    d=pd.read_csv(E/'injected_fault_controls.csv');tests=d.test.drop_duplicates().tolist()
    labels=['Clean future-target mutation','All-visit scaler leakage','Duplicate subject','Reused sample','Cross-subject mispairing','Nonfinite training expression','Train/test subject overlap']
    arr=[]
    for t in tests:
        row=[]
        for c in ['GSE86216','GSE113873']:
            r=d[(d.test==t)&(d.cohort==c)].iloc[0];row.extend([int(r.expected_detection),int(r.detected),int(r.passed)])
        arr.append(row)
    arr=np.array(arr);fig=p.canvas(6.4);header(fig,'Engineered control outcomes and positive-control effect')
    ax=fig.add_axes([.405,.535,.55,.345]);label(ax,'A','Statin / PAD control checks');ax.imshow(arr,cmap='Blues',vmin=0,vmax=1,aspect='auto');ax.set_yticks(range(7),labels);ax.set_xticks(range(6),['Expect','Detect','Pass']*2);ax.tick_params(length=0);ax.axvline(2.5,color='white',lw=3)
    for i in range(7):
        for j in range(6):ax.text(j,i,'Yes' if arr[i,j] else 'No',ha='center',va='center',fontsize=8.2,color='white' if arr[i,j] else C['ink'])
    fig.text(.49,.49,'Statin',fontsize=9,fontweight='bold');fig.text(.78,.49,'PAD',fontsize=9,fontweight='bold')
    ax=fig.add_axes([.18,.15,.40,.24]);label(ax,'B','Leaky-scaler positive control');q=d[d.test=='all_visit_scaler_future_leakage'];ax.bar(range(2),q.observed_statistic,color=[C['blue'],C['orange']],width=.5);ax.set_xticks([0,1],['Statin','PAD']);ax.set_ylim(0,2.15);ax.set_ylabel('Maximum baseline-state change',fontsize=8.4);p.clean(ax,'y')
    for i,v in enumerate(q.observed_statistic):ax.text(i,v+.06,f'{v:.3f}',ha='center',fontsize=8.5)
    fig.text(.66,.35,'14 finite control cases\n\n2 clean controls invariant\n12 injected faults detected\n\nNot general sensitivity\nor biological validity',fontsize=8.8,va='top',linespacing=1.4,color=C['gray'])
    p.note(fig,'Clean mutation checks: first fold of the first repetition; tuning held fixed.\nPositive control deliberately contaminates scaling with future expression.',y=.014)
    save(fig,'FigureS6_control_details')

def crossbed():
    dr=pd.read_csv(E/'corrected_group_module_directions.csv')
    cr=pd.read_csv(E/'corrected_cross_bed_concordance.csv')
    assert len(cr)==10 and (cr.conditional_ci95_low<0).all() and (cr.conditional_ci95_high>0).all()
    fig=p.canvas(7.2);header(fig,'Cross-context comparisons and their uncertainty')
    ax=fig.add_axes([.31,.52,.64,.34]);label(ax,'A','All ten direction cosines')
    for i,(_,r) in enumerate(cr.iterrows()):
        ax.errorbar(r.cosine,i,xerr=[[r.cosine-r.conditional_ci95_low],[r.conditional_ci95_high-r.cosine]],fmt='o',color=C['blue'],ms=4,lw=1.1,capsize=2)
    ax.set_yticks(range(10),[f'{SHORT[GROUPS.index(r.group_a)]} / {SHORT[GROUPS.index(r.group_b)]}' for _,r in cr.iterrows()],fontsize=8.5)
    ax.set_ylim(9.6,-.6);ax.set_xlim(-1.05,1.05);ax.axvline(0,color=C['gray'],ls='--',lw=.8);ax.set_xlabel('Cosine');p.clean(ax)
    q=dr[dr.group=='exercise_minus_noexercise_paired_change'].set_index('module').reindex(MOD)
    ax=fig.add_axes([.31,.15,.64,.23]);label(ax,'B','Exercise minus no exercise: 5 versus 9 pairs')
    for i,(_,r) in enumerate(q.iterrows()):
        ax.errorbar(r.mean_direction,i,xerr=[[r.mean_direction-r.conditional_ci95_low],[r.conditional_ci95_high-r.mean_direction]],fmt='o',color=C['orange'],ms=4,lw=1.1,capsize=2)
    ax.set_yticks(range(8),LAB);ax.set_ylim(7.6,-.6);ax.axvline(0,color=C['gray'],ls='--',lw=.8)
    ax.set_xlabel('Difference in paired changes (full-series standardized units)',fontsize=8.3);ax.xaxis.set_major_locator(MaxNLocator(4));p.clean(ax)
    p.note(fig,'Conditional 95% intervals: 5,000 subject bootstraps; normalization and modules fixed.\nAll ten cosine intervals include zero. Module directions are in Figure S5.\nExploratory contrasts; no transport, causal-treatment or device inference.',y=.014)
    save(fig,'FigureS7_cross_bed')

if __name__=='__main__':
    scatter('GSE86216','FigureS1_statin_module_predictions');outcome();scatter('GSE113873','FigureS3_PAD_module_predictions');tuning();directions();controls();crossbed()
    (OUT/'supplementary_figure_manifest.json').write_text(json.dumps(p.MAN,indent=2),encoding='utf-8')
    print(json.dumps([{'figure':m['figure'],'height_mm':m['height_mm'],'minimum_font_pt':m['minimum_font_pt']} for m in p.MAN],indent=2))
```
