## Supplementary material for "VascFlux-GEO: Comparator selection and auditability in vascular transcriptome module prediction": Analysis code and aggregate results: README.txt

VascFlux-GEO: code and aggregate submission attachment

Prepared for submission and author review on 25 October 2026. This archive
does not establish final submission or public posting; portal status is
recorded separately.
CC BY manuscript distribution was authorized by the user on 25 October 2026.
This manuscript permission does not independently license software or third-party
GEO data. No independent software licence is granted by this archive.
Actual portal licensing and submission actions are recorded separately.

This subset contains unchanged analysis/acquisition/verification code, numerical
dependency records, frozen module definitions, 35 result CSVs (34 current
aggregate or synthetic result tables plus the historical permutation diagnostic),
the current plotting code, a study-level data registry, and interpretation notes. It deliberately omits source expression/platform payloads, all individual
records, patient/sample identifiers, individual ages, linked folds and individual
predictions. The complete saved-output audit package is a separate private file.

Scientific scope
The manuscript compares within-cohort predictions in 72 rosuvastatin pairs and
14 PAD pairs, plus separate descriptive sources. Do not sum heterogeneous source
counts as an independently confirmed patient total. Simulations are synthetic.
All extensions are post hoc. Conditional bootstrap fields describe fixed-output
resampling ranges; their nominal level is not validated population coverage.
See reproducibility/STATISTICAL_DATA_DICTIONARY.md and dataset_registry.tsv.

Folder layout and numerical rebuilding
Keep reproducibility/ and figures/ as siblings. Use Python 3.12.14 for the recorded
numerical environment. From the reproducibility directory:

  python -m pip install -r requirements-analysis.txt
  python inputs/download_geo.py --out geo_cache
  python inputs/acquire_canonical_platforms.py --source-cache geo_cache --out canonical_cache
  python reproduce.py --full --data-dir canonical_cache --source-manifest canonical_cache/canonical_manifest.json

Acquisition requires the network and verifies seven exact complete files plus
three complete normalized platform tables. The two official family streams are
checked through the complete first platform table, not the entire family archive.
The former truncated GPL570 cache is not a permitted input. Future official data
changes can make acquisition fail; do not replace expected hashes to force success.
Use official source conditions and citations; public visibility is not a new
redistribution licence for third-party data.

After a successful full numerical rebuild, python reproduce.py --verify-only
checks the locally generated individual and aggregate outputs. The distributed
aggregate subset ALONE does not satisfy --verify-only and is not claimed to do so.
Numerical rebuilding creates individual-level local files; those generated files
are NOT part of this submission attachment and should not be uploaded with it.
The optional historical first-partition prediction comparison is skipped if the
excluded original results/oof_subject_errors.csv anchor is absent.

Plotting and historical inputs
The plotting environment is separate: Python 3.11.9 and requirements-figures.txt.
Current plotting code is supplied without modifying its analysis logic. Rebuild
the numerical stages before running plots. Some figures require those generated
JSON summaries and per-person predictions. Supplementary Figure S2 additionally
requires the original results/oof_subject_errors.csv anchor, which is excluded
from this attachment. Consequently --full --figures and complete all-15-figure
redrawing are not promised from this subset alone. The private review package
contains that historical anchor and records the full figure reconstruction.

reproducibility/results/summary.json is an EXPLICIT DERIVED AGGREGATE SUBSET of
the frozen historical summary, not the original byte-identical summary. It keeps
only module_names, dataset sample/coverage counts, paired_counts and the original
permutation diagnostic needed by the plotting code. It excludes historical
cross-context estimates superseded by the complete GPL570 correction. Its own
_attachment_derivation field and ATTACHMENT_MANIFEST.json record the original
source hash and selected keys. The permutation_pooled_mae.csv is an unchanged
200-repetition historical diagnostic aggregate, not a calibrated hypothesis test.
Current corrected cross-context results are in results_extended/.

Validation boundary
The attachment audit checks a closed file list, hashes, CSV fields, known
identifier patterns and Python syntax/import dependencies. Recorded smoke tests
are listed in the separate author audit. Passing these checks is not a complete
de-identification certification or proof of scientific generalization. No online
availability, DOI, portal preview or final submission is implied by this folder.
