## Supplementary material for "VascFlux-GEO: Comparator selection and auditability in vascular transcriptome module prediction": Analysis code and aggregate results: acquire_canonical_platforms.py

```
"""Validate fresh official platform tables, then build a canonical analysis cache.

The two family URLs and GPL570 text URL are streamed through their first complete platform table.
This is deliberately NOT a full family-archive download or integrity test. Normal
TLS validation is mandatory. Whole normalized table bytes and the actual analysis
probe mapping must match the recorded expected digests before installation.
No third-party source table or derived source-data snapshot is bundled here.
The remaining seven inputs are copied from a separately hash-verified local cache.
"""
import argparse
import contextlib
import gzip
import hashlib
import json
import os
import shutil
import ssl
import sys
import tempfile
import urllib.request
from datetime import datetime, timezone
from pathlib import Path

ROOT = Path(__file__).resolve().parents[1]
sys.path.insert(0, str(ROOT))
from run_extended_benchmarks import read_platform
from download_geo import require_official_https, sha, verify
from validate_inputs import inspect_table


def write_soft(table, output):
    with output.open('wb') as raw:
        stream = gzip.GzipFile(fileobj=raw, mode='wb', mtime=0, filename='') if output.suffix == '.gz' else contextlib.nullcontext(raw)
        with stream as out:
            out.write(b'!platform_table_begin\n')
            out.write(table)
            out.write(b'!platform_table_end\n')


def mapping_digest(path):
    mapping, ambiguous = read_platform(path)
    blob = json.dumps(mapping, sort_keys=True, separators=(',', ':')).encode()
    return {'mapped_probe_count': len(mapping), 'ambiguous_probe_count': ambiguous,
            'probe_to_module_gene_sha256': hashlib.sha256(blob).hexdigest()}


def validate_expected_table(table, evidence, expected, accession):
    """Require the entire normalized table, not only mapped genes, to match."""
    if hashlib.sha256(table).hexdigest() != expected['table_uncompressed_sha256']:
        raise ValueError(f'Fresh official table differs from archived table: {accession}; fail closed.')
    if len(table) != expected['table_uncompressed_bytes'] or evidence['table_data_rows'] != expected['table_data_rows']:
        raise ValueError(f'Fresh official table size or row count differs: {accession}')


def fetch_table(url, timeout, compressed=True):
    require_official_https(url)
    req = urllib.request.Request(url, headers={'User-Agent': 'VascFlux-GEO-reproducibility/1.2', 'Accept-Encoding': 'identity'})
    begin = end = False
    lines = []
    with urllib.request.urlopen(req, timeout=timeout, context=ssl.create_default_context()) as response:
        require_official_https(response.geturl())
        if 'text/html' in response.headers.get('Content-Type', '').lower():
            raise ValueError('Official endpoint returned HTML; no input installed.')
        observed_length = response.headers.get('Content-Length')
        last_modified = response.headers.get('Last-Modified')
        etag = response.headers.get('ETag')
        with gzip.GzipFile(fileobj=response, mode='rb') if compressed else contextlib.nullcontext(response) as stream:
            for line in stream:
                if line.startswith(b'!platform_table_begin'):
                    begin = True
                    continue
                if begin and line.startswith(b'!platform_table_end'):
                    end = True
                    break
                if begin:
                    lines.append(line.rstrip(b'\r\n') + b'\n')
                    if len(lines) > 1000000:
                        raise ValueError('Platform row safety limit exceeded.')
        # Exiting closes the response immediately after table completion, including
        # at most the gzip reader's bounded read-ahead; family trailer is not read.
    if not (begin and end and len(lines) > 1):
        raise ValueError('No complete platform table received.')
    return b''.join(lines), {'platform_table_begin_found': begin,
        'platform_table_end_found': end, 'table_data_rows': len(lines)-1,
        'remote_export_advertised_bytes': int(observed_length) if observed_length else None,
        'remote_last_modified': last_modified, 'remote_etag': etag,
        'retrieved_utc': datetime.now(timezone.utc).isoformat(),
        'whole_family_downloaded_or_integrity_verified': False,
        'TLS_certificate_verification': True}


def main():
    parser = argparse.ArgumentParser(description=__doc__)
    parser.add_argument('--source-cache', type=Path, required=True,
                        help='Hash-verified source for the OTHER seven unchanged inputs')
    parser.add_argument('--out', type=Path, default=ROOT / 'canonical_cache')
    parser.add_argument('--timeout', type=float, default=120)
    parser.add_argument('--report', type=Path, default=ROOT / 'provenance/canonical_acquisition.json')
    args = parser.parse_args()
    args.out.mkdir(parents=True, exist_ok=True)
    args.report.parent.mkdir(parents=True, exist_ok=True)
    public = json.loads((ROOT / 'inputs/source_manifest_public.json').read_text(encoding='utf-8'))
    snapshots = json.loads((ROOT / 'inputs/canonical_platform_expected.json').read_text(encoding='utf-8'))['platforms']
    report = {'status': 'running', 'utc': datetime.now(timezone.utc).isoformat(),
        'scope': 'Three fresh official complete first-platform-table validations plus seven hash-verified local copies; not ten fresh full downloads.',
        'platforms': {}, 'unchanged_inputs': {}, 'full_family_downloads': 0}
    canonical = {}
    try:
        with tempfile.TemporaryDirectory(prefix='canonical-platform-', dir=args.out) as tmp:
            temp = Path(tmp)
            for accession, rec in snapshots.items():
                name = rec['source_filename']
                fresh_table, evidence = fetch_table(public[name]['url'], args.timeout, rec.get('remote_format', 'gzip') == 'gzip')
                table_hash = hashlib.sha256(fresh_table).hexdigest()
                evidence.update({'url': public[name]['url'], 'table_uncompressed_bytes': len(fresh_table),
                    'table_uncompressed_sha256': table_hash,
                    'matches_archived_complete_table': table_hash == rec['table_uncompressed_sha256']})
                report['platforms'][accession] = evidence
                validate_expected_table(fresh_table, evidence, rec, accession)
                fresh_soft = temp / name
                write_soft(fresh_table, fresh_soft)
                structure = inspect_table(fresh_soft)
                evidence['structure'] = structure
                expected, actual = rec['expected_analysis_mapping'], mapping_digest(fresh_soft)
                evidence.update({'archived_mapping': expected, 'fresh_mapping': actual,
                                 'analysis_mapping_equal': expected == actual})
                if expected != actual:
                    raise ValueError(f'Actual analysis mapping differs: {accession}')
                # Install a deterministic SOFT wrapper from the fresh table with verified digests.
                target = args.out / name
                os.replace(fresh_soft, target)
                canonical[name] = {'filename': name, 'bytes': target.stat().st_size,
                    'sha256': sha(target), 'url': public[name]['url'], 'format': 'gzip' if name.endswith('.gz') else 'canonical_platform_soft',
                    'input_kind': 'derived_complete_first_platform_table_SOFT_not_family_archive',
                    'transformation': 'complete normalized table matching frozen digest; deterministic begin/end SOFT wrapper; gzip only for .gz names',
                    'source_table_sha256': table_hash,
                    'normalized_table_sha256': table_hash,
                    'expected_table_rows': rec['table_data_rows'],
                    'fresh_official_complete_table_and_analysis_mapping_equal': True}
            for name, rec in public.items():
                if name in canonical:
                    continue
                source = args.source_cache / name
                verified = verify(source, rec)
                target = args.out / name
                if target.resolve() == source.resolve():
                    raise ValueError('Canonical output must be separate from the frozen source cache.')
                shutil.copyfile(source, target)
                verify(target, rec)
                canonical[name] = dict(rec, filename=name, acquisition_this_run='copied_from_hash_verified_local_cache_not_fresh_download')
                report['unchanged_inputs'][name] = dict(verified, acquisition='verified_local_copy')
        (args.out / 'canonical_manifest.json').write_text(json.dumps(canonical, indent=2)+'\n', encoding='utf-8')
        report.update(status='passed', fresh_complete_platform_tables_compared=len(snapshots),
                      verified_local_inputs_copied=len(report['unchanged_inputs']),
                      canonical_manifest_sha256=sha(args.out / 'canonical_manifest.json'))
    except Exception as exc:
        report.update(status='failed_closed', error=f'{type(exc).__name__}: {exc}')
        # No ready manifest is left for a partial or failed acquisition.
        (args.out / 'canonical_manifest.json').unlink(missing_ok=True)
        raise
    finally:
        args.report.write_text(json.dumps(report, indent=2)+'\n', encoding='utf-8')
        print(json.dumps(report, indent=2), flush=True)


if __name__ == '__main__':
    main()
```
