## Supplementary material for "VascFlux-GEO: Comparator selection and auditability in vascular transcriptome module prediction": Analysis code and aggregate results: download_geo.py

```
"""Obtain frozen official GEO inputs, or verify a cache without networking.

HTTPS certificate verification uses Python's default SSL context. A newly fetched
file is installed only after its size, format signature, and SHA-256 all match.
No source normalization is performed: the manifest describes exact input bytes.
"""
import argparse
import hashlib
import json
import os
import platform
import ssl
import tempfile
import urllib.error
import urllib.parse
import urllib.request
import sys
from datetime import datetime, timezone
from pathlib import Path

MANIFEST = Path(__file__).with_name("source_manifest_public.json")
sys.path.insert(0, str(Path(__file__).resolve().parents[1]))
from validate_inputs import inspect_table
OFFICIAL_HOSTS = {"www.ncbi.nlm.nih.gov", "ftp.ncbi.nlm.nih.gov"}


def utc_now():
    return datetime.now(timezone.utc).isoformat()


def sha(path):
    digest = hashlib.sha256()
    with path.open("rb") as stream:
        for block in iter(lambda: stream.read(1024 * 1024), b""):
            digest.update(block)
    return digest.hexdigest()


def require_official_https(url):
    parsed = urllib.parse.urlparse(url)
    if parsed.scheme != "https" or parsed.hostname not in OFFICIAL_HOSTS:
        raise ValueError(f"Expected an official NCBI HTTPS URL, got {url}")


def check_signature(prefix, rec):
    text = prefix.lstrip().lower()
    if text.startswith((b"<!doctype html", b"<html")) or b"ncbi website access" in text or b"checkpoint" in text:
        raise ValueError("NCBI returned an HTML/access checkpoint page, not the frozen GEO input; complete access requirements through the official site, then retry.")
    expected_format = rec.get("format", "gzip" if rec["url"].endswith(".gz") else "geo_platform_text")
    if expected_format == "gzip" and not prefix.startswith(b"\x1f\x8b"):
        raise ValueError("Response is not a gzip file; it may be an access, rate-limit, or error page.")
    if expected_format == "geo_platform_text" and not prefix.startswith(b"^PLATFORM = "):
        raise ValueError("Response is not a GEO full platform text export beginning with ^PLATFORM.")


def verify(path, rec):
    size = path.stat().st_size
    if size != rec["bytes"]:
        raise ValueError(f"BYTE SIZE MISMATCH: expected {rec['bytes']}, got {size}")
    with path.open("rb") as stream:
        check_signature(stream.read(1024), rec)
    actual = sha(path)
    if actual != rec["sha256"]:
        raise ValueError(f"SHA256 MISMATCH: expected {rec['sha256']}, got {actual}; do not replace the frozen hash without scientific revalidation.")
    structure = inspect_table(path) if rec.get('validate_geo_table', False) else {}
    return {"bytes": size, "sha256": actual, **structure}


def download_atomic(path, rec, timeout):
    require_official_https(rec["url"])
    request = urllib.request.Request(rec["url"], headers={"User-Agent": "VascFlux-GEO-reproducibility/1.2", "Accept-Encoding": "identity"})
    temporary_path = None
    try:
        with urllib.request.urlopen(request, timeout=timeout, context=ssl.create_default_context()) as response:
            require_official_https(response.geturl())
            if "text/html" in response.headers.get("Content-Type", "").lower():
                raise ValueError("NCBI returned an HTML/access checkpoint page instead of the frozen input; no cache file was installed.")
            remote_size = response.headers.get("Content-Length")
            if remote_size is not None and int(remote_size) != rec["bytes"]:
                raise ValueError(f"REMOTE BYTE SIZE MISMATCH: frozen {rec['bytes']}, server {remote_size}; download stopped before reading the body. Review provenance or source drift before continuing.")
            with tempfile.NamedTemporaryFile(mode="wb", prefix=path.name + ".", suffix=".part", dir=path.parent, delete=False) as target:
                temporary_path = Path(target.name)
                written = 0
                while True:
                    block = response.read(1024 * 1024)
                    if not block:
                        break
                    if written == 0:
                        check_signature(block[:1024], rec)
                    written += len(block)
                    if written > rec["bytes"]:
                        raise ValueError("Response exceeds the frozen byte length; possible source drift or wrong response.")
                    target.write(block)
                target.flush()
                os.fsync(target.fileno())
        verified = verify(temporary_path, rec)
        os.replace(temporary_path, path)
        temporary_path = None
        return verified
    finally:
        if temporary_path is not None:
            temporary_path.unlink(missing_ok=True)


def main():
    parser = argparse.ArgumentParser(description=__doc__)
    parser.add_argument("--out", type=Path, default=Path("geo_cache"), help="Input cache directory")
    parser.add_argument("--verify-only", action="store_true", help="Read and hash every required cached file; never make network requests")
    parser.add_argument("--files", nargs="+", help="Optional exact-byte input filenames; default is seven exact files. Obtain the other three with acquire_canonical_platforms.py.")
    parser.add_argument("--report", type=Path, help="Write a machine-readable verification report")
    parser.add_argument("--timeout", type=float, default=120, help="Per socket operation timeout in seconds")
    args = parser.parse_args()
    manifest = json.loads(MANIFEST.read_text(encoding="utf-8"))
    names = args.files or [name for name, rec in manifest.items() if rec.get('acquisition_method') != 'canonical_complete_platform_table']
    if len(names) != len(set(names)) or any(name not in manifest for name in names):
        parser.error("--files must contain unique filenames present in the manifest")
    if any(manifest[name].get('acquisition_method') == 'canonical_complete_platform_table' for name in names):
        parser.error('These dynamic platform exports require acquire_canonical_platforms.py; historical archive hashes are not a current acquisition route.')
    if any(Path(name).name != name or "/" in name or "\\" in name for name in manifest):
        parser.error("Manifest filenames must be plain basenames")
    if not args.verify_only:
        args.out.mkdir(parents=True, exist_ok=True)
    report = {"started_utc": utc_now(), "python": platform.python_version(), "mode": "cache_verification_only" if args.verify_only else "download_missing_and_verify", "manifest_sha256": sha(MANIFEST), "source_normalization": "none; exact bytes", "ssl_certificate_verification": "default enabled", "files": {}, "network_downloads_completed": 0}
    for name in names:
        rec, path = manifest[name], args.out / name
        try:
            if path.exists():
                details = verify(path, rec)
                action = "existing_cache_verified"
            elif args.verify_only:
                raise FileNotFoundError("Missing cached input; --verify-only makes no network requests")
            else:
                details = download_atomic(path, rec, args.timeout)
                action = "fresh_download_verified"
                report["network_downloads_completed"] += 1
            report["files"][name] = {"status": "passed", "action": action, **details}
            print(f"PASS {name}: {action}")
        except (OSError, ValueError, urllib.error.URLError) as error:
            report["files"][name] = {"status": "failed", "error": str(error), "cache_policy": "existing files preserved; failed new downloads removed"}
            print(f"FAIL {name}: {error}")
    report["finished_utc"] = utc_now()
    report["status"] = "passed" if all(rec["status"] == "passed" for rec in report["files"].values()) else "failed"
    if args.report:
        args.report.parent.mkdir(parents=True, exist_ok=True)
        args.report.write_text(json.dumps(report, indent=2) + "\n", encoding="utf-8")
    raise SystemExit(0 if report["status"] == "passed" else 1)


if __name__ == "__main__":
    main()
```
