## Supplementary material for "VascFlux-GEO: Comparator selection and auditability in vascular transcriptome module prediction": Analysis code and aggregate results: reproduce.py

```
"""Portable orchestrator for v2 analysis versus archived-output checking.

--verify-only checks saved artifacts and NEVER refits models. --full runs the
requested stages (default: all numerical stages), replacing their generated
outputs. Work on an extracted copy when comparing with archived results.
"""
import argparse
import json
import os
import subprocess
import sys
from datetime import datetime, timezone
from pathlib import Path

ROOT = Path(__file__).resolve().parent
NUMERICAL = ['core', 'descriptive', 'fair', 'simulation', 'robustness', 'external']
FIGURE_SCRIPTS = ['redraw_main_figures.py', 'redraw_supplement_figures.py', 'finalize_figures.py']


def format_provenance_path(token, root=ROOT, path_module=os.path):
    """Display paths without preventing a valid Windows cross-drive run."""
    token = str(token)
    if path_module.isabs(token):
        try:
            token = path_module.relpath(token, str(root))
        except ValueError:
            pass  # Different Windows mounts have no relative path.
        token = token.replace('\\', '/')
    return token


def main():
    p = argparse.ArgumentParser(description=__doc__)
    mode = p.add_mutually_exclusive_group(required=True)
    mode.add_argument('--verify-only', action='store_true')
    mode.add_argument('--full', action='store_true')
    p.add_argument('--data-dir', type=Path, default=ROOT.parent / 'data')
    p.add_argument('--source-manifest', type=Path,
                   help='Required for --full: filename-dict complete input hashes from the canonical acquisition')
    p.add_argument('--input-manifest', type=Path,
                   help='External GSE281841 input manifest; defaults to source-manifest if supplied, otherwise inputs/source_manifest_public.json')
    p.add_argument('--stages', nargs='+', choices=NUMERICAL, default=NUMERICAL)
    p.add_argument('--figures', action='store_true', help='Regenerate the current eight main and seven supplementary figures after numerical execution')
    p.add_argument('--figure-dir', type=Path, default=ROOT.parent/'figures', help='Sibling V07 figure code directory; its sibling reproducibility directory supplies results')
    p.add_argument('--plot-python', default=sys.executable, help='Interpreter for the separate plotting environment')
    p.add_argument('--dry-run', action='store_true', help='Print the commands; do not run them')
    p.add_argument('--report', type=Path, default=ROOT / 'provenance/reproduction_run.json')
    args = p.parse_args()
    if args.verify_only and args.figures:
        p.error('--verify-only cannot regenerate figures; use --full --figures')
    commands = []
    def add(script, extra=(), python=None):
        commands.append([python or sys.executable, str(ROOT / script), *map(str,extra)])
    if args.verify_only:
        add('verify_extended_results.py')
        add('verify_revision_outputs.py')
        interpretation = 'Archived output consistency checks only; no inputs acquired and no models refitted.'
    else:
        if not args.source_manifest:
            p.error('--full requires --source-manifest for the audited complete input set; historical truncated inputs must not be used.')
        data = ['--data-dir', args.data_dir.resolve()]
        manifest = ['--source-manifest', args.source_manifest.resolve()] if args.source_manifest else []
        add('validate_inputs.py', data + manifest + ['--report', ROOT / 'provenance/input_structure_validation.json'])
        stages = set(args.stages)
        if 'robustness' in stages and manifest:
            robustness_source = (ROOT/'run_representation_sensitivity.py').read_text(encoding='utf-8')
            if "'--source-manifest'" not in robustness_source and '"--source-manifest"' not in robustness_source:
                p.error('The current robustness script requires the original frozen cache. Use the default manifest for --full, or explicitly limit --stages core descriptive fair external for the canonical cache.')
        if 'core' in stages:
            add('run_extended_benchmarks.py', data+manifest)
        if 'descriptive' in stages:
            add('run_extended_descriptive.py', data+manifest)
        if {'core','descriptive'}.issubset(stages):
            add('verify_extended_results.py')
        if 'fair' in stages:
            add('run_fair_comparator_extension.py', data+manifest)
        if 'simulation' in stages:
            add('run_known_truth_simulation.py')
        if 'robustness' in stages:
            add('run_representation_sensitivity.py', data+manifest)
        if 'external' in stages:
            external_manifest = args.input_manifest or args.source_manifest or ROOT/'inputs/source_manifest_public.json'
            add('run_external_direction_holdout.py',data+['--input-manifest',external_manifest.resolve()])
        if {'fair','simulation','robustness'}.issubset(stages):
            add('verify_revision_outputs.py')
        if args.figures:
            if args.figure_dir.resolve().parent != ROOT.parent.resolve():
                p.error('--figure-dir must share the package parent with reproducibility; the figure scripts use that explicit layout.')
            for script in FIGURE_SCRIPTS:
                add(args.figure_dir.resolve()/script,python=args.plot_python)
        interpretation = ('Executed requested numerical stages from supplied inputs; simulation uses synthetic data. '
                          'Input acquisition is separate. This does not itself establish fresh-download or clean-OS reproducibility.')
    if args.dry_run:
        interpretation = 'Command plan only: no command executed, no inputs acquired, and no models refitted.'
    record = {'utc':datetime.now(timezone.utc).isoformat(),
              'mode':'archived_check' if args.verify_only else 'numerical_rebuild',
              'requested_stages':[] if args.verify_only else args.stages,
              'scope':interpretation,'dry_run':args.dry_run,'commands':[],'status':'running'}
    try:
        for command in commands:
            print(subprocess.list2cmdline(command),flush=True)
            public_command = ['<plot-python>' if Path(command[1]).name in FIGURE_SCRIPTS else '<analysis-python>']
            for token in command[1:]:
                public_command.append(format_provenance_path(token))
            item = {'argv_for_reproduction':public_command, 'executed':not args.dry_run}
            record['commands'].append(item)
            if not args.dry_run:
                result = subprocess.run(command,cwd=ROOT,check=False)
                item['returncode']=result.returncode
                if result.returncode:
                    raise RuntimeError(f'Stage failed: {Path(command[1]).name}, exit {result.returncode}')
        record['status']='commands_printed_only' if args.dry_run else 'passed'
    except Exception as exc:
        record.update(status='failed',error=str(exc))
        raise
    finally:
        args.report.parent.mkdir(parents=True,exist_ok=True)
        args.report.write_text(json.dumps(record,indent=2)+'\n',encoding='utf-8')
    print(interpretation,flush=True)


if __name__=='__main__':
    main()
```
