## Supplementary material for "VascFlux-GEO: Comparator selection and auditability in vascular transcriptome module prediction": Analysis code and aggregate results: requirements-analysis.txt

# Executed numerical analysis environment: Python 3.12.14.
# Used by run_extended_benchmarks.py, run_extended_descriptive.py,
# and verify_extended_results.py.
numpy==2.3.5
pandas==3.0.1
python-dateutil==3.0.0
six==1.17.0
tzdata==2026.5

# Figure generation was run separately with Python 3.11.9 and:
# numpy==2.4.6
# pandas==2.3.3
# matplotlib==3.10.9
# Do not install both version sets into the same environment.
