## Supplementary material for "VascFlux-GEO: Comparator selection and auditability in vascular transcriptome module prediction": Analysis code and aggregate results: run_extended_benchmarks.py

```
"""Subject-held-out prediction sensitivity and coupling diagnostics, 10 Oct 2026.

Run: python run_extended_benchmarks.py --data-dir ../data
Outputs are confined to results_extended. No download or manuscript mutation.
"""
import argparse
import ast
import csv
import gzip
import hashlib
import io
import json
import os
import platform
import re
from pathlib import Path

import numpy as np
import pandas as pd
from validate_inputs import inspect_table

ROOT = Path(__file__).resolve().parent
GRID = np.array([0.01, 0.1, 1., 10., 100., 1000.])
SHRINK_GRID = np.array([0., .25, .5, .75, 1.])
N_REPEATS = 20
N_NULL = 200
N_BOOTSTRAP = 5000
SEEDS = {'outer_first': 20261007, 'outer_repeats_start': 2026101001,
         'inner': 2026101011, 'null': 2026101021,
         'bootstrap': 2026101031, 'sample_size': 2026101041}
# Read the frozen dictionary without importing the original top-level pipeline.
_tree = ast.parse((ROOT / 'run_vascflux_public.py').read_text(encoding='utf-8'))
MODULES = ast.literal_eval(next(n.value for n in _tree.body
    if isinstance(n, ast.Assign) and any(isinstance(t, ast.Name) and t.id == 'MODULES'
                                         for t in n.targets)))
MOD = list(MODULES)
TARGET = set(sum(MODULES.values(), []))
METHODS = ['mean_delta', 'mean_followup', 'persistence',
           'group_mean_delta', 'group_mean_followup',
           'delta_ridge_fixed_0.01', 'followup_ridge_fixed_0.01',
           'delta_ridge_nested', 'followup_ridge_nested',
           'pca2_followup_nested', 'shrunken_state_followup_nested']


def load_source_manifest(path):
    """Read either legacy records or filename-keyed canonical input records."""
    records = json.loads(Path(path).read_text(encoding='utf-8-sig'))
    return {r['filename']: r for r in records} if isinstance(records, list) else records


def read_platform(path):
    """Retain original first-hit mapping, including module-preferred ambiguous probes."""
    inspect_table(path)  # Refuse malformed/truncated tables even outside reproduce.py.
    opener = gzip.open if str(path).endswith('.gz') else open
    mapping, ambiguous, active = {}, 0, False
    with opener(path, 'rt', errors='replace') as h:
        for line in h:
            if line.startswith('!platform_table_begin'):
                header = next(h).rstrip('\n').split('\t')
                cols = {k: i for i, k in enumerate(header)}
                symcol = next(c for c in ['Gene Symbol', 'Gene symbol', 'Symbol',
                    'Gene Symbol (human)', 'gene_assignment'] if c in cols)
                active = True
                continue
            if line.startswith('!platform_table_end'):
                break
            if not active or not line.strip():
                continue
            parts = line.rstrip('\n').split('\t')
            if len(parts) <= max(cols['ID'], cols[symcol]):
                continue
            raw = parts[cols[symcol]].strip()
            if symcol == 'gene_assignment':
                symbols = [x.split('//')[1].strip() for x in raw.split('///')
                    if len(x.split('//')) >= 2 and x.split('//')[1].strip()
                    and x.split('//')[1].strip() not in ('---', '--- //')]
            else:
                symbols = [x.strip() for x in re.split(r'[;|, ]+', raw.replace('///', ';'))
                           if x.strip() and x.strip() not in ('---', 'NA', 'null', 'nan')]
            ambiguous += int(len(set(symbols)) > 1)
            hit = next((s for s in symbols if s in TARGET), symbols[0] if symbols else None)
            if hit in TARGET:
                mapping[parts[cols['ID']].strip()] = hit
    return mapping, ambiguous


def validate_pairing(rec, sample_subject=None):
    assert len({r['subject'] for r in rec}) == len(rec), 'Duplicate paired subject'
    assert len({r[k] for r in rec for k in ['baseline', 'followup']}) == 2 * len(rec), 'Reused paired sample'
    if sample_subject is not None:
        assert all(sample_subject[r[k]] == r['subject'] for r in rec
                   for k in ['baseline', 'followup']), 'Metadata subject/pair mismatch'


def load_cohort(data, gse, platfile, subject_label, visit_label, treatment_label, expression_only=False):
    mapping, ambiguous = read_platform(data / platfile)
    inspect_table(data / f'{gse}_series_matrix.txt.gz')
    rows, metadata, charrows, active = [], {}, [], False
    with gzip.open(data / f'{gse}_series_matrix.txt.gz', 'rt', errors='replace') as h:
        for line in h:
            if line.startswith('!Sample_'):
                p = next(csv.reader([line], delimiter='\t'))
                if p[0] == '!Sample_characteristics_ch1':
                    charrows.append(p[1:])
                else:
                    metadata[p[0][8:]] = p[1:]
            if line.startswith('!series_matrix_table_begin'):
                active = True
                continue
            if line.startswith('!series_matrix_table_end'):
                break
            if active:
                rows.append(line)
    table = pd.read_csv(io.StringIO(''.join(rows)), sep='\t').copy()
    samples = list(table.columns[1:])
    assert samples == metadata['geo_accession'], 'Metadata/matrix order differs'
    table['symbol'] = table.iloc[:, 0].astype(str).map(mapping)
    table = table[table.symbol.notna()]
    raw = table.drop(columns=[table.columns[0], 'symbol']).apply(pd.to_numeric, errors='coerce')
    raw['symbol'] = table.symbol.values
    raw = raw.groupby('symbol', sort=False).median(numeric_only=True)
    assert np.isfinite(raw.to_numpy()).all(), 'Nonfinite source module-gene expression'
    fields = {}
    for rr in charrows:
        assert len(rr) == len(samples)
        labels = {x.split(':', 1)[0].strip() for x in rr}
        assert len(labels) == 1, 'Inconsistent characteristics labels'
        label = rr[0].split(':', 1)[0].strip()
        assert label not in fields, 'Ambiguous duplicate field label'
        fields[label] = dict(zip(samples, rr))
    if expression_only:
        return {'raw_frame': raw, 'fields': fields, 'samples': samples}
    groups = {}
    for s in samples:
        sub = fields[subject_label][s]
        assert sub.split(':', 1)[1].strip()
        groups.setdefault(sub, []).append(s)
    rec, excluded = [], []
    for sub, ss in groups.items():
        b = [s for s in ss if 'baseline' in fields[visit_label][s].lower()]
        f = [s for s in ss if any(x in fields[visit_label][s].lower() for x in ['final', 'followup'])]
        assert len(b) <= 1 and len(f) <= 1, f'Ambiguous pairing: {sub}'
        if b and f:
            tr = fields[treatment_label][f[0]]
            allowed = 'rosuvastatin' in tr.lower() if gse == 'GSE86216' else 'exercise' in tr.lower()
            if allowed:
                rec.append({'subject': sub, 'baseline': b[0], 'followup': f[0], 'treatment': tr})
            else:
                excluded.append({'subject': sub, 'reason': 'no eligible paired regimen'})
        else:
            excluded.append({'subject': sub, 'reason': 'missing baseline or followup'})
    validate_pairing(rec, fields[subject_label])
    module_index = [np.array([raw.index.get_loc(g) for g in genes if g in raw.index], int)
                    for genes in MODULES.values()]
    assert all(len(x) for x in module_index)
    sample_index = {s: i for i, s in enumerate(raw.columns)}
    cohort = {'name': gse, 'raw': raw.to_numpy(float), 'records': rec,
              'base': np.array([sample_index[r['baseline']] for r in rec]),
              'follow': np.array([sample_index[r['followup']] for r in rec]),
              'module_index': module_index, 'treatment': np.array([r['treatment'] for r in rec]),
              'sample_subject': fields[subject_label],
              'categories': sorted({r['treatment'] for r in rec}) if gse != 'GSE86216' else []}
    cohort['audit'] = {'n_source_samples': len(samples), 'n_source_subjects': len(groups),
        'n_pairs': len(rec), 'excluded_subjects': excluded,
        'paired_unique_subjects': True, 'unique_samples': True,
        'module_gene_counts': {m: len(x) for m, x in zip(MOD, module_index)},
        'n_module_gene_union': len(raw), 'ambiguous_platform_probes': ambiguous,
        'probe_mapping_rule': 'original module-preferred first symbol; median across probes',
        'treatment_counts': {t: int(sum(cohort['treatment'] == t)) for t in set(cohort['treatment'])}}
    return cohort


def states(cohort, train, raw=None):
    a = cohort['raw'] if raw is None else raw
    mu = a[:, cohort['base'][train]].mean(1)
    sd = a[:, cohort['base'][train]].std(1, ddof=0)
    sd[sd == 0] = 1.
    z = (a - mu[:, None]) / sd[:, None]
    scores = np.vstack([np.median(z[ix], 0) for ix in cohort['module_index']]).T
    b, f = scores[cohort['base']], scores[cohort['follow']]
    assert np.isfinite(b).all() and np.isfinite(f).all()
    return b, f, mu, sd


def features(cohort, b, train, test, normalized, pca=False):
    bt, be = b[train], b[test]
    if normalized:
        mu, sd = bt.mean(0), bt.std(0, ddof=0)
        sd[sd < 1e-12] = 1.
        bt, be = (bt - mu) / sd, (be - mu) / sd
    if pca:
        _, _, v = np.linalg.svd(bt, full_matrices=False)
        bt, be = bt @ v[:2].T, be @ v[:2].T
    # All categories are prespecified from public treatment metadata; no outcome use.
    action = np.array([[int(t == cat) for cat in cohort['categories']] for t in cohort['treatment']])
    return np.column_stack([np.ones(len(train)), bt, action[train]]), np.column_stack([np.ones(len(test)), be, action[test]])


def ridge(x, y, lam):
    penalty = np.eye(x.shape[1]) * float(lam)
    penalty[0, 0] = 0.
    return np.linalg.solve(x.T @ x + penalty, x.T @ y)


def splits(n, seed, k=5):
    ix = np.arange(n)
    np.random.default_rng(seed).shuffle(ix)
    return [np.array(x, int) for x in np.array_split(ix, k)]


def inner_cache(cohort, train, seed):
    parts = splits(len(train), seed, 4 if len(train) >= 24 else 3)
    out = []
    for part in parts:
        iv = train[part]
        it = np.array([i for i in train if i not in set(iv)], int)
        b, f, _, _ = states(cohort, it)
        out.append((it, iv, b, f))
    return out


def tune(cohort, cache, kind, grid=GRID):
    losses = []
    for lam in grid:
        errors = []
        for train, test, b, f in cache:
            if kind == 'shrunken':
                pred = f[train].mean(0) + lam * (b[test] - b[train].mean(0))
            else:
                x, xe = features(cohort, b, train, test, True, kind == 'pca2')
                target = f[train] - b[train] if kind == 'delta' else f[train]
                pred = xe @ ridge(x, target, lam)
                if kind == 'delta':
                    pred += b[test]
            errors.extend(np.abs(f[test] - pred).mean(1))
        losses.append(float(np.mean(errors)))
    best = int(np.argmin(losses))
    return float(grid[best]), losses


def predict_all(cohort, train, test, params, b=None, f=None):
    if b is None:
        b, f, _, _ = states(cohort, train)
    d = f - b
    p = {'mean_delta': b[test] + d[train].mean(0),
         'mean_followup': np.tile(f[train].mean(0), (len(test), 1)),
         'persistence': b[test].copy()}
    for label, target in [('group_mean_delta', d), ('group_mean_followup', f)]:
        gp = []
        for i in test:
            match = train[cohort['treatment'][train] == cohort['treatment'][i]]
            match = match if len(match) else train
            gp.append(target[match].mean(0) + (b[i] if label.endswith('delta') else 0))
        p[label] = np.array(gp)
    for normalized, labels in [(False, [('delta', 'delta_ridge_fixed_0.01', .01),
                                      ('followup', 'followup_ridge_fixed_0.01', .01)]),
                               (True, [('delta', 'delta_ridge_nested', params['delta']),
                                       ('followup', 'followup_ridge_nested', params['followup']),
                                       ('pca2', 'pca2_followup_nested', params['pca2'])])]:
        for kind, label, lam in labels:
            x, xe = features(cohort, b, train, test, normalized, kind == 'pca2')
            target = d[train] if kind == 'delta' else f[train]
            pred = xe @ ridge(x, target, lam)
            p[label] = pred + b[test] if kind == 'delta' else pred
    p['shrunken_state_followup_nested'] = f[train].mean(0) + params['shrunken'] * (b[test] - b[train].mean(0))
    assert all(np.isfinite(v).all() for v in p.values())
    return p, b, f


def pooled_metrics(y, p):
    e = y - p
    return {'MAE': float(np.mean(np.abs(e))), 'RMSE': float(np.sqrt(np.mean(e ** 2))),
            'R2': float(1. - np.sum(e ** 2) / (np.sum((y - y.mean(0)) ** 2) + 1e-12))}


def write_csv(out, name, rows):
    pd.DataFrame(rows).to_csv(out / name, index=False)


def repeated_benchmark(cohorts, out):
    oof, folds_rows, tune_rows, fit_cache, split_rows = [], [], [], {}, []
    # First run exactly reproduces the original RNG sequence for both cohorts.
    original_rng = np.random.default_rng(SEEDS['outer_first'])
    original = {}
    for c in cohorts:
        ix = np.arange(len(c['records']))
        original_rng.shuffle(ix)
        original[c['name']] = list(np.array_split(ix, 5))
    for ci, cohort in enumerate(cohorts):
        n, name = len(cohort['records']), cohort['name']
        print(f'{name}: {n} paired subjects, repeated nested CV', flush=True)
        for repeat in range(N_REPEATS):
            seed = SEEDS['outer_first'] if repeat == 0 else SEEDS['outer_repeats_start'] + repeat
            folds = original[name] if repeat == 0 else splits(n, seed + 100 * ci)
            for fold, test in enumerate(folds):
                # Preserve original concatenate-by-fold order for exact reproduction.
                train = np.concatenate([x for j, x in enumerate(folds) if j != fold])
                assert not set(train).intersection(test)
                cache = inner_cache(cohort, train, SEEDS['inner'] + ci * 10000 + repeat * 10 + fold)
                params = {}
                for kind, grid in [('delta', GRID), ('followup', GRID), ('pca2', GRID), ('shrunken', SHRINK_GRID)]:
                    params[kind], losses = tune(cohort, cache, kind, grid)
                    for candidate, loss in zip(grid, losses):
                        tune_rows.append({'cohort': name, 'repeat': repeat, 'fold': fold,
                            'target_model': kind, 'candidate': candidate, 'inner_pooled_MAE': loss,
                            'selected': bool(candidate == params[kind]), 'inner_folds': len(cache),
                            'n_outer_train': len(train)})
                predictions, b, f = predict_all(cohort, train, test, params)
                fit_cache[(name, repeat, fold)] = (train, test, params, b, f)
                for i in test:
                    r = cohort['records'][i]
                    split_rows.append({'cohort': name, 'repeat': repeat, 'seed': seed + 100 * ci if repeat else seed,
                        'fold': fold, 'subject': r['subject'], 'baseline_sample': r['baseline'],
                        'followup_sample': r['followup'], 'treatment': r['treatment']})
                for method, pred in predictions.items():
                    for mi, module in enumerate(MOD):
                        e = f[test, mi] - pred[:, mi]
                        folds_rows.append({'cohort': name, 'repeat': repeat, 'fold': fold,
                            'method': method, 'module': module, 'n_train': len(train), 'n_test': len(test),
                            'MAE': float(np.abs(e).mean()), 'RMSE': float(np.sqrt((e ** 2).mean()))})
                    for j, i in enumerate(test):
                        r = cohort['records'][i]
                        for mi, module in enumerate(MOD):
                            oof.append({'cohort': name, 'repeat': repeat, 'fold': fold,
                                'subject': r['subject'], 'treatment': r['treatment'], 'module': module,
                                'method': method, 'baseline': b[i, mi], 'observed_followup': f[i, mi],
                                'observed_delta': f[i, mi] - b[i, mi], 'predicted_followup': pred[j, mi],
                                'predicted_delta': pred[j, mi] - b[i, mi],
                                'absolute_error': abs(f[i, mi] - pred[j, mi]),
                                'squared_error': (f[i, mi] - pred[j, mi]) ** 2})
            if repeat in (0, 9, 19):
                print(f'  complete {repeat + 1}/{N_REPEATS}', flush=True)
    write_csv(out, 'oof_predictions.csv', oof)
    write_csv(out, 'fold_module_errors.csv', folds_rows)
    write_csv(out, 'nested_tuning.csv', tune_rows)
    write_csv(out, 'subject_folds.csv', split_rows)
    return pd.DataFrame(oof), fit_cache


def summarize_oof(df, cohorts, out):
    metrics_rows, uncertainty_rows, subject_rows, summary = [], [], [], {}
    for ci, c in enumerate(cohorts):
        name = c['name']
        sub = df[df.cohort == name]
        for (repeat, method), s in sub.groupby(['repeat', 'method'], sort=False):
            y, p = s.observed_followup.to_numpy().reshape(-1, len(MOD)), s.predicted_followup.to_numpy().reshape(-1, len(MOD))
            yd, pdelt = s.observed_delta.to_numpy().reshape(-1, len(MOD)), s.predicted_delta.to_numpy().reshape(-1, len(MOD))
            meanf = sub[(sub.repeat == repeat) & (sub.method == 'mean_followup')].squared_error.sum()
            metrics_rows.append({'cohort': name, 'repeat': repeat, 'method': method,
                'n_subjects': len(c['records']), 'n_module_predictions': len(s),
                'MAE': abs(y - p).mean(), 'RMSE': np.sqrt(((y - p) ** 2).mean()),
                'pooled_followup_R2': 1 - np.sum((y - p) ** 2) / np.sum((y - y.mean(0)) ** 2),
                'pooled_delta_R2': 1 - np.sum((yd - pdelt) ** 2) / np.sum((yd - yd.mean(0)) ** 2),
                'followup_Q2_vs_training_mean': 1 - np.sum((y - p) ** 2) / meanf})
        # Average each subject over repetitions and modules before resampling subjects.
        errors = sub.groupby(['subject', 'method']).absolute_error.mean().unstack('method')
        rng = np.random.default_rng(SEEDS['bootstrap'] + ci)
        boot_ix = rng.integers(0, len(errors), (N_BOOTSTRAP, len(errors)))
        summary[name] = {'n_subjects': len(errors), 'methods': {}, 'comparisons': {}}
        for method in METHODS:
            er = errors[method].to_numpy()
            ci95 = np.quantile(er[boot_ix].mean(1), [.025, .975])
            rep = [r['MAE'] for r in metrics_rows if r['cohort'] == name and r['method'] == method]
            summary[name]['methods'][method] = {'MAE_mean_over_repeats': float(er.mean()),
                'conditional_subject_bootstrap_ci95': ci95.tolist(),
                'split_variability_central95': np.quantile(rep, [.025, .975]).tolist()}
            uncertainty_rows.append({'cohort': name, 'method': method, 'comparator': '',
                'estimand': 'subject_mean_repeated_OOF_MAE', 'estimate': er.mean(),
                'conditional_ci95_low': ci95[0], 'conditional_ci95_high': ci95[1],
                'n_subjects': len(errors), 'n_repeats': N_REPEATS, 'bootstrap_draws': N_BOOTSTRAP})
            for subject, err in zip(errors.index, er):
                subject_rows.append({'cohort': name, 'subject': subject, 'method': method,
                                     'mean_MAE_over_repeats_and_modules': err})
            for comparator in ['mean_delta', 'mean_followup', 'group_mean_followup']:
                if method == comparator:
                    continue
                diff = errors[comparator].to_numpy() - er
                cint = np.quantile(diff[boot_ix].mean(1), [.025, .975])
                result = {'MAE_improvement': float(diff.mean()),
                          'conditional_subject_bootstrap_ci95': cint.tolist()}
                summary[name]['comparisons'][f'{method}_vs_{comparator}'] = result
                uncertainty_rows.append({'cohort': name, 'method': method, 'comparator': comparator,
                    'estimand': 'comparator_MAE_minus_method_MAE', 'estimate': diff.mean(),
                    'conditional_ci95_low': cint[0], 'conditional_ci95_high': cint[1],
                    'n_subjects': len(errors), 'n_repeats': N_REPEATS, 'bootstrap_draws': N_BOOTSTRAP})
    write_csv(out, 'repeat_metrics.csv', metrics_rows)
    write_csv(out, 'conditional_subject_intervals.csv', uncertainty_rows)
    write_csv(out, 'subject_average_errors.csv', subject_rows)
    # Fold-standardized module errors; bootstrap resamples subjects, not modules or repeats.
    module_rows = []
    for (name, method, module), s in df.groupby(['cohort', 'method', 'module'], sort=False):
        er = s.groupby('subject').absolute_error.mean().to_numpy()
        rng = np.random.default_rng(SEEDS['bootstrap'] + 99)
        q = np.quantile(er[rng.integers(0, len(er), (N_BOOTSTRAP, len(er)))].mean(1), [.025, .975])
        module_rows.append({'cohort': name, 'method': method, 'module': module,
            'MAE': er.mean(), 'RMSE': np.sqrt(s.squared_error.mean()),
            'conditional_ci95_low': q[0], 'conditional_ci95_high': q[1], 'n_subjects': len(er)})
    write_csv(out, 'module_errors.csv', module_rows)
    return summary


def randomized_followup_null(cohorts, fit_cache, out):
    """Split-local random re-pairing: no source subject crosses train/test.

    Baselines stay observed; followups are permuted within outer training and,
    independently, outer test sets. Delta is then recomputed F_pi - B, preserving
    the -B term. This is a diagnostic distribution, not a formal joint permutation
    test, because permutations differ across overlapping outer folds.
    """
    rows, summary = [], {}
    for ci, c in enumerate(cohorts):
        name = c['name']
        rng = np.random.default_rng(SEEDS['null'] + ci)
        observed = {}
        print(f'{name}: {N_NULL} coupling-preserving re-pairing diagnostics', flush=True)
        for rep in range(-1, N_NULL):
            predall, actualall, baseall, fixed_counts = {}, [], [], 0
            for fold in range(5):
                tr, te, params, b, f = fit_cache[(name, 0, fold)]
                fp = f.copy()
                if rep >= 0:
                    # Within treatment strata in PAD, preserving group structure.
                    for partition in [tr, te]:
                        for treatment in np.unique(c['treatment'][partition]):
                            subset = partition[c['treatment'][partition] == treatment]
                            perm = rng.permutation(subset)
                            fp[subset] = f[perm]
                            fixed_counts += int(sum(subset == perm))
                # Fixed-lambda model is the original diagnostic target; no post-null tuning.
                predictions, _, _ = predict_all(c, tr, te, params, b, fp)
                for method in ['mean_delta', 'mean_followup', 'delta_ridge_fixed_0.01', 'followup_ridge_fixed_0.01']:
                    predall.setdefault(method, []).append(predictions[method])
                actualall.append(fp[te])
                baseall.append(b[te])
            y, b = np.vstack(actualall), np.vstack(baseall)
            for method, pp in predall.items():
                p = np.vstack(pp)
                value = float(np.abs(y - p).mean())
                rows.append({'cohort': name, 'null_repetition': rep, 'method': method,
                    'MAE': value, 'is_observed': rep == -1,
                    'fixed_assignments_across_train_and_test_partitions': fixed_counts,
                    'n_subject_predictions': len(y)})
                if rep == -1:
                    observed[method] = value
        frame = pd.DataFrame([r for r in rows if r['cohort'] == name]).pivot(index='null_repetition', columns='method', values='MAE')
        null = frame.loc[frame.index >= 0]
        summary[name] = {'n_diagnostics': N_NULL, 'outer_repeats': 1,
            'observed_MAE': observed, 'comparisons': {},
            'null_MAE_central95': {m: np.quantile(null[m], [.025, .975]).tolist() for m in null.columns}}
        for comparator in ['mean_delta', 'mean_followup']:
            for method in ['delta_ridge_fixed_0.01', 'followup_ridge_fixed_0.01']:
                vals = null[comparator] - null[method]
                obs = observed[comparator] - observed[method]
                summary[name]['comparisons'][f'{method}_vs_{comparator}'] = {
                    'observed_improvement': obs, 'randomized_pairing_mean_improvement': float(vals.mean()),
                    'randomized_pairing_central95': np.quantile(vals, [.025, .975]).tolist(),
                    'fraction_null_improvements_at_least_observed_plus_one': float((1 + sum(vals >= obs)) / (N_NULL + 1)),
                    'fraction_is_calibrated_p_value': False}
    write_csv(out, 'coupling_preserving_null.csv', rows)
    return summary


def sensitivity(cohorts, fit_cache, out):
    reg_rows, size_rows = [], []
    for ci, c in enumerate(cohorts):
        name = c['name']
        rng = np.random.default_rng(SEEDS['sample_size'] + ci)
        for repeat in range(5):
            for fold in range(5):
                train, test, params, b, f = fit_cache[(name, repeat, fold)]
                for normalized in [False, True]:
                    x, xe = features(c, b, train, test, normalized)
                    for lam in GRID:
                        for target in ['delta', 'followup']:
                            y = f[train] - b[train] if target == 'delta' else f[train]
                            pp = xe @ ridge(x, y, lam)
                            pp += b[test] if target == 'delta' else 0
                            reg_rows.append({'cohort': name, 'repeat': repeat, 'fold': fold,
                                'target': target, 'lambda': lam, 'normalized_module_features': normalized,
                                'n_train': len(train), 'n_test': len(test),
                                'MAE': float(np.abs(f[test] - pp).mean())})
                sizes = [12, 24, 36, 48, len(train)] if name == 'GSE86216' else [6, 8, 10, len(train)]
                for size in sorted(set(s for s in sizes if s <= len(train))):
                    # Class-preserving subsets when both PAD treatment groups occur in outer training.
                    if c['categories']:
                        required = []
                        for treatment in np.unique(c['treatment'][train]):
                            pool = train[c['treatment'][train] == treatment]
                            required.extend(rng.choice(pool, min(2, len(pool)), replace=False))
                        left = np.array([i for i in train if i not in set(required)])
                        subset = np.array(required + list(rng.choice(left, size - len(required), replace=False)))
                    else:
                        subset = rng.choice(train, size, replace=False)
                    if size == len(train):
                        subset = train.copy()
                    cb = inner_cache(c, subset, SEEDS['sample_size'] + ci * 10000 + repeat * 100 + fold * 10 + size)
                    par = {kind: tune(c, cb, kind, grid)[0] for kind, grid in
                           [('delta', GRID), ('followup', GRID), ('pca2', GRID), ('shrunken', SHRINK_GRID)]}
                    predictions, bs, fs = predict_all(c, subset, test, par)
                    for method in ['mean_delta', 'mean_followup', 'delta_ridge_fixed_0.01', 'followup_ridge_nested', 'shrunken_state_followup_nested']:
                        size_rows.append({'cohort': name, 'repeat': repeat, 'fold': fold,
                            'n_train': size, 'n_test': len(test), 'method': method,
                            'MAE': float(np.abs(fs[test] - predictions[method]).mean())})
    write_csv(out, 'regularization_sensitivity.csv', reg_rows)
    write_csv(out, 'sample_size_sensitivity.csv', size_rows)
    summaries = {}
    for key, rows, groups in [('regularization', reg_rows, ['cohort', 'target', 'lambda', 'normalized_module_features']),
                              ('sample_size', size_rows, ['cohort', 'n_train', 'method'])]:
        frame = pd.DataFrame(rows)
        frame['sum_abs_error'] = frame.MAE * frame.n_test
        agg = frame.groupby(groups, as_index=False).agg(sum_abs_error=('sum_abs_error', 'sum'),
            n_test_predictions=('n_test', 'sum'), n_folds=('fold', 'count'))
        agg['MAE'] = agg.sum_abs_error / agg.n_test_predictions
        agg.to_csv(out / f'{key}_sensitivity_summary.csv', index=False)
        summaries[key] = {'n_outer_repeats': 5, 'n_outer_folds_per_repeat': 5,
            'uncertainty': 'descriptive only; overlapping subjects and changing gene-score scales'}
    return summaries


def executable_audit(cohorts, fit_cache, df, out):
    rows, fault_rows = [], []
    for c in cohorts:
        name = c['name']
        tr, te, params, b, f = fit_cache[(name, 0, 0)]
        original, _, _ = predict_all(c, tr, te, params, b, f)
        mutated = c['raw'].copy()
        mutated[:, c['follow'][te]] = mutated[:, c['follow'][te]] * 1000 + 10000
        bm, fm, mum, sdm = states(c, tr, mutated)
        _, _, mu, sd = states(c, tr)
        pred_mutated, _, _ = predict_all(c, tr, te, params, bm, fm)
        max_diff = max(float(np.max(np.abs(original[m] - pred_mutated[m]))) for m in METHODS)
        assert max_diff == 0 and np.array_equal(mu, mum) and np.array_equal(sd, sdm)
        rows.append({'cohort': name, 'test': 'heldout_followup_extreme_mutation',
            'passed': True, 'max_prediction_difference': max_diff,
            'scope': 'fitted outer-fold predictions and train-baseline gene scaler'})
        fault_rows.append({'cohort': name, 'test': 'heldout_followup_extreme_mutation',
            'injected_failure': False, 'expected_detection': False, 'detected': False,
            'passed': True, 'observed_statistic': max_diff,
            'scope': 'clean control: predictions remain invariant under future target mutation'})
        # Positive control deliberately fits gene scaling on all sample visits.
        def leaky_scores(a):
            z = (a - a.mean(1)[:, None]) / a.std(1, ddof=0)[:, None]
            return np.vstack([np.median(z[ix], 0) for ix in c['module_index']]).T
        leaky_diff = float(np.max(np.abs(leaky_scores(mutated)[c['base'][tr]] - leaky_scores(c['raw'])[c['base'][tr]])))
        assert leaky_diff > 1e-6
        fault_rows.append({'cohort': name, 'test': 'all_visit_scaler_future_leakage',
            'injected_failure': True, 'expected_detection': True, 'detected': True,
            'passed': True, 'observed_statistic': leaky_diff,
            'scope': 'deliberately contaminated scaler; mutation reveals changed training baseline states'})
        for label, rec in [('duplicate_subject', c['records'] + [dict(c['records'][0])]),
                           ('reused_sample', [dict(r) for r in c['records']]),
                           ('cross_subject_mispairing', [dict(r) for r in c['records']])]:
            if label == 'reused_sample':
                rec[1]['baseline'] = rec[0]['baseline']
            if label == 'cross_subject_mispairing':
                rec[0]['followup'], rec[1]['followup'] = rec[1]['followup'], rec[0]['followup']
            try:
                validate_pairing(rec, c['sample_subject'])
                detected = False
            except AssertionError:
                detected = True
            assert detected
            fault_rows.append({'cohort': name, 'test': label, 'injected_failure': True,
                'expected_detection': True, 'detected': detected, 'passed': True,
                'observed_statistic': 1, 'scope': 'pairing validator rejects injected ambiguous record'})
        corrupt = c['raw'].copy()
        corrupt[0, c['base'][tr[0]]] = np.nan
        try:
            states(c, tr, corrupt)
            detected = False
        except AssertionError:
            detected = True
        assert detected
        fault_rows.append({'cohort': name, 'test': 'nonfinite_training_expression',
            'injected_failure': True, 'expected_detection': True, 'detected': detected,
            'passed': True, 'observed_statistic': 1, 'scope': 'scoring validator rejects injected NaN'})
        bad_train = np.r_[tr, te[0]]
        detected = bool(set(bad_train).intersection(te))
        assert detected
        fault_rows.append({'cohort': name, 'test': 'subject_overlap_between_train_and_test',
            'injected_failure': True, 'expected_detection': True, 'detected': detected,
            'passed': True, 'observed_statistic': len(set(bad_train).intersection(te)),
            'scope': 'same split-disjointness predicate used during benchmark'})
        # Demonstrate algebraic equivalence at unpenalized OLS (minimum-norm solution).
        x, xe = features(c, b, tr, te, False)
        pdelta = xe @ np.linalg.lstsq(x, f[tr] - b[tr], rcond=None)[0] + b[te]
        pfollow = xe @ np.linalg.lstsq(x, f[tr], rcond=None)[0]
        diff = float(np.max(np.abs(pdelta - pfollow)))
        # Rank deficient tiny PAD folds can invalidate out-of-sample identity.
        full_rank = np.linalg.matrix_rank(x) == x.shape[1]
        # Redundant action dummy/intercept is allowed; baseline columns' exact reconstruction is checked.
        baseline_reconstruction = float(np.max(np.abs(xe @ np.linalg.lstsq(x, b[tr], rcond=None)[0] - b[te])))
        rows.append({'cohort': name, 'test': 'OLS_delta_plus_baseline_vs_direct_followup',
            'passed': bool(diff < 1e-8), 'max_prediction_difference': diff,
            'baseline_reconstruction_max_error': baseline_reconstruction,
            'design_full_column_rank': bool(full_rank),
            'scope': 'algebra check; ridge penalties define different priors'})
        # Equal absolute errors for direct followup and its implied change are an identity.
        check = np.max(np.abs(np.abs(f[te] - original['followup_ridge_nested']) -
            np.abs((f[te] - b[te]) - (original['followup_ridge_nested'] - b[te]))))
        assert check < 1e-12
        rows.append({'cohort': name, 'test': 'followup_vs_implied_delta_absolute_error_identity',
                     'passed': True, 'max_prediction_difference': float(check), 'scope': 'numerical identity'})
    original_path = ROOT / 'results' / 'oof_subject_errors.csv'
    if original_path.exists():
        previous = pd.read_csv(original_path)
        lookup = {'GSE86216_statin': 'GSE86216', 'GSE113873_PAD_exercise': 'GSE113873'}
        previous['cohort'] = previous.cohort.map(lookup)
        now = df[(df.repeat == 0) & (df.method == 'delta_ridge_fixed_0.01')]
        joined = previous.merge(now, on=['cohort', 'fold', 'subject', 'module'], validate='one_to_one')
        diff = float(np.max(np.abs(joined.delta_ridge_prediction - joined.predicted_delta)))
        assert len(joined) == 8 * sum(len(c['records']) for c in cohorts)
        assert diff < 1e-8
        rows.append({'cohort': 'both', 'test': 'original_fixed_lambda_OOF_reproduction',
            'passed': True, 'max_prediction_difference': diff,
            'n_compared_module_predictions': len(joined), 'scope': 'same first folds, raw features, lambda .01'})
    write_csv(out, 'executable_audit.csv', rows)
    write_csv(out, 'injected_fault_controls.csv', fault_rows)
    return rows


def main():
    parser = argparse.ArgumentParser()
    parser.add_argument('--data-dir', type=Path, default=Path(os.environ.get('VASCFLUX_DATA_DIR', ROOT.parent / 'data')))
    parser.add_argument('--output-dir', type=Path, default=ROOT / 'results_extended')
    parser.add_argument('--source-manifest', type=Path, default=ROOT / 'source_cache_manifest.json')
    args = parser.parse_args()
    out = args.output_dir.resolve()
    out.mkdir(parents=True, exist_ok=True)
    cohorts = [load_cohort(args.data_dir, 'GSE86216', 'GPL14951_family.soft.gz',
                          'sibship (sampleid)', 'time', 'treatment'),
               load_cohort(args.data_dir, 'GSE113873', 'GPL15207_full2.txt',
                          'subject', 'visit', 'treatment group')]
    assert [len(c['records']) for c in cohorts] == [72, 14]
    write_csv(out, 'paired_records.csv', [dict(cohort=c['name'], **r) for c in cohorts for r in c['records']])
    provenance = []
    original_manifest = load_source_manifest(args.source_manifest)
    for filename in ['GPL14951_family.soft.gz', 'GPL15207_full2.txt',
                     'GSE86216_series_matrix.txt.gz', 'GSE113873_series_matrix.txt.gz']:
        path = args.data_dir / filename
        sha = hashlib.sha256()
        with path.open('rb') as h:
            while block := h.read(8 * 1024 * 1024):
                sha.update(block)
        match = sha.hexdigest() == original_manifest[filename]['sha256']
        assert match, f'Cached input changed: {filename}'
        provenance.append({'filename': filename, 'bytes': path.stat().st_size,
                           'sha256': sha.hexdigest(), 'matches_selected_manifest': match,
                           'manifest_filename': args.source_manifest.name,
                           'input_kind': original_manifest[filename].get('input_kind', 'frozen_original_input')})
    df, fit_cache = repeated_benchmark(cohorts, out)
    results = summarize_oof(df, cohorts, out)
    audit = executable_audit(cohorts, fit_cache, df, out)
    null = randomized_followup_null(cohorts, fit_cache, out)
    sensitivities = sensitivity(cohorts, fit_cache, out)
    summary = {'analysis_date': '2026-10-10', 'post_hoc_extension': True,
        'n_outer_repeats': N_REPEATS, 'n_outer_folds': 5,
        'n_null_diagnostics': N_NULL, 'n_bootstrap': N_BOOTSTRAP, 'seeds': SEEDS,
        'lambda_grid': GRID.tolist(), 'shrunken_state_grid': SHRINK_GRID.tolist(),
        'methods': METHODS, 'input_provenance': provenance,
        'runtime': {'python': platform.python_version(), 'numpy': np.__version__, 'pandas': pd.__version__},
        'cohort_audit': {c['name']: c['audit'] for c in cohorts},
        'repeated_cv': results, 'coupling_preserving_null': null,
        'sensitivity': sensitivities, 'executable_audit': audit,
        'injected_fault_controls': pd.read_csv(out / 'injected_fault_controls.csv').to_dict(orient='records'),
        'estimand': 'subject-held-out module MAE in train-baseline gene-standardized units; equally average modules, subjects, and repeated splits',
        'uncertainty_scope': 'Conditional subject bootstrap after averaging each subject over all fitted repeated OOF predictions. Repeats/folds/modules are not independent observations. Does not include model-refitting uncertainty.',
        'null_scope': '200 complete five-fold split-local independent followup re-pairings within training/test and treatment partitions; outcomes recomputed as F_permuted-B. Diagnostic fractions are not calibrated permutation p values.',
        'limitations': [
            'Small PAD cohort (14 subjects; 5 exercise/9 no exercise) makes nested tuning highly variable. It is a stress test, not comparative treatment-effect estimation.',
            'Processed public expression and eight overlapping curated modules provide no clinical or device endpoint.',
            'Primary statin cohort has a single observed regimen; intervention/action effects are not identifiable.',
            'Baseline-change coupling survives followup re-pairing; outperforming mean change or delta target permutation is insufficient evidence of biological response information.',
            'Gene standardization changes across splits and sample sizes; repeated/sensitivity errors are descriptive on their corresponding fold scales.',
            'PCA uses two fixed training-only components; this bounded comparison is not a comprehensive search over representations.',
            'Original ambiguous-probe module-preferred annotation policy is retained for exact reproduction; annotation and module selection uncertainty are not included.',
            'Analyses were designed after inspecting original results; no prospective registration or confirmatory multiplicity control is claimed.',
            'No reinforcement learning, learned clinical world model, device simulation, or causal policy evaluation is performed.']}
    (out / 'summary_extended.json').write_text(json.dumps(summary, indent=2, allow_nan=False), encoding='utf-8')
    print(json.dumps({'completed': True, 'output': str(out), 'repeated_cv': results,
                      'coupling_preserving_null': null}, indent=2), flush=True)


if __name__ == '__main__':
    main()
```
