## Supplementary material for "VascFlux-GEO: Comparator selection and auditability in vascular transcriptome module prediction": Analysis code and aggregate results: run_extended_descriptive.py

```
"""Corrected action-specific descriptive module directions and conditional CIs.

No prediction models are fit here. Full-series normalization is explicitly
descriptive. Bootstrap resamples subjects conditional on those module scores.
"""
import argparse
import hashlib
import itertools
import json
from pathlib import Path

import numpy as np
import pandas as pd

from run_extended_benchmarks import ROOT, MOD, MODULES, load_cohort, load_source_manifest

B = 5000
SEED = 2026101051


def cosine(a, b):
    return np.sum(a * b, axis=-1) / (np.linalg.norm(a, axis=-1) * np.linalg.norm(b, axis=-1) + 1e-12)


def expression(data, gse, platform):
    inputs = load_cohort(data, gse, platform, None, None, None, expression_only=True)
    raw = inputs['raw_frame']
    a = raw.to_numpy(float)
    sd = a.std(1, ddof=0)
    sd[sd < 1e-8] = 1.
    z = pd.DataFrame((a - a.mean(1)[:, None]) / sd[:, None], index=raw.index, columns=raw.columns)
    scores = pd.DataFrame({m: z.loc[[g for g in genes if g in z.index]].median(0)
                           for m, genes in MODULES.items()}, index=raw.columns)
    return scores, inputs['fields']


def value(text):
    return text.split(':', 1)[1].strip().lower()


def paired_direction(scores, fields, subject, visit, baseline_term, follow_term, treatment=None):
    result = []
    for sub in dict.fromkeys(fields[subject].values()):
        ss = [s for s in scores.index if fields[subject][s] == sub]
        bs = [s for s in ss if baseline_term in value(fields[visit][s])]
        fs = [s for s in ss if follow_term in value(fields[visit][s])]
        assert len(bs) <= 1 and len(fs) <= 1
        if bs and fs:
            result.append({'subject': sub, 'baseline': bs[0], 'followup': fs[0],
                'treatment': value(fields[treatment][fs[0]]) if treatment else '',
                'delta': (scores.loc[fs[0]] - scores.loc[bs[0]]).to_numpy(float)})
    return result


def main():
    parser = argparse.ArgumentParser()
    parser.add_argument('--data-dir', type=Path, default=ROOT.parent / 'data')
    parser.add_argument('--output-dir', type=Path, default=ROOT / 'results_extended')
    parser.add_argument('--source-manifest', type=Path, default=ROOT / 'source_cache_manifest.json')
    args = parser.parse_args()
    out = args.output_dir.resolve()
    out.mkdir(parents=True, exist_ok=True)
    stat, ms = expression(args.data_dir, 'GSE86216', 'GPL14951_family.soft.gz')
    ex, me = expression(args.data_dir, 'GSE113873', 'GPL15207_full2.txt')
    plaque, mp = expression(args.data_dir, 'GSE43292', 'GPL6244.annot.gz')
    pad, md = expression(args.data_dir, 'GSE27034', 'GPL570_full.txt')
    sr = paired_direction(stat, ms, 'sibship (sampleid)', 'time', 'baseline', 'followup', 'treatment')
    er = paired_direction(ex, me, 'subject', 'visit', 'baseline', 'final', 'treatment group')
    pr = paired_direction(plaque, mp, 'patient', 'tissue', 'intact', 'atheroma')
    sr = [r for r in sr if r['treatment'] == '40 mg rosuvastatin every day for 8-12 weeks']
    exercise = [r for r in er if r['treatment'] == 'exercise']
    noexercise = [r for r in er if r['treatment'] == 'no exercise']
    assert [len(sr), len(exercise), len(noexercise), len(pr)] == [72, 5, 9, 32]
    pds = [s for s in pad.index if 'peripheral' in value(md['disease state'][s])]
    controls = [s for s in pad.index if 'normal' in value(md['disease state'][s])]
    assert [len(pds), len(controls)] == [19, 18]
    groups = {'statin': {'cohort': 'GSE86216', 'a': np.array([r['delta'] for r in sr]), 'b': None},
              'exercise_only': {'cohort': 'GSE113873', 'a': np.array([r['delta'] for r in exercise]), 'b': None},
              'no_exercise_only': {'cohort': 'GSE113873', 'a': np.array([r['delta'] for r in noexercise]), 'b': None},
              'carotid_plaque_minus_intact': {'cohort': 'GSE43292', 'a': np.array([r['delta'] for r in pr]), 'b': None},
              'PAD_PBMC_minus_control': {'cohort': 'GSE27034', 'a': pad.loc[pds].to_numpy(float), 'b': pad.loc[controls].to_numpy(float)}}
    rng = np.random.default_rng(SEED)
    directions, draws, rows, subject_rows = {}, {}, [], []
    for group, g in groups.items():
        a, b = g['a'], g['b']
        boot = a[rng.integers(0, len(a), (B, len(a)))].mean(1)
        observed = a.mean(0)
        if b is not None:
            boot -= b[rng.integers(0, len(b), (B, len(b)))].mean(1)
            observed -= b.mean(0)
        directions[group], draws[group] = observed, boot
        intervals = np.quantile(boot, [.025, .975], axis=0)
        for j, module in enumerate(MOD):
            rows.append({'group': group, 'cohort': g['cohort'], 'module': module,
                'n_subjects': len(a), 'n_control_subjects': len(b) if b is not None else 0,
                'mean_direction': observed[j], 'conditional_ci95_low': intervals[0, j],
                'conditional_ci95_high': intervals[1, j], 'bootstrap_draws': B,
                'normalization': 'full-series gene z-score; descriptive only'})
    # Difference of observed paired changes, explicitly descriptive, not a causal exercise effect.
    difference = directions['exercise_only'] - directions['no_exercise_only']
    difference_draws = draws['exercise_only'] - draws['no_exercise_only']
    q = np.quantile(difference_draws, [.025, .975], axis=0)
    for j, module in enumerate(MOD):
        rows.append({'group': 'exercise_minus_noexercise_paired_change', 'cohort': 'GSE113873',
            'module': module, 'n_subjects': 5, 'n_control_subjects': 9,
            'mean_direction': difference[j], 'conditional_ci95_low': q[0, j],
            'conditional_ci95_high': q[1, j], 'bootstrap_draws': B,
            'normalization': 'full-series gene z-score; descriptive difference in paired changes'})
    for group, rec in [('statin', sr), ('exercise_only', exercise), ('no_exercise_only', noexercise),
                        ('carotid_plaque_minus_intact', pr)]:
        for r in rec:
            for j, module in enumerate(MOD):
                subject_rows.append({'group': group, 'subject': r['subject'], 'baseline_sample': r['baseline'],
                    'followup_sample': r['followup'], 'treatment': r['treatment'],
                    'module': module, 'direction': r['delta'][j]})
    for group, ss in [('PAD_PBMC', pds), ('PBMC_control', controls)]:
        for s in ss:
            for j, module in enumerate(MOD):
                subject_rows.append({'group': group, 'subject': s, 'baseline_sample': s,
                    'followup_sample': '', 'treatment': value(md['disease state'][s]),
                    'module': module, 'direction': pad.loc[s, module]})
    concordance = []
    for ga, gb in itertools.combinations(groups, 2):
        estimate = float(cosine(directions[ga], directions[gb]))
        samples = cosine(draws[ga], draws[gb])
        interval = np.quantile(samples, [.025, .975])
        concordance.append({'group_a': ga, 'group_b': gb, 'n_a': len(groups[ga]['a']),
            'n_b': len(groups[gb]['a']), 'n_control_a': len(groups[ga]['b']) if groups[ga]['b'] is not None else 0,
            'n_control_b': len(groups[gb]['b']) if groups[gb]['b'] is not None else 0,
            'cosine': estimate, 'conditional_ci95_low': interval[0],
            'conditional_ci95_high': interval[1], 'bootstrap_draws': B,
            'conditional_on_full_series_normalization': True})
    pd.DataFrame(rows).to_csv(out / 'corrected_group_module_directions.csv', index=False)
    pd.DataFrame(subject_rows).to_csv(out / 'corrected_subject_module_directions.csv', index=False)
    pd.DataFrame(concordance).to_csv(out / 'corrected_cross_bed_concordance.csv', index=False)
    # Historical combined direction reproduces the previously mislabeled exercise contrast.
    combined = np.array([r['delta'] for r in exercise + noexercise]).mean(0)
    legacy = {'combined_n': 14, 'exercise_only_n': 5, 'noexercise_only_n': 9,
        'combined_direction': dict(zip(MOD, combined.tolist())),
        'historical_combined_vs_PAD_PBMC_cosine': float(cosine(combined, directions['PAD_PBMC_minus_control'])),
        'historical_combined_vs_statin_cosine': float(cosine(combined, directions['statin'])),
        'correction': "Original substring 'exercise' retained both exercise and no exercise; exact parsed action matching now separates groups."}
    provenance = []
    manifest = load_source_manifest(args.source_manifest)
    for fn in ['GPL14951_family.soft.gz', 'GPL15207_full2.txt', 'GSE86216_series_matrix.txt.gz', 'GSE113873_series_matrix.txt.gz',
               'GPL6244.annot.gz', 'GPL570_full.txt', 'GSE43292_series_matrix.txt.gz', 'GSE27034_series_matrix.txt.gz']:
        h = hashlib.sha256()
        path = args.data_dir / fn
        with path.open('rb') as source:
            while block := source.read(8 * 1024 * 1024):
                h.update(block)
        assert h.hexdigest() == manifest[fn]['sha256']
        provenance.append({'filename': fn, 'bytes': path.stat().st_size, 'sha256': h.hexdigest(),
                           'matches_selected_manifest': True, 'manifest_filename': args.source_manifest.name,
                           'input_kind': manifest[fn].get('input_kind', 'frozen_original_input')})
    summary = {'analysis_date': '2026-10-10', 'bootstrap_draws': B, 'seed': SEED,
        'action_matching': 'exact value after metadata label; no substring matching',
        'group_counts': {g: {'n': len(v['a']), 'n_control': len(v['b']) if v['b'] is not None else 0}
                         for g, v in groups.items()},
        'historical_label_correction': legacy, 'cross_bed_concordance': concordance,
        'additional_input_provenance': provenance,
        'uncertainty_scope': 'Bootstrap resamples paired subjects within groups; PAD PBMC resamples disease/control subjects independently. Normalization and annotation are held fixed. Overlapping modules are not resampled or treated as independent.',
        'limitations': ['Full-series gene scaling is permitted only for these descriptive contrasts and is separate from predictive train-only scaling.',
            'Small action groups (5 exercise, 9 no exercise) give uncertain direction and cosine estimates.',
            'Cosine reflects cross-cohort descriptive alignment and cannot identify a common mechanism or treatment effect.',
            'Eight curated modules overlap in genes; no independence or confirmatory multiple-testing claims are made.',
            'PBMC and carotid tissue contrasts differ in tissue, population and contrast semantics.']}
    (out / 'summary_descriptive_corrected.json').write_text(json.dumps(summary, indent=2, allow_nan=False), encoding='utf-8')
    print(json.dumps(summary, indent=2), flush=True)


if __name__ == '__main__':
    main()
```
