## Supplementary material for "VascFlux-GEO: Comparator selection and auditability in vascular transcriptome module prediction": Analysis code and aggregate results: run_external_direction_holdout.py

```
"""Post hoc independent-source cross-sectional description of GSE281841.

The historical filename is retained for compatibility. No follow-up predictor
is evaluated; this is not a longitudinal holdout or clinical/causal evidence.
"""
import argparse
import csv
import gzip
import hashlib
import json
import re
from pathlib import Path
import numpy as np
import pandas as pd
from run_extended_benchmarks import MODULES, MOD, load_cohort

ROOT = Path(__file__).resolve().parent
B, SEED = 5000, 2026101511


def sha256(path):
    h = hashlib.sha256()
    with path.open('rb') as f:
        for chunk in iter(lambda: f.read(1024 * 1024), b''):
            h.update(chunk)
    return h.hexdigest()


def verify_inputs(data, manifest_path):
    manifest = json.loads(manifest_path.read_text(encoding='utf-8-sig'))
    records = []
    for name in ['GPL96_family.soft.gz', 'GSE281841_series_matrix.txt.gz']:
        expected = manifest.get(name, {}).get('sha256', '')
        if not re.fullmatch(r'[0-9a-f]{64}', expected):
            raise RuntimeError(f'Missing frozen SHA-256: {name}')
        actual = sha256(data / name)
        if actual != expected:
            raise RuntimeError(f'Frozen input SHA-256 mismatch: {name}')
        records.append({'filename': name, 'bytes': (data / name).stat().st_size,
                        'sha256': actual, 'expected_sha256': expected, 'hash_match': True,
                        'url': manifest[name].get('url')})
    return records


def read_metadata(path):
    meta = {}
    with gzip.open(path, 'rt', errors='strict') as f:
        for line in f:
            if line.startswith('!series_matrix_table_begin'):
                break
            if line.startswith('!Sample_') and not line.startswith('!Sample_characteristics_ch1'):
                row = next(csv.reader([line], delimiter='\t'))
                key = row[0][8:]
                if key not in {'geo_accession', 'title', 'source_name_ch1'}:
                    continue
                if key in meta:
                    raise RuntimeError(f'Duplicate metadata: {key}')
                meta[key] = row[1:]
    return meta


def patient_id(text):
    found = re.findall(r'\b(P\d{3,4}|Pt\d{2,3})\b', text)
    if len(found) != 1:
        raise RuntimeError(f'Ambiguous/missing patient ID: {text}')
    return found[0]  # Do not merge P and Pt forms without a source crosswalk.


def scores_and_coverage(raw):
    target = set().union(*map(set, MODULES.values()))
    present = set(raw.index)
    coverage = pd.DataFrame([{'module': m, 'requested_genes': len(set(genes)),
        'mapped_genes': len(set(genes) & present),
        'coverage_fraction': len(set(genes) & present) / len(set(genes)),
        'mapped_symbols': ';'.join(sorted(set(genes) & present)),
        'missing_symbols': ';'.join(sorted(set(genes) - present))} for m, genes in MODULES.items()])
    if target - present:
        raise RuntimeError('Incomplete frozen coverage: ' + ','.join(sorted(target - present)))
    a = raw.to_numpy(float)
    if not np.isfinite(a).all():
        raise RuntimeError('Nonfinite expression')
    sd = a.std(1, ddof=0)
    sd[sd < 1e-8] = 1.0
    z = pd.DataFrame((a - a.mean(1)[:, None]) / sd[:, None], index=raw.index, columns=raw.columns)
    scores = pd.DataFrame({m: z.loc[genes].median(axis=0) for m, genes in MODULES.items()}, index=raw.columns)
    return scores, coverage, len(target), len(target & present)


def contrast(patients, seed=SEED):
    hi = patients.loc[patients.group == 'high_4_5', MOD].to_numpy(float)
    lo = patients.loc[patients.group == 'low_0_1', MOD].to_numpy(float)
    if min(len(hi), len(lo)) < 2:
        raise RuntimeError('At least two patients per group required for a bootstrap contrast')
    rng = np.random.default_rng(seed)
    draws = hi[rng.integers(0, len(hi), (B, len(hi)))].mean(1) - lo[rng.integers(0, len(lo), (B, len(lo)))].mean(1)
    ci = np.quantile(draws, [.025, .975], axis=0)
    return pd.DataFrame({'module': MOD, 'mean_high_minus_low': hi.mean(0) - lo.mean(0),
        'conditional_ci95_low': ci[0], 'conditional_ci95_high': ci[1],
        'n_high_patients': len(hi), 'n_low_patients': len(lo), 'bootstrap_draws': B,
        'normalization': 'full-series gene z-score; descriptive only; normalization held fixed',
        'external_role': 'post hoc independent-source cross-sectional descriptive contrast',
        'interval_status': 'conditional; no multiplicity correction'})


def main():
    ap = argparse.ArgumentParser()
    ap.add_argument('--data-dir', type=Path, default=ROOT.parent / 'data')
    ap.add_argument('--output-dir', type=Path, default=ROOT / 'results_extended')
    ap.add_argument('--input-manifest', type=Path, default=ROOT / 'inputs/source_manifest_public.json')
    args = ap.parse_args()
    data, out = args.data_dir.resolve(), args.output_dir.resolve()
    provenance = verify_inputs(data, args.input_manifest.resolve())
    cohort = load_cohort(data, 'GSE281841', 'GPL96_family.soft.gz', None, None, None, expression_only=True)
    scores, coverage, requested, mapped = scores_and_coverage(cohort['raw_frame'])
    meta = read_metadata(data / 'GSE281841_series_matrix.txt.gz')
    if list(scores.index) != meta['geo_accession']:
        raise RuntimeError('Sample metadata order mismatch')
    rows = []
    for i, sample in enumerate(scores.index):
        title, source = meta['title'][i], meta['source_name_ch1'][i]
        pid, source_pid = patient_id(title), patient_id(source)
        if pid != source_pid:
            raise RuntimeError(f'Patient ID mismatch: {sample}')
        value = cohort['fields']['stenosis score'][sample].split(':', 1)[1].strip().lower()
        score = int(value) if re.fullmatch(r'\d+', value) else None
        group = 'high_4_5' if score in {4, 5} else ('low_0_1' if score in {0, 1} else 'ineligible')
        artery_match = re.fullmatch(r'(.+?)\s+artery\s+from\s+Patient\s+.+', source, flags=re.IGNORECASE)
        prefix_match = re.search(r'\[(\d+)_', title)
        if not artery_match or not prefix_match:
            raise RuntimeError(f'Unparsed artery or title token: {sample}')
        artery = ' '.join(artery_match.group(1).lower().split())
        age = cohort['fields']['patient age'][sample].split(':', 1)[1].strip()
        if not re.fullmatch(r'\d+', age):
            raise RuntimeError(f'Non-numeric source age: {sample}')
        rows.append({'source_column_1based': i + 1, 'sample': sample, 'patient': pid,
            'patient_id_title': pid, 'patient_id_source_name': source_pid,
            'patient_id_status': 'exact title/source agreement; original identifier preserved',
            'title': title, 'source_name_ch1': source, 'stenosis_score_raw': value,
            'stenosis_score': score, 'score_parse_status': 'numeric' if score is not None else 'missing_or_non_numeric',
            'group': group, 'extreme_score_eligible': group != 'ineligible', 'patient_age': int(age),
            'artery': artery, 'segment': 'proximal' if artery.startswith('proximal ') else 'not separately specified',
            'title_prefix_proxy': prefix_match.group(1), 'proxy_status': 'title-derived token; not confirmed batch',
            **{m: float(scores.loc[sample, m]) for m in MOD}})
    sample_df = pd.DataFrame(rows)
    for field in ['patient_age', 'title_prefix_proxy']:
        if (sample_df.groupby('patient')[field].nunique() > 1).any():
            raise RuntimeError(f'Within-patient {field} inconsistency')
    extreme = sample_df.loc[sample_df.extreme_score_eligible]
    groups = extreme.groupby('patient').group.nunique()
    mixed = sorted(groups[groups > 1].index)
    sample_df['included_primary'] = sample_df.extreme_score_eligible & ~sample_df.patient.isin(mixed)
    sample_df['selection_reason'] = np.where(sample_df.included_primary, 'included: extreme score; one grade group per patient',
        np.where(sample_df.patient.isin(mixed), 'excluded: patient has both high and low grades',
                 'excluded: missing or non-extreme stenosis score'))
    clean = sample_df.loc[sample_df.included_primary].copy()
    patients = clean.groupby(['patient', 'group'], as_index=False)[MOD].mean()
    patient_meta = clean.groupby('patient', as_index=False).agg(patient_age=('patient_age', 'first'),
        title_prefix_proxy=('title_prefix_proxy', 'first'), n_samples=('sample', 'count'),
        arteries=('artery', lambda v: ';'.join(sorted(set(v)))), source_samples=('sample', lambda v: ';'.join(v)))
    patient_export = patients.merge(patient_meta, on='patient', validate='one_to_one')
    actual = (len(sample_df), sample_df.patient.nunique(), len(clean), len(patients),
              int((patients.group == 'high_4_5').sum()), int((patients.group == 'low_0_1').sum()))
    if actual != (41, 28, 38, 26, 18, 8):
        raise RuntimeError(f'Unexpected frozen-source selection flow: {actual}')
    primary = contrast(patients)
    confounding = []
    for group, sub in clean.groupby('group'):
        ages = sub.groupby('patient').patient_age.first()
        for stat, value in [('n', len(ages)), ('mean', ages.mean()), ('sd', ages.std(ddof=1)), ('min', ages.min()), ('max', ages.max())]:
            confounding.append({'variable': 'patient_age', 'category': stat, 'group': group,
                                'unit': 'patient', 'value': value, 'denominator': len(ages)})
        for field in ['artery', 'segment', 'title_prefix_proxy']:
            for value in sorted(clean[field].unique()):
                selected = sub.loc[sub[field] == value]
                confounding.append({'variable': field, 'category': value, 'group': group,
                    'unit': 'sample', 'value': len(selected), 'denominator': len(sub)})
                confounding.append({'variable': field, 'category': value, 'group': group,
                    'unit': 'patient_with_at_least_one_sample', 'value': selected.patient.nunique(),
                    'denominator': sub.patient.nunique()})
    # Two restrained sensitivity families, selected by source metadata overlap,
    # not by effect size: same artery, and same unverified title prefix.
    sensitivities, overlap = [], []
    for field in ['artery', 'title_prefix_proxy']:
        for value, sub in clean.groupby(field, sort=True):
            pp = sub.groupby(['patient', 'group'], as_index=False)[MOD].mean()
            nh, nl = int((pp.group == 'high_4_5').sum()), int((pp.group == 'low_0_1').sum())
            eligible = min(nh, nl) >= 2
            overlap.append({'restriction': field, 'stratum': value, 'n_high_patients': nh,
                'n_low_patients': nl, 'n_samples': len(sub), 'estimated': eligible,
                'reason': 'both groups have at least two patients' if eligible else 'not estimated: sparse or absent group overlap'})
            if eligible:
                estimate = contrast(pp, SEED + 100 + len(sensitivities))
                estimate.insert(0, 'stratum', value)
                estimate.insert(0, 'restriction', field)
                estimate['status'] = 'exploratory within observed stratum; unadjusted; correlated comparisons'
                sensitivities.append(estimate)
    summary = {'study': 'GSE281841', 'platform': 'GPL96', 'source_samples': len(sample_df),
        'source_patient_ids': int(sample_df.patient.nunique()), 'usable_extreme_samples': len(extreme),
        'patient_count_before_excluding_mixed': int(extreme.patient.nunique()),
        'mixed_grade_patients_excluded': mixed, 'unscored_sample_ids': sample_df.loc[sample_df.stenosis_score.isna(), 'sample'].tolist(),
        'retained_samples': len(clean), 'patient_count_after_excluding_mixed': len(patients),
        'high_4_5_patients': actual[-2], 'low_0_1_patients': actual[-1],
        'high_4_5_samples': int((clean.group == 'high_4_5').sum()), 'low_0_1_samples': int((clean.group == 'low_0_1').sum()),
        'mapping': {'requested_unique_genes': requested, 'mapped_unique_genes': mapped,
                    'missing_symbols': [], 'per_module': coverage.to_dict(orient='records')},
        'bootstrap_draws': B, 'seed': SEED, 'input_manifest_sha256': sha256(args.input_manifest.resolve()),
        'provenance': provenance, 'sensitivity_families': ['same artery', 'same unverified title prefix'],
        'interpretation': 'Post hoc independent-source cross-sectional description only; not longitudinal prediction holdout, clinical utility, device evidence, or causal inference.',
        'limitations': [
            'Matrix has 41 columns / 28 exact patient IDs; GEO summary reports 39 samples / 27 patients. The discrepancy is unresolved.',
            'One unscored sample and two samples from one mixed-grade identifier are excluded. Retained 38 samples are averaged to 26 literal source IDs.',
            'P-prefixed and Pt-prefixed IDs are not merged without an author-supplied crosswalk; true patient identity cannot be independently verified.',
            'Lower-grade arteries belong to an end-stage PAD/amputation series; they are not healthy control individuals.',
            'Age is source metadata; artery is parsed from source_name_ch1. Only proximal is an explicit segment qualifier; other segment details are missing.',
            '0092 and 0521 are title-derived proxies only; they are not confirmed assay or biological batches.',
            'Different artery/age/proxy composition and cell composition may confound contrasts. Restrictions do not remove residual confounding.',
            'Full-series normalization is descriptive. Bootstrap intervals hold normalization/mapping/modules fixed and are not multiplicity-adjusted.',
            'Sparse overlap does not support a robust multivariable adjusted model; none is fitted.',
            'All contrasts and sensitivities are post hoc exploratory; no treatment, causal, device or clinical inference is supported.']}
    out.mkdir(parents=True, exist_ok=True)
    exports = {'direction_holdout': primary, 'sample_audit': sample_df, 'patient_scores': patient_export,
        'module_coverage': coverage, 'confounding': pd.DataFrame(confounding),
        'restricted_sensitivities': pd.concat(sensitivities, ignore_index=True), 'stratum_overlap_audit': pd.DataFrame(overlap)}
    for suffix, frame in exports.items():
        frame.to_csv(out / f'external_GSE281841_{suffix}.csv', index=False)
    (out / 'summary_external_GSE281841.json').write_text(json.dumps(summary, indent=2), encoding='utf-8')
    print(json.dumps({k: summary[k] for k in ['source_samples', 'source_patient_ids', 'usable_extreme_samples',
        'retained_samples', 'patient_count_after_excluding_mixed', 'high_4_5_patients', 'low_0_1_patients']}, indent=2))


if __name__ == '__main__':
    main()
```
