## Supplementary material for "VascFlux-GEO: Comparator selection and auditability in vascular transcriptome module prediction": Analysis code and aggregate results: run_fair_comparator_extension.py

```
"""Post hoc MAE-aligned comparators on the immutable exported repeated folds.

Run: python run_fair_comparator_extension.py --data-dir ../data
Only results_reviewer_revision/fair_* and summary_fair_comparators.json are written.
Original benchmark predictions are read and never overwritten. All intervals are
conditional subject bootstraps of fixed repeated OOF errors, not refit intervals.
"""
import argparse
import hashlib
import json
import platform
from pathlib import Path

import numpy as np
import pandas as pd

from run_extended_benchmarks import MOD, load_cohort, states, load_source_manifest

ROOT = Path(__file__).resolve().parent
N_REPEATS = 20
N_FOLDS = 5
N_BOOTSTRAP = 5000
BOOTSTRAP_SEED = 2026101517
NEW_METHODS = ['median_followup', 'group_median_followup',
               'median_delta', 'group_median_delta']
REFERENCE_METHODS = ['followup_ridge_nested',
                     'shrunken_state_followup_nested', 'mean_followup']
SIMPLE_METHODS = ['mean_delta', 'mean_followup', 'persistence',
                  'group_mean_delta', 'group_mean_followup'] + NEW_METHODS
KEY = ['cohort', 'repeat', 'fold', 'subject', 'module']


def sha256(path):
    h = hashlib.sha256()
    with Path(path).open('rb') as handle:
        for block in iter(lambda: handle.read(8 * 1024 * 1024), b''):
            h.update(block)
    return h.hexdigest()


def dump_json(path, value):
    path.write_text(json.dumps(value, indent=2, allow_nan=False), encoding='utf-8')


def main():
    parser = argparse.ArgumentParser()
    parser.add_argument('--data-dir', type=Path, default=ROOT.parent / 'data')
    parser.add_argument('--output-dir', type=Path, default=ROOT / 'results_reviewer_revision')
    parser.add_argument('--source-manifest', type=Path, default=ROOT / 'source_cache_manifest.json')
    args = parser.parse_args()
    out = args.output_dir.resolve()
    out.mkdir(parents=True, exist_ok=True)
    original_paths = [ROOT / 'results_extended' / name for name in
                      ['oof_predictions.csv', 'subject_folds.csv', 'nested_tuning.csv']]
    original_hashes = {p.name: sha256(p) for p in original_paths}
    source_files = ['GPL14951_family.soft.gz', 'GPL15207_full2.txt',
                    'GSE86216_series_matrix.txt.gz', 'GSE113873_series_matrix.txt.gz']
    source_manifest = load_source_manifest(args.source_manifest)
    source_hashes = {}
    for name in source_files:
        actual = sha256(args.data_dir / name)
        assert actual == source_manifest[name]['sha256'], f'Source hash mismatch: {name}'
        source_hashes[name] = actual
    original = pd.read_csv(original_paths[0])
    folds = pd.read_csv(original_paths[1])
    assert not original.duplicated(KEY + ['method']).any()
    assert not folds.duplicated(['cohort', 'repeat', 'subject']).any()
    assert original.method.nunique() == 11
    assert set(folds['repeat']) == set(range(N_REPEATS))
    assert set(folds['fold']) == set(range(N_FOLDS))
    cohorts = [load_cohort(args.data_dir, 'GSE86216', 'GPL14951_family.soft.gz',
                           'sibship (sampleid)', 'time', 'treatment'),
               load_cohort(args.data_dir, 'GSE113873', 'GPL15207_full2.txt',
                           'subject', 'visit', 'treatment group')]
    assert [len(c['records']) for c in cohorts] == [72, 14]
    assert len(folds) == 86 * N_REPEATS
    all_rows, invariant_rows = [], []
    for cohort in cohorts:
        name = cohort['name']
        index = {r['subject']: i for i, r in enumerate(cohort['records'])}
        cf = folds[folds.cohort == name]
        for repeat in range(N_REPEATS):
            repeat_rows = cf[cf['repeat'] == repeat]
            assert set(repeat_rows.subject) == set(index)
            fold_arrays = [np.array([index[s] for s in
                           repeat_rows[repeat_rows.fold == fold].subject], dtype=int)
                           for fold in range(N_FOLDS)]
            for fold, test in enumerate(fold_arrays):
                # Identical concatenation and within-fold order to the exported run.
                train = np.concatenate([a for k, a in enumerate(fold_arrays) if k != fold])
                assert not set(train).intersection(test)
                assert set(train).union(test) == set(range(len(index)))
                b, f, _, _ = states(cohort, train)
                d = f - b
                predictions = {
                    'median_followup': np.tile(np.median(f[train], axis=0), (len(test), 1)),
                    'median_delta': b[test] + np.median(d[train], axis=0),
                }
                fallback_count = 0
                for method, target in [('group_median_followup', f), ('group_median_delta', d)]:
                    pred = []
                    for i in test:
                        matching = train[cohort['treatment'][train] == cohort['treatment'][i]]
                        if len(matching) == 0:
                            matching = train
                            fallback_count += 1
                        pred.append(np.median(target[matching], axis=0) +
                                    (b[i] if method == 'group_median_delta' else 0))
                    predictions[method] = np.array(pred)
                max_state_error = 0.0
                max_mean_prediction_error = 0.0
                orig_fold = original[(original.cohort == name) &
                    (original['repeat'] == repeat) & (original.fold == fold) &
                    (original.method == 'mean_followup')].set_index(['subject', 'module'])
                assert len(orig_fold) == len(test) * len(MOD)
                for j, i in enumerate(test):
                    record = cohort['records'][i]
                    recorded = repeat_rows[repeat_rows.subject == record['subject']].iloc[0]
                    assert recorded.baseline_sample == record['baseline']
                    assert recorded.followup_sample == record['followup']
                    assert recorded.treatment == record['treatment']
                    for mi, module in enumerate(MOD):
                        old = orig_fold.loc[(record['subject'], module)]
                        state_error = max(abs(old.baseline - b[i, mi]),
                                          abs(old.observed_followup - f[i, mi]))
                        mean_error = abs(old.predicted_followup - f[train, mi].mean())
                        max_state_error = max(max_state_error, float(state_error))
                        max_mean_prediction_error = max(max_mean_prediction_error, float(mean_error))
                        for method in NEW_METHODS:
                            pred = predictions[method][j, mi]
                            all_rows.append({'cohort': name, 'repeat': repeat, 'fold': fold,
                                'subject': record['subject'], 'treatment': record['treatment'],
                                'module': module, 'method': method, 'baseline': b[i, mi],
                                'observed_followup': f[i, mi], 'observed_delta': d[i, mi],
                                'predicted_followup': pred, 'predicted_delta': pred - b[i, mi],
                                'absolute_error': abs(f[i, mi] - pred),
                                'squared_error': (f[i, mi] - pred) ** 2})
                assert max_state_error < 1e-12, f'Transformed outcome mismatch: {name}/{repeat}/{fold}'
                assert max_mean_prediction_error < 1e-12, 'Training reconstruction differs'
                invariant_rows.append({'cohort': name, 'repeat': repeat, 'fold': fold,
                    'n_train': len(train), 'n_test': len(test), 'subject_disjoint': True,
                    'exported_fold_and_sample_identity': True, 'finite_predictions':
                    all(np.isfinite(p).all() for p in predictions.values()),
                    'max_state_difference_from_original': max_state_error,
                    'max_mean_followup_prediction_difference': max_mean_prediction_error,
                    'group_fallback_calls_across_two_group_methods': fallback_count})
        print(f'{name}: reconstructed all 100 outer folds; 4 median comparators', flush=True)
    new = pd.DataFrame(all_rows)
    expected_new_rows = 86 * N_REPEATS * len(MOD) * len(NEW_METHODS)
    assert len(new) == expected_new_rows == 55040
    assert not new.duplicated(KEY + ['method']).any()
    assert np.isfinite(new.select_dtypes(include='number')).all().all()
    assert len(invariant_rows) == 200
    assert all(row['finite_predictions'] for row in invariant_rows)
    assert (new.groupby(['cohort', 'subject', 'method', 'module']).size() == N_REPEATS).all()
    joined = new.merge(folds[['cohort', 'repeat', 'fold', 'subject']],
                       on=['cohort', 'repeat', 'subject'], suffixes=('', '_exported'), validate='many_to_one')
    assert len(joined) == len(new) and (joined.fold == joined.fold_exported).all()
    new.to_csv(out / 'fair_oof_predictions.csv', index=False)
    pd.DataFrame(invariant_rows).to_csv(out / 'fair_fold_invariants.csv', index=False)
    combined = pd.concat([original, new], ignore_index=True)
    assert combined.method.nunique() == 15
    subject_errors = combined.groupby(['cohort', 'subject', 'method'], sort=True).absolute_error.mean()
    subject_table = subject_errors.rename('mean_MAE_over_repeats_and_modules').reset_index()
    subject_table.to_csv(out / 'fair_subject_average_errors.csv', index=False)
    repeat_metrics = combined.groupby(['cohort', 'repeat', 'method'], sort=True).agg(
        MAE=('absolute_error', 'mean'), mean_squared_error=('squared_error', 'mean'),
        n_subjects=('subject', 'nunique'), n_module_predictions=('module', 'size')).reset_index()
    repeat_metrics['RMSE'] = np.sqrt(repeat_metrics.pop('mean_squared_error'))
    repeat_metrics.to_csv(out / 'fair_repeat_metrics.csv', index=False)
    paired_rows, subject_difference_rows, summary = [], [], {}
    for ci, cohort in enumerate(cohorts):
        name = cohort['name']
        error = subject_errors.loc[name].unstack('method').sort_index()
        assert len(error) == len(cohort['records']) and error.notna().all().all()
        rng = np.random.default_rng(BOOTSTRAP_SEED + ci)
        boot_ix = rng.integers(0, len(error), (N_BOOTSTRAP, len(error)))
        methods = {}
        for method in error.columns:
            values = error[method].to_numpy()
            ci95 = np.quantile(values[boot_ix].mean(axis=1), [.025, .975])
            methods[method] = {'MAE': float(values.mean()),
                'conditional_subject_bootstrap_ci95': ci95.tolist()}
        # The empirically strongest constant/change baseline is selected only for
        # descriptive comparison. Intervals do not correct for this selection.
        best_simple = min(SIMPLE_METHODS, key=lambda m: (methods[m]['MAE'], SIMPLE_METHODS.index(m)))
        comparisons = [(method, comparator, 'new_MAE_aligned_comparator')
                       for method in REFERENCE_METHODS for comparator in NEW_METHODS]
        comparisons += [('followup_ridge_nested', 'shrunken_state_followup_nested',
                         'direct_ridge_vs_scalar'),
                        ('followup_ridge_nested', best_simple, 'empirically_best_simple_baseline'),
                        ('shrunken_state_followup_nested', best_simple, 'empirically_best_simple_baseline')]
        for method, comparator, comparison_role in comparisons:
            diff = (error[comparator] - error[method]).to_numpy()
            ci95 = np.quantile(diff[boot_ix].mean(axis=1), [.025, .975])
            result = {'cohort': name, 'method': method, 'comparator': comparator,
                'comparison_role': comparison_role, 'estimand': 'comparator_MAE_minus_method_MAE',
                'estimate': float(diff.mean()), 'conditional_ci95_low': float(ci95[0]),
                'conditional_ci95_high': float(ci95[1]), 'n_subjects': len(error),
                'n_repeats': N_REPEATS, 'bootstrap_draws': N_BOOTSTRAP,
                'bootstrap_seed': BOOTSTRAP_SEED + ci, 'post_hoc_nonconfirmatory': True}
            paired_rows.append(result)
            for subject, difference in zip(error.index, diff):
                subject_difference_rows.append({'cohort': name, 'subject': subject,
                    'method': method, 'comparator': comparator, 'comparison_role': comparison_role,
                    'comparator_MAE_minus_method_MAE': difference})
        summary[name] = {'n_subjects': len(error), 'methods': methods,
            'empirically_best_simple_baseline': best_simple,
            'simple_baseline_selection_candidates': SIMPLE_METHODS,
            'comparisons': [row for row in paired_rows if row['cohort'] == name]}
    pd.DataFrame(paired_rows).to_csv(out / 'fair_paired_differences.csv', index=False)
    pd.DataFrame(subject_difference_rows).to_csv(out / 'fair_subject_paired_differences.csv', index=False)
    assert {p.name: sha256(p) for p in original_paths} == original_hashes, 'Original benchmark changed'
    checks = {'passed': True, 'new_prediction_rows': len(new), 'new_methods': len(NEW_METHODS),
        'subjects_total': 86, 'module_count': len(MOD), 'repeats': N_REPEATS,
        'cohort_repeat_fold_checks': len(invariant_rows), 'same_exported_folds': True,
        'original_benchmark_files_unchanged': True, 'source_hashes_match': True,
        'all_outputs_finite': True, 'max_state_difference_from_original':
        max(r['max_state_difference_from_original'] for r in invariant_rows),
        'max_training_mean_prediction_difference':
        max(r['max_mean_followup_prediction_difference'] for r in invariant_rows)}
    dump_json(out / 'fair_output_checks.json', checks)
    summary_document = {'analysis_date': '2026-10-15', 'post_hoc_nonconfirmatory': True,
        'new_methods': NEW_METHODS, 'bootstrap_seed_base': BOOTSTRAP_SEED,
        'n_bootstrap': N_BOOTSTRAP, 'original_output_sha256': original_hashes,
        'source_sha256': source_hashes, 'script_sha256': sha256(__file__),
        'runtime': {'python': platform.python_version(), 'numpy': np.__version__, 'pandas': pd.__version__},
        'transformation': 'Original states(): training-baseline gene mean/SD, then frozen module medians',
        'group_rule': 'Training subjects sharing recorded treatment; pooled training fallback if absent',
        'uncertainty_scope': 'Subject bootstrap conditional on all fixed OOF fits and transformations. '
            'Average modules and 20 repeats within subject before resampling. '
            'Does not include refitting, tuning, representation or selected-comparator uncertainty; '
            'no multiplicity correction; not confirmatory.',
        'simple_baseline_selection_scope': 'Per-cohort smallest empirical repeated OOF MAE among '
            'listed nine simple specifications. This outcome-informed selection is descriptive '
            'and does not define a locked deployable selection strategy.',
        'results': summary, 'checks': checks}
    dump_json(out / 'summary_fair_comparators.json', summary_document)
    lines = ['METHODS INSERT (post hoc extension)',
        'In a post hoc, nonconfirmatory comparator-completeness analysis, we added pooled '
        'median follow-up, treatment-group median follow-up, pooled median change, and '
        'treatment-group median change predictors. Each used the identical exported 20-by-5 '
        'subject-held-out splits and training-baseline standardization as the original 11 '
        'specifications. Follow-up medians predict each module from training follow-up '
        'scores; median-change methods add the training median change to the held-out '
        'baseline. Group versions use only training subjects in the same recorded '
        'treatment group, with pooled fallback if that group is absent. We averaged '
        'absolute errors over modules and repeats within each subject before 5,000 paired '
        'subject bootstrap draws (seed 2026101517 for GSE86216 and 2026101518 for GSE113873). '
        'Positive differences denote comparator MAE minus model MAE. Intervals condition '
        'on the fitted predictions and exclude refitting and comparator-selection '
        'uncertainty; neither repeats nor modules were treated as independent subjects. '
        'The best simple comparator was selected descriptively from the nine listed '
        'constant, persistence, mean-change and median-change specifications.', '',
        'RESULTS INSERT']
    for name, result in summary.items():
        methods = result['methods']
        medians = '; '.join(f"{m} {methods[m]['MAE']:.4f}" for m in NEW_METHODS)
        best = result['empirically_best_simple_baseline']
        lines.append(f"{name} (n={result['n_subjects']}): {medians}. The empirically lowest-error "
                     f"simple comparator was {best} (MAE {methods[best]['MAE']:.4f}).")
        for row in result['comparisons']:
            if row['comparison_role'] in ['direct_ridge_vs_scalar', 'empirically_best_simple_baseline']:
                lines.append(f"{row['method']} versus {row['comparator']}: comparator-minus-model "
                    f"MAE difference {row['estimate']:.4f}, conditional 95% subject-bootstrap interval "
                    f"[{row['conditional_ci95_low']:.4f}, {row['conditional_ci95_high']:.4f}].")
    lines.extend(['', 'INTERPRETATION',
        'The direct ridge-versus-scalar paired interval includes zero in both cohorts. '
        'These post hoc comparisons do not establish superiority of the full matrix model '
        'over scalar shrinkage and do not constitute independent study-level external validation. '
        'The PAD median comparator results are unfavorable to the nested models; retain them. '
        'All 200 cohort/repeat/fold reconstructions passed sample identity, subject disjointness, '
        'finite-value, and original transformed-target/training-mean reconstruction checks.'])
    (out / 'fair_comparator_insert.txt').write_text('\n'.join(lines) + '\n', encoding='utf-8')
    print(json.dumps({'checks': checks, 'results': summary}, indent=2), flush=True)


if __name__ == '__main__':
    main()
```
