## Supplementary material for "VascFlux-GEO: Comparator selection and auditability in vascular transcriptome module prediction": Analysis code and aggregate results: run_known_truth_simulation.py

```
"""Post hoc known-truth comparator simulation; no patient data are used.

Run with bundled Python: python run_known_truth_simulation.py
The fixed sim_protocol.json is written and hashed BEFORE any outcome is simulated.
This is not a preregistered experiment and is not external clinical validation.
"""
import hashlib
import json
import math
import platform
from pathlib import Path

import numpy as np
import pandas as pd

ROOT = Path(__file__).resolve().parent
OUT = ROOT / "results_reviewer_revision"
P = 8
TRAIN_NS = [72, 300]
TEST_N = 2000
REPEATS = 100
SEED = 2026101531
SCALAR_GRID = [0.0, 0.25, 0.5, 0.75, 1.0]
RIDGE_GRID = [0.01, 0.1, 1.0, 10.0, 100.0, 1000.0]
METHODS = ["mean_followup", "median_followup", "mean_change", "persistence",
           "scalar_inner_selected", "ridge_inner_selected"]
SCENARIOS = ["independent_followup", "scalar_persistence",
             "off_diagonal_state", "transport_shift"]


def dump(path, obj):
    path.write_text(json.dumps(obj, indent=2, allow_nan=False), encoding="utf-8")


def sha(path):
    return hashlib.sha256(path.read_bytes()).hexdigest()


def cov(rho, scale=1.0):
    return scale ** 2 * rho ** np.abs(np.subtract.outer(np.arange(P), np.arange(P)))


def scenario(name):
    identity = np.eye(P)
    rotation = np.roll(identity, 1, axis=1)
    zero = np.zeros((P, P))
    a = {"independent_followup": zero, "scalar_persistence": 0.75 * identity,
         "off_diagonal_state": 0.1 * identity + 0.75 * rotation,
         "transport_shift": 0.75 * identity}[name]
    return {
        "train_A": a, "test_A": (-0.25 * identity if name == "transport_shift" else a),
        "train_B_mean": np.zeros(P),
        "test_B_mean": (np.full(P, 0.5) if name == "transport_shift" else np.zeros(P)),
        "train_B_cov": cov(0.4),
        "test_B_cov": (cov(0.7, 1.35) if name == "transport_shift" else cov(0.4)),
        "train_F_offset": np.zeros(P),
        "test_F_offset": (np.full(P, 0.5) if name == "transport_shift" else np.zeros(P)),
        "noise_sd": 1.0,
    }


def sample(spec, n, rng, split):
    b = rng.multivariate_normal(spec[f"{split}_B_mean"], spec[f"{split}_B_cov"], size=n)
    noise = rng.standard_normal((n, P)) * spec["noise_sd"]
    f = b @ spec[f"{split}_A"].T + spec[f"{split}_F_offset"] + noise
    return b, f


def ridge_fit(b, f, penalty):
    bm, fm = b.mean(axis=0), f.mean(axis=0)
    bc, fc = b - bm, f - fm
    coefficient = np.linalg.solve(bc.T @ bc + penalty * np.eye(P), bc.T @ fc)
    return coefficient, bm, fm


def ridge_predict(fitted, b):
    coef, bm, fm = fitted
    return (b - bm) @ coef + fm


def tune(b, f, rng):
    # Every inner split uses only training observations, including centering.
    folds = np.array_split(rng.permutation(len(b)), 5)
    scalar_losses = np.zeros(len(SCALAR_GRID))
    ridge_losses = np.zeros(len(RIDGE_GRID))
    for k, val in enumerate(folds):
        tr = np.concatenate([folds[j] for j in range(5) if j != k])
        bt, ft = b[tr], f[tr]
        bm, fm = bt.mean(axis=0), ft.mean(axis=0)
        for j, alpha in enumerate(SCALAR_GRID):
            pred = fm + alpha * (b[val] - bm)
            scalar_losses[j] += np.abs(f[val] - pred).sum()
        for j, penalty in enumerate(RIDGE_GRID):
            pred = ridge_predict(ridge_fit(bt, ft, penalty), b[val])
            ridge_losses[j] += np.abs(f[val] - pred).sum()
    # Deterministic first grid entry wins ties. No test data are available here.
    si, ri = int(np.argmin(scalar_losses)), int(np.argmin(ridge_losses))
    return SCALAR_GRID[si], RIDGE_GRID[ri], scalar_losses[si] / (len(b) * P), ridge_losses[ri] / (len(b) * P)


def aggregate(values):
    return {"mc_mean": float(np.mean(values)),
            "mc_standard_error": float(np.std(values, ddof=1) / math.sqrt(len(values))),
            "mc_sd": float(np.std(values, ddof=1)),
            "replicate_q025": float(np.quantile(values, .025)),
            "replicate_q975": float(np.quantile(values, .975)),
            "monte_carlo_repeats": len(values)}


def main():
    OUT.mkdir(exist_ok=True)
    specs = {name: scenario(name) for name in SCENARIOS}
    serial_specs = {name: {key: value.tolist() if isinstance(value, np.ndarray) else value
                          for key, value in spec.items()} for name, spec in specs.items()}
    protocol = {
        "label": "Post hoc reviewer-oriented known-truth simulation; not preregistered",
        "purpose": "Assess comparator dependence and transport failure under explicit known truths; no patient inference",
        "design_fixed_before_execution": True, "seed": SEED, "dimensions": P,
        "train_n": TRAIN_NS, "independent_test_n_per_repeat": TEST_N,
        "monte_carlo_repeats_per_cell": REPEATS, "methods": METHODS,
        "scenario_order": SCENARIOS, "data_generating_models": serial_specs,
        "formula": "F = B @ A.T + offset + independent standard Gaussian noise",
        "inner_cv": {"folds": 5, "selection_metric": "pooled validation MAE",
                     "scalar_grid": SCALAR_GRID, "ridge_grid": RIDGE_GRID,
                     "centering": "inner-training means only; full-training refit after selection",
                     "test_data_never_used_for_selection": True},
        "all_scenarios_reported": True,
        "mean_change_prediction": "B_test + mean(F_train - B_train)",
        "scalar_prediction": "mean(F_train) + alpha * (B_test - mean(B_train))",
        "ridge_objective": "sum squared residuals + lambda * squared Frobenius coefficient norm; unpenalized intercept",
        "uncertainty": "Monte Carlo standard error and empirical replicate distribution; not a patient confidence interval",
        "in_scope": "Fresh independent training/test generation and full model refitting in every replicate",
        "out_of_scope": ["external clinical validation", "true-patient bootstrap refitting", "clinical utility", "causal or device effects"],
        "analytic_null": {
            "A": "exactly zero; B and F independent in population",
            "oracle_MAE": math.sqrt(2 / math.pi),
            "mean_followup_expected_MAE": "sqrt(2/pi) * sqrt(1 + 1/n)",
            "mean_change_expected_MAE": "sqrt(2/pi) * sqrt(2 + 2/n)",
            "median_followup_expected_MAE": "No exact finite-sample formula asserted; converges to sqrt(2/pi) as n grows",
        },
    }
    protocol_path = ROOT / "sim_protocol.json"
    dump(protocol_path, protocol)
    protocol_hash = sha(protocol_path)
    rows, selections, identity_errors = [], [], []
    for si, name in enumerate(SCENARIOS):
        spec = specs[name]
        for ni, n in enumerate(TRAIN_NS):
            for r in range(REPEATS):
                children = np.random.SeedSequence([SEED, si, ni, r]).spawn(3)
                btr, ftr = sample(spec, n, np.random.default_rng(children[0]), "train")
                bte, fte = sample(spec, TEST_N, np.random.default_rng(children[1]), "test")
                alpha, penalty, scalar_cv, ridge_cv = tune(btr, ftr, np.random.default_rng(children[2]))
                mean_f, mean_b = ftr.mean(axis=0), btr.mean(axis=0)
                predictions = {
                    "mean_followup": np.broadcast_to(mean_f, fte.shape),
                    "median_followup": np.broadcast_to(np.median(ftr, axis=0), fte.shape),
                    "mean_change": bte + np.mean(ftr - btr, axis=0),
                    "persistence": bte,
                    "scalar_inner_selected": mean_f + alpha * (bte - mean_b),
                    "ridge_inner_selected": ridge_predict(ridge_fit(btr, ftr, penalty), bte),
                }
                oracle = bte @ spec["test_A"].T + spec["test_F_offset"]
                oracle_mae = float(np.abs(fte - oracle).mean())
                for method, pred in predictions.items():
                    assert np.isfinite(pred).all()
                    followup_error = fte - pred
                    change_error = (fte - bte) - (pred - bte)
                    invariant_error = float(np.max(np.abs(followup_error - change_error)))
                    identity_errors.append(invariant_error)
                    rows.append({"scenario": name, "train_n": n, "test_n": TEST_N,
                                 "repeat": r, "method": method,
                                 "MAE": float(np.abs(followup_error).mean()),
                                 "RMSE": float(np.sqrt(np.mean(followup_error ** 2))),
                                 "test_population_oracle_MAE": oracle_mae,
                                 "max_followup_change_residual_difference": invariant_error})
                selections.append({"scenario": name, "train_n": n, "repeat": r,
                                   "selected_scalar": alpha, "selected_ridge_penalty": penalty,
                                   "selected_scalar_inner_MAE": scalar_cv,
                                   "selected_ridge_inner_MAE": ridge_cv})
            print(f"{name}, n={n}: {REPEATS} independent train/test refits complete", flush=True)
    results = pd.DataFrame(rows)
    selection = pd.DataFrame(selections)
    results.to_csv(OUT / "simulation_replicate_metrics.csv", index=False)
    selection.to_csv(OUT / "simulation_training_selection.csv", index=False)
    summaries = []
    for (name, n, method), group in results.groupby(["scenario", "train_n", "method"], sort=False):
        summaries.append({"scenario": name, "train_n": n, "method": method,
                          **aggregate(group.MAE.to_numpy())})
    means = pd.DataFrame(summaries)
    means.to_csv(OUT / "simulation_summary_metrics.csv", index=False)
    pair_rows = []
    for (name, n), group in results.groupby(["scenario", "train_n"], sort=False):
        pivot = group.pivot(index="repeat", columns="method", values="MAE")
        for comparator, method in [("mean_change", "mean_followup"),
                                   ("mean_followup", "median_followup"),
                                   ("mean_followup", "scalar_inner_selected"),
                                   ("mean_followup", "ridge_inner_selected"),
                                   ("scalar_inner_selected", "ridge_inner_selected")]:
            diff = pivot[comparator] - pivot[method]
            pair_rows.append({"scenario": name, "train_n": n, "comparator": comparator,
                              "method": method, "estimand": "comparator_MAE_minus_method_MAE",
                              **aggregate(diff.to_numpy()), "fraction_replicates_method_lower_MAE": float((diff > 0).mean())})
    paired = pd.DataFrame(pair_rows)
    paired.to_csv(OUT / "simulation_paired_differences.csv", index=False)
    expected = []
    for n in TRAIN_NS:
        for method, variance in [("mean_followup", 1 + 1 / n), ("mean_change", 2 + 2 / n)]:
            row = means[(means.scenario == "independent_followup") & (means.train_n == n) & (means.method == method)].iloc[0]
            truth = math.sqrt(2 / math.pi) * math.sqrt(variance)
            tolerance = 5 * float(row.mc_standard_error) + .002
            expected.append({"scenario": "independent_followup", "train_n": n,
                             "method": method, "analytic_expected_MAE": truth,
                             "observed_MC_mean": float(row.mc_mean),
                             "absolute_difference": abs(float(row.mc_mean) - truth),
                             "tolerance_5_MCSE_plus_0p002": tolerance,
                             "passed": bool(abs(float(row.mc_mean) - truth) < tolerance)})
    pd.DataFrame(expected).to_csv(OUT / "simulation_analytic_null_checks.csv", index=False)
    assert len(results) == len(SCENARIOS) * len(TRAIN_NS) * REPEATS * len(METHODS) == 4800
    assert len(selection) == 800
    assert not results.duplicated(["scenario", "train_n", "repeat", "method"]).any()
    assert np.isfinite(results.select_dtypes(include="number")).all().all()
    assert max(identity_errors) < 1e-12
    assert all(row["passed"] for row in expected), "Null Monte Carlo result differs unexpectedly from analytic expectation"
    assert np.count_nonzero(specs["independent_followup"]["train_A"]) == 0
    assert np.count_nonzero(specs["independent_followup"]["test_A"]) == 0
    assert sha(protocol_path) == protocol_hash, "Protocol changed during simulation"
    summary = {
        "status": "passed", "analysis_type": "post hoc known-truth simulation, not preregistered",
        "protocol_sha256": protocol_hash, "protocol_path": "sim_protocol.json",
        "script_sha256": sha(Path(__file__)), "python": platform.python_version(),
        "numpy": np.__version__, "pandas": pd.__version__, "seed": SEED,
        "scenarios": SCENARIOS, "train_ns": TRAIN_NS, "test_n": TEST_N,
        "repeats_per_cell": REPEATS, "complete_train_test_refits": 800,
        "six_method_metric_rows": 4800, "inner_training_cv_only": True,
        "all_scenarios_and_repeats_reported": True,
        "analytic_zero_signal_checks": expected,
        "max_prediction_change_identity_error": max(identity_errors),
        "finite_outputs": True,
        "uncertainty_label": "MCSE and replicate quantiles; not patient confidence intervals",
        "all_metrics": summaries, "all_paired_differences": pair_rows,
        "does_not_resolve": ["true longitudinal external cohort validation",
                             "real-patient training-sample/refit uncertainty", "causal inference"],
    }
    dump(OUT / "summary_known_truth_simulation.json", summary)
    def cell(s, n, m):
        row = means[(means.scenario == s) & (means.train_n == n) & (means.method == m)].iloc[0]
        return f"{row.mc_mean:.4f} (Monte Carlo SE {row.mc_standard_error:.4f})"
    insertion = (
        "METHODS INSERT: Post hoc known-truth comparator simulation\n"
        "We added a reviewer-oriented simulation after examining the real-data results; it was not preregistered. "
        "The full design was saved to sim_protocol.json before execution. Eight correlated Gaussian baseline states "
        "were generated under four explicit scenarios: independent follow-up (no temporal signal), scalar persistence, "
        "off-diagonal state dependence, and joint covariate/relationship transport shift. Each scenario used 100 independent "
        "training/test replicates at training n=72 and n=300, with an independently generated test n=2,000. "
        "Mean follow-up, median follow-up, mean change, persistence, scalar shrinkage, and multivariate ridge were compared. "
        "Scalar and ridge hyperparameters were selected by five-fold training-only MAE and refitted before test evaluation. "
        "All scenarios and repeats are retained. Uncertainty is summarized as Monte Carlo standard error and replicate "
        "quantiles, not patient confidence intervals. Full refitting here describes the simulation only and does not "
        "repair conditional uncertainty in the real-data analysis.\n\n"
        "RESULTS INSERT: Known-truth comparator behavior\n"
        f"With no true baseline-to-follow-up signal and n=72, MAE was {cell('independent_followup',72,'mean_followup')} "
        f"for mean follow-up and {cell('independent_followup',72,'mean_change')} for mean change. "
        f"At n=300, mean and median follow-up errors were {cell('independent_followup',300,'mean_followup')} and "
        f"{cell('independent_followup',300,'median_followup')}, respectively, near the Gaussian population oracle "
        f"MAE of {math.sqrt(2/math.pi):.4f}. Thus, improvement over mean change can occur without any temporal signal. "
        f"For scalar-persistence truth at n=300, scalar and ridge MAE were {cell('scalar_persistence',300,'scalar_inner_selected')} "
        f"and {cell('scalar_persistence',300,'ridge_inner_selected')}; for off-diagonal truth they were "
        f"{cell('off_diagonal_state',300,'scalar_inner_selected')} and {cell('off_diagonal_state',300,'ridge_inner_selected')}. "
        f"Under the fixed transport-shift scenario at n=300, mean follow-up, scalar and ridge MAE were "
        f"{cell('transport_shift',300,'mean_followup')}, {cell('transport_shift',300,'scalar_inner_selected')}, and "
        f"{cell('transport_shift',300,'ridge_inner_selected')}. These deliberately controlled settings identify "
        "conditions under which full-matrix structure is useful and illustrate failure after transport; they do not "
        "establish the truth of those conditions in the observed vascular cohorts. Complete 48-cell results and "
        "40 paired contrasts are exported, including all n=72 results. Analytic null expectations and the "
        "follow-up/change residual identity passed numerical checks.\n\n"
        "LIMITATION: The simulation is methodological evidence about comparator dependence, not a substitute for "
        "an independent longitudinal cohort, biological validation, or clinical utility evidence. The four scenarios "
        "are illustrative and are not an exhaustive robustness envelope.\n"
    )
    (OUT / "simulation_insert.txt").write_text(insertion, encoding="utf-8")
    print(json.dumps({"status": "passed", "refits": 800, "metrics": 4800,
                      "protocol_sha256": protocol_hash,
                      "max_residual_identity_error": max(identity_errors)}, indent=2))


if __name__ == "__main__":
    main()
```
