## Supplementary material for "VascFlux-GEO: Comparator selection and auditability in vascular transcriptome module prediction": Analysis code and aggregate results: run_representation_sensitivity.py

```
"""Post-hoc annotation, target-weight, and full nested leakage checks.

Run: python run_representation_sensitivity.py --data-dir ../data
Only new robustness_* files, summary_representation_sensitivity.json, and
robustness_insert.txt in results_reviewer_revision are written. No original
benchmark code, manuscript, or CMPB submission is changed.
"""
import argparse
import copy
import gzip
import hashlib
import json
import platform
import re
from pathlib import Path

import numpy as np
import pandas as pd
import run_extended_benchmarks as core

ROOT = Path(__file__).resolve().parent
N_REPEATS = 5
METHODS = ['mean_followup', 'median_followup', 'followup_ridge_nested',
           'shrunken_state_followup_nested']
SPECS = [('GSE86216', 'GPL14951_family.soft.gz', 'sibship (sampleid)', 'time', 'treatment'),
         ('GSE113873', 'GPL15207_full2.txt', 'subject', 'visit', 'treatment group')]


def sha256(path):
    hh = hashlib.sha256()
    with path.open('rb') as handle:
        while chunk := handle.read(8 * 1024 * 1024):
            hh.update(chunk)
    return hh.hexdigest()


def unique_symbol_mapping(path):
    """Same token parser as frozen pipeline, retain only one distinct symbol.

    This does not claim updated HGNC canonicalization. Ambiguity is evaluated
    over all parsed symbols, BEFORE restricting to the frozen module dictionary.
    Multiple tokens repeating the same symbol are not considered ambiguous.
    """
    opener = gzip.open if str(path).endswith('.gz') else open
    mapping, total, ambiguous, eligible_ambiguous, active = {}, 0, 0, 0, False
    with opener(path, 'rt', errors='replace') as handle:
        for line in handle:
            if line.startswith('!platform_table_begin'):
                cols = {key: i for i, key in enumerate(next(handle).rstrip('\n').split('\t'))}
                symcol = next(key for key in ['Gene Symbol', 'Gene symbol', 'Symbol',
                    'Gene Symbol (human)', 'gene_assignment'] if key in cols)
                active = True
                continue
            if line.startswith('!platform_table_end'):
                break
            if not active or not line.strip():
                continue
            fields = line.rstrip('\n').split('\t')
            if len(fields) <= max(cols['ID'], cols[symcol]):
                continue
            total += 1
            raw = fields[cols[symcol]].strip()
            if symcol == 'gene_assignment':
                symbols = [part.split('//')[1].strip() for part in raw.split('///')
                    if len(part.split('//')) >= 2 and part.split('//')[1].strip()
                    and part.split('//')[1].strip() not in ('---', '--- //')]
            else:
                symbols = [part.strip() for part in re.split(r'[;|, ]+', raw.replace('///', ';'))
                           if part.strip() and part.strip() not in ('---', 'NA', 'null', 'nan')]
            distinct = set(symbols)
            ambiguous += int(len(distinct) > 1)
            eligible_ambiguous += int(len(distinct) > 1 and bool(distinct & core.TARGET))
            if len(distinct) == 1:
                symbol = next(iter(distinct))
                if symbol in core.TARGET:
                    mapping[fields[cols['ID']].strip()] = symbol
    return mapping, ambiguous, {'platform_rows_parsed': total,
        'platform_ambiguous_rows': ambiguous,
        'ambiguous_rows_intersecting_module_genes': eligible_ambiguous,
        'unique_symbol_target_probes': len(mapping),
        'unique_symbol_target_genes': len(set(mapping.values()))}


def load_schemes(data, source_manifest=ROOT / 'source_cache_manifest.json'):
    original_reader = core.read_platform
    schemes, coverage, provenance = {}, [], []
    selected_manifest = core.load_source_manifest(source_manifest)
    expected = {name: row['sha256'] for name, row in selected_manifest.items()}
    for gse, plat, *_ in SPECS:
        for filename in [plat, f'{gse}_series_matrix.txt.gz']:
            path = data / filename
            digest = sha256(path)
            assert digest == expected[filename], f'Input hash mismatch: {filename}'
            provenance.append({'filename': filename, 'sha256': digest,
                               'bytes': path.stat().st_size, 'matches_selected_manifest': True,
                               'manifest_filename': Path(source_manifest).name,
                               'input_kind': selected_manifest[filename].get('input_kind', 'frozen_original_input')})
    for gse, plat, subject, visit, treatment in SPECS:
        preferred, ambig = original_reader(data / plat)
        unique, _, audit = unique_symbol_mapping(data / plat)
        for scheme, mapping in [('module_preferred', preferred), ('unique_symbol_only', unique)]:
            # Local dependency injection solely to reuse the frozen pairing and
            # probe-to-gene median code; original source file remains unchanged.
            core.read_platform = lambda _path, mm=mapping, aa=ambig: (mm, aa)
            try:
                cohort = core.load_cohort(data, gse, plat, subject, visit, treatment)
            finally:
                core.read_platform = original_reader
            assert len(cohort['records']) == (72 if gse == 'GSE86216' else 14)
            cohort['mapping_scheme'] = scheme
            schemes[(gse, scheme)] = cohort
            for module, count in cohort['audit']['module_gene_counts'].items():
                coverage.append({'cohort': gse, 'mapping_scheme': scheme, 'platform': plat,
                    'module': module, 'frozen_genes': len(core.MODULES[module]),
                    'observed_genes': count, 'union_observed_genes': cohort['audit']['n_module_gene_union'],
                    'retained_target_probes_in_platform': len(mapping), **audit})
        assert schemes[(gse, 'module_preferred')]['records'] == schemes[(gse, 'unique_symbol_only')]['records']
    return schemes, coverage, provenance


def fold_plan(schemes):
    rng = np.random.default_rng(core.SEEDS['outer_first'])
    first = {}
    for gse, *_ in SPECS:
        idx = np.arange(len(schemes[(gse, 'module_preferred')]['records']))
        rng.shuffle(idx)
        first[gse] = list(np.array_split(idx, 5))
    plan = {}
    for ci, (gse, *_) in enumerate(SPECS):
        n = len(schemes[(gse, 'module_preferred')]['records'])
        for repeat in range(N_REPEATS):
            seed = core.SEEDS['outer_first'] if repeat == 0 else core.SEEDS['outer_repeats_start'] + repeat + 100 * ci
            folds = first[gse] if repeat == 0 else core.splits(n, seed)
            for fold, test in enumerate(folds):
                train = np.concatenate([part for j, part in enumerate(folds) if j != fold])
                assert not set(train) & set(test)
                plan[(gse, repeat, fold)] = (train, test, core.SEEDS['inner'] + ci * 10000 + repeat * 10 + fold)
    return plan


def predict_selected(cohort, train, test, params):
    baseline, followup, mu, sd = core.states(cohort, train)
    x, xe = core.features(cohort, baseline, train, test, True)
    predictions = {
        'mean_followup': np.tile(followup[train].mean(0), (len(test), 1)),
        'median_followup': np.tile(np.median(followup[train], axis=0), (len(test), 1)),
        'followup_ridge_nested': xe @ core.ridge(x, followup[train], params['followup']),
        'shrunken_state_followup_nested': followup[train].mean(0) + params['shrunken'] *
                                        (baseline[test] - baseline[train].mean(0))}
    assert all(np.isfinite(value).all() for value in predictions.values())
    return predictions, baseline, followup, mu, sd


def run_mapping_fits(schemes, plan):
    oof, tuning, splits = [], [], []
    for (gse, scheme), cohort in schemes.items():
        print(f'{gse} / {scheme}: 5 x 5 outer folds with full inner tuning', flush=True)
        for repeat in range(N_REPEATS):
            for fold in range(5):
                train, test, inner_seed = plan[(gse, repeat, fold)]
                cache = core.inner_cache(cohort, train, inner_seed)
                params = {}
                for kind, grid in [('followup', core.GRID), ('shrunken', core.SHRINK_GRID)]:
                    params[kind], losses = core.tune(cohort, cache, kind, grid)
                    assert np.isfinite(losses).all()
                    for candidate, loss in zip(grid, losses):
                        tuning.append({'cohort': gse, 'mapping_scheme': scheme, 'repeat': repeat, 'fold': fold,
                            'kind': kind, 'candidate': candidate, 'inner_MAE': loss,
                            'selected': bool(candidate == params[kind]), 'inner_folds': len(cache)})
                predictions, baseline, followup, _, _ = predict_selected(cohort, train, test, params)
                for j, subject_idx in enumerate(test):
                    subject = cohort['records'][subject_idx]['subject']
                    splits.append({'cohort': gse, 'mapping_scheme': scheme, 'repeat': repeat,
                                   'fold': fold, 'subject': subject, 'inner_seed': inner_seed})
                    for method, prediction in predictions.items():
                        for mi, module in enumerate(core.MOD):
                            oof.append({'cohort': gse, 'mapping_scheme': scheme, 'repeat': repeat,
                                'fold': fold, 'subject': subject, 'module': module, 'method': method,
                                'observed_followup': followup[subject_idx, mi],
                                'predicted_followup': prediction[j, mi],
                                'absolute_error': abs(followup[subject_idx, mi] - prediction[j, mi])})
    result = pd.DataFrame(oof)
    assert len(result) == 2 * 5 * (72 + 14) * 8 * len(METHODS)
    assert not result.duplicated(['cohort', 'mapping_scheme', 'repeat', 'subject', 'module', 'method']).any()
    assert np.isfinite(result[['observed_followup', 'predicted_followup', 'absolute_error']]).all().all()
    return result, tuning, splits


def summarize_mapping(oof):
    rows = []
    for (gse, scheme), sub in oof.groupby(['cohort', 'mapping_scheme'], sort=False):
        err = sub.groupby(['subject', 'method']).absolute_error.mean().unstack('method')
        n = len(err)
        rng = np.random.default_rng(2026101506)
        ix = rng.integers(0, n, (5000, n))
        for method in METHODS:
            for comparator in ['mean_followup', 'median_followup']:
                diff = err[comparator].to_numpy() - err[method].to_numpy()
                q = np.quantile(diff[ix].mean(1), [.025, .975])
                rows.append({'cohort': gse, 'mapping_scheme': scheme, 'method': method,
                    'comparator': comparator, 'n_subjects': n, 'n_repeats': N_REPEATS,
                    'method_MAE': err[method].mean(), 'comparator_MAE': err[comparator].mean(),
                    'within_scheme_MAE_improvement': diff.mean(),
                    'within_scheme_fractional_MAE_improvement': diff.mean() / err[comparator].mean(),
                    'conditional_subject_ci95_low': q[0], 'conditional_subject_ci95_high': q[1],
                    'bootstrap_draws': 5000,
                    'uncertainty_scope': 'conditional on fixed fitted OOF predictions; no bootstrap refitting',
                    'interpretation': 'compare improvement within mapping; mapping changes target definition'})
    return pd.DataFrame(rows)


def target_weight_sensitivity(oof):
    rows = []
    # A scoring sensitivity, not an input/target ablation: predictions, all
    # training targets, the fitted models, and the hyperparameters remain fixed.
    for (gse, scheme), sub in oof.groupby(['cohort', 'mapping_scheme'], sort=False):
        for dropped in ['none'] + core.MOD:
            selected = sub if dropped == 'none' else sub[sub.module != dropped]
            errors = selected.groupby(['subject', 'method']).absolute_error.mean().unstack('method')
            for method in METHODS:
                for comparator in ['mean_followup', 'median_followup']:
                    rows.append({'cohort': gse, 'mapping_scheme': scheme, 'zero_weight_target_module': dropped,
                        'remaining_target_modules': 8 if dropped == 'none' else 7,
                        'method': method, 'comparator': comparator, 'n_subjects': len(errors),
                        'n_repeats': N_REPEATS, 'method_MAE': errors[method].mean(),
                        'within_scheme_MAE_improvement': (errors[comparator] - errors[method]).mean(),
                        'model_refitted': False, 'input_module_removed': False,
                        'scope': 'fixed OOF target weighting only; not predictive module ablation'})
    assert len(rows) == 2 * 2 * 9 * 4 * 2
    return pd.DataFrame(rows)


def input_module_ablation(schemes, plan, full_oof):
    """Actual nested refitting, original mapping only, all eight targets fixed."""
    oof_rows, tuning_rows = [], []
    for gse, *_ in SPECS:
        cohort = schemes[(gse, 'module_preferred')]
        print(f'{gse}: nested ridge input-module deletion, all 8 targets retained', flush=True)
        for repeat in range(N_REPEATS):
            for fold in range(5):
                train, test, inner_seed = plan[(gse, repeat, fold)]
                cache = core.inner_cache(cohort, train, inner_seed)
                baseline, followup, _, _ = core.states(cohort, train)
                outer_x, outer_xe = core.features(cohort, baseline, train, test, True)
                for mi, module in enumerate(core.MOD):
                    # Intercept is column 0; module columns 1..8 precede
                    # prespecified treatment columns. Targets are never removed.
                    columns = [j for j in range(outer_x.shape[1]) if j != mi + 1]
                    candidate_losses = []
                    for lam in core.GRID:
                        errors = []
                        for it, iv, bb, ff in cache:
                            x, xe = core.features(cohort, bb, it, iv, True)
                            prediction = xe[:, columns] @ core.ridge(x[:, columns], ff[it], lam)
                            errors.extend(np.abs(ff[iv] - prediction).mean(1))
                        candidate_losses.append(float(np.mean(errors)))
                    assert np.isfinite(candidate_losses).all()
                    selected = float(core.GRID[np.argmin(candidate_losses)])
                    for candidate, loss in zip(core.GRID, candidate_losses):
                        tuning_rows.append({'cohort': gse, 'removed_input_module': module,
                            'repeat': repeat, 'fold': fold, 'candidate': candidate,
                            'inner_MAE_all_eight_targets': loss, 'selected': candidate == selected})
                    pred = outer_xe[:, columns] @ core.ridge(outer_x[:, columns], followup[train], selected)
                    assert np.isfinite(pred).all()
                    for jj, subject_idx in enumerate(test):
                        for target_idx, target in enumerate(core.MOD):
                            oof_rows.append({'cohort': gse, 'mapping_scheme': 'module_preferred',
                                'removed_input_module': module, 'repeat': repeat, 'fold': fold,
                                'subject': cohort['records'][subject_idx]['subject'], 'target_module': target,
                                'selected_lambda': selected, 'observed_followup': followup[subject_idx, target_idx],
                                'predicted_followup': pred[jj, target_idx],
                                'absolute_error': abs(followup[subject_idx, target_idx] - pred[jj, target_idx])})
    frame = pd.DataFrame(oof_rows)
    assert len(frame) == (72 + 14) * 5 * 8 * 8
    assert not frame.duplicated(['cohort', 'removed_input_module', 'repeat', 'subject', 'target_module']).any()
    summary_rows = []
    for (gse, removed), sub in frame.groupby(['cohort', 'removed_input_module'], sort=False):
        err = sub.groupby('subject').absolute_error.mean()
        reference = full_oof[(full_oof.cohort == gse) &
                            (full_oof.mapping_scheme == 'module_preferred')].groupby(
                                ['subject', 'method']).absolute_error.mean().unstack('method')
        assert err.index.equals(reference.index)
        for comparator in ['mean_followup', 'median_followup', 'followup_ridge_nested']:
            diff = reference[comparator].to_numpy() - err.to_numpy()
            summary_rows.append({'cohort': gse, 'mapping_scheme': 'module_preferred',
                'removed_input_module': removed, 'retained_input_modules': 7, 'retained_target_modules': 8,
                'comparator': comparator, 'n_subjects': len(err), 'n_repeats': N_REPEATS,
                'model_refitted': True, 'complete_inner_tuning_rerun': True,
                'ablated_MAE': err.mean(), 'comparator_MAE': reference[comparator].mean(),
                'comparator_MAE_minus_ablated_MAE': diff.mean(),
                'scope': 'exploratory input sensitivity, not gene importance or biological necessity'})
    return frame, tuning_rows, pd.DataFrame(summary_rows)


def full_nested_future_mutation(schemes, plan):
    records, candidates = [], []
    for (gse, scheme), cohort in schemes.items():
        for fold in [0, 4]:
            train, test, inner_seed = plan[(gse, 0, fold)]
            altered = copy.copy(cohort)
            altered['raw'] = cohort['raw'].copy()
            # Only outer heldout followup expression is changed. The same train
            # membership and inner splits are recomputed from scratch below.
            original_values = altered['raw'][:, cohort['follow'][test]].copy()
            altered['raw'][:, cohort['follow'][test]] = original_values * -7.0 + 1000.0
            assert np.array_equal(altered['raw'][:, cohort['base']], cohort['raw'][:, cohort['base']])
            assert np.array_equal(altered['raw'][:, cohort['follow'][train]], cohort['raw'][:, cohort['follow'][train]])
            before, after, max_candidate_difference = {}, {}, 0.
            caches = [core.inner_cache(cc, train, inner_seed) for cc in [cohort, altered]]
            for kind, grid in [('delta', core.GRID), ('followup', core.GRID),
                               ('pca2', core.GRID), ('shrunken', core.SHRINK_GRID)]:
                before[kind], loss0 = core.tune(cohort, caches[0], kind, grid)
                after[kind], loss1 = core.tune(altered, caches[1], kind, grid)
                assert np.isfinite(loss0).all() and np.isfinite(loss1).all()
                diff = float(np.max(np.abs(np.array(loss0) - np.array(loss1))))
                max_candidate_difference = max(max_candidate_difference, diff)
                assert before[kind] == after[kind] and diff < 1e-12
                for value, one, two in zip(grid, loss0, loss1):
                    candidates.append({'cohort': gse, 'mapping_scheme': scheme, 'repeat': 0, 'fold': fold,
                        'kind': kind, 'candidate': value, 'before_inner_MAE': one,
                        'after_inner_MAE': two, 'absolute_difference': abs(one - two),
                        'selected_before': bool(value == before[kind]),
                        'selected_after': bool(value == after[kind]), 'inner_folds': len(caches[0])})
            pp0, b0, f0 = core.predict_all(cohort, train, test, before)
            pp1, b1, f1 = core.predict_all(altered, train, test, after)
            _, _, mu0, sd0 = core.states(cohort, train)
            _, _, mu1, sd1 = core.states(altered, train)
            max_prediction_difference = max(float(np.max(np.abs(pp0[method] - pp1[method]))) for method in pp0)
            future_change = float(np.max(np.abs(f0[test] - f1[test])))
            assert max_prediction_difference < 1e-12 and future_change > 1
            assert np.array_equal(mu0, mu1) and np.array_equal(sd0, sd1)
            assert np.array_equal(b0, b1) and np.array_equal(f0[train], f1[train])
            records.append({'cohort': gse, 'mapping_scheme': scheme, 'repeat': 0, 'fold': fold,
                'n_outer_train': len(train), 'n_outer_test': len(test), 'inner_folds': len(caches[0]),
                'all_four_tuning_families_refit': True, 'number_candidate_losses_per_run': 23,
                'selected_hyperparameters_unchanged': before == after,
                'max_candidate_loss_difference': max_candidate_difference,
                'number_prediction_methods_checked': len(pp0),
                'max_prediction_difference': max_prediction_difference,
                'heldout_followup_score_max_difference': future_change,
                'train_gene_scalers_unchanged': True, 'passed': True,
                'scope': 'representative outer-fold full-inner-tuning invariance check, not universal leakage proof'})
    assert len(records) == 8 and len(candidates) == 8 * 23
    return records, candidates


def main():
    parser = argparse.ArgumentParser()
    parser.add_argument('--data-dir', type=Path, default=ROOT.parent / 'data')
    parser.add_argument('--output-dir', type=Path, default=ROOT / 'results_reviewer_revision')
    parser.add_argument('--source-manifest', type=Path, default=ROOT / 'source_cache_manifest.json')
    args = parser.parse_args()
    args.output_dir.mkdir(exist_ok=True, parents=True)
    schemes, coverage, provenance = load_schemes(args.data_dir, args.source_manifest)
    plan = fold_plan(schemes)
    oof, tuning, split_rows = run_mapping_fits(schemes, plan)
    summary = summarize_mapping(oof)
    weight = target_weight_sensitivity(oof)
    input_oof, input_tuning, input_summary = input_module_ablation(schemes, plan, oof)
    invariance, candidate_checks = full_nested_future_mutation(schemes, plan)
    baseline = pd.read_csv(ROOT / 'results_extended' / 'oof_predictions.csv')
    match = oof[(oof.mapping_scheme == 'module_preferred') & (oof.method != 'median_followup')].merge(
        baseline[baseline.repeat < N_REPEATS],
        on=['cohort', 'repeat', 'fold', 'subject', 'module', 'method'],
        suffixes=('_sensitivity', '_original'), validate='one_to_one')
    expected = (72 + 14) * 5 * 8 * 3
    assert len(match) == expected
    reproduction = float(np.max(np.abs(match.predicted_followup_sensitivity - match.predicted_followup_original)))
    assert reproduction < 1e-10
    outputs = {
        'robustness_annotation_coverage.csv': coverage,
        'robustness_annotation_oof.csv': oof,
        'robustness_annotation_tuning.csv': tuning,
        'robustness_annotation_splits.csv': split_rows,
        'robustness_annotation_summary.csv': summary,
        'robustness_target_weight_sensitivity.csv': weight,
        'robustness_input_module_ablation_oof.csv': input_oof,
        'robustness_input_module_ablation_tuning.csv': input_tuning,
        'robustness_input_module_ablation_summary.csv': input_summary,
        'robustness_full_nested_future_mutation.csv': invariance,
        'robustness_full_nested_future_mutation_candidates.csv': candidate_checks}
    for filename, obj in outputs.items():
        frame = obj if isinstance(obj, pd.DataFrame) else pd.DataFrame(obj)
        assert not frame.empty
        numeric = frame.select_dtypes(include='number')
        assert np.isfinite(numeric).all().all(), filename
        frame.to_csv(args.output_dir / filename, index=False)
    compact = summary[summary.method.isin(METHODS[2:]) & (summary.comparator == 'mean_followup')].to_dict(orient='records')
    scope = [
        'Post-hoc exploratory robustness checks, not preregistered or confirmatory analyses.',
        'Only the first five of the frozen twenty repeat plans were refitted for two annotation schemes.',
        'Unique annotation means one distinct parsed platform symbol before module filtering; no new HGNC synonym curation.',
        'Probe rules change both input representation and target definition. Absolute MAE is not directly comparable between rules; compare each model with its within-rule baseline.',
        'Target leave-one-module-out only reweights fixed OOF errors: models were not refitted and input modules were not removed.',
        'The separate input-module deletion analysis actually refits and retunes ridge after each of eight input removals, original mapping only, retaining all eight target modules. Correlated overlapping modules prevent a gene-importance or biological-necessity interpretation.',
        'Full nested future-outcome mutation covers eight representative outer fits (two folds per cohort and mapping); it is an executable scoped invariance check, not proof against all possible leakage.',
        'Subject bootstrap intervals condition on fitted OOF predictions; they omit refitting uncertainty and do not support a new confirmatory claim.',
        'No independent longitudinal study, clinical outcome, intervention effect, or device validation is supplied by these checks.']
    record = {'status': 'passed', 'analysis_date': '2026-10-15', 'post_hoc_exploratory': True,
        'n_repeats': 5, 'outer_folds': 5, 'mapping_schemes': ['module_preferred', 'unique_symbol_only'],
        'methods': METHODS, 'input_provenance': provenance,
        'runtime': {'python': platform.python_version(), 'numpy': np.__version__, 'pandas': pd.__version__},
        'mapping_comparisons': compact, 'module_coverage': coverage,
        'unique_oof_rows': len(oof), 'duplicate_oof_keys': 0, 'all_outputs_finite': True,
        'input_module_ablation': {'nested_outer_fits': 2 * 5 * 5 * 8,
            'n_oof_module_predictions': len(input_oof), 'all_eight_targets_retained': True,
            'summary': input_summary.to_dict(orient='records')},
        'frozen_OOF_reproduction': {'n_compared_predictions': expected, 'max_absolute_difference': reproduction},
        'future_mutation': {'complete_nested_runs_checked': len(invariance),
            'candidate_loss_comparisons': len(candidate_checks), 'all_passed': True,
            'max_candidate_loss_difference': max(x['max_candidate_loss_difference'] for x in invariance),
            'max_prediction_difference': max(x['max_prediction_difference'] for x in invariance)},
        'limitations': scope,
        'script_sha256': sha256(Path(__file__)),
        'outputs': {filename: sha256(args.output_dir / filename) for filename in outputs}}
    (args.output_dir / 'summary_representation_sensitivity.json').write_text(
        json.dumps(record, indent=2, allow_nan=False), encoding='utf-8')
    lines = [
        'POST-HOC EXPLORATORY REPRESENTATION ROBUSTNESS — INSERTION DRAFT',
        'Methods. Two probe-to-gene rules were compared using the same first five repeated five-fold subject splits: the frozen module-preferred mapping and a rule retaining only probes with one distinct parsed symbol before module filtering. Probe-median gene aggregation, training-baseline gene standardization, eight module targets, and inner folds were otherwise unchanged. Each scheme refitted nested direct-follow-up ridge and scalar shrinkage, together with training-only mean and componentwise-median follow-up baselines. Because annotation changes the target definition as well as the inputs, interpretation is based on improvement over a within-scheme comparator, not cross-scheme absolute MAE.',
        'Results.']
    for row in compact:
        lines.append(f"{row['cohort']} / {row['mapping_scheme']} / {row['method']}: mean-follow-up MAE improvement {row['within_scheme_MAE_improvement']:.6f} ({100*row['within_scheme_fractional_MAE_improvement']:.2f}%); conditional subject-bootstrap interval [{row['conditional_subject_ci95_low']:.6f}, {row['conditional_subject_ci95_high']:.6f}].")
    for row in summary[(summary.method == 'followup_ridge_nested') & (summary.comparator == 'median_followup')].to_dict(orient='records'):
        lines.append(f"Median comparator: {row['cohort']} / {row['mapping_scheme']}, ridge MAE improvement over the componentwise training-median baseline is {row['within_scheme_MAE_improvement']:.6f}. Negative values favor the median baseline; the PAD model does not outperform this baseline.")
    lines.append('Coverage. GSE86216 retained 141 module-target probes and 83 of 84 union genes under both annotation rules because no ambiguous platform probe intersected a module gene. GSE113873 retained 290 probes and 84 union genes under the frozen rule versus 282 probes and 83 union genes under the unique-symbol rule; oxidative_stress coverage changed from 9 to 8 genes. Thus the statin annotation sensitivity is an identity control, while the PAD comparison exercises a changed representation and target.')
    for (gse, scheme, method), ss in weight[(weight.zero_weight_target_module != 'none') &
            weight.method.isin(METHODS[2:]) & (weight.comparator == 'mean_followup')].groupby(['cohort', 'mapping_scheme', 'method']):
        lines.append(f"Fixed-OOF target-weight sensitivity: {gse} / {scheme} / {method}, leaving one module out of the loss gave within-scheme MAE improvements from {ss.within_scheme_MAE_improvement.min():.6f} to {ss.within_scheme_MAE_improvement.max():.6f}; this did not refit any model or remove any input module.")
    lines.extend([
        'Separate input ablation. On the frozen original annotation, each baseline module was removed from the model input in turn while all eight follow-up target modules remained fixed. Ridge was fully retuned within every outer training fold across the same five repeats (400 outer fits; 27,520 OOF target predictions). This differs from the fixed-OOF target-weight check above. Overlapping and correlated modules limit interpretation to predictive sensitivity, not biological necessity.',
        f'Leakage invariance. In {len(invariance)} representative outer fits (folds 0 and 4 from repeat 0 for both cohorts and both annotation schemes), only outer-held-out follow-up expression was perturbed. Complete inner tuning was rerun for all four original tuned families. All 184 candidate-loss comparisons, selected hyperparameters, and predictions from 11 original methods were unchanged within 1e-12, while held-out follow-up targets changed. This scopes the audit to future-target use in these exercised paths.',
        f'Reproduction and validity. The frozen mapping reproduced {expected} original OOF module predictions with maximum absolute difference {reproduction:.3g}. All {len(oof)} newly fitted OOF rows were uniquely keyed and finite. These are post-hoc checks; neither the conditional intervals nor the target-weight sensitivity establishes independent longitudinal validity.',
        'See robustness_annotation_coverage.csv for per-module gene coverage and robustness_annotation_summary.csv for comparisons with the median baseline.'])
    for gse, subset in input_summary[input_summary.comparator == 'followup_ridge_nested'].groupby('cohort'):
        lines.append(f"Actual input deletion: {gse}, full-input MAE minus ablated-input MAE ranges from {subset.comparator_MAE_minus_ablated_MAE.min():.6f} to {subset.comparator_MAE_minus_ablated_MAE.max():.6f}; positive values favor the ablated input set. No input subset was selected as a new primary model.")
    (args.output_dir / 'robustness_insert.txt').write_text('\n\n'.join(lines) + '\n', encoding='utf-8')
    print(json.dumps({'status': record['status'], 'unique_oof_rows': len(oof),
        'frozen_OOF_reproduction': record['frozen_OOF_reproduction'],
        'future_mutation': record['future_mutation'], 'comparisons': compact}, indent=2), flush=True)


if __name__ == '__main__':
    main()
```
