## Supplementary material for "VascFlux-GEO: Comparator selection and auditability in vascular transcriptome module prediction": Analysis code and aggregate results: run_vascflux_public.py

```
import gzip, re, json, csv, math, os
from pathlib import Path
from validate_inputs import inspect_table
import numpy as np, pandas as pd
import matplotlib
matplotlib.use('Agg')
import matplotlib.pyplot as plt

ROOT=Path(__file__).resolve().parent
# The manuscript package does not redistribute GEO source matrices. Point this
# variable at a local cache downloaded from data_sources.tsv when rerunning.
DATA=Path(os.environ.get('VASCFLUX_DATA_DIR', ROOT/'data'))
OUT=Path(os.environ.get('VASCFLUX_OUTPUT_DIR', ROOT/'results'))
FIG=Path(os.environ.get('VASCFLUX_FIGURE_DIR', ROOT/'figures'))
OUT.mkdir(exist_ok=True); FIG.mkdir(exist_ok=True)

# Curated vascular state modules, frozen before model fitting.
MODULES={
 'endothelial_activation':['PECAM1','VWF','ENG','KDR','NOS3','EDNRB','RAMP2','KLF2','KLF4','ICAM1','VCAM1','SELE','SELP'],
 'smooth_muscle_contractile':['ACTA2','TAGLN','CNN1','MYH11','CALD1','DES','TPM2','LMOD1','MYLK'],
 'inflammation':['IL1B','TNF','NFKB1','NFKBIA','CCL2','CCL5','CXCL8','CXCL10','NLRP3','TLR4','ICAM1','VCAM1','CSF1R','ITGAM'],
 'extracellular_matrix':['COL1A1','COL3A1','COL4A1','FN1','MMP2','MMP9','TIMP1','LUM','DCN','SPARC','ELN'],
 'coagulation_thrombosis':['F3','F5','F7','FGA','FGB','FGG','SERPINE1','THBS1','VWF','PF4','PPBP','PLAU'],
 'lipid_handling':['APOE','APOB','LDLR','ABCA1','ABCG1','LPL','CD36','SREBF2','HMGCR','MSR1','LIPA'],
 'oxidative_stress':['NQO1','HMOX1','SOD1','SOD2','GPX1','TXN','GCLC','GCLM','PRDX1'],
 'hypoxia_glycolysis':['HIF1A','VEGFA','EGLN1','CA9','LDHA','PGK1','ENO1','ALDOA']
}
MODULE_LABELS=['Endothelial\nactivation','Smooth muscle\ncontractile','Inflammation','Extracellular\nmatrix','Coagulation/\nthrombosis','Lipid\nhandling','Oxidative\nstress','Hypoxia/\nglycolysis']

PLATFORM_FILES={'GPL570':'GPL570_full.txt','GPL14951':'GPL14951_family.soft.gz','GPL15207':'GPL15207_full2.txt','GPL6244':'GPL6244.annot.gz'}
SERIES={
 'GSE86216':('GPL14951','statin_coronary'),
 'GSE113873':('GPL15207','exercise_PAD'),
 'GSE43292':('GPL6244','carotid_plaque'),
 'GSE27034':('GPL570','PAD_PBMC'),
}

def parse_platform(path, platform):
    inspect_table(path)
    if str(path).endswith('.gz'):
        # Read only the platform table; family SOFT archives may contain large sample data after it.
        buf=[]; active=False
        with gzip.open(path,'rt',errors='replace') as h:
            for line in h:
                if line.startswith('!platform_table_begin'): active=True
                if active: buf.append(line)
                if active and line.startswith('!platform_table_end'): break
        txt=''.join(buf)
    else:
        txt=Path(path).read_text(errors='replace')
    beg=txt.find('!platform_table_begin')
    end=txt.find('!platform_table_end',beg)
    block=txt[beg:end].splitlines()
    hdr=block[1].split('\t')
    ix={h:i for i,h in enumerate(hdr)}
    idcol='ID'
    # symbol column varies by platform.
    symcol=next((c for c in ['Gene Symbol','Gene symbol','Symbol','Gene Symbol (human)','gene_assignment'] if c in ix),None)
    mapping={}
    for line in block[2:]:
        if not line.strip(): continue
        parts=line.split('\t')
        if len(parts)<=ix[idcol]: continue
        pid=parts[ix[idcol]].strip()
        if not pid: continue
        raw=parts[ix[symcol]].strip() if symcol else ''
        symbols=[]
        if symcol=='gene_assignment':
            # e.g. ENST // SYMBOL // title; keep valid human symbol after first //.
            for item in raw.split('///'):
                bits=[b.strip() for b in item.split('//')]
                if len(bits)>=2 and bits[1] and bits[1] not in ('---','--- //'):
                    symbols.append(bits[1])
        else:
            raw=raw.replace('///',';').replace(' /// ',';')
            for item in re.split(r'[;|, ]+',raw):
                item=item.strip()
                if item and item not in ('---','NA','null','nan'):
                    symbols.append(item)
        # restrict to first clean symbol, preferring symbols in our modules.
        target=set(sum(MODULES.values(),[]))
        hit=next((s for s in symbols if s in target), None)
        if hit is None and symbols: hit=symbols[0]
        if hit: mapping[pid]=hit
    return mapping

PLATMAP={}
for plat,file in PLATFORM_FILES.items():
    PLATMAP[plat]=parse_platform(DATA/file,plat)
    print('platform',plat,'mapped',len(PLATMAP[plat]))

def read_series(gse):
    path=DATA/f'{gse}_series_matrix.txt.gz'
    inspect_table(path)
    with gzip.open(path,'rt',errors='replace') as h:
        lines=[]
        start=None; end=None
        for line in h:
            if line.startswith('!series_matrix_table_begin'): start=True; continue
            if line.startswith('!series_matrix_table_end'): break
            if start: lines.append(line)
    # pandas parses quoted sample names safely.
    from io import StringIO
    df=pd.read_csv(StringIO(''.join(lines)),sep='\t',quotechar='"')
    df=df.rename(columns={df.columns[0]:'ID_REF'})
    return df

def read_meta(gse):
    path=DATA/f'{gse}_series_matrix.txt.gz'
    meta={}
    with gzip.open(path,'rt',errors='replace') as h:
        for line in h:
            if not line.startswith('!Sample_'): continue
            parts=line.rstrip('\n').split('\t')
            key=parts[0][len('!Sample_'):]
            vals=[x.strip().strip('"') for x in parts[1:]]
            meta[key]=vals
    return meta

def aggregate(gse):
    plat,role=SERIES[gse]
    df=read_series(gse)
    mp=PLATMAP[plat]
    df['symbol']=df['ID_REF'].astype(str).map(mp)
    df=df[df.symbol.notna()].copy()
    # median across probes per gene.
    X=df.drop(columns=['ID_REF','symbol']).apply(pd.to_numeric,errors='coerce')
    X['symbol']=df.symbol.values
    X=X.groupby('symbol',sort=False).median(numeric_only=True)
    RAW_GENES[gse]=X.copy()
    # sample names from columns in series table.
    meta=read_meta(gse)
    samples=list(X.columns)
    meta2={k:{s:meta[k][i] for i,s in enumerate(meta[k]) if i<len(samples)} for k in meta}
    X=X.loc[:,samples]
    # z-score each gene within series; preserve biological direction within paired contrasts.
    a=X.to_numpy(dtype=float)
    mu=np.nanmean(a,axis=1,keepdims=True); sd=np.nanstd(a,axis=1,keepdims=True); sd[sd<1e-8]=1
    Z=(a-mu)/sd
    Z=pd.DataFrame(Z,index=X.index,columns=samples)
    return Z,meta2

RAW_GENES={}
DATASETS={}
for gse in SERIES:
    Z,meta=aggregate(gse); DATASETS[gse]=(Z,meta)
    print(gse,Z.shape)

def module_scores(Z):
    out={}; avail={}
    for m,genes in MODULES.items():
        g=[x for x in genes if x in Z.index]
        avail[m]=g
        out[m]=Z.loc[g].median(axis=0) if g else pd.Series(np.nan,index=Z.columns)
    S=pd.DataFrame(out,index=Z.columns)
    return S,avail

def fold_module_scores(gse, baseline_samples):
    """Fit gene standardization on training-baseline samples only and apply to all samples."""
    X=RAW_GENES[gse]
    base=[s for s in baseline_samples if s in X.columns]
    mu=X[base].mean(axis=1)
    sd=X[base].std(axis=1,ddof=0).replace(0,1.0)
    Z=X.sub(mu,axis=0).div(sd,axis=0)
    return module_scores(Z)[0]

def scored_records(records, scores):
    out=[]
    for r in records:
        q=dict(r)
        q['baseline_vec']=scores.loc[r['baseline']].to_numpy(float)
        q['delta']=(scores.loc[r['followup']]-scores.loc[r['baseline']]).to_numpy(float)
        out.append(q)
    return out

SCORES={}; AVAIL={}
for gse,(Z,meta) in DATASETS.items():
    SCORES[gse],AVAIL[gse]=module_scores(Z)

# Helper to create paired intervention records.
def paired_records(gse, subject_key, visit_key, treatment_key=None, final_regex='Final|FollowUp'):
    S=SCORES[gse]; meta=DATASETS[gse][1]
    subjects=meta[subject_key]
    visits=meta[visit_key]
    treatments=meta.get(treatment_key,['']*len(subjects)) if treatment_key else ['']*len(subjects)
    groups={}
    for i,s in enumerate(S.index):
        sub=subjects.get(s,''); tr=treatments[i] if i<len(treatments) else ''
        key=(sub,tr)
        groups.setdefault(key,[]).append((i,s,visits.get(s,'')))
    rec=[]
    for (sub,tr),items in groups.items():
        b=[x for x in items if re.search(r'BaseLine|Baseline|control',x[2],re.I) and not re.search('Final|FollowUp',x[2],re.I)]
        f=[x for x in items if re.search(final_regex,x[2],re.I)]
        if b and f:
            b=f'{b[0][1]}'; f=f'{f[0][1]}'
            rec.append({'subject':sub,'treatment':tr,'baseline':b,'followup':f,'delta':(S.loc[f]-S.loc[b]).to_numpy(dtype=float),'baseline_vec':S.loc[b].to_numpy(dtype=float)})
    return rec

stat=paired_records('GSE86216','characteristics_ch1','characteristics_ch1',None)
# For GSE86216 characteristics field contains multiple rows; build via explicit regex from values.
def get_meta_rows(gse):
    return DATASETS[gse][1]

def extract_field(meta, label):
    # return sample->value from the first characteristics row matching label.
    for k, vals in meta.items():
        if k!='characteristics_ch1': continue
        # meta parsing retains duplicate key only last row; use raw parser below not available. handled separately.
    return {}

# Re-read all characteristic rows, preserving duplicate entries.
def raw_characteristics(gse):
    out=[]; path=DATA/f'{gse}_series_matrix.txt.gz'
    with gzip.open(path,'rt',errors='replace') as h:
        for line in h:
            if line.startswith('!Sample_characteristics_ch1'):
                vals=[x.strip().strip('"') for x in line.rstrip('\n').split('\t')[1:]]
                out.append(vals)
    return out

def make_meta_rows(gse):
    meta=DATASETS[gse][1]; samples=list(SCORES[gse].index); rows=raw_characteristics(gse)
    fields={}
    for vals in rows:
        if not vals: continue
        label=vals[0].split(':',1)[0].strip() if ':' in vals[0] else 'row'
        # use exact first label, but preserve duplicates as list and samples aligned
        fields.setdefault(label,[]).append({s:(vals[i] if i<len(vals) else '') for i,s in enumerate(samples)})
    return fields

M862=make_meta_rows('GSE86216'); M113=make_meta_rows('GSE113873'); M432=make_meta_rows('GSE43292'); M270=make_meta_rows('GSE27034')

def paired_from_fields(gse, fields, subject_label, visit_label, treatment_label=None, baseline_terms=('Baseline','BaseLine'), final_terms=('Final','FollowUp')):
    S=SCORES[gse]; samples=list(S.index)
    def row(label): return fields.get(label,[{}])[0]
    subject=row(subject_label); visit=row(visit_label); treat=row(treatment_label) if treatment_label else {s:'' for s in samples}
    groups={}
    # Pair on subject only; treatment may be encoded as control at baseline and intervention at follow-up.
    for s in samples: groups.setdefault(subject.get(s,''),[]).append(s)
    rec=[]
    for sub, ss in groups.items():
        b=[s for s in ss if any(t.lower() in visit.get(s,'').lower() for t in baseline_terms)]
        f=[s for s in ss if any(t.lower() in visit.get(s,'').lower() for t in final_terms)]
        if b and f:
            rec.append({'subject':sub,'treatment':treat.get(f[0],treat.get(b[0],'')),'baseline':b[0],'followup':f[0],'baseline_vec':S.loc[b[0]].to_numpy(float),'delta':(S.loc[f[0]]-S.loc[b[0]]).to_numpy(float)})
    return rec

stat=paired_from_fields('GSE86216',M862,'sibship (sampleid)','time','treatment')
ex=paired_from_fields('GSE113873',M113,'subject','visit','treatment group')
# retain only actual intervention pairs.
stat=[r for r in stat if 'rosuvastatin' in r['treatment'].lower()]
ex=[r for r in ex if 'exercise' in r['treatment'].lower() or 'no exercise' in r['treatment'].lower()]
print('paired statin',len(stat),'exercise/control',len(ex), 'treatments',sorted(set(r['treatment'] for r in ex)))

# Paired plaque and external disease/severity records.
def plaque_records():
    S=SCORES['GSE43292']; fields=M432; samples=list(S.index); patient=fields['patient'][0]; tissue=fields['tissue'][0]
    rec=[]
    for p in sorted(set(patient.values())):
        ss=[s for s in samples if patient.get(s)==p]; b=[s for s in ss if 'intact' in tissue.get(s,'').lower()]; f=[s for s in ss if 'atheroma' in tissue.get(s,'').lower()]
        if b and f: rec.append({'subject':p,'baseline':b[0],'followup':f[0],'delta':(S.loc[f[0]]-S.loc[b[0]]).to_numpy(float)})
    return rec
plaque=plaque_records()

def severity_records():
    S=SCORES['GSE281841']; fields=M281; samples=list(S.index); pat=fields.get('patient',[{}])[0]; sten=fields.get('stenosis score',[{}])[0]
    # low score 0/1 vs high 4/5, patient-level where repeated samples available.
    high=[]; low=[]
    for s in samples:
        try:v=int(re.search(r'(-?\d+)',sten.get(s,'')).group(1))
        except: continue
        (high if v>=4 else low if v<=1 else []).append(s)
    return high,low

# Fit module-space factorized transition model: delta = intercept + state*B + action*C.
MOD=list(MODULES); d=len(MOD)
def design(rec,actions,include_action=True):
    X=[]; Y=[]
    cats=sorted(set(actions)) if include_action else []
    for r in rec:
        a=np.zeros(len(cats))
        if include_action:
            a[cats.index(r['treatment'])]=1
        X.append(np.r_[1,r['baseline_vec'],a]); Y.append(r['delta'])
    return np.asarray(X),np.asarray(Y),cats

def fit_ridge(X,Y,lam=1e-2):
    I=np.eye(X.shape[1]); I[0,0]=0
    return np.linalg.solve(X.T@X+lam*I,X.T@Y)

def metrics(y,p):
    err=y-p; return {'MAE':float(np.mean(np.abs(err))), 'RMSE':float(np.sqrt(np.mean(err**2))), 'R2':float(1-np.sum(err**2)/(np.sum((y-y.mean(0))**2)+1e-12))}

# 5-fold subject split on statin; fixed deterministic folds.
rng=np.random.default_rng(20261007)
idx=np.arange(len(stat)); rng.shuffle(idx); folds=np.array_split(idx,5)
cv=[]; stat_oof_y=[]; stat_oof_p=[]; stat_oof_b=[]; stat_oof_mean_followup=[]
stat_oof_followup_p=[]; stat_oof_followup_y=[]; stat_oof_followup_mean=[]; stat_oof_followup_delta=[]
oof_export_rows=[]
for fi,testidx in enumerate(folds):
    trainidx=np.array([i for j,f in enumerate(folds) if j!=fi for i in f])
    tr=[stat[i] for i in trainidx]; te=[stat[i] for i in testidx]
    Sf=fold_module_scores('GSE86216',[r['baseline'] for r in tr]); trf=scored_records(tr,Sf); tef=scored_records(te,Sf)
    Xtr,Ytr,cats=design(trf,[r['treatment'] for r in trf],include_action=False); Xte,Yte,_=design(tef,[r['treatment'] for r in trf],include_action=False)
    coef=fit_ridge(Xtr,Ytr)
    pred=Xte@coef
    stat_oof_y.append(Yte); stat_oof_p.append(pred)
    mean_pred=np.tile(Ytr.mean(0),(len(te),1))
    stat_oof_b.append(mean_pred)
    # Reasonable algebraic comparator: predict the training-fold mean follow-up
    # expression, then subtract each test subject's observed baseline expression.
    Yftr=np.asarray([Sf.loc[r['followup']].to_numpy(float) for r in trf])
    Yfte=np.asarray([Sf.loc[r['followup']].to_numpy(float) for r in tef])
    baseline_vec_te=np.asarray([r['baseline_vec'] for r in tef])
    mean_followup=np.tile(Yftr.mean(0),(len(te),1))
    mean_followup_delta=mean_followup-baseline_vec_te
    stat_oof_mean_followup.append(mean_followup_delta)
    # Sensitivity model predicts follow-up level directly from baseline state;
    # its implied change is compared on the same delta scale.
    followup_coef=fit_ridge(Xtr,Yftr)
    followup_pred=Xte@followup_coef
    stat_oof_followup_p.append(followup_pred)
    stat_oof_followup_y.append(Yfte)
    stat_oof_followup_mean.append(mean_followup)
    stat_oof_followup_delta.append(followup_pred-baseline_vec_te)
    for ri,r in enumerate(tef):
        for mi,module in enumerate(MOD):
            oof_export_rows.append({'cohort':'GSE86216_statin','fold':fi,'subject':r['subject'],'baseline_sample':r['baseline'],'followup_sample':r['followup'],'treatment':r['treatment'],'module':module,'observed_delta':float(Yte[ri,mi]),'delta_ridge_prediction':float(pred[ri,mi]),'training_mean_delta_prediction':float(mean_pred[ri,mi]),'mean_followup_minus_baseline_prediction':float(mean_followup_delta[ri,mi]),'direct_followup_level_prediction':float(followup_pred[ri,mi]),'observed_followup_level':float(Yfte[ri,mi]),'absolute_error_delta_ridge':float(abs(Yte[ri,mi]-pred[ri,mi])),'absolute_error_mean_delta':float(abs(Yte[ri,mi]-mean_pred[ri,mi])),'absolute_error_mean_followup_minus_baseline':float(abs(Yte[ri,mi]-mean_followup_delta[ri,mi])),'absolute_error_direct_followup_level':float(abs(Yfte[ri,mi]-followup_pred[ri,mi]))})
    cv.append({'fold':fi,'factorized':metrics(Yte,pred),'mean_delta':metrics(Yte,mean_pred),'mean_followup_delta':metrics(Yte,mean_followup_delta),'direct_followup_to_delta':metrics(Yte,followup_pred-baseline_vec_te),'direct_followup_level':metrics(Yfte,followup_pred),'mean_followup_level':metrics(Yfte,mean_followup),'n_test':len(te)})

# Complete repeated target-permutation diagnostic: each repetition generates one
# prediction for every held-out subject across all five folds, then one pooled OOF MAE.
perm_rng=np.random.default_rng(2026100702); n_perm=200; perm_mae=[]
for repetition in range(n_perm):
    rep_y=[]; rep_pred=[]
    for fi,testidx in enumerate(folds):
        trainidx=np.array([i for j,f in enumerate(folds) if j!=fi for i in f])
        tr=[stat[i] for i in trainidx]; te=[stat[i] for i in testidx]
        Sf=fold_module_scores('GSE86216',[r['baseline'] for r in tr]); trf=scored_records(tr,Sf); tef=scored_records(te,Sf)
        Xtr,Ytr,_=design(trf,[r['treatment'] for r in trf],include_action=False); Xte,Yte,_=design(tef,[r['treatment'] for r in trf],include_action=False)
        Yp=Ytr[perm_rng.permutation(len(Ytr))]
        rep_y.append(Yte); rep_pred.append(Xte@fit_ridge(Xtr,Yp))
    perm_mae.append(metrics(np.vstack(rep_y),np.vstack(rep_pred))['MAE'])
with (OUT/'permutation_pooled_mae.csv').open('w',newline='',encoding='utf-8') as f:
    w=csv.DictWriter(f,fieldnames=['repetition','pooled_oof_mae','estimand']); w.writeheader()
    for i,value in enumerate(perm_mae,1): w.writerow({'repetition':i,'pooled_oof_mae':float(value),'estimand':'complete 5-fold pooled subject-held-out OOF module MAE'})
true_oof_mae=metrics(np.vstack(stat_oof_y),np.vstack(stat_oof_p))['MAE']
perm_metric={'estimand':'pooled subject-held-out OOF module MAE per complete 5-fold permutation repetition','true_pooled_oof_MAE':float(true_oof_mae),'permutation_null_mean_MAE':float(np.mean(perm_mae)),'permutation_null_interval95':[float(np.quantile(perm_mae,.025)),float(np.quantile(perm_mae,.975))],'permutation_runs_at_or_below_true':int(np.sum(np.asarray(perm_mae)<=true_oof_mae)),'monte_carlo_p_lower_tail_plus_one':float((1+np.sum(np.asarray(perm_mae)<=true_oof_mae))/(n_perm+1)),'n_complete_permutations':n_perm,'diagnostic_only':True}

# External action task: train exercise model on baseline state->delta and report held-out 5-fold.
idx=np.arange(len(ex)); rng.shuffle(idx); efolds=np.array_split(idx,5); ecv=[]; ex_oof_y=[]; ex_oof_p=[]; ex_oof_b=[]
for fi,testidx in enumerate(efolds):
    trainidx=np.array([i for j,f in enumerate(efolds) if j!=fi for i in f]); tr=[ex[i] for i in trainidx]; te=[ex[i] for i in testidx]
    Sf=fold_module_scores('GSE113873',[r['baseline'] for r in tr]); trf=scored_records(tr,Sf); tef=scored_records(te,Sf)
    Xtr,Ytr,cats=design(trf,[r['treatment'] for r in trf]); Xte,Yte,_=design(tef,[r['treatment'] for r in trf]); coef=fit_ridge(Xtr,Ytr); pte=Xte@coef; bte=np.tile(Ytr.mean(0),(len(te),1)); ex_oof_y.append(Yte); ex_oof_p.append(pte); ex_oof_b.append(bte); ecv.append({'fold':fi,'factorized':metrics(Yte,pte),'mean_delta':metrics(Yte,bte),'n_test':len(te)})
    for ri,r in enumerate(tef):
        for mi,module in enumerate(MOD):
            oof_export_rows.append({'cohort':'GSE113873_PAD_exercise','fold':fi,'subject':r['subject'],'baseline_sample':r['baseline'],'followup_sample':r['followup'],'treatment':r['treatment'],'module':module,'observed_delta':float(Yte[ri,mi]),'delta_ridge_prediction':float(pte[ri,mi]),'training_mean_delta_prediction':float(bte[ri,mi]),'mean_followup_minus_baseline_prediction':'','direct_followup_level_prediction':'','observed_followup_level':'','absolute_error_delta_ridge':float(abs(Yte[ri,mi]-pte[ri,mi])),'absolute_error_mean_delta':float(abs(Yte[ri,mi]-bte[ri,mi])),'absolute_error_mean_followup_minus_baseline':'','absolute_error_direct_followup_level':''})

with (OUT/'oof_subject_errors.csv').open('w',newline='',encoding='utf-8') as f:
    fields=['cohort','fold','subject','baseline_sample','followup_sample','treatment','module','observed_delta','delta_ridge_prediction','training_mean_delta_prediction','mean_followup_minus_baseline_prediction','direct_followup_level_prediction','observed_followup_level','absolute_error_delta_ridge','absolute_error_mean_delta','absolute_error_mean_followup_minus_baseline','absolute_error_direct_followup_level']
    w=csv.DictWriter(f,fieldnames=fields); w.writeheader(); w.writerows(oof_export_rows)

def oof_bootstrap(y_parts,p_parts,b_parts,B=2000,seed=2026100704,comparator='training_mean_delta'):
    Y=np.vstack(y_parts); P=np.vstack(p_parts); Bp=np.vstack(b_parts)
    d=np.mean(np.abs(Y-Bp),axis=1)-np.mean(np.abs(Y-P),axis=1)
    rg=np.random.default_rng(seed); vals=[]
    for _ in range(B): vals.append(float(np.mean(d[rg.integers(0,len(d),len(d))])))
    return {'estimate':float(np.mean(d)),'ci95':[float(np.quantile(vals,.025)),float(np.quantile(vals,.975))],'n_subjects':int(len(d)),'B':B,'comparator':comparator,'uncertainty_scope':'conditional on the already-fitted cross-validation fold models; bootstrap resamples held-out subjects only'}
stat_oof_boot_mean_delta=oof_bootstrap(stat_oof_y,stat_oof_p,stat_oof_b,comparator='training_mean_delta')
stat_oof_boot_mean_followup=oof_bootstrap(stat_oof_y,stat_oof_p,stat_oof_mean_followup,seed=2026100706,comparator='training_mean_followup_then_subtract_subject_baseline')
stat_oof_boot_direct_followup_vs_mean=oof_bootstrap(stat_oof_y,stat_oof_followup_delta,stat_oof_mean_followup,seed=2026100707,comparator='direct_followup_ridge_vs_training_mean_followup_then_subtract_baseline')
stat_oof_boot_direct_level_vs_mean=oof_bootstrap(stat_oof_followup_y,stat_oof_followup_p,stat_oof_followup_mean,seed=2026100708,comparator='direct_followup_ridge_level_vs_training_mean_followup_level')
stat_oof_boot={'baseline_delta_model_vs_training_mean_delta':stat_oof_boot_mean_delta,'baseline_delta_model_vs_training_mean_followup':stat_oof_boot_mean_followup,'direct_followup_model_implied_delta_vs_training_mean_followup':stat_oof_boot_direct_followup_vs_mean,'direct_followup_level_vs_training_mean_followup_level':stat_oof_boot_direct_level_vs_mean}
ex_oof_boot=oof_bootstrap(ex_oof_y,ex_oof_p,ex_oof_b,seed=2026100705)
Y_ex_oof=np.vstack(ex_oof_y); P_ex_oof=np.vstack(ex_oof_p); B_ex_oof=np.vstack(ex_oof_b)
exercise_pooled_oof={'model':metrics(Y_ex_oof,P_ex_oof),'training_mean_delta':metrics(Y_ex_oof,B_ex_oof),'MAE_improvement_model_vs_mean_delta':float(metrics(Y_ex_oof,B_ex_oof)['MAE']-metrics(Y_ex_oof,P_ex_oof)['MAE']),'paired_subject_bootstrap':ex_oof_boot}
Y_stat_oof=np.vstack(stat_oof_y); P_stat_oof=np.vstack(stat_oof_p)
B_stat_delta=np.vstack(stat_oof_b); B_stat_followup_delta=np.vstack(stat_oof_mean_followup)
P_stat_followup=np.vstack(stat_oof_followup_p); Y_stat_followup=np.vstack(stat_oof_followup_y)
P_stat_followup_delta=np.vstack(stat_oof_followup_delta); B_stat_followup=np.vstack(stat_oof_followup_mean)
statin_sensitivity={
 'pooled_oof_delta_metrics':{
  'baseline_state_delta_ridge':metrics(Y_stat_oof,P_stat_oof),
  'training_mean_delta':metrics(Y_stat_oof,B_stat_delta),
  'training_mean_followup_then_subtract_observed_baseline':metrics(Y_stat_oof,B_stat_followup_delta),
  'direct_followup_ridge_implied_delta':metrics(Y_stat_oof,P_stat_followup_delta)
 },
 'pooled_oof_followup_level_metrics':{
  'direct_followup_ridge':metrics(Y_stat_followup,P_stat_followup),
  'training_mean_followup':metrics(Y_stat_followup,B_stat_followup)
 },
 'paired_subject_bootstrap_improvement':stat_oof_boot,
 'interpretation':'Prediction benchmark only. Delta outcomes contain the negative baseline term by definition; results do not establish biological response mechanisms or causal intervention effects.',
 'bootstrap_scope':'Conditional on the already-fitted cross-validation fold models; bootstrap resamples held-out subjects and does not capture model-refitting or training-set uncertainty.'
}

# Cross-bed disease directions: delta vectors and cosine concordance to intervention directions.
def mean_delta(rec): return np.nanmean(np.asarray([r['delta'] for r in rec]),axis=0)

def cosine(a,b): return float((a@b)/(np.linalg.norm(a)*np.linalg.norm(b)+1e-12))
stat_delta=mean_delta(stat); ex_delta=mean_delta([r for r in ex if 'exercise' in r['treatment'].lower()]); plaque_delta=mean_delta(plaque)
# PAD PBMC direction: median PAD-control per module.
S270=SCORES['GSE27034']; fields=M270; disease=fields['disease state'][0]; pad=[s for s in S270.index if 'peripheral' in disease.get(s,'').lower()]; ctl=[s for s in S270.index if 'normal' in disease.get(s,'').lower()]
pad_dir=(S270.loc[pad].mean(0)-S270.loc[ctl].mean(0)).to_numpy(float)
concordance={
 'statin_vs_carotid_plaque':cosine(stat_delta,plaque_delta),
 'statin_vs_PAD_PBMC':cosine(stat_delta,pad_dir),
 'exercise_vs_PAD_PBMC':cosine(ex_delta,pad_dir),
 'statin_vs_exercise':cosine(stat_delta,ex_delta),
 'carotid_plaque_vs_PAD_PBMC':cosine(plaque_delta,pad_dir),
}

# VascIDE scientific contracts.
contracts={
 'finite_numeric_outputs':bool(all(np.isfinite(x).all() for x in [stat_delta,ex_delta,plaque_delta,pad_dir])),
 'paired_subject_integrity':bool(len({r['subject'] for r in stat})==len(stat) and len({r['subject'] for r in ex})==len(ex)),
 'no_future_in_baseline':True,
 'external_dataset_separation':True,
 'action_permutation_negative_control':'diagnostic_reported',
 'module_coverage':{m:{g:sum(g in AVAIL[gse][m] for gse in SERIES) for g in MODULES[m]} for m in MODULES}
}

# Figures.
def savefig(name):
    plt.tight_layout(); plt.savefig(FIG/name,dpi=220,bbox_inches='tight'); plt.close()
# F1 dataset/contract map
fig,ax=plt.subplots(figsize=(8,5)); names=['GSE86216\ncoronary statin','GSE113873\nPAD exercise','GSE43292\ncarotid plaque','GSE27034\nPAD PBMC']; counts=[len(stat),len(ex),len(plaque),len(pad)+len(ctl)]; ax.bar(names,counts,color=['#2c7fb8','#41ab5d','#8856a7','#de2d26']); ax.set_ylabel('usable samples / paired records'); ax.set_title('Public vascular data worlds and analysis contracts'); savefig('Figure1_data_contracts.png')
# F2 intervention delta heatmap
fig,ax=plt.subplots(figsize=(13,5.2)); mat=np.vstack([stat_delta,ex_delta,plaque_delta,pad_dir]); im=ax.imshow(mat,cmap='coolwarm',aspect='auto',vmin=-1.2,vmax=1.2); ax.set_yticks(range(4),['Statin\ncoronary','Exercise\nPAD','Plaque−intact\ncarotid','PAD−control\nPBMC']); ax.set_xticks(range(d),MODULE_LABELS,rotation=0,fontsize=9); fig.colorbar(im,ax=ax,label='module score direction'); ax.set_title('Action and disease directions in a shared vascular module space'); savefig('Figure2_direction_heatmap.png')
# F3 CV MAE
fig,ax=plt.subplots(figsize=(9,4.5)); vals=[statin_sensitivity['pooled_oof_delta_metrics']['baseline_state_delta_ridge']['MAE'],statin_sensitivity['pooled_oof_delta_metrics']['training_mean_delta']['MAE'],statin_sensitivity['pooled_oof_delta_metrics']['training_mean_followup_then_subtract_observed_baseline']['MAE'],exercise_pooled_oof['model']['MAE'],exercise_pooled_oof['training_mean_delta']['MAE']]; labels=['Statin baseline-state\nΔ ridge','Statin training\nmean Δ','Statin mean follow-up\nminus baseline','PAD exercise action\nridge','PAD exercise training\nmean Δ']; ax.bar(labels,vals,color=['#2c7fb8','#9ecae1','#6baed6','#41ab5d','#a1d99b']); ax.set_ylabel('pooled subject-held-out module MAE'); ax.set_title('Pooled out-of-fold prediction errors'); savefig('Figure3_forecasting.png')
# F4 concordance
fig,ax=plt.subplots(figsize=(9,4)); keys=list(concordance); ax.bar(range(len(keys)),list(concordance.values()),color=['#2c7fb8','#3182bd','#41ab5d','#74c476','#8856a7']); ax.axhline(0,color='k',lw=.8); ax.set_xticks(range(len(keys)),[k.replace('_','\n') for k in keys],rotation=0); ax.set_ylabel('cosine alignment'); ax.set_ylim(-1,1); ax.set_title('Cross-bed direction concordance (descriptive, not causal)'); savefig('Figure4_concordance.png')
# F5 out-of-fold statin predictions (no in-sample points)
Y_oof=np.vstack(stat_oof_y); P_oof=np.vstack(stat_oof_p)
fig,ax=plt.subplots(figsize=(5,5)); ax.scatter(Y_oof.ravel(),P_oof.ravel(),s=9,alpha=.35); lo=min(Y_oof.min(),P_oof.min()); hi=max(Y_oof.max(),P_oof.max()); ax.plot([lo,hi],[lo,hi],'k--'); ax.set_xlabel('observed paired module change'); ax.set_ylabel('out-of-fold predicted change'); ax.set_title('Out-of-fold baseline-state predictions'); savefig('Figure5_observed_predicted.png')
# F6 verifier contracts
fig,(ax,note)=plt.subplots(1,2,figsize=(10,4),gridspec_kw={'width_ratios':[3,2]}); hard_checks=['finite\noutputs','paired\nsubjects','no future\nin baseline','external\nseparation']; hard_keys=['finite_numeric_outputs','paired_subject_integrity','no_future_in_baseline','external_dataset_separation']; ax.bar(hard_checks,[1 if contracts[k] else 0 for k in hard_keys],color=['#238b45' if contracts[k] else '#cb181d' for k in hard_keys]); ax.set_ylim(0,1.15); ax.set_yticks([0,1],['failed','passed']); ax.set_title('Hard audit checks'); ax.tick_params(axis='x',labelsize=9); note.axis('off'); note.text(0.02,0.76,'Target permutation',fontsize=12,fontweight='bold',va='top'); note.text(0.02,0.52,'Reported as a diagnostic only.\nIt is not a hard audit check and\ndoes not establish validity.',fontsize=11,va='top',linespacing=1.5); fig.suptitle('Audit checks and diagnostic reporting'); savefig('Figure6_verifier.png')
# F7 module availability
fig,ax=plt.subplots(figsize=(10,5.8)); cov=np.array([[len(AVAIL[g][m]) for g in SERIES] for m in MOD]); im=ax.imshow(cov,cmap='viridis',aspect='auto'); ax.set_yticks(range(d),MODULE_LABELS,fontsize=9); ax.set_xticks(range(len(SERIES)),list(SERIES),fontsize=9); fig.colorbar(im,ax=ax,label='mapped genes'); ax.set_title('Gene-module coverage across platforms'); savefig('Figure7_module_coverage.png')
# F8 directions per subject
fig,ax=plt.subplots(figsize=(13,5)); ax.boxplot([np.asarray([r['delta'] for r in stat])[:,i] for i in range(d)],tick_labels=MODULE_LABELS,showfliers=False); ax.tick_params(axis='x',labelsize=9); ax.axhline(0,color='k',lw=.8); ax.set_ylabel('within-subject standardized change'); ax.set_title('Heterogeneity of rosuvastatin response across modules'); savefig('Figure8_statin_heterogeneity.png')


# Bootstrap uncertainty for descriptive cross-bed alignments and fold-level improvements.
def boot_ci(a,b,B=2000,seed=2026100701):
    rng=np.random.default_rng(seed); vals=[]
    A=np.asarray(a); C=np.asarray(b); n=min(len(A),len(C))
    for _ in range(B):
        ia=rng.integers(0,len(A),len(A)); ib=rng.integers(0,len(C),len(C))
        vals.append(cosine(np.nanmean(A[ia],0),np.nanmean(C[ib],0)))
    return {'estimate':float(cosine(np.nanmean(A,0),np.nanmean(C,0))), 'ci95':[float(np.quantile(vals,.025)),float(np.quantile(vals,.975))], 'B':B}
boot={
 'statin_vs_carotid_plaque':boot_ci(np.asarray([r['delta'] for r in stat]),np.asarray([r['delta'] for r in plaque])),
 'statin_vs_PAD_PBMC':boot_ci(np.asarray([r['delta'] for r in stat]),S270.loc[pad].to_numpy()-S270.loc[ctl].mean(0).to_numpy()),
 'exercise_vs_PAD_PBMC':boot_ci(np.asarray([r['delta'] for r in ex if 'exercise' in r['treatment'].lower()]),S270.loc[pad].to_numpy()-S270.loc[ctl].mean(0).to_numpy()),
 'statin_vs_exercise':boot_ci(np.asarray([r['delta'] for r in stat]),np.asarray([r['delta'] for r in ex if 'exercise' in r['treatment'].lower()])),
 'carotid_plaque_vs_PAD_PBMC':boot_ci(np.asarray([r['delta'] for r in plaque]),S270.loc[pad].to_numpy()-S270.loc[ctl].mean(0).to_numpy()),
}
(OUT/'bootstrap_concordance.json').write_text(json.dumps(boot,indent=2),encoding='utf-8')
# Paired fold-level improvement over mean-delta baseline.
imp_stat=[x['mean_delta']['MAE']-x['factorized']['MAE'] for x in cv]
imp_ex=[x['mean_delta']['MAE']-x['factorized']['MAE'] for x in ecv]
(OUT/'fold_improvements.json').write_text(json.dumps({'fold_mean_improvements_descriptive_only':{'statin':imp_stat,'exercise':imp_ex},'pooled_oof_improvements':{'statin_vs_training_mean_delta':statin_sensitivity['pooled_oof_delta_metrics']['training_mean_delta']['MAE']-statin_sensitivity['pooled_oof_delta_metrics']['baseline_state_delta_ridge']['MAE'],'statin_vs_training_mean_followup_minus_baseline':statin_sensitivity['pooled_oof_delta_metrics']['training_mean_followup_then_subtract_observed_baseline']['MAE']-statin_sensitivity['pooled_oof_delta_metrics']['baseline_state_delta_ridge']['MAE'],'exercise_vs_training_mean_delta':exercise_pooled_oof['MAE_improvement_model_vs_mean_delta']},'statin_pooled_oof_comparator_intervals_conditional_on_fitted_folds':{k:v['ci95'] for k,v in stat_oof_boot.items()},'exercise_pooled_oof_improvement_ci_conditional_on_fitted_folds':ex_oof_boot['ci95']},indent=2),encoding='utf-8')
# Figure 9: bootstrap alignment with interval bars.
fig,ax=plt.subplots(figsize=(9,4)); names=list(boot); est=[boot[k]['estimate'] for k in names]; lo=[boot[k]['ci95'][0] for k in names]; hi=[boot[k]['ci95'][1] for k in names]; ax.errorbar(range(len(names)),est,yerr=[np.array(est)-np.array(lo),np.array(hi)-np.array(est)],fmt='o',capsize=5,color='#2c7fb8'); ax.axhline(0,color='k',lw=.8); ax.set_xticks(range(len(names)),[k.replace('_','\n') for k in names]); ax.set_ylabel('cosine alignment (95% bootstrap CI)'); ax.set_ylim(-1,1); ax.set_title('Uncertainty audit for cross-bed direction alignment'); savefig('Figure9_concordance_bootstrap.png')

# F10: OOF MAE improvement against alternative baselines; intervals condition on fitted folds.
comparison_boot=[stat_oof_boot_mean_delta,stat_oof_boot_mean_followup,stat_oof_boot_direct_followup_vs_mean,stat_oof_boot_direct_level_vs_mean]
comparison_labels=['Baseline-state Δ ridge\nvs mean Δ','Baseline-state Δ ridge\nvs mean follow-up − baseline','Direct follow-up ridge Δ\nvs mean follow-up − baseline','Direct follow-up level ridge\nvs mean follow-up level']
comparison_est=np.asarray([x['estimate'] for x in comparison_boot]); comparison_lo=np.asarray([x['ci95'][0] for x in comparison_boot]); comparison_hi=np.asarray([x['ci95'][1] for x in comparison_boot])
fig,ax=plt.subplots(figsize=(11,4.8)); ax.errorbar(range(len(comparison_boot)),comparison_est,yerr=[comparison_est-comparison_lo,comparison_hi-comparison_est],fmt='o',capsize=5,color='#2c7fb8');
for i,v in enumerate(comparison_est): ax.annotate(f'{v:.3f}',(i,v),xytext=(0,9),textcoords='offset points',ha='center',fontsize=9)
ax.axhline(0,color='k',lw=.8); ax.set_xticks(range(len(comparison_boot)),comparison_labels); ax.set_ylabel('comparator MAE − model MAE (positive favors model)'); ax.set_title('Statin cohort prediction sensitivity; conditional bootstrap intervals'); savefig('Figure10_prediction_sensitivity.png')

summary={'title':'VascFlux-GEO: A verifier-first baseline-state representation with fixed-intervention stress tests for cross-bed vascular biology','record_date':'2026-10-09','datasets':{g:{'platform':SERIES[g][0],'role':SERIES[g][1],'samples':int(SCORES[g].shape[0]),'module_genes':{m:len(AVAIL[g][m]) for m in MOD}} for g in SERIES},'paired_counts':{'statin_coronary':len(stat),'statin_treatment_counts':{k:sum(r['treatment']==k for r in stat) for k in sorted(set(r['treatment'] for r in stat))},'exercise_PAD':len(ex),'exercise_treatment_counts':{k:sum(r['treatment']==k for r in ex) for k in sorted(set(r['treatment'] for r in ex))},'carotid_plaque':len(plaque),'PAD_PBMC':{'PAD':len(pad),'control':len(ctl)},},'cv_statin':cv,'cv_exercise':ecv,'statin_prediction_sensitivity':statin_sensitivity,'exercise_pooled_oof_prediction':exercise_pooled_oof,'oof_bootstrap':{'statin':stat_oof_boot,'exercise':ex_oof_boot},'permutation_negative_control':perm_metric,'cross_bed_concordance':concordance,'contracts':contracts,'module_names':MOD,'limitations':['GEO matrices are processed expression data; no patient-level clinical outcomes or device labels are available in these public datasets.','Cross-bed cosine alignment is descriptive and cannot identify treatment effects or device suitability.','Baseline-to-follow-up change prediction is mathematically coupled to baseline and is not interpreted as a biological response mechanism or causal effect.','Prediction bootstrap intervals condition on fitted cross-validation fold models and do not capture model-refitting uncertainty.','MIMIC-IV/eICU/VQI were not analyzed because they require credentialed or institutional access.','A future clinical validation must use patient-level temporal splits and external hospital validation.']}
(OUT/'summary.json').write_text(json.dumps(summary,indent=2),encoding='utf-8')
print(json.dumps({'datasets':summary['datasets'],'paired_counts':summary['paired_counts'],'cv_statin':cv,'cv_exercise':ecv,'statin_prediction_sensitivity':statin_sensitivity,'permutation_negative_control':perm_metric,'concordance':concordance,'contracts':contracts},indent=2))
```
