## Supplementary material for "VascFlux-GEO: Comparator selection and auditability in vascular transcriptome module prediction": Analysis code and aggregate results: STATISTICAL_DATA_DICTIONARY.md

# Statistical output interpretation

This dictionary clarifies existing output names. It does not rename columns, change values, rerun analyses or confer inferential validity on historical names.

| Output / field | Meaning and limits |
|---|---|
| `results_extended/conditional_subject_intervals.csv`, `results_reviewer_revision/fair_paired_differences.csv`, and corresponding representation-sensitivity outputs: `conditional_ci95_low`, `conditional_ci95_high` | Lower and upper percentile endpoints from 5,000 subject bootstrap draws of fixed observed OOF losses. Subject errors are averaged across modules and executed repetitions before resampling, with the same subject weights across methods. The endpoints form a descriptive central 95% resampling range. The procedure does not refit preprocessing, tuning or models, and does not repeat post hoc comparator development/selection. Shared training observations induce dependence between subject losses; averaging within subject does not remove it. The nominal level is not a validated conditional or population coverage guarantee. The `ci95` spelling is retained solely for compatibility and traceability. |
| `estimate` in paired temporal comparisons | Observed comparator MAE minus method MAE on the matched fixed predictions. A positive number indicates lower observed loss for the named method; it does not by itself establish population superiority, equivalence or noninferiority. The empirical best simple comparator is a descriptive ranking selected after observing results, not a locked selection procedure. |
| Complete-repeat central ranges | Variation across the 20 executed repeated partitions. The repetitions share subjects and are dependent; these ranges are not confidence intervals based on 20 independent studies. |
| Cross-sectional module and cosine bootstrap endpoints | Descriptive ranges obtained while holding source normalization, mapping and modules fixed. Whole pairs or recorded identifier clusters are resampled as specified in the manuscript. The GSE281841 cluster definition depends on literal recorded identifiers whose distinct-person interpretation is unconfirmed. These ranges do not resolve confounding, identity uncertainty or longitudinal validation. |
| `results_reviewer_revision/simulation_replicate_metrics.csv`: `test_population_oracle_MAE` | Historical field name for **empirical finite-test oracle MAE**, not an exact population constant. For each generated replicate, the code forms `oracle = B_test @ test_A.T + test_F_offset`, then averages `abs(F_test - oracle)` over the 2,000 test observations and eight modules. The value varies with the generated test sample and is repeated across the six method rows sharing that replicate. Those repeated method rows are not independent oracle measurements. No column or saved result is renamed. |
| Analytic population-oracle MAE | In the independent-follow-up, unit-noise Gaussian scenario, the expectation is `sqrt(2/pi) = 0.7979` when rounded to four decimals. This analytic expectation is distinct from the finite-test empirical oracle field above. The finite-training expected MAEs are `sqrt(2/pi) * sqrt(1 + 1/n)` for mean follow-up and `sqrt(2/pi) * sqrt(2 + 2/n)` for mean change. |
| Simulation MCSE | The standard deviation across 100 independent generated training/test replicates divided by `sqrt(100)`, calculated within the scenario/training-size/method cell. Paired differences use matched methods within each replicate. MCSE describes simulation precision, not uncertainty for a patient population. It is distinct from simulation replicate quantiles and from real-data fixed-loss resampling ranges. |

The source implementation for the oracle field is `run_known_truth_simulation.py`; it remains unchanged. All numerical result files and their column names remain unchanged by this clarification.
