## Supplementary material for "VascFlux-GEO: Comparator selection and auditability in vascular transcriptome module prediction": Analysis code and aggregate results: validate_inputs.py

```
"""Validate GEO input hashes and complete table structure before any refitting.

A matching hash authenticates a frozen file, not its completeness. This guard was
added after a truncated historical GPL570 export was discovered on 2026-10-25.
"""
import argparse,gzip,hashlib,json
from pathlib import Path

def inspect_table(path):
    path=Path(path)
    matrix='_series_matrix' in path.name
    begin=b'!series_matrix_table_begin' if matrix else b'!platform_table_begin'
    end=b'!series_matrix_table_end' if matrix else b'!platform_table_end'
    with path.open('rb') as signature:
        compressed=signature.read(2)==b'\x1f\x8b'
    opener=gzip.open if compressed else open
    active=False;ended=False;rows=0;header=None;digest=hashlib.sha256();seen=set();width=None
    with opener(path,'rb') as stream:
        for line in stream:
            if not active and line.startswith(begin):active=True;continue
            if active and line.startswith(end):ended=True;break
            if active:
                normalized=line.rstrip(b'\r\n')+b'\n'
                digest.update(normalized)
                fields=normalized.rstrip(b'\n').split(b'\t')
                if header is None:
                    header=normalized; width=len(fields)
                    names=[x.strip(b'"') for x in fields]
                    if names[0] != (b'ID_REF' if matrix else b'ID') or width < 2:
                        raise ValueError(f'Unexpected table header: {path.name}')
                    if len(names) != len(set(names)):
                        raise ValueError(f'Duplicate column header: {path.name}')
                    if not matrix and not any(x in names for x in (b'Gene Symbol',b'Gene symbol',b'Symbol',b'Gene Symbol (human)',b'gene_assignment')):
                        raise ValueError(f'Missing gene annotation column: {path.name}')
                else:
                    rows+=1
                    if len(fields)!=width:
                        raise ValueError(f'Ragged table row {rows}: {path.name}; expected {width} columns, found {len(fields)}')
                    key=fields[0].strip(b'" ')
                    if not key or key in seen:
                        raise ValueError(f'Empty or duplicate probe ID at row {rows}: {path.name}')
                    seen.add(key)
    if not active or not ended or header is None or rows==0:
        raise ValueError(f'Incomplete GEO table: {path.name}; begin={active}, end={ended}, rows={rows}')
    if not header.lstrip(b'"').startswith(b'ID'):
        raise ValueError(f'Unexpected table header: {path.name}')
    return {'table_rows':rows,'table_columns':width,'unique_probe_ids':len(seen),'table_end_present':True,'normalized_table_sha256':digest.hexdigest()}

def validate(data,manifest_path):
    manifest=json.loads(Path(manifest_path).read_text(encoding='utf-8-sig'))
    if isinstance(manifest,list):manifest={r['filename']:r for r in manifest}
    checks=[]
    for name,rec in manifest.items():
        path=Path(data)/name
        h=hashlib.sha256()
        with path.open('rb') as f:
            for block in iter(lambda:f.read(1024*1024),b''):h.update(block)
        if h.hexdigest()!=rec['sha256'] or path.stat().st_size!=rec['bytes']:
            raise ValueError(f'Frozen input hash/length mismatch: {name}')
        info=inspect_table(path)
        if rec.get('expected_table_rows') is not None and info['table_rows']!=rec['expected_table_rows']:
            raise ValueError(f'Table row count mismatch: {name}')
        if rec.get('normalized_table_sha256') and info['normalized_table_sha256']!=rec['normalized_table_sha256']:
            raise ValueError(f'Table content mismatch: {name}')
        checks.append({'filename':name,'sha256':h.hexdigest(),'bytes':path.stat().st_size,**info})
    return {'status':'passed','scope':'Selected input hash, complete first table, optional row count and normalized table digest; source-study metadata truth and upstream preprocessing are not certified.','files':checks}

def main():
    p=argparse.ArgumentParser();p.add_argument('--data-dir',type=Path,required=True);p.add_argument('--source-manifest',type=Path,required=True);p.add_argument('--report',type=Path,required=True)
    a=p.parse_args();report=validate(a.data_dir,a.source_manifest);a.report.parent.mkdir(parents=True,exist_ok=True);a.report.write_text(json.dumps(report,indent=2)+'\n',encoding='utf-8');print(json.dumps(report,indent=2))
if __name__=='__main__':main()
```
