## Supplementary material for "VascFlux-GEO: Comparator selection and auditability in vascular transcriptome module prediction": Analysis code and aggregate results: verify_extended_results.py

```
"""Verify completed numerical artifacts after both extended analysis scripts."""
import argparse
import json
from pathlib import Path

import numpy as np
import pandas as pd


def main():
    parser = argparse.ArgumentParser()
    parser.add_argument('--output-dir', type=Path, default=Path(__file__).resolve().parent / 'results_extended')
    p = parser.parse_args().output_dir
    summary = json.loads((p / 'summary_extended.json').read_text())
    d = pd.read_csv(p / 'oof_predictions.csv')
    assert len(d) == (72 + 14) * 20 * 8 * 11
    assert d.groupby(['cohort', 'repeat', 'method', 'subject']).size().eq(8).all()
    assert d.groupby(['cohort', 'repeat', 'subject', 'module']).fold.nunique().eq(1).all()
    assert np.isfinite(d[['observed_followup', 'observed_delta', 'predicted_followup',
                         'predicted_delta', 'absolute_error', 'squared_error']].to_numpy()).all()
    assert np.allclose(d.absolute_error, np.abs(d.observed_delta - d.predicted_delta), atol=1e-12)
    assert len(pd.read_csv(p / 'subject_folds.csv')) == 86 * 20
    assert len(pd.read_csv(p / 'nested_tuning.csv')) == 2 * 20 * 5 * (6 * 3 + 5)
    null = pd.read_csv(p / 'coupling_preserving_null.csv')
    assert len(null) == 2 * 201 * 4
    assert null.groupby(['cohort', 'null_repetition']).size().eq(4).all()
    fault = pd.read_csv(p / 'injected_fault_controls.csv')
    assert len(fault) == 14 and fault.passed.all()
    assert fault[fault.injected_failure].detected.all()
    assert (~fault[~fault.injected_failure].detected).all()
    con = pd.read_csv(p / 'corrected_cross_bed_concordance.csv')
    assert len(con) == 10
    assert np.isfinite(con[['cosine', 'conditional_ci95_low', 'conditional_ci95_high']]).all().all()
    checks = {'all_checks_passed': True, 'n_oof_module_rows': len(d),
        'n_distinct_prediction_subjects': 86, 'n_subject_fold_rows': 1720,
        'n_methods': len(summary['methods']), 'n_repeats': summary['n_outer_repeats'],
        'n_null_rows': len(null), 'n_injected_faults': 12, 'n_clean_mutation_controls': 2,
        'n_corrected_concordance_pairs': len(con),
        'checks': ['each subject has 8 module rows per method per repeat',
                   'identical held-out folds across methods', 'finite numeric predictions and errors',
                   'followup/implied-delta absolute errors identical', 'complete nested-tuning candidate grid',
                   'complete null repetitions', 'all injected faults detected and clean controls invariant',
                   'ten corrected descriptive group comparisons']}
    (p / 'output_checks.json').write_text(json.dumps(checks, indent=2), encoding='utf-8')
    print(json.dumps(checks, indent=2))


if __name__ == '__main__':
    main()
```
