## Supplementary material for "VascFlux-GEO: Comparator selection and auditability in vascular transcriptome module prediction": Analysis code and aggregate results: verify_revision_outputs.py

```
"""Check archived v2 numerical artifacts; this does not refit any model."""
import argparse
import hashlib
import json
from pathlib import Path
import numpy as np
import pandas as pd

ROOT = Path(__file__).resolve().parent


def sha(path):
    return hashlib.sha256(path.read_bytes()).hexdigest()


def prediction_check(frame, keys, expected_rows):
    assert len(frame) == expected_rows, (len(frame), expected_rows)
    assert not frame.duplicated(keys).any(), 'Duplicate OOF keys'
    columns = ['observed_followup', 'predicted_followup', 'absolute_error']
    assert np.isfinite(frame[columns].to_numpy(float)).all()
    expected_error = np.abs(frame.observed_followup-frame.predicted_followup)
    assert np.allclose(frame.absolute_error, expected_error, atol=1e-12, rtol=0)
    return {'rows': len(frame), 'unique_keys': True, 'finite': True,
            'error_recomputed_from_prediction': True}


def external_selection_check(sample, summary):
    """Reconstruct the exclusion predicate from recorded source scores, not IDs."""
    assert not sample['sample'].duplicated().any(), 'Duplicate source sample'
    assert sample.patient.eq(sample.patient_id_title).all()
    assert sample.patient.eq(sample.patient_id_source_name).all()
    raw_score = pd.to_numeric(sample.stenosis_score_raw, errors='coerce')
    assert np.allclose(raw_score, sample.stenosis_score, equal_nan=True)
    expected_group = pd.Series(np.where(raw_score.isin([4, 5]), 'high_4_5',
        np.where(raw_score.isin([0, 1]), 'low_0_1', 'ineligible')), index=sample.index)
    assert sample.group.eq(expected_group).all(), 'Grade-group mismatch'
    eligible = expected_group.ne('ineligible')
    assert sample.extreme_score_eligible.eq(eligible).all(), 'Eligibility mismatch'
    grade_counts = sample.loc[eligible].groupby('patient').group.nunique()
    mixed = set(grade_counts[grade_counts > 1].index)
    expected_inclusion = eligible & ~sample.patient.isin(mixed)
    assert sample.included_primary.eq(expected_inclusion).all(), 'Exclusion predicate mismatch'
    expected_reason = np.where(expected_inclusion,
        'included: extreme score; one grade group per patient',
        np.where(sample.patient.isin(mixed), 'excluded: patient has both high and low grades',
                 'excluded: missing or non-extreme stenosis score'))
    assert sample.selection_reason.eq(expected_reason).all(), 'Exclusion reason mismatch'
    assert set(summary['mixed_grade_patients_excluded']) == mixed
    assert set(summary['unscored_sample_ids']) == set(sample.loc[raw_score.isna(), 'sample'])
    assert int(raw_score.isna().sum()) == 1 and len(mixed) == 1
    assert int(sample.patient.isin(mixed).sum()) == 2
    selected = sample.loc[expected_inclusion]
    assert not selected.patient.isin(mixed).any()
    assert selected.stenosis_score.isin([0, 1, 4, 5]).all()
    assert len(sample) == summary['source_samples'] == 41
    assert sample.patient.nunique() == summary['source_patient_ids'] == 28
    assert int(eligible.sum()) == summary['usable_extreme_samples'] == 40
    assert sample.loc[eligible].patient.nunique() == summary['patient_count_before_excluding_mixed'] == 27
    assert len(selected) == summary['retained_samples'] == 38
    assert selected.patient.nunique() == summary['patient_count_after_excluding_mixed'] == 26
    return selected


def main():
    p = argparse.ArgumentParser(description=__doc__)
    p.add_argument('--core-dir', type=Path, default=ROOT / 'results_extended')
    p.add_argument('--revision-dir', type=Path, default=ROOT / 'results_reviewer_revision')
    p.add_argument('--report', type=Path, default=ROOT / 'provenance/revision_output_verification.json')
    args = p.parse_args()
    core, rev = args.core_dir.resolve(), args.revision_dir.resolve()
    checks = {}
    fair = pd.read_csv(rev / 'fair_oof_predictions.csv')
    keys = ['cohort', 'repeat', 'fold', 'subject', 'module', 'method']
    checks['fair'] = prediction_check(fair, keys, 86*20*8*4)
    assert fair.groupby(['cohort', 'repeat', 'subject', 'method']).size().eq(8).all()
    assert np.allclose(fair.predicted_followup-fair.baseline, fair.predicted_delta, atol=1e-12)
    expected_folds = pd.read_csv(core/'subject_folds.csv')[['cohort','repeat','subject','fold']]
    joined = fair.merge(expected_folds, on=['cohort','repeat','subject'], suffixes=('', '_expected'), validate='many_to_one')
    assert len(joined) == len(fair) and joined.fold.eq(joined.fold_expected).all()
    checks['fair']['same_core_subject_folds'] = True
    sim = pd.read_csv(rev / 'simulation_replicate_metrics.csv')
    assert len(sim) == 4*2*100*6
    assert not sim.duplicated(['scenario', 'train_n', 'repeat', 'method']).any()
    assert sim.groupby(['scenario','train_n','method']).size().eq(100).all()
    assert np.isfinite(sim[['MAE','RMSE','test_population_oracle_MAE']].to_numpy()).all()
    assert sim.max_followup_change_residual_difference.max() < 1e-12
    analytic = pd.read_csv(rev / 'simulation_analytic_null_checks.csv')
    assert (np.abs(analytic.analytic_expected_MAE-analytic.observed_MC_mean) <= analytic.tolerance_5_MCSE_plus_0p002).all()
    checks['simulation'] = {'metric_rows':len(sim), 'all_48_cells_have_100_replicates':True,
                            'analytic_null_tolerances_pass':True, 'not_patient_validation':True}
    robust = pd.read_csv(rev / 'robustness_annotation_oof.csv')
    checks['annotation'] = prediction_check(robust, ['mapping_scheme']+keys, 86*5*8*4*2)
    ablation = pd.read_csv(rev / 'robustness_input_module_ablation_oof.csv')
    checks['input_ablation'] = prediction_check(ablation,
        ['cohort','repeat','fold','subject','removed_input_module','target_module'],86*5*8*8)
    mutation = pd.read_csv(rev / 'robustness_full_nested_future_mutation.csv')
    assert len(mutation) == 8 and mutation.passed.all()
    assert (mutation.max_candidate_loss_difference <= 1e-12).all()
    assert (mutation.max_prediction_difference <= 1e-12).all()
    assert (mutation.heldout_followup_score_max_difference > 0).all()
    checks['future_outcome_mutation'] = {'representative_fits':8,'nested_tuning_invariance':True,
                                       'scope':'these exercised paths only'}
    record = json.loads((rev/'summary_representation_sensitivity.json').read_text())
    for filename, expected in record['outputs'].items():
        assert sha(rev/filename) == expected, f'Robustness result hash changed: {filename}'
    checks['robustness_output_manifest_matches'] = True
    sample = pd.read_csv(core/'external_GSE281841_sample_audit.csv')
    patients = pd.read_csv(core/'external_GSE281841_patient_scores.csv')
    contrast = pd.read_csv(core/'external_GSE281841_direction_holdout.csv')
    modules = list(contrast.module)
    external_summary = json.loads((core/'summary_external_GSE281841.json').read_text(encoding='utf-8'))
    selected = external_selection_check(sample, external_summary)
    assert len(sample)==41 and sample.patient.nunique()==28
    assert len(selected)==38 and len(patients)==26 and not patients.patient.duplicated().any()
    assert patients.groupby('group').size().to_dict()=={'high_4_5':18,'low_0_1':8}
    recomputed = selected.groupby(['patient','group'])[modules].mean().sort_index()
    archived = patients.set_index(['patient','group'])[modules].sort_index()
    assert recomputed.index.equals(archived.index)
    assert np.allclose(recomputed.to_numpy(),archived.to_numpy(),atol=1e-12,rtol=0)
    high=patients[patients.group=='high_4_5'][modules].mean()
    low=patients[patients.group=='low_0_1'][modules].mean()
    assert np.allclose(contrast.mean_high_minus_low,high-low,atol=1e-12,rtol=0)
    checks['external_description']={'source_columns':41,'exact_patient_ids':28,
        'retained_samples':38,'retained_patient_ids':26,'patient_aggregation_recomputed':True,
        'exclusion_predicate_recomputed_from_source_scores':True,
        'exclusion_reasons_and_summary_consistent':True,
        'group_contrast_recomputed':True,'role':'cross-sectional description only'}
    result = {'status':'passed','mode':'archived_output_consistency_only_no_refit', 'checks':checks}
    args.report.parent.mkdir(parents=True, exist_ok=True)
    args.report.write_text(json.dumps(result, indent=2)+'\n',encoding='utf-8')
    print(json.dumps(result, indent=2))


if __name__ == '__main__':
    main()
```
